# Consensus recommendations for supporting people waiting for total hip and knee arthroplasty in Scotland: A modified Delphi study

**DOI:** 10.64898/2026.02.09.26345536

**Authors:** Kay Cooper, Emma Stage, Erin Hart-Winks, Paul Swinton, Lyndsay Alexander, Jo Shim, Tom Herbert, Stephen Bridgman

**Author notes:** Corresponding author: Kay Cooper, School of Health, Ishbel Gordon Building, Robert Gordon University, Garthdee Road, Aberdeen, AB10 7QG.

## Abstract

**Background:** Many people experience long waiting times for hip and knee replacement surgery, negatively impacting physical conditioning and quality of life, and therefore need evidence-informed strategies to support them while they are waiting.

**Objective:** To develop consensus-based recommendations for supporting people waiting for hip or knee replacement in Scotland.

**Design:** Three-round online modified Delphi study involving 17 people with experience of waiting for hip or knee replacement and 30 professionals involved in supporting them.

**Methods:** Two rounds of online survey and one online workshop. Participants rated agreement with 113 (round 1), 40 (round 2) and 20 (round 3) statements on a 5-point Likert scale, with consensus based on ≥70% agreement. Items were modified and added in subsequent rounds based on content analysis of participant comments. The final recommendations represent items that reached consensus in both participant groups after 3 rounds.

**Results:** 47 participants took part in round 1 (17 patients, 30 professionals), 41 in round 2 (13 patients, 28 professionals) and 23 in round 3 (8 patients, 15 professionals). Eighty-two of 113 statements reached consensus in round one, 20 of 40 in round two and 6 of 20 in round 3. The final recommendations comprise 108 statements relating to: preoperative education; patient optimisation; other interventions to support people waiting; and, strategies to support people waiting a long time for surgery.

**Conclusions:** These findings are an important step towards developing best practice guidance for supporting people waiting for hip and knee replacement in Scotland.

## Introduction

Osteoarthritis is the most common form of arthritis in adults and a leading cause of chronic pain and long-term disability (Steinmetz et al., 2023). Osteoarthritis affects an estimated 7.6% of the global population(Steinmetz et al., 2023), and case numbers are projected to increase by nearly 50% between 2020 and 2050 (Jennison, MacGregor, & Goldberg, 2023). Arthroplasty is recommended for the hip, knee and shoulder where non-surgical management of osteoarthritis is ineffective, and symptoms substantially impair quality of life (NICE 2022) (National Institute for Health and Care Excellence, 2022; The Royal Australian College of General Practitioners, 2025). Total hip arthroplasty (THA) and total knee arthroplasty (TKA) are among the most frequently performed surgical procedures in the UK NICE, 2020). In Scotland alone, 7,968 THA and 7,124 TKA surgeries were performed in 2023 (Public Health Scotland, 2024). Both procedures are predicted to rise over the next 20-30 years in line with changing population demographics (Farrow, McLoughlin, Gaba, & Ashcroft, 2023; Shichman et al., 2023).

Many health systems face challenges in meeting the demand for THA and TKA, a situation exacerbated by the surgical backlog caused by the COVID-19 pandemic (French et al., 2024; T W Wainwright, Immins, & Middleton, 2024; Yapp, Clarke, Moran, Simpson, & Scott, 2021). In Scotland this has led to unprecedented waiting list pressures, with thousands of patients on waiting lists and the 18-week referral-to-treatment standard frequently not being met (Farrow et al., 2023; Public Health Scotland, 2025). Prolonged waiting times not only extend the duration of disability and reduced quality of life but also increase the risk of physical deconditioning, thereby increasing the risk of surgical complications and adverse post-operative outcomes (Ackerman, Bennell, & Osborne, 2011). It is therefore important that patients waiting for THA and TKA are appropriately supported to optimise their health during this period.

Preoperative education and prehabilitation for THA and TKA are widely adopted evidence-based interventions (Omar, Wylde, Fogg, Whitehouse, & Bertram, 2025); however, they were developed and implemented when waiting times were not at the levels currently being experienced in Scotland and elsewhere. Waiting several months, or in some cases over a year for surgery, may require a different approach to supporting people, to prevent excessive deconditioning and maintain quality of life as far as possible. A survey of 16 Scottish Health Boards (14 regional; 2 special) undertaken in preparation for this study identified a range of interventions, in addition to preoperative education and rehabilitation, that had been developed by various health boards to support people waiting for THA and TKA. These included one-off ‘waiting well’ consultations; interventions aimed at weight management, smoking cessation, alcohol reduction and supporting mental wellbeing; provision of social support, and signposting to online resources and third sector organisations. Provision was highly variable, with no single health board providing all intervention types, indicating a lack of consensus on the optimal approach to supporting people waiting for these procedures.

The purpose of this modified Delphi study was therefore to develop recommendations for supporting people waiting for THA and TKA in Scotland based on evidence and consensus. Previous consensus studies have addressed TKA education and prehabilitation (Anderson et al., 2021), occupational advice for patients undergoing THA and TKA (Baker et al., 2020), and aspects of THA and TKA care including wound closure and dressing management (Ainslie-Garcia et al., 2024) and perioperative care (Plenge et al., 2018). Farrow et al. (2023) identified patients’ and surgeons’ preferences for prioritising patients waiting for THA and TKA. However, no previous consensus-based studies have focused supporting patients throughout the entire waiting period for such surgery.

## Methods

We conducted a three-round online modified Delphi study (Figure 1), with rounds one and two using online surveys and round three conducted as workshop discussions. The study was guided by methodological recommendations (Beiderbeck, Frevel, von der Gracht, Schmidt, & Schweitzer, 2021) and previous research in the field (Ainslie-Garcroia et al., 2024; Fearon et al., 2024). Round 1 (Supplementary file 1) was informed by: i) an evidence review conducted by the study team of more than 450 evidence sources (manuscripts in preparation); ii) the survey of Scottish Health Boards described above; iii) a desktop review of publicly available information aimed at supporting people waiting for THA and TKA in Scotland, and iv) previous consensus statements and recommendations (Anderson et al., 2021; Thomas W Wainwright et al., 2019). The study is reported in line with guidance on conducting and reporting Delphi studies (CREDES)(Jünger, Payne, Brine, Radbruch, & Brearley, 2017). The project was overseen by a Project Advisory Group (PAG) comprising the project team (n=9), individuals with lived experience of waiting for THA or TKA (n=4), healthcare professionals (n=11), researchers (n=2), policy makers (n=5) and third sector representatives (n=4). The PAG met six times during the study, including reviewing the draft recommendations. Ethical approval was granted by the Robert Gordon University School of Health Sciences Research Ethics Committee (Ref: SHS 24/25) and the study was approved by the NHS Research Scotland Permissions Coordinating Centre (Ref: NRS24/344449).

**Figure 1:**
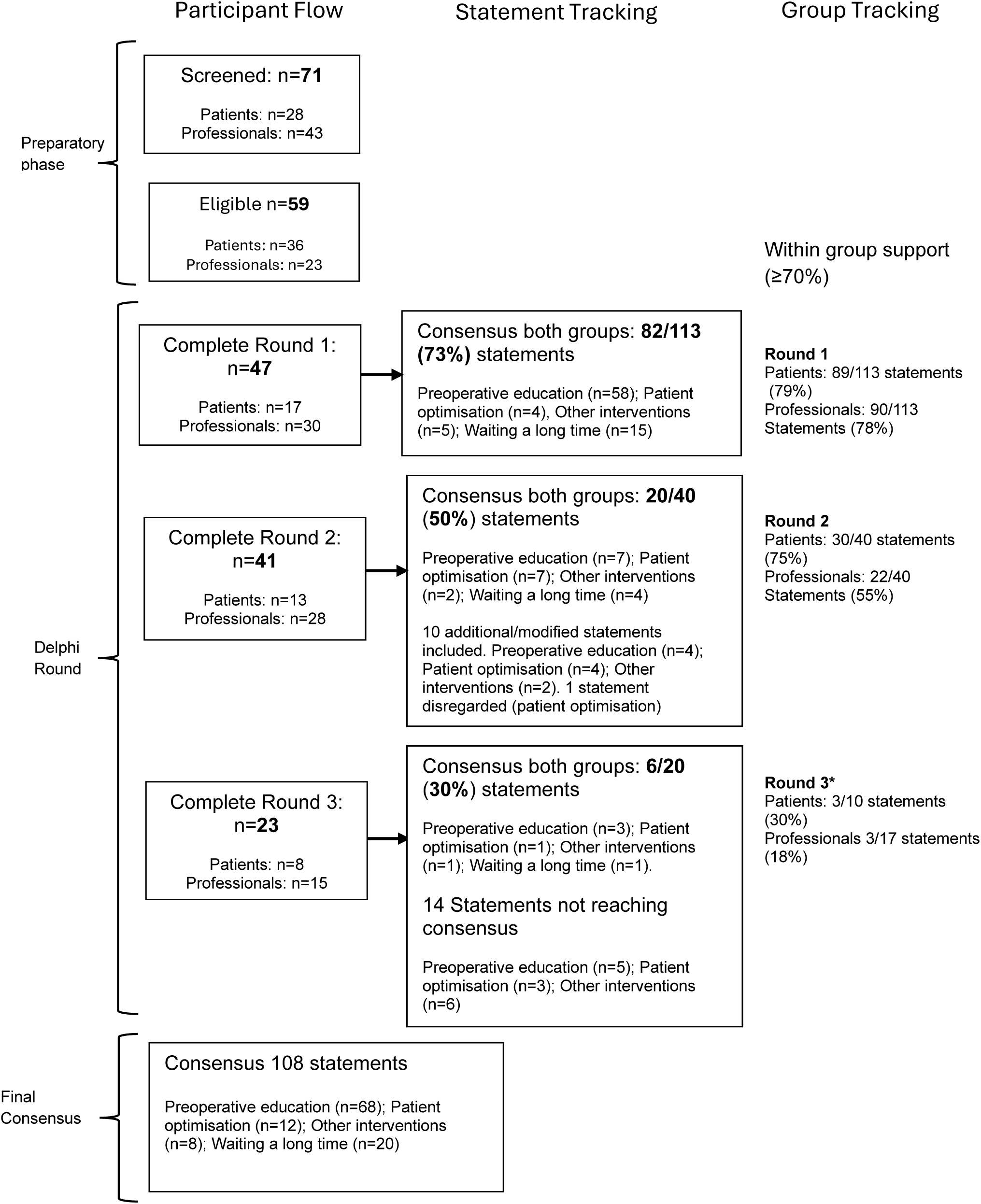
Flowchart of Delphi Process. *Different questions discussed in Patients & Professionals workshops respectively.

### Expert panel

We recruited two sub-groups of experts to our panel: i) people with lived experience of waiting for THA or TKA by way of currently being on a waiting list or having had surgery following a wait (hereafter ‘patients’), and ii) professionals with expertise in the management of people waiting for THA and TKA, including orthopaedic surgeons, physiotherapists, occupational therapists, nurses, service managers and funders, and published researchers in the field (hereafter ‘professionals’). As this study related to the Scottish context, due to the devolved nature of health services in Scotland, participants were mostly drawn from the Scottish population. However, we also included professionals from outwith Scotland to reduce bias. Similar to other consensus-based studies (Ainslie-Garcia et al., 2024), PAG members were eligible to take part as they could provide highly relevant expertise. Sample sizes for Delphi studies are highly variable, with 30 widely recommended as the minimum for statistical analysis purposes (de Villiers, de Villiers, & Kent, 2005), 15 per group for subgroup analyses (Beiderbeck et al., 2021) and no recommended maximum. We aimed to recruit at least 15 participants in each population sub-group. Table 1 outlines the inclusion criteria and recruitment strategy for each subgroup. Patients were recruited through public channels (social media, health board public involvement networks, third sector organisations) and professionals by email invitation followed by up to 2 reminders. Participants who responded were provided with a participant information sheet, unique identifier and link to provide consent and demographic information via the GDPR-compliant Jisc online survey platform (Supplementary file 1). The unique identifiers were used by each participant on each survey round and enabled the research team to send targeted reminders. Participants could complete a paper copy of the survey and return by freepost if they preferred; no participants requested this option. The survey was piloted by the research team, PAG members and four patient representatives. Minor wording and structural changes were made based on piloting, and the average completion time was estimated at 30 minutes.

**Table 1:** Inclusion criteria and recruitment strategy.

|  | **Inclusion Criteria** | **Recruitment strategy** |
| --- | --- | --- |
| **People with lived experience of waiting for THA/TKA** | Listed for THA/TKA or had THA/TKA past 6-months<br>Aged 18+<br>Resident in Scotland | Social media accounts of project team & PAG (X & Facebook) & UK THA/TKA Facebook groups<br>Health Board Public Involvement Networks<br>Third sector organisations e.g., Versus Arthritis, The Alliance |
| **Experts in the management of people waiting for THA/TKA** | HCPs with experience of managing people on waiting lists for THA/TKA (e.g., Orthopaedic consultant, GP, Allied Health Professional, Public health/Health Promotion/Health Improvement specialist, Psychologist, Social worker)<br>Published researchers in the field of preoperative interventions for THA/TKA<br>Service managers & clinical directors at Health Board level; Health Board Directors<br>Individuals from Public Health Scotland, Scottish Arthroplasty Project, Scottish Government, Healthcare Improvement Scotland | Email invitation from research team to personal contacts in relevant organisations + snowballing<br>Email invitation from research team to PAG + snowballing<br>Email invitation to published researchers (based on publication record) identified by recent scoping review conducted by project team<br>Email invitation from research team & PAG members to professional groups + snowballing |

Key: TKA – total knee arthroplasty; THA – total hip arthroplasty; PAG – Project advisory group

### Data collection

#### Round one

Data were collected between August and November 2024. Panellists in round one received a Jisc online survey comprising 113 statements relevant to preoperative education (n=69), patient optimisation (n=12), other interventions to support people waiting for THA and TKA surgery (n=12) and extended waiting times for surgery (n=20). Panellists were asked to rate their level of agreement using a 5-point Likert scale (strongly disagree, disagree, neither agree nor disagree, agree, strongly agree). Statements were considered to have reached consensus when 70% or more agreement (agree or strongly agree) was achieved in both panel groups (Niederberger & Spranger, 2020). Statements that received 50% or greater disagreement (strongly disagree or disagree) from either group were removed from further consideration. Statements receiving less than 70% agreement in either group progressed to round two for reconsideration. Open-text boxes were included at the end of each set of statements to allow for panellists to suggest additional statements.

#### Round two

All panellists who completed round one were invited to participate in round two, conducted between December 2024 and January 2025. Panellists received a summary of their individual responses from round one along with a link to the round two survey (Supplementary file 1). All statements that reached consensus in the previous round were presented at the beginning of each survey section.

Items that did not reach consensus were accompanied by two charts showing the level of agreement from the patient and professional groups. Panellists were asked to reconsider these items using the same 5-point Likert scale. Round two included additional or modified statements based on the obtained open-text suggestions: preoperative education (n=4), Patient optimisation (n=4), other supportive interventions (n=2). Free-text boxes were also included in the round two survey for panellists to make additional suggestions, although no new items were generated. Statements that did not meet consensus progressed to round three.

#### Round three

Panellists who completed both surveys were invited to online workshops in February 2025. Six workshops (three per group) were held asynchronously. Prior to the workshops, panellists were provided with the presentation slides (Supplementary file 1) summarising results from the previous rounds, including consensus statements, exclusions, and items requiring further consideration. As the workshops were held separately for patients and professionals, each group only considered statements that had not reached consensus within their group.

Seventeen statements were considered by the professional group and ten by the patient group. Following discussion of each statement, panellists indicated their level of agreement using the same 5-point Likert scale via Microsoft Teams polls. In patient workshops, technical difficulties required responses to be given verbally and recorded by the research team.

### Data processing and analysis

Quantitative data were analysed descriptively using R (R Core Team, 2024). For each statement, the proportions of panellists selecting “agree” or “strongly agree” were combined, as were the proportions selecting “disagree” or “strongly disagree.” Statements that did not meet the threshold for agreement or removal due to disagreement were carried forward. Where applicable, comparisons between patient and professional responses were explored descriptively (see supplementary file 2). No imputation was applied for missing data.

For each survey, free text responses were analysed using content analysis (by ES, TH, EHW) to generate additional statements and discussion points for the workshops. Additional statements were finalised by a project co-lead (KC/LA).

### Equity, diversity and inclusion statement

When recruiting panel members every effort was made to ensure equity and diversity. This was monitored via demographic information including gender, age, sociodemographic background, THA and TKA type, professional discipline, and professional experience, collected for each panellist.

## Results

### Participants

Seventy-one people indicated an interest in taking part, and 59 subsequently consented (36 patients and 23 professionals) and received the round one survey link. Reasons for dropout at this stage included non-response, lack of relevant lived experience, and not being an eligible healthcare professional. Of the 59 panellists, 47 (patients = 17, professionals = 30) completed round one, 41 completed round two (patients = 13, professionals = 28) and 23 completed round three (patients = 8, professionals = 15). Table 2 describes participant demographics.

**Table 2:**
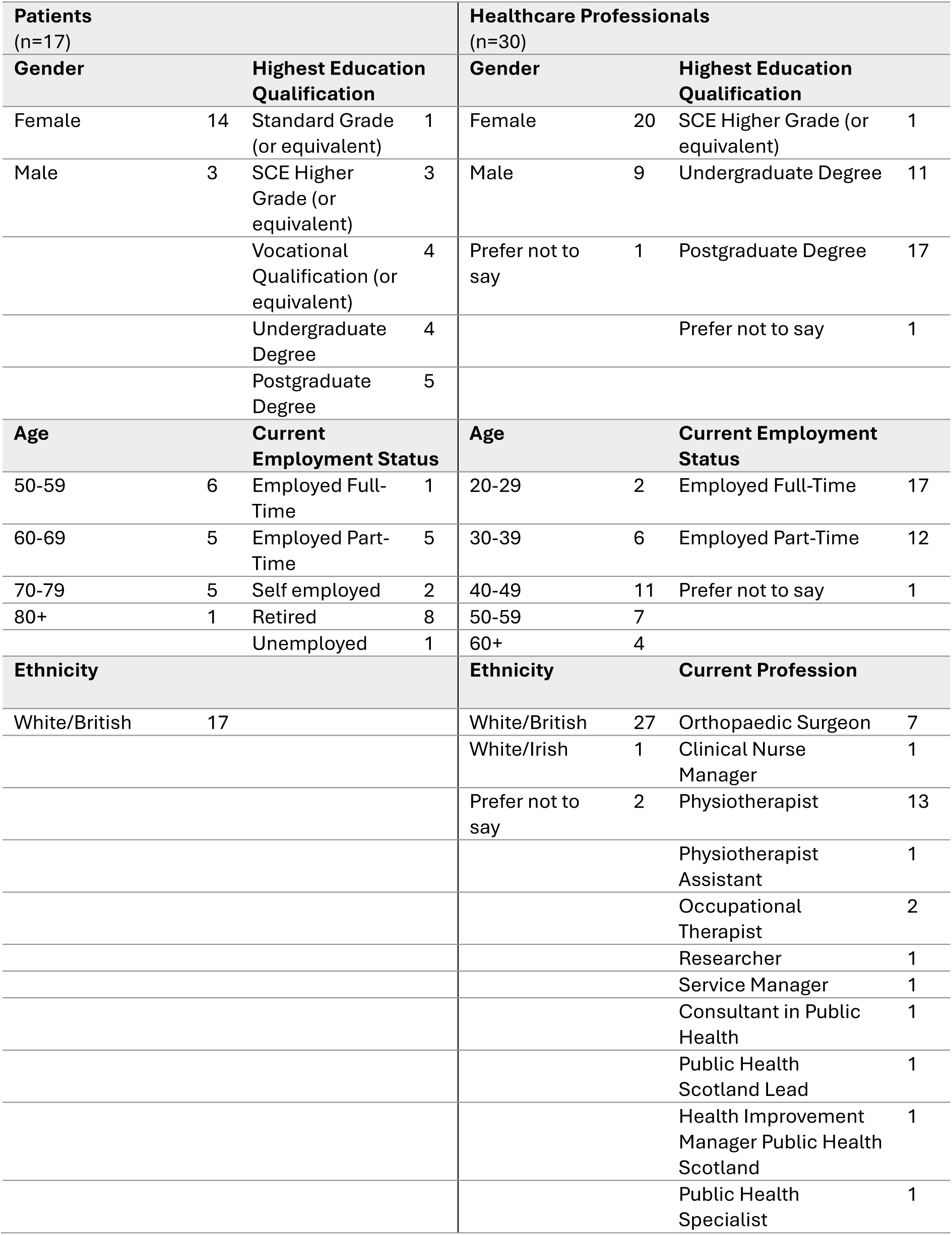
Characteristics of the panellists.

### Findings

Table 3 provides a summary of items reaching agreement in each round, and tables 4-7 provide a summary for each domain (preoperative education, patient optimisations, other interventions, waiting a long time).

**Table 3:** Summary of recommended items.

|  | Number of recommended items per round |  |  |  |  |  |  |  |  |  |  |  |
| --- | --- | --- | --- | --- | --- | --- | --- | --- | --- | --- | --- | --- |
|  | Round 1 |  |  |  | Round 2 |  |  |  | Round 3 |  |  |  |
|  | Total<sup>a</sup> | Pts | Profs | Consensus reached<sup>b</sup> | Total<sup>a</sup> | Pts | Profs | Consensus Reached<sup>b</sup> | Total<sup>a</sup> | Pts | Profs | Consensus Reached<sup>b</sup> |
| Preoperative education | 69 | 61 | 62 | 58 | 15 | 14 | 8 | 7 | 8 | 1 | 2 | 3 |
| Patient optimisation | 12 | 4 | 4 | 4 | 11 | 9 | 7 | 7 | 4 | 0 | 1 | 1 |
| Other interventions to support people waiting | 12 | 7 | 5 | 5 | 9 | 3 | 3 | 2 | 7 | 1 | 0 | 1 |
| Waiting a long time for surgery | 20 | 17 | 17 | 15 | 5 | 4 | 4 | 4 | 1 | 1 | 0 | 1 |
| **All items** | **113** | **89** | **90** | **82** | **40\*** | **30** | **22** | **20** | **20** | **3** | **3** | **6** |

Pts: Patient panellists; Profs: Professional panellists; <sup>a</sup> Total number of statements included in each round; <sup>b</sup> Total number of statements that reached consensus by both patients and professionals in each round. Consensus is defined as statements reaching ≥70% as suggested by panellists indicating “agree” or “strongly agree” for each statement; \*In Round 1 item 88 was voted “disagree” or “strongly disagree” in both groups and was excluded from the recommendations

**Table 4:**
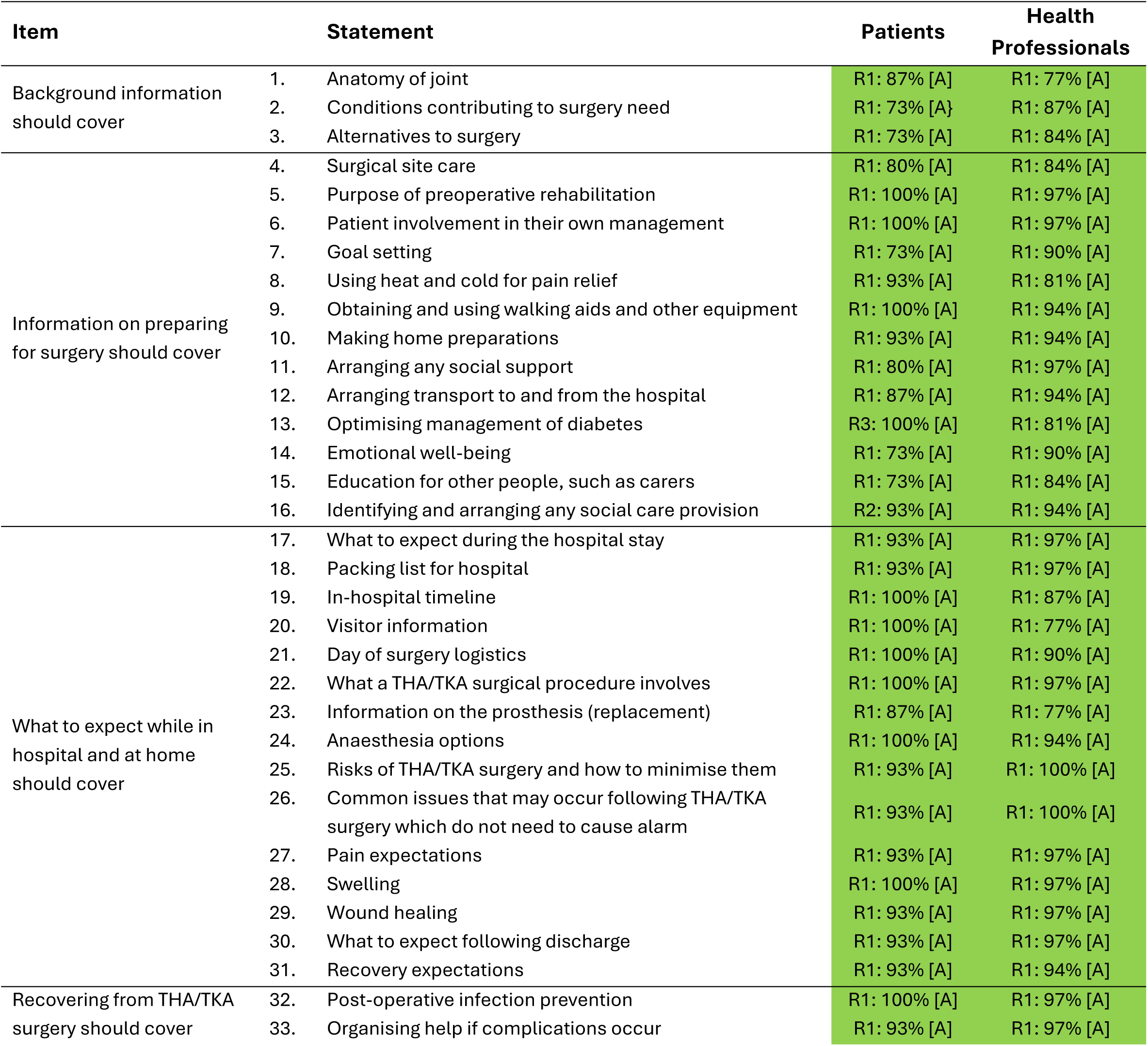
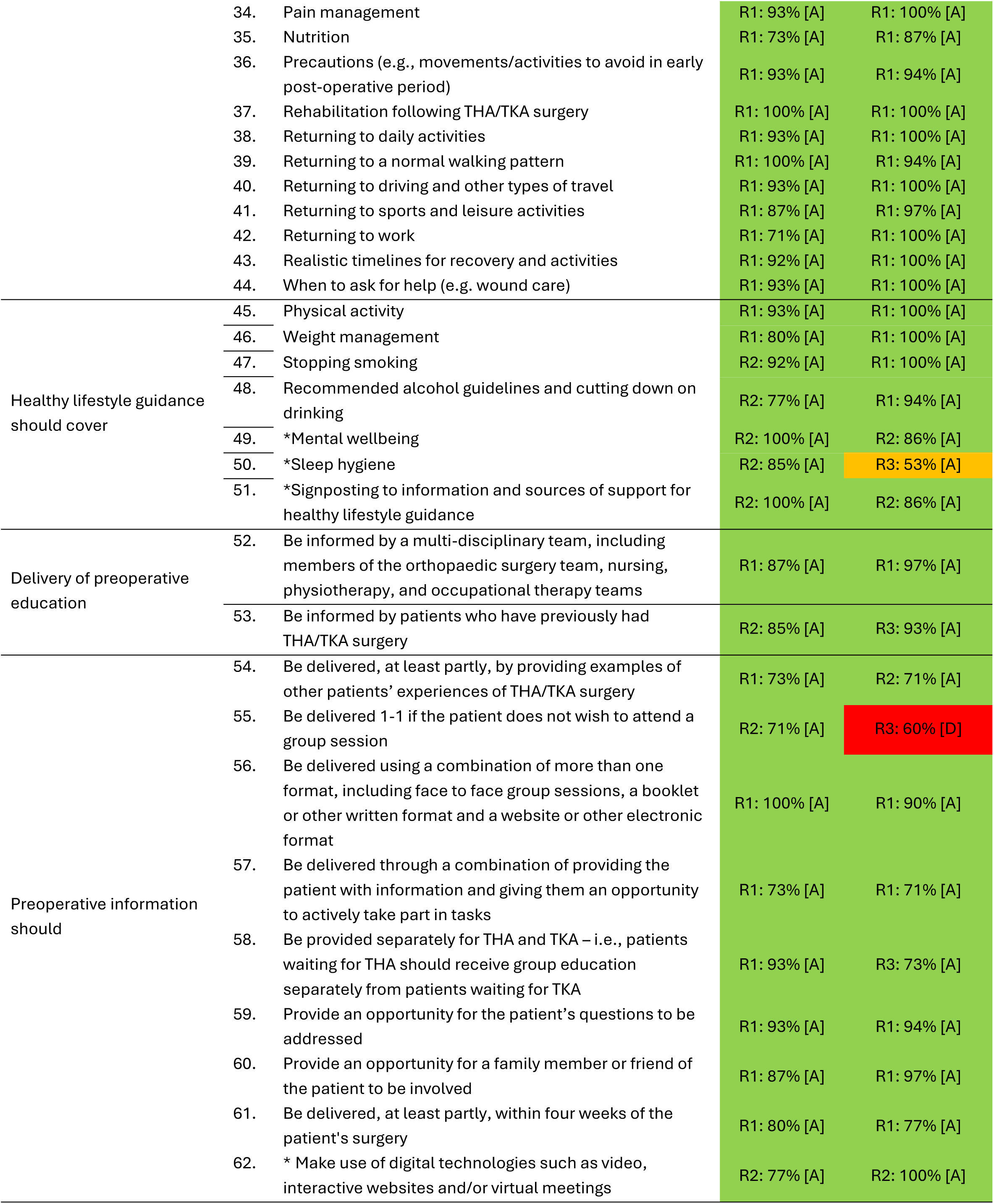
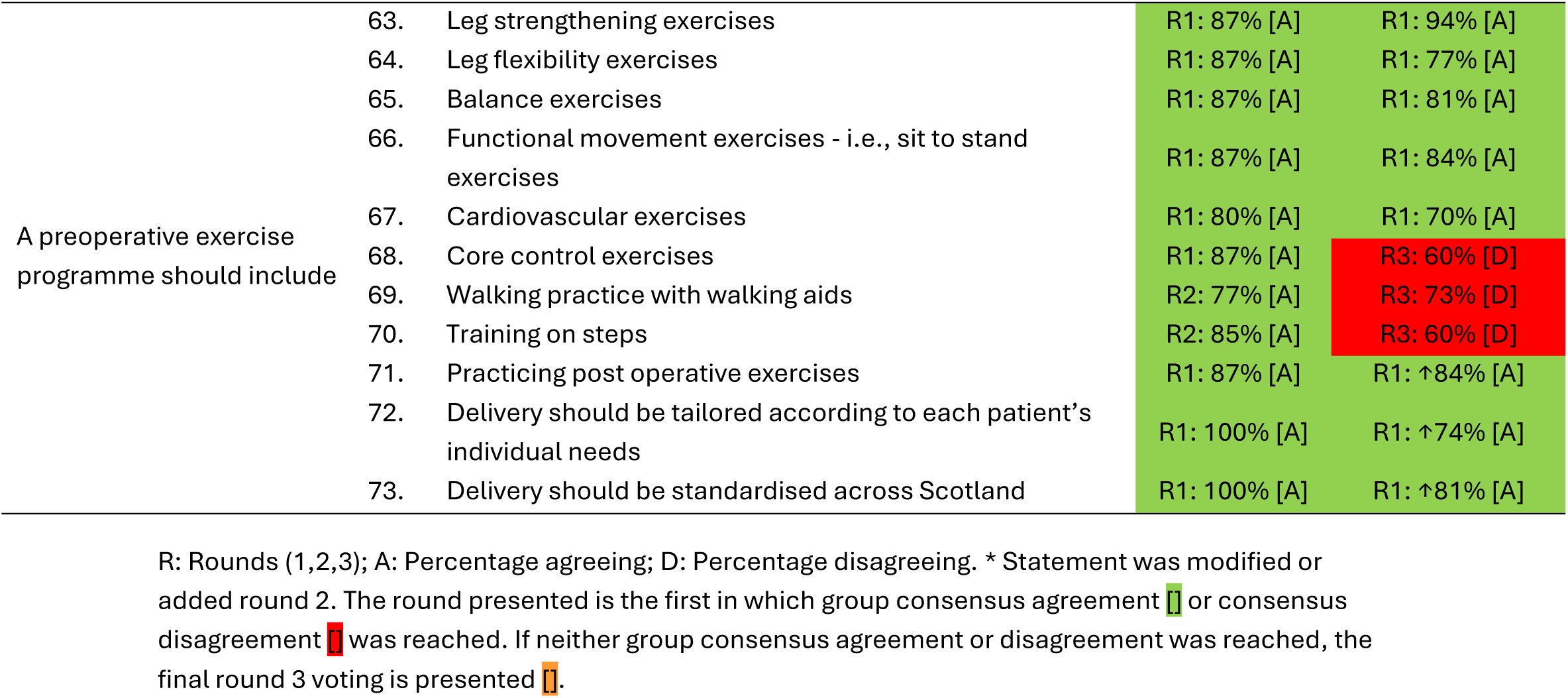
Summary of Delphi voting results from patients and health professionals on “preoperative education” statements.

| Item | Statement | Patients | Health Professionals |
| --- | --- | --- | --- |
| Background information should cover | 1. Anatomy of joint | R1: 87% [A] | R1: 77% [A] |
|  | 2. Conditions contributing to surgery need | R1: 73% [A] | R1: 87% [A] |
|  | 3. Alternatives to surgery | R1: 73% [A] | R1: 84% [A] |
| Information on preparing for surgery should cover | 4. Surgical site care | R1: 80% [A] | R1: 84% [A] |
|  | 5. Purpose of preoperative rehabilitation | R1: 100% [A] | R1: 97% [A] |
|  | 6. Patient involvement in their own management | R1: 100% [A] | R1: 97% [A] |
|  | 7. Goal setting | R1: 73% [A] | R1: 90% [A] |
|  | 8. Using heat and cold for pain relief | R1: 93% [A] | R1: 81% [A] |
|  | 9. Obtaining and using walking aids and other equipment | R1: 100% [A] | R1: 94% [A] |
|  | 10. Making home preparations | R1: 93% [A] | R1: 94% [A] |
|  | 11. Arranging any social support | R1: 80% [A] | R1: 97% [A] |
|  | 12. Arranging transport to and from the hospital | R1: 87% [A] | R1: 94% [A] |
|  | 13. Optimising management of diabetes | R3: 100% [A] | R1: 81% [A] |
|  | 14. Emotional well-being | R1: 73% [A] | R1: 90% [A] |
|  | 15. Education for other people, such as carers | R1: 73% [A] | R1: 84% [A] |
|  | 16. Identifying and arranging any social care provision | R2: 93% [A] | R1: 94% [A] |
| What to expect while in hospital and at home should cover | 17. What to expect during the hospital stay | R1: 93% [A] | R1: 97% [A] |
|  | 18. Packing list for hospital | R1: 93% [A] | R1: 97% [A] |
|  | 19. In-hospital timeline | R1: 100% [A] | R1: 87% [A] |
|  | 20. Visitor information | R1: 100% [A] | R1: 77% [A] |
|  | 21. Day of surgery logistics | R1: 100% [A] | R1: 90% [A] |
|  | 22. What a THA/TKA surgical procedure involves | R1: 100% [A] | R1: 97% [A] |
|  | 23. Information on the prosthesis (replacement) | R1: 87% [A] | R1: 77% [A] |
|  | 24. Anaesthesia options | R1: 100% [A] | R1: 94% [A] |
|  | 25. Risks of THA/TKA surgery and how to minimise them | R1: 93% [A] | R1: 100% [A] |
|  | 26. Common issues that may occur following THA/TKA surgery which do not need to cause alarm | R1: 93% [A] | R1: 100% [A] |
|  | 27. Pain expectations | R1: 93% [A] | R1: 97% [A] |
|  | 28. Swelling | R1: 100% [A] | R1: 97% [A] |
|  | 29. Wound healing | R1: 93% [A] | R1: 97% [A] |
|  | 30. What to expect following discharge | R1: 93% [A] | R1: 97% [A] |
|  | 31. Recovery expectations | R1: 93% [A] | R1: 94% [A] |
| Recovering from THA/TKA surgery should cover | 32. Post-operative infection prevention | R1: 100% [A] | R1: 97% [A] |
|  | 33. Organising help if complications occur | R1: 93% [A] | R1: 97% [A] |

|  |  |  |  |  |
| --- | --- | --- | --- | --- |
|  | 34. | Pain management | R1: 93% [A] | R1: 100% [A] |
|  | 35. | Nutrition | R1: 73% [A] | R1: 87% [A] |
|  | 36. | Precautions (e.g., movements/activities to avoid in early post-operative period) | R1: 93% [A] | R1: 94% [A] |
|  | 37. | Rehabilitation following THA/TKA surgery | R1: 100% [A] | R1: 100% [A] |
|  | 38. | Returning to daily activities | R1: 93% [A] | R1: 100% [A] |
|  | 39. | Returning to a normal walking pattern | R1: 100% [A] | R1: 94% [A] |
|  | 40. | Returning to driving and other types of travel | R1: 93% [A] | R1: 100% [A] |
|  | 41. | Returning to sports and leisure activities | R1: 87% [A] | R1: 97% [A] |
|  | 42. | Returning to work | R1: 71% [A] | R1: 100% [A] |
|  | 43. | Realistic timelines for recovery and activities | R1: 92% [A] | R1: 100% [A] |
|  | 44. | When to ask for help (e.g. wound care) | R1: 93% [A] | R1: 100% [A] |
| Healthy lifestyle guidance should cover | 45. | Physical activity | R1: 93% [A] | R1: 100% [A] |
|  | 46. | Weight management | R1: 80% [A] | R1: 100% [A] |
|  | 47. | Stopping smoking | R2: 92% [A] | R1: 100% [A] |
|  | 48. | Recommended alcohol guidelines and cutting down on drinking | R2: 77% [A] | R1: 94% [A] |
|  | 49. | *Mental wellbeing | R2: 100% [A] | R2: 86% [A] |
|  | 50. | *Sleep hygiene | R2: 85% [A] | R3: 53% [A] |
|  | 51. | *Signposting to information and sources of support for healthy lifestyle guidance | R2: 100% [A] | R2: 86% [A] |
| Delivery of preoperative education | 52. | Be informed by a multi-disciplinary team, including members of the orthopaedic surgery team, nursing, physiotherapy, and occupational therapy teams | R1: 87% [A] | R1: 97% [A] |
|  | 53. | Be informed by patients who have previously had THA/TKA surgery | R2: 85% [A] | R3: 93% [A] |
| Preoperative information should | 54. | Be delivered, at least partly, by providing examples of other patients' experiences of THA/TKA surgery | R1: 73% [A] | R2: 71% [A] |
|  | 55. | Be delivered 1-1 if the patient does not wish to attend a group session | R2: 71% [A] | R3: 60% [D] |
|  | 56. | Be delivered using a combination of more than one format, including face to face group sessions, a booklet or other written format and a website or other electronic format | R1: 100% [A] | R1: 90% [A] |
|  | 57. | Be delivered through a combination of providing the patient with information and giving them an opportunity to actively take part in tasks | R1: 73% [A] | R1: 71% [A] |
|  | 58. | Be provided separately for THA and TKA – i.e., patients waiting for THA should receive group education separately from patients waiting for TKA | R1: 93% [A] | R3: 73% [A] |
|  | 59. | Provide an opportunity for the patient's questions to be addressed | R1: 93% [A] | R1: 94% [A] |
|  | 60. | Provide an opportunity for a family member or friend of the patient to be involved | R1: 87% [A] | R1: 97% [A] |
|  | 61. | Be delivered, at least partly, within four weeks of the patient's surgery | R1: 80% [A] | R1: 77% [A] |
|  | 62. | * Make use of digital technologies such as video, interactive websites and/or virtual meetings | R2: 77% [A] | R2: 100% [A] |
| A preoperative exercise programme should include | 63. | Leg strengthening exercises | R1: 87% [A] | R1: 94% [A] |
|  | 64. | Leg flexibility exercises | R1: 87% [A] | R1: 77% [A] |
|  | 65. | Balance exercises | R1: 87% [A] | R1: 81% [A] |
|  | 66. | Functional movement exercises - i.e., sit to stand exercises | R1: 87% [A] | R1: 84% [A] |
|  | 67. | Cardiovascular exercises | R1: 80% [A] | R1: 70% [A] |
|  | 68. | Core control exercises | R1: 87% [A] | R3: 60% [D] |
|  | 69. | Walking practice with walking aids | R2: 77% [A] | R3: 73% [D] |
|  | 70. | Training on steps | R2: 85% [A] | R3: 60% [D] |
|  | 71. | Practicing post operative exercises | R1: 87% [A] | R1: ↑84% [A] |
|  | 72. | Delivery should be tailored according to each patient's individual needs | R1: 100% [A] | R1: ↑74% [A] |
|  | 73. | Delivery should be standardised across Scotland | R1: 100% [A] | R1: ↑81% [A] |

R: Rounds (1,2,3); A: Percentage agreeing; D: Percentage disagreeing. \* Statement was modified or added round 2. The round presented is the first in which group consensus agreement or consensus disagreement was reached. If neither group consensus agreement or disagreement was reached, the final round 3 voting is presented .

### Preoperative education (Table 4)

Sixty-nine statements on preoperative education were included in round one. Sixty-one reached consensus for patients and 62 for professionals, with 58 statements reaching consensus for both groups (Table 3). Of the four statements reaching consensus among professionals only, these included items related to diabetes management, social care provision, smoking cessation and alcohol advice (Items 13, 16, 47, 48). Of the three statements reaching consensus amongst patients only, these included items related to preoperative information being delivered, at least partly, by providing examples of other patients’ experiences (Item 54), being provided separately for THA and TKA (Item 58), and the inclusion of core control exercises (Item 68).

In round two, 15 statements (including four new or modified statements) were included. Fourteen achieved consensus for patients and eight for professionals, with seven reaching consensus for both groups. Three statements that had reached consensus by professionals in round one went on to achieve consensus for patients (Items 16, 47, 48). Three statements added in round two achieved consensus for both groups and were related to mental wellbeing advice, signposting to information and support, and the use of digital technologies (items 49, 51, 62). Five statements reached consensus by patients only and were related to sleep hygiene (Item 50), delivery of preoperative education (Item 53), preoperative information (Item 55) and preoperative exercise (Items 69, 70). Preoperative information including examples of patients’ experiences of THA/TKA which had reached consensus by patients in round one, reached consensus by professionals in round 2 (Item 54).

In round three (workshops), eight statements were included (Table 3). Patients reconsidered one statement (Item 13) that had reached consensus by professionals in round one and professionals reconsidered seven statements that had reached consensus in earlier rounds by patients (Items: 5, 53, 55, 58, 68-70). This resulted in three statements reaching consensus among both groups (items 13, 53, 58) which related to optimising diabetes management (Item 13), preoperative education being informed by people who previously had THA/TKA (Item 53), and preoperative information being provided separately for THA and TKA (Item 58). Discussion within the workshops revealed that patients were initially not confident to make a decision regarding optimising management of diabetes, which may have influenced responses in earlier rounds. However, once the context was discussed and clarified, patients identified the importance of managing diabetes, leading to consensus for this statement. Within the professionals’ workshops, five statements that had achieved consensus in earlier rounds by patients did not reach consensus within this group. These related to sleep hygiene, preoperative information being delivered one-to-one, and preoperative exercise types (Items 50, 55, 68-70). Sleep hygiene was a topic of debate with differing perspectives among panellists. There was agreement among both groups that lack of sleep is influenced by night pain. However, there was conflicting opinion on the importance of providing sleep hygiene guidance, with professionals expressing concerns about overloading patients with preoperative information.

### Patient Optimisation (table 5)

Twelve statements were included in round one, with four statements achieving consensus by both panellist groups (Table 3): These related to referral to smoking cessation programmes (Items 74, 75), referral to alcohol cessation programmes (Item 78), and investigating preoperative anaemia prior to THA/TKA surgery (Item 83).

**Table 5.**
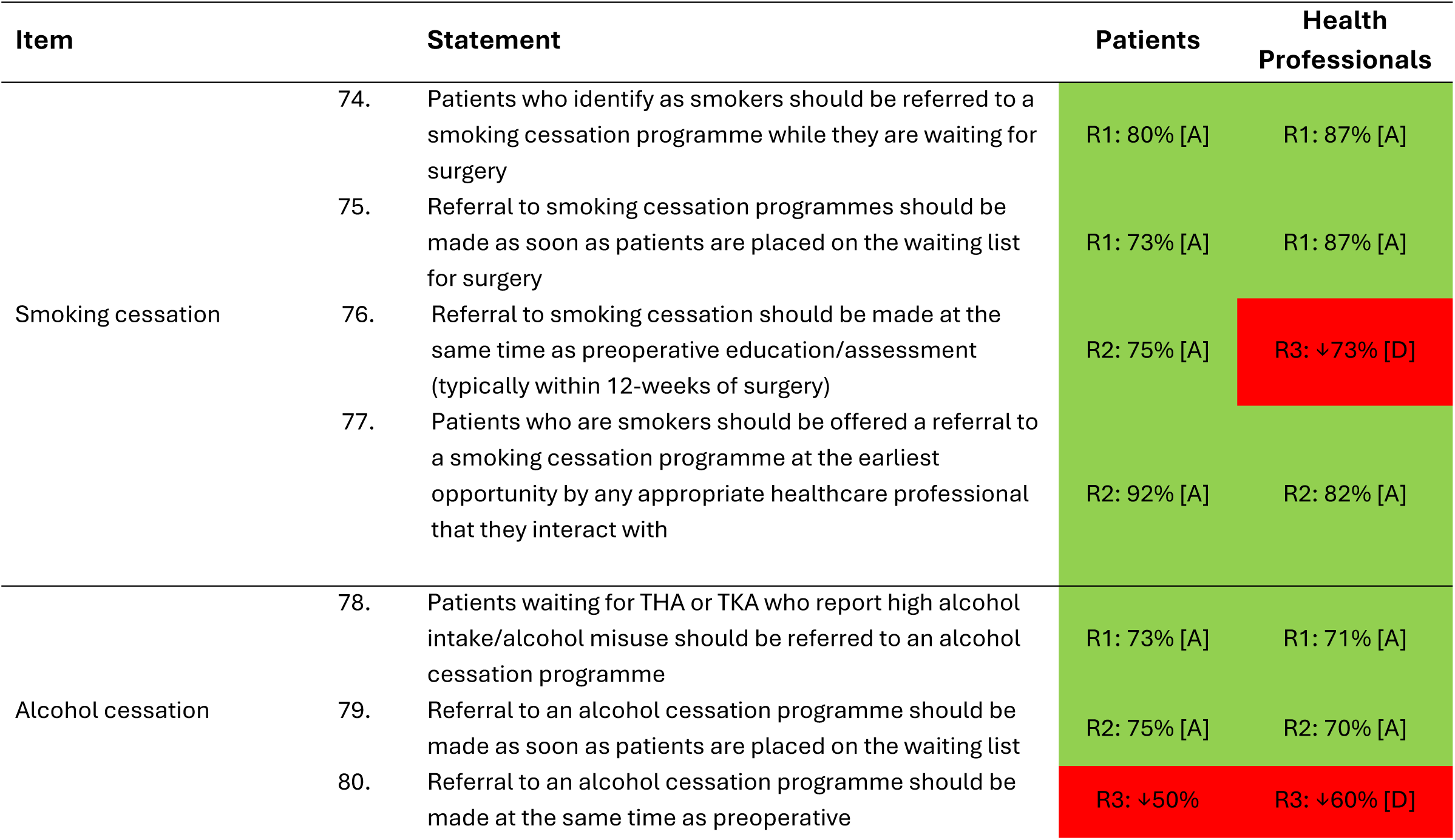
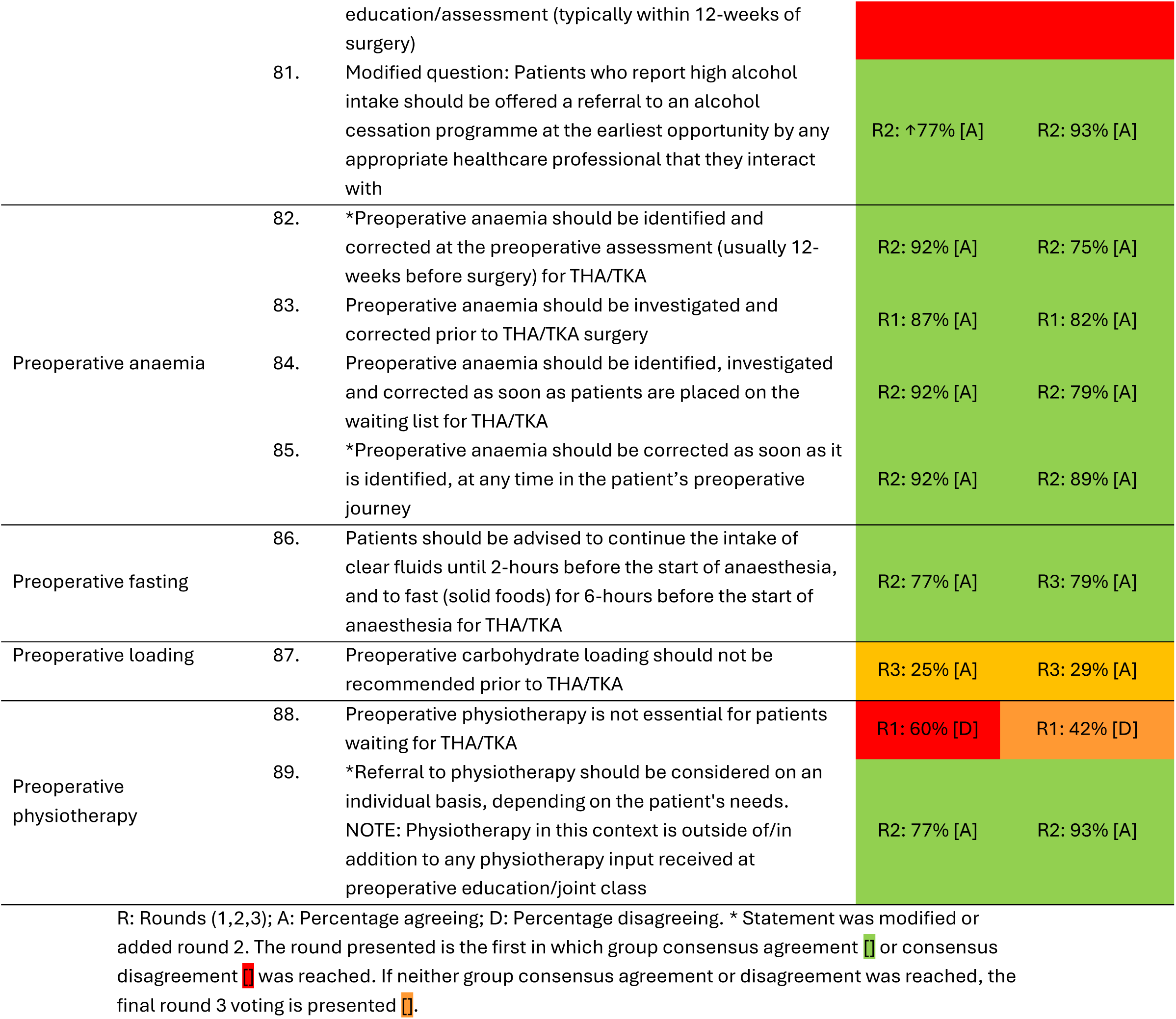
Summary of Delphi voting results from patients and health professionals on “patient optimisation” statements.

| Item | Statement | Patients | Health Professionals |
| --- | --- | --- | --- |
| Smoking cessation | 74. Patients who identify as smokers should be referred to a smoking cessation programme while they are waiting for surgery | R1: 80% [A] | R1: 87% [A] |
|  | 75. Referral to smoking cessation programmes should be made as soon as patients are placed on the waiting list for surgery | R1: 73% [A] | R1: 87% [A] |
|  | 76. Referral to smoking cessation should be made at the same time as preoperative education/assessment (typically within 12-weeks of surgery) | R2: 75% [A] | R3: ↓73% [D] |
|  | 77. Patients who are smokers should be offered a referral to a smoking cessation programme at the earliest opportunity by any appropriate healthcare professional that they interact with | R2: 92% [A] | R2: 82% [A] |
| Alcohol cessation | 78. Patients waiting for THA or TKA who report high alcohol intake/alcohol misuse should be referred to an alcohol cessation programme | R1: 73% [A] | R1: 71% [A] |
|  | 79. Referral to an alcohol cessation programme should be made as soon as patients are placed on the waiting list | R2: 75% [A] | R2: 70% [A] |
|  | 80. Referral to an alcohol cessation programme should be made at the same time as preoperative | R3: ↓50% | R3: ↓60% [D] |

|  |  |  |  |  |
| --- | --- | --- | --- | --- |
|  |  | education/assessment (typically within 12-weeks of surgery) |  |  |
|  | 81. | Modified question: Patients who report high alcohol intake should be offered a referral to an alcohol cessation programme at the earliest opportunity by any appropriate healthcare professional that they interact with | R2: ↑77% [A] | R2: 93% [A] |
| Preoperative anaemia | 82. | *Preoperative anaemia should be identified and corrected at the preoperative assessment (usually 12-weeks before surgery) for THA/TKA | R2: 92% [A] | R2: 75% [A] |
|  | 83. | Preoperative anaemia should be investigated and corrected prior to THA/TKA surgery | R1: 87% [A] | R1: 82% [A] |
|  | 84. | Preoperative anaemia should be identified, investigated and corrected as soon as patients are placed on the waiting list for THA/TKA | R2: 92% [A] | R2: 79% [A] |
|  | 85. | *Preoperative anaemia should be corrected as soon as it is identified, at any time in the patient's preoperative journey | R2: 92% [A] | R2: 89% [A] |
| Preoperative fasting | 86. | Patients should be advised to continue the intake of clear fluids until 2-hours before the start of anaesthesia, and to fast (solid foods) for 6-hours before the start of anaesthesia for THA/TKA | R2: 77% [A] | R3: 79% [A] |
| Preoperative loading | 87. | Preoperative carbohydrate loading should not be recommended prior to THA/TKA | R3: 25% [A] | R3: 29% [A] |
| Preoperative physiotherapy | 88. | Preoperative physiotherapy is not essential for patients waiting for THA/TKA | R1: 60% [D] | R1: 42% [D] |
|  | 89. | *Referral to physiotherapy should be considered on an individual basis, depending on the patient's needs. NOTE: Physiotherapy in this context is outside of/in addition to any physiotherapy input received at preoperative education/joint class | R2: 77% [A] | R2: 93% [A] |

R: Rounds (1,2,3); A: Percentage agreeing; D: Percentage disagreeing. \* Statement was modified or added round 2. The round presented is the first in which group consensus agreement or consensus disagreement was reached. If neither group consensus agreement or disagreement was reached, the final round 3 voting is presented .

In round two, 11 statements (including four new or modified statements) were included, with seven reaching consensus in both groups. In round one, both groups disagreed with the statement that preoperative physiotherapy is not essential for patients waiting for THA/TKA (item 88) and therefore a new statement was introduced (Referral to physiotherapy should be considered on an individual basis, depending on the patient’s needs: Item 89). This statement subsequently reached consensus in both groups.

Two statements relating to smoking cessation referrals within 12-weeks of surgery (item 76), and patients being advised to continue intake of clear fluids until two hours before anaesthesia (Item 86) achieved consensus among patients but not professionals.

Four statements were included in round three, with one reaching consensus (Table 3). All panellists reconsidered two statements that had not achieved consensus in earlier rounds (Items 80, 87). Additionally, professionals evaluated two statements that had previously reached consensus by patients in round two (Items 76, 86). Neither group achieved consensus on statements related to referring patients to an alcohol cessation programme within 12-weeks of surgery (Item 80) and recommending preoperative carbohydrate loading prior to THA/TKA (Item 87). Most professionals disagreed with referring patients to smoking cessation within 12-weeks of surgery (Item 76). There was, however, consensus among both groups that referrals to either alcohol or smoking cessation programmes should be offered at the earliest opportunity and should be reassessed at each point along the pathway, rather than only within 12-weeks of surgery.

### Other interventions to support people waiting (Table 6)

Twelve statements were included in round one, with five reaching consensus in both groups (Table 3). Two statements reached consensus for patients in round one, but not for professionals: prehabilitation being offered with twelve weeks of surgery (Item 91) and patients with a body mass index (BMI) ≥27 kg/m² being offered a referral to a weight management programme (Item 102).

**Table 6.**
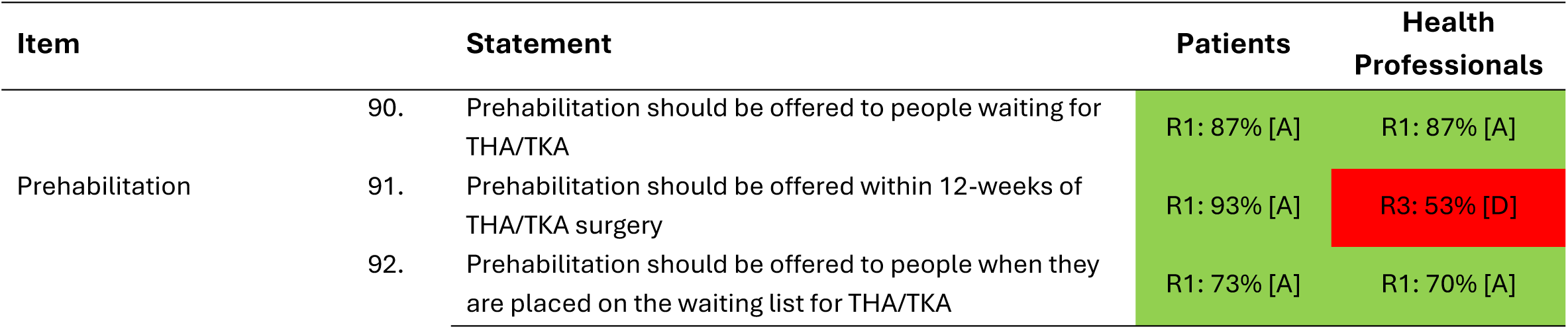
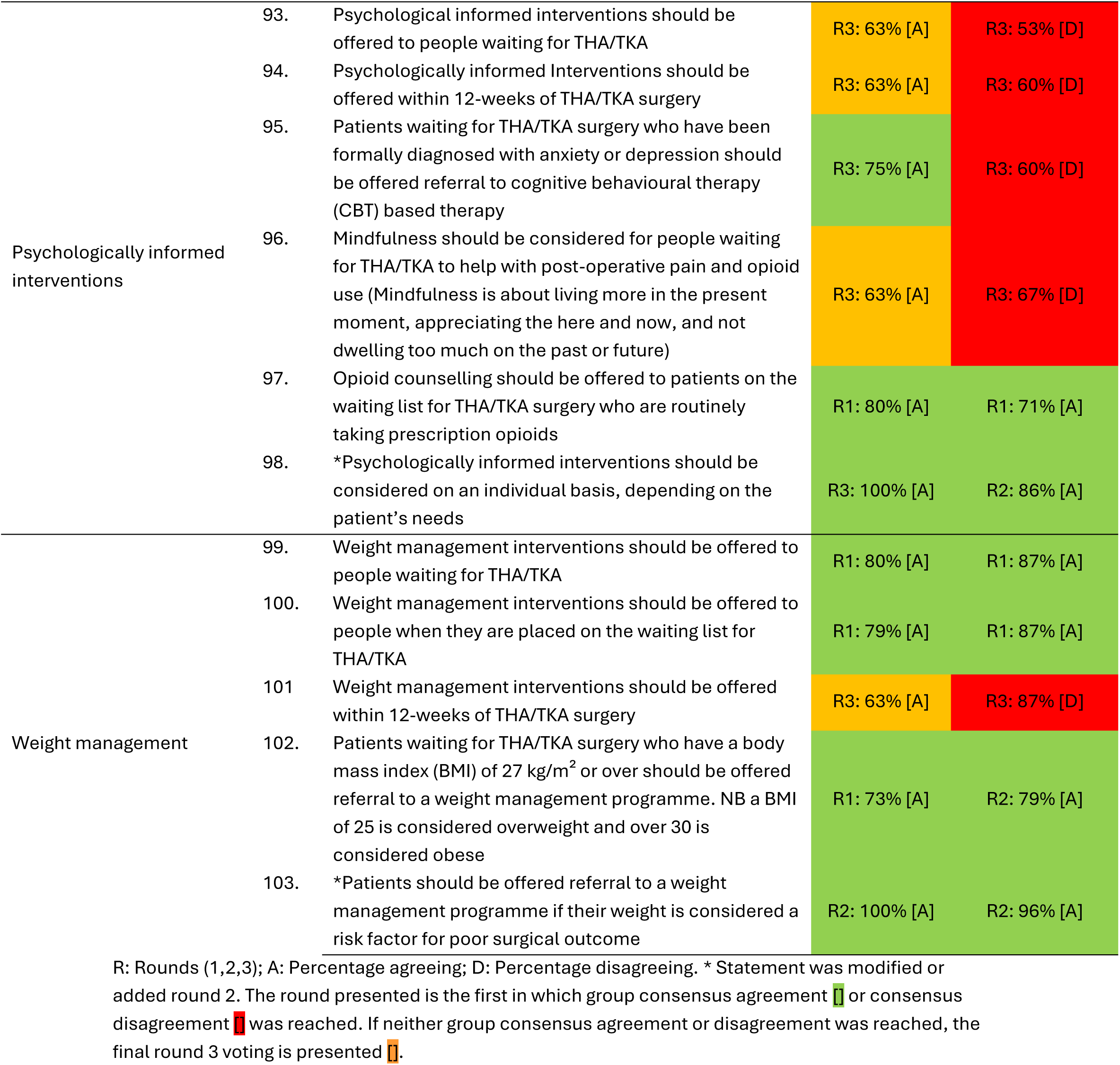
Summary of Delphi voting results from patients and health professionals on “other interventions to support people waiting” statements.

| Item | Statement | Patients | Health Professionals |
| --- | --- | --- | --- |
| Prehabilitation | 90. Prehabilitation should be offered to people waiting for THA/TKA | R1: 87% [A] | R1: 87% [A] |
|  | 91. Prehabilitation should be offered within 12-weeks of THA/TKA surgery | R1: 93% [A] | R3: 53% [D] |
|  | 92. Prehabilitation should be offered to people when they are placed on the waiting list for THA/TKA | R1: 73% [A] | R1: 70% [A] |

|  |  |  |  |  |
| --- | --- | --- | --- | --- |
| Psychologically informed interventions | 93. | Psychological informed interventions should be offered to people waiting for THA/TKA | R3: 63% [A] | R3: 53% [D] |
|  | 94. | Psychologically informed Interventions should be offered within 12-weeks of THA/TKA surgery | R3: 63% [A] | R3: 60% [D] |
|  | 95. | Patients waiting for THA/TKA surgery who have been formally diagnosed with anxiety or depression should be offered referral to cognitive behavioural therapy (CBT) based therapy | R3: 75% [A] | R3: 60% [D] |
|  | 96. | Mindfulness should be considered for people waiting for THA/TKA to help with post-operative pain and opioid use (Mindfulness is about living more in the present moment, appreciating the here and now, and not dwelling too much on the past or future) | R3: 63% [A] | R3: 67% [D] |
|  | 97. | Opioid counselling should be offered to patients on the waiting list for THA/TKA surgery who are routinely taking prescription opioids | R1: 80% [A] | R1: 71% [A] |
|  | 98. | *Psychologically informed interventions should be considered on an individual basis, depending on the patient's needs | R3: 100% [A] | R2: 86% [A] |
| Weight management | 99. | Weight management interventions should be offered to people waiting for THA/TKA | R1: 80% [A] | R1: 87% [A] |
|  | 100. | Weight management interventions should be offered to people when they are placed on the waiting list for THA/TKA | R1: 79% [A] | R1: 87% [A] |
|  | 101. | Weight management interventions should be offered within 12-weeks of THA/TKA surgery | R3: 63% [A] | R3: 87% [D] |
|  | 102. | Patients waiting for THA/TKA surgery who have a body mass index (BMI) of 27 kg/m<sup>2</sup> or over should be offered referral to a weight management programme. NB a BMI of 25 is considered overweight and over 30 is considered obese | R1: 73% [A] | R2: 79% [A] |
|  | 103. | *Patients should be offered referral to a weight management programme if their weight is considered a risk factor for poor surgical outcome | R2: 100% [A] | R2: 96% [A] |

R: Rounds (1,2,3); A: Percentage agreeing; D: Percentage disagreeing. \* Statement was modified or added round 2. The round presented is the first in which group consensus agreement or consensus disagreement was reached. If neither group consensus agreement or disagreement was reached, the final round 3 voting is presented .

In round two, nine statements were included (including two new or modified statements), with three reaching consensus by both groups. Professionals achieved consensus on referring patients with a BMI of ≥27 kg/m² to a weight management programme (item 102). Both groups also agreed that patients should be referred to a weight management programme if their BMI was considered a risk factor for poor surgical outcome (item 103). However, only professionals agreed that psychologically informed interventions should be considered on an individual basis, depending on the patient’s needs (Item 98).

Seven statements were included in round three, with only one statement reaching consensus. Both groups reconsidered five statements that had not achieved consensus in earlier rounds (Items 93-96, 101). Additionally, patients reconsidered one statement (Item 98), that had had achieved consensus among professionals in round two, while professionals reconsidered one statement that had achieved consensus by patients in round one (Item 91) Neither group agreed with referring patients to weight management or alcohol and smoking cessation programmes within 12-weeks of surgery. The interventions themselves were not questioned; rather, the timeframe, with 12-weeks from surgery being considered too late.

Consensus was not reached in either group for: offering psychological interventions to people waiting for THA/TKA (item 93), offering psychological interventions within 12-weeks of surgery (item 94), and considering mindfulness for post-operative pain and opioid use (Item 96). However, patients reached consensus that psychologically informed interventions should be considered on an individual basis, depending on the patient’s needs (Item 98). Patients also reached consensus on offering patients formally diagnosed with anxiety or depression a referral to cognitive behavioural therapy (CBT) (Item 95), but this did not achieve consensus among professionals. Whilst all panellists discussed the importance of psychological interventions, professionals highlighted service constraints such as large waiting lists for mental health services. They also noted time limitations related to writing referrals and ensuring patients are signposted to appropriate services.

### Waiting a long time for surgery (table 7)

Twenty statements relating to waiting a long time for surgery were included and all reached consensus: 15 in round one, and four more in round two (Table 3). The final statement relating to signposting people waiting for THA/TKA surgery to third sector organisations reached consensus by professionals in round one, and patients in round three (item 123).

**Table 7.**
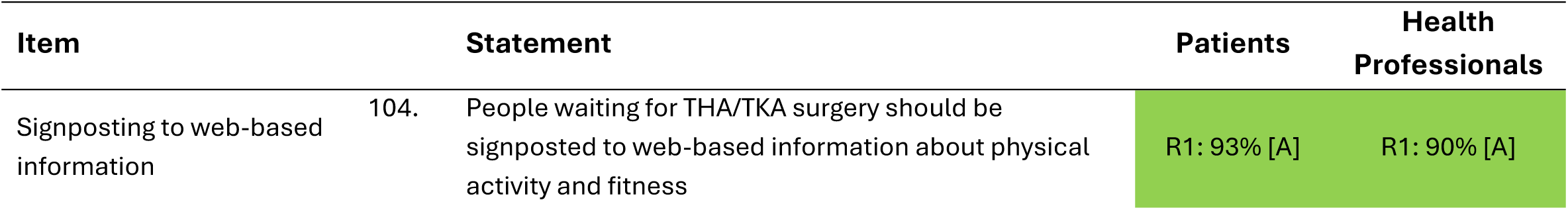
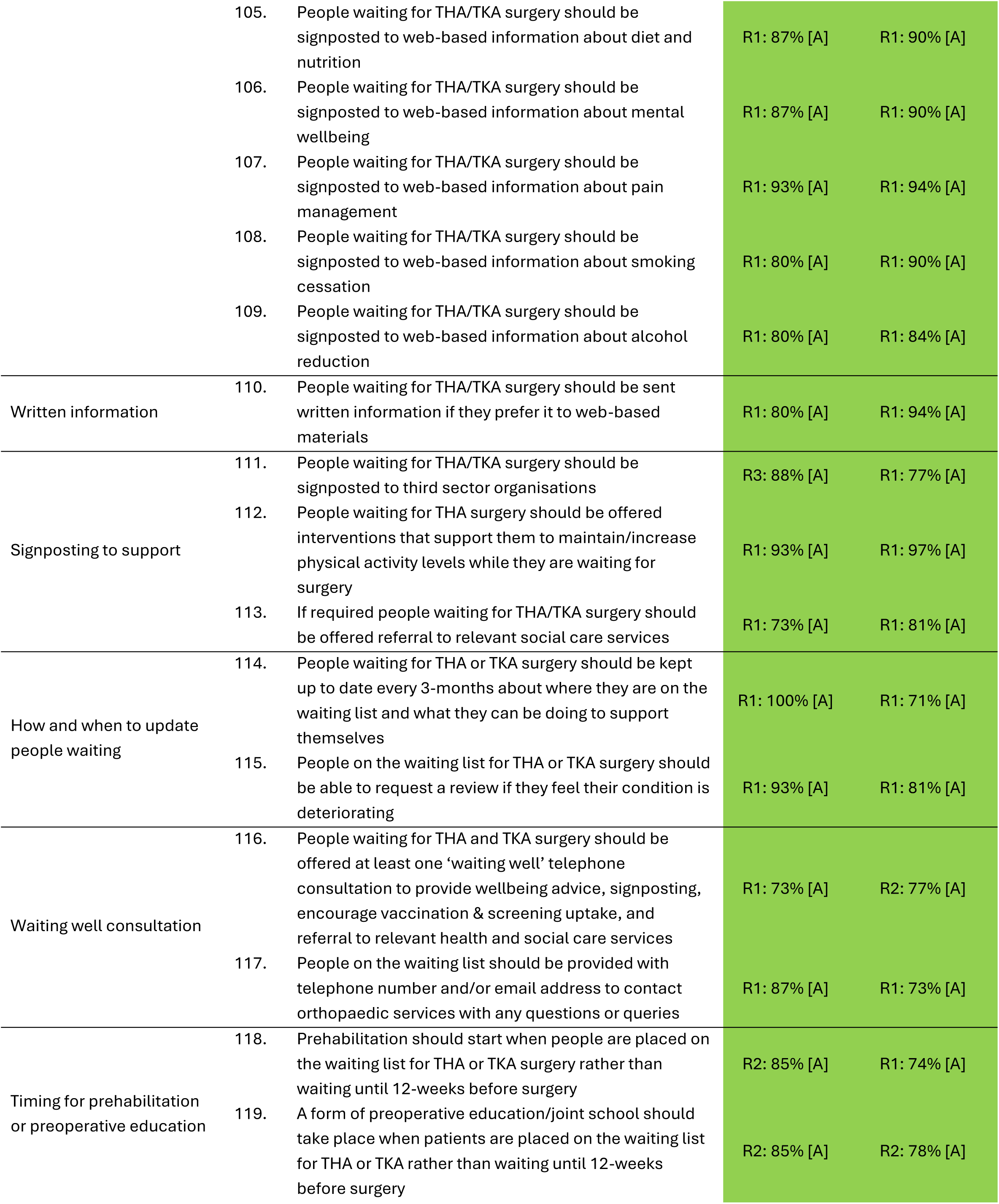
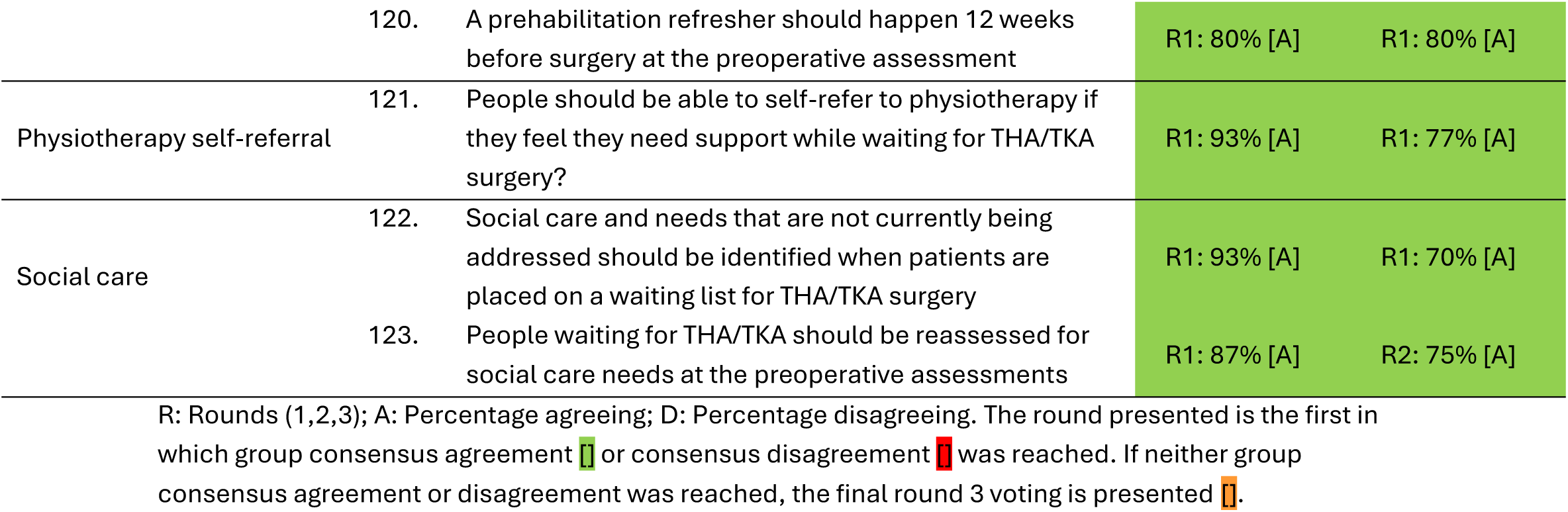
Summary of Delphi voting results from patients and health professionals on “waiting a long time for surgery” statements.

| Item | Statement | Patients | Health Professionals |
| --- | --- | --- | --- |
| Signposting to web-based information | 104. People waiting for THA/TKA surgery should be signposted to web-based information about physical activity and fitness | R1: 93% [A] | R1: 90% [A] |

|  |  |  |  |  |
| --- | --- | --- | --- | --- |
|  | 105. | People waiting for THA/TKA surgery should be signposted to web-based information about diet and nutrition | R1: 87% [A] | R1: 90% [A] |
|  | 106. | People waiting for THA/TKA surgery should be signposted to web-based information about mental wellbeing | R1: 87% [A] | R1: 90% [A] |
|  | 107. | People waiting for THA/TKA surgery should be signposted to web-based information about pain management | R1: 93% [A] | R1: 94% [A] |
|  | 108. | People waiting for THA/TKA surgery should be signposted to web-based information about smoking cessation | R1: 80% [A] | R1: 90% [A] |
|  | 109. | People waiting for THA/TKA surgery should be signposted to web-based information about alcohol reduction | R1: 80% [A] | R1: 84% [A] |
| Written information | 110. | People waiting for THA/TKA surgery should be sent written information if they prefer it to web-based materials | R1: 80% [A] | R1: 94% [A] |
| Signposting to support | 111. | People waiting for THA/TKA surgery should be signposted to third sector organisations | R3: 88% [A] | R1: 77% [A] |
|  | 112. | People waiting for THA surgery should be offered interventions that support them to maintain/increase physical activity levels while they are waiting for surgery | R1: 93% [A] | R1: 97% [A] |
|  | 113. | If required people waiting for THA/TKA surgery should be offered referral to relevant social care services | R1: 73% [A] | R1: 81% [A] |
| How and when to update people waiting | 114. | People waiting for THA or TKA surgery should be kept up to date every 3-months about where they are on the waiting list and what they can be doing to support themselves | R1: 100% [A] | R1: 71% [A] |
|  | 115. | People on the waiting list for THA or TKA surgery should be able to request a review if they feel their condition is deteriorating | R1: 93% [A] | R1: 81% [A] |
| Waiting well consultation | 116. | People waiting for THA and TKA surgery should be offered at least one 'waiting well' telephone consultation to provide wellbeing advice, signposting, encourage vaccination & screening uptake, and referral to relevant health and social care services | R1: 73% [A] | R2: 77% [A] |
|  | 117. | People on the waiting list should be provided with telephone number and/or email address to contact orthopaedic services with any questions or queries | R1: 87% [A] | R1: 73% [A] |
| Timing for prehabilitation or preoperative education | 118. | Prehabilitation should start when people are placed on the waiting list for THA or TKA surgery rather than waiting until 12-weeks before surgery | R2: 85% [A] | R1: 74% [A] |
|  | 119. | A form of preoperative education/joint school should take place when patients are placed on the waiting list for THA or TKA rather than waiting until 12-weeks before surgery | R2: 85% [A] | R2: 78% [A] |
|  | 120. | A prehabilitation refresher should happen 12 weeks before surgery at the preoperative assessment | R1: 80% [A] | R1: 80% [A] |
| Physiotherapy self-referral | 121. | People should be able to self-refer to physiotherapy if they feel they need support while waiting for THA/TKA surgery? | R1: 93% [A] | R1: 77% [A] |
| Social care | 122. | Social care and needs that are not currently being addressed should be identified when patients are placed on a waiting list for THA/TKA surgery | R1: 93% [A] | R1: 70% [A] |
|  | 123. | People waiting for THA/TKA should be reassessed for social care needs at the preoperative assessments | R1: 87% [A] | R2: 75% [A] |

R: Rounds (1,2,3); A: Percentage agreeing; D: Percentage disagreeing. The round presented is the first in which group consensus agreement or consensus disagreement was reached. If neither group consensus agreement or disagreement was reached, the final round 3 voting is presented .

In summary, the panel in this three-round modified Delphi study reached consensus on 108 statements across preoperative education (n=68), patient optimisation (n=12), other interventions to support people waiting (n=8) and waiting a long time for surgery (n=20) (Appendix A).

## Discussion

This national modified Delphi study developed a comprehensive set of recommendations for interventions and support for people waiting for hip or knee replacement surgery. Of the 108 statements included in the final recommendations (Appendix A), most focused on preoperative patient education (68 statements, Table 4) followed by waiting a long time for surgery (20 statements; Table 7), patient optimisation (12 statements, Table 5) and other interventions (8 statements, Table 6). Of the ten modified or new statements introduced following feedback in round one, nine reached consensus. The addition of these statements allowed the perspectives of patients and professionals to be more fully reflected.

The current findings and recommendations align with Anderson et al. (2021) Delphi study on preoperative education and prehabilitation for people waiting for TKA highlighting the importance of preoperative education and weight management interventions. However, contrary to Anderson et al. (2021), the current Delphi study reached consensus with patients but not with professionals regarding CBT-based therapy. In addition, suggestions were made by both patients and professionals to modify the statements related to psychological interventions so that such approaches could be tailored to individual needs.

Although most statements achieved consensus, fifteen statements failed to meet the 70% threshold in both groups. These statements spanned preoperative education, patient optimisation and other interventions to support people waiting. There were varying levels of agreement between the groups, with patients achieving agreement in all items within preoperative education, and interventions to support people waiting a long time for surgery. Some statements received high agreement (≥70%) from patients but did not reach agreement by professionals, such as core control exercises, walking aid practice and training with steps being part of preoperative exercise. Additionally, all twenty statements within interventions to support people waiting a long time for surgery reached a high agreement (≥70%) amongst both panellist groups signifying universality of importance.

Notably, timing of interventions (delivery within 12-weeks of surgery), and psychological interventions were among those that generated the most uncertainty and varied opinions, indicating a need for further clarification around efficient delivery. Specifically, there was a need identified by all panellists to consider the timing of these interventions. Professionals highlighted the importance of introducing interventions earlier along the surgical pathway, especially those supporting behavioural change, which may have minimal effect if delivered too close to surgery (Burns et al., 2016; N. Singh, Stewart, & Benatar, 2019). As behavioural change takes time, the opportunity for patients to engage earlier and gradually adopt and maintain health behaviours (such as smoking cessation, weight reduction, and reduced alcohol consumption) prior to surgery was considered important. It was highlighted that a ‘drip feed’ approach, with information and support introduced from the point of being listed for surgery, would provide an opportunity for patients to adopt health behaviours to enhance engagement and optimise health. Grocott, Plumb, Edwards, Fecher-Jones, & Levett (2017) highlighted the importance of engaging patients in collaborative decision making at “the moment of contemplation of surgery”, where there is potential for patients to be guided towards successful behavioural change earlier in the pathway.

The panel’s opinion aligns with the transtheoretical model of change (Prochaska, DiClemente, & Norcross, 1992) which emphasises the importance of delivering interventions at the most appropriate time. It may therefore be useful to evaluate an individual’s stage to assess their readiness to change. Singh, Murphy, Maher, & Smith (2024) systematic review on habit formation also highlights that behavioural change varies considerably across individuals and can be influenced by factors such as frequency, timing, and the type of habit being formed. While evidence supports modifying behaviours prior to surgery for optimal benefit (Fong et al., 2023), this suggests that not all patients will be ready to engage in behavioural change interventions when offered 12-weeks before surgery.

Healthcare professionals also raised concerns about the feasibility of delivering interventions under current system pressures. Barriers around staff capacity and workload pressures highlighted the importance of exploring alternative methods of delivery such as signposting to digital technologies and community/third sector partners. This has been demonstrated using digital technology for smoking cessation and reducing alcohol intake (Tønnesen et al., 2023) (Doiron-Cadrin et al., 2020; Miller, Mohammadi, Watson, Crocker, & Westby, 2021; Straat et al., 2023) and prehabilitation. However, further investigation into digital technology for delivering effective interventions is required. Studies have described the strategies that have been used to operate when system pressures exceed capacity (Manning & Islam, 2023; Page, Irving, Amalberti, & Vincent, 2024). Further research is needed to identify efficient and effective ways for interventions to be integrated into existing pathways without placing strain on an already over-stretched system.

### Strengths and limitations

A key strength of this study was the robust use of a modified Delphi methodology and the development of round one statements based on extensive evidence review. The use of online surveys and Microsoft Teams to host asynchronous virtual workshops facilitated inclusion of panellists from across Scotland and researchers from the whole of UK, with online voting enabling anonymity. Involvement of all panellists to refine and develop statements and to review and comment on the development of the final recommendations ensured diverse opinions informed each step. In addition, the final recommendations were determined by a combination of expert consensus and a comprehensive evidence review.

This study also had limitations. Firstly, all statements were equally weighted, and no prioritisation process was conducted, which may need to be addressed prior to implementation. While online methods were utilised to maximise engagement with geographically dispersed individuals, some may not have had the opportunity to participate, and the recommendations may not be truly representative of the Scottish population waiting for THA or TKA. Some patient participants experienced technical limitations within Microsoft Teams and were unable to submit votes anonymously in the workshops, which may have affected their responses. In addition, participants were mainly female, with all patients (100%) and 90% of professionals identifying as White British. Whilst we acknowledge some potential panellists may have been excluded due to digital challenges, all patients had support from the study team to pilot accessing the survey and the workshops. Although we did not reach our target sample size for the patient group across all rounds, attempts were made to keep participants informed in between each round. Finally, panellists were given the opportunity to choose mixed workshops (both professionals and patients) or separate workshops (workshops specifically for professionals and patients respectively).

The patient panellists opted to have separate workshops and discussions between the two groups may have influenced the final recommendations in a different way.

### Recommendations for practice and future research

Expert panellists expressed concerns in relation to the timing and delivery of behavioural change interventions prior to surgery under service pressures. Further research is required to determine optimal delivery of appropriate and effective interventions that are timed appropriately for patients, and to evaluate how intervention implementation can be adapted to the Scottish context under existing resource and system pressures. Further research is also required to explore psychological interventions for this population. This Delphi study has generated findings that will inform the development of best practice guidance for supporting people waiting for hip or knee replacement surgery that can guide clinical practice and policy in Scotland and that may be applicable in other regions with similar services and structures.

### Conclusion

This modified Delphi study was informed by key stakeholders with expertise in supporting people waiting for hip or knee replacement surgery, alongside those with lived experience. The study identified a comprehensive set of 108 recommendations organised into four key domains which can be used as a foundation to guide clinical practice and the preoperative management of people while they wait for hip or knee replacement surgery.

## Author statement

**Kay Cooper:** Conceptualisation, Methodology, Formal Analysis, Investigation, Writing – Original Draft, Writing – Review and Editing, Supervision, Project administration, Funding acquisition. **Emma Stage:** Investigation, Data Curation, Formal Analysis, Writing - Original Draft, Writing – Review and Editing. **Erin Hart-Winks:** Investigation, Data Curation, Formal Analysis. **Paul Swinton:** Conceptualisation, Methodology, Data Curation, Formal Analysis, Visualisation, Writing – Review and Editing, Supervision. **Lyndsay Alexander:** Conceptualisation, Methodology, Formal Analysis, Investigation, Writing – Original Draft, Writing – Review and Editing, Supervision. **Jo Shim:** Conceptualisation, Methodology, Data Curation, Investigation, Formal Analysis, Visualisation. **Tom Herbert:** Formal Analysis. **Stephen Bridgman:** Conceptualisation, Methodology, Funding acquisition, Writing - Review and Editing.

## Supporting information

Supplementary File 3_Equator Checklist

Supplementary File_CREDES Checklist

## Data Availability

All data produced in the present study are available upon reasonable request to the authors

## Acknowledgements

The study was funded by a project grant from Public Health Scotland (PHS2023-24R012)

The study was approved by Robert Gordon University School of Health Sciences Research Ethics Committee (Ref: SHS 24/25) and the NHS Research Scotland Permissions Coordinating Centre (Ref: NRS24/344449).

Thank you to the members of our Project Advisory Committee for their expert guidance and project oversight.

## Supplementary Files

Supplementary file 1: Round 1 & 2 Surveys and Round 3 Workshop slides Supplementary file 2: Descriptive comparison

Supplementary file 3: Equator checklist Supplementary file 4: CREDES checklist

## Appendix A. Final 108 statements reaching consensus

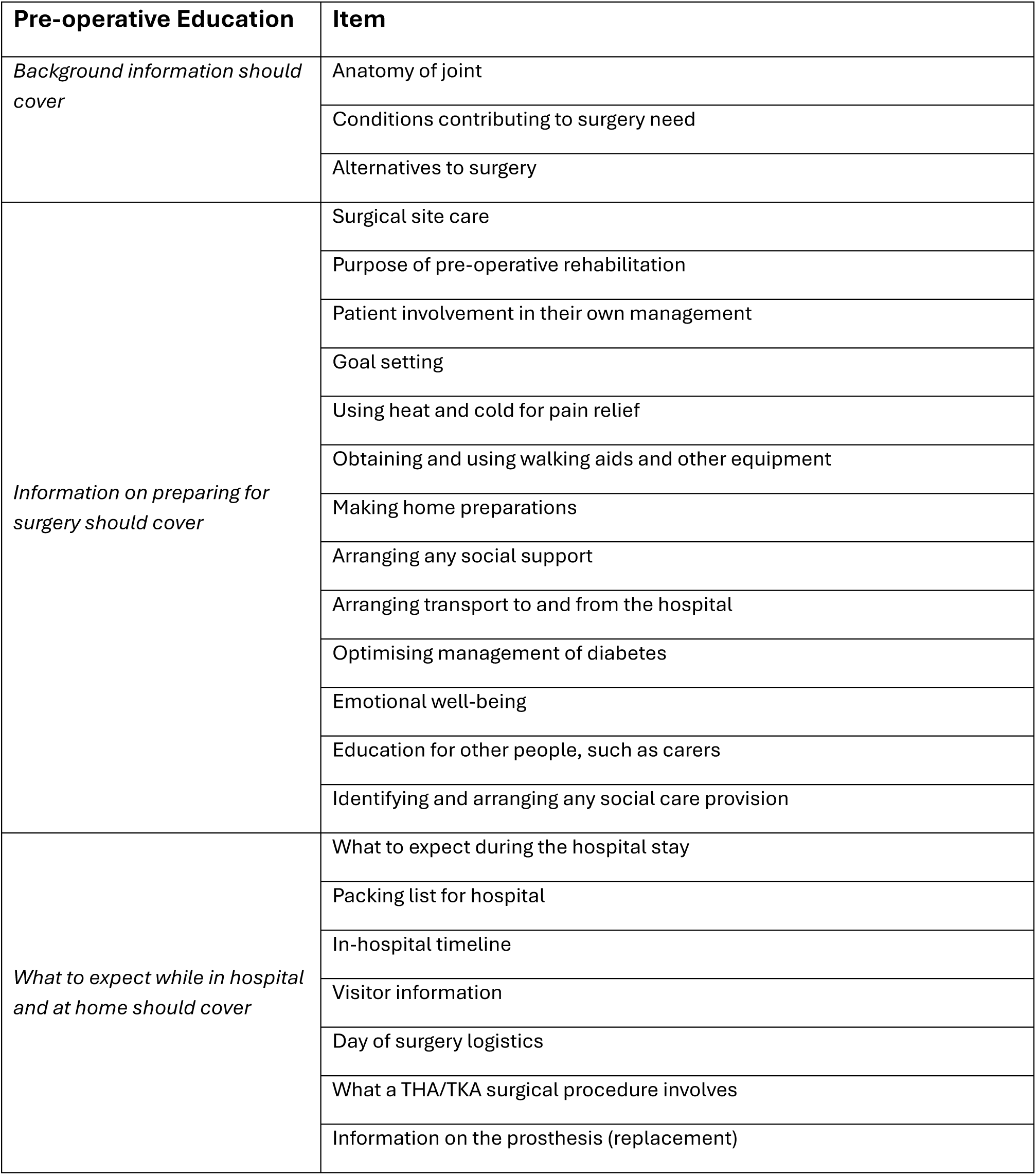
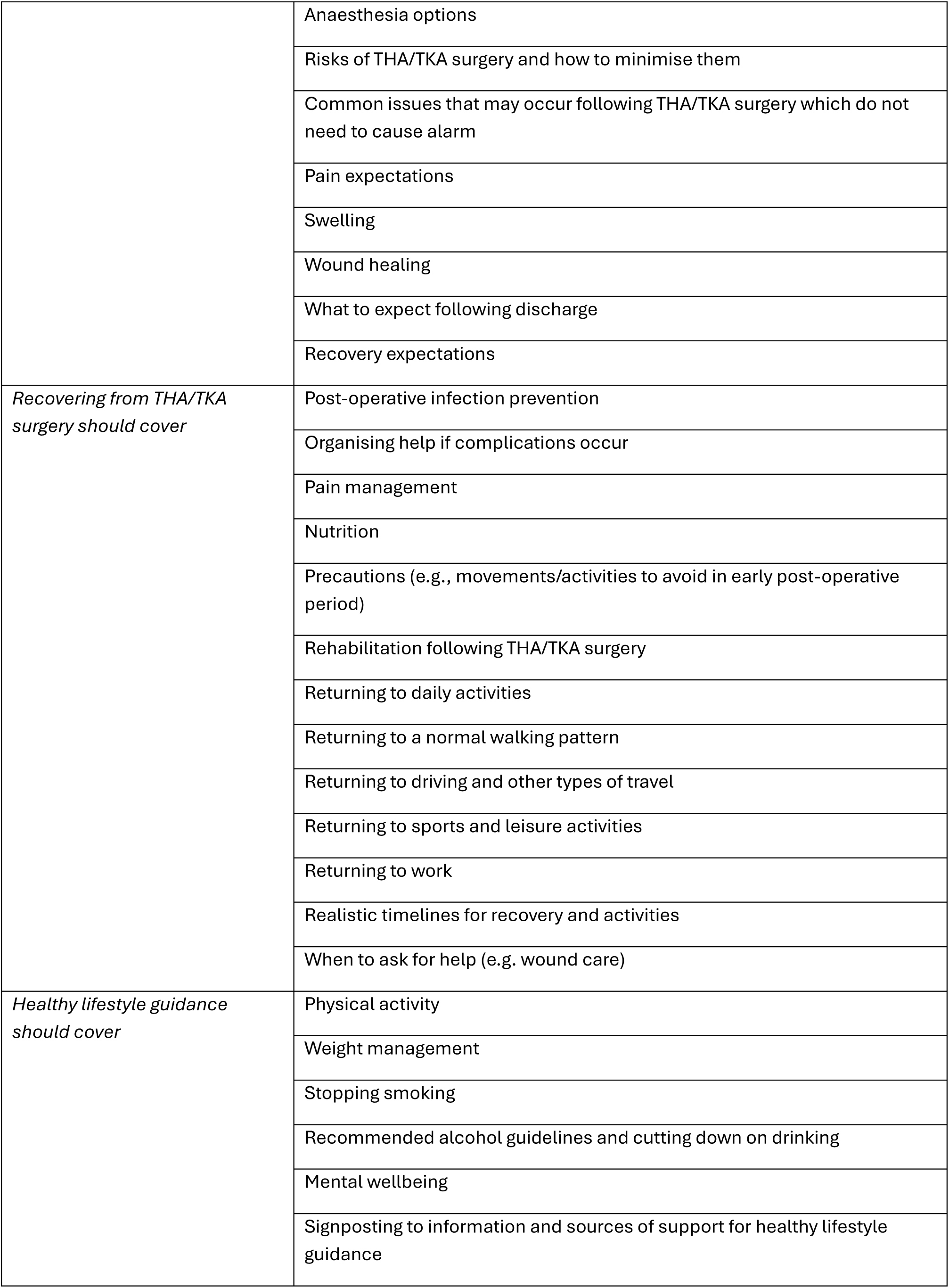
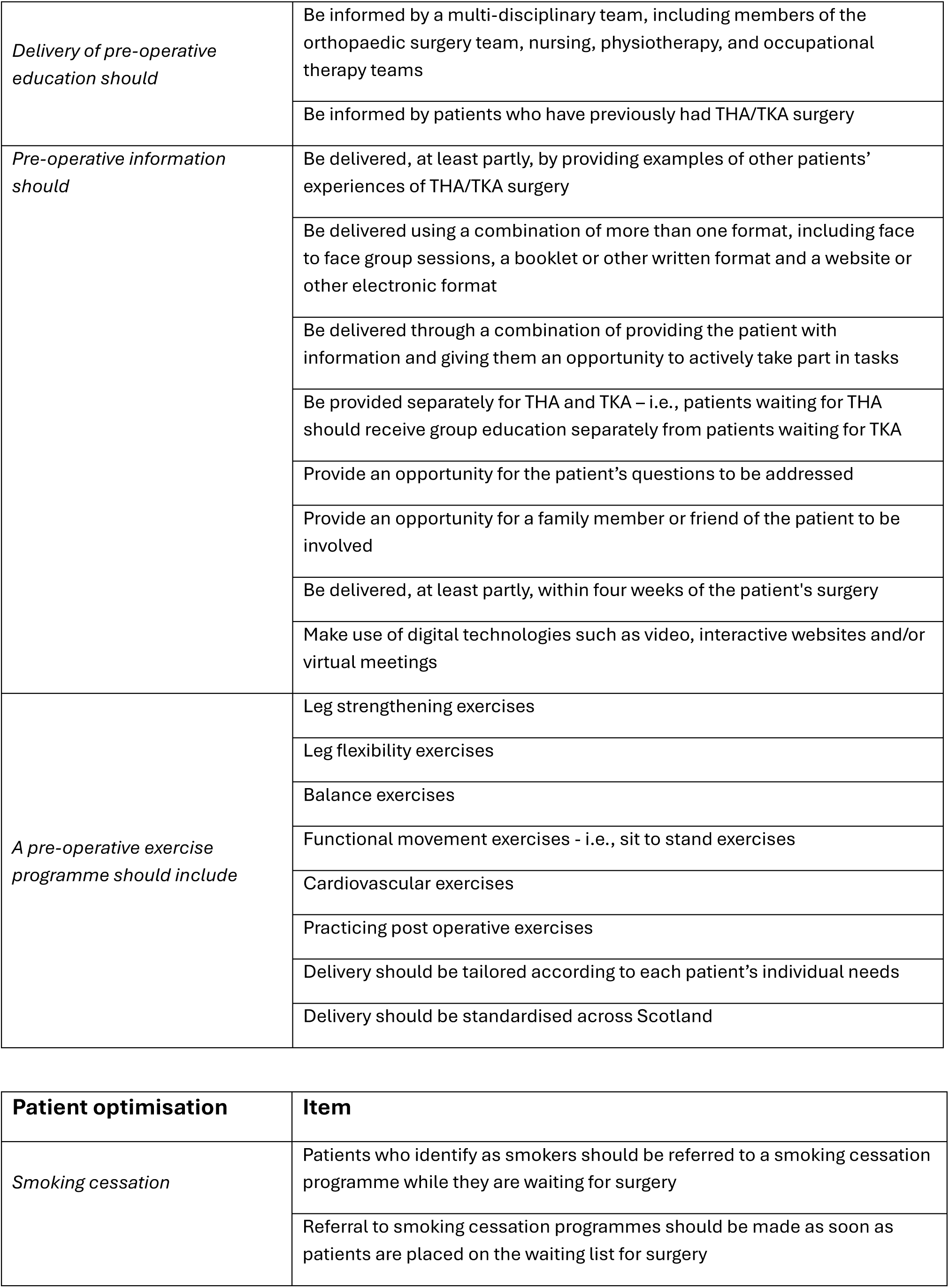
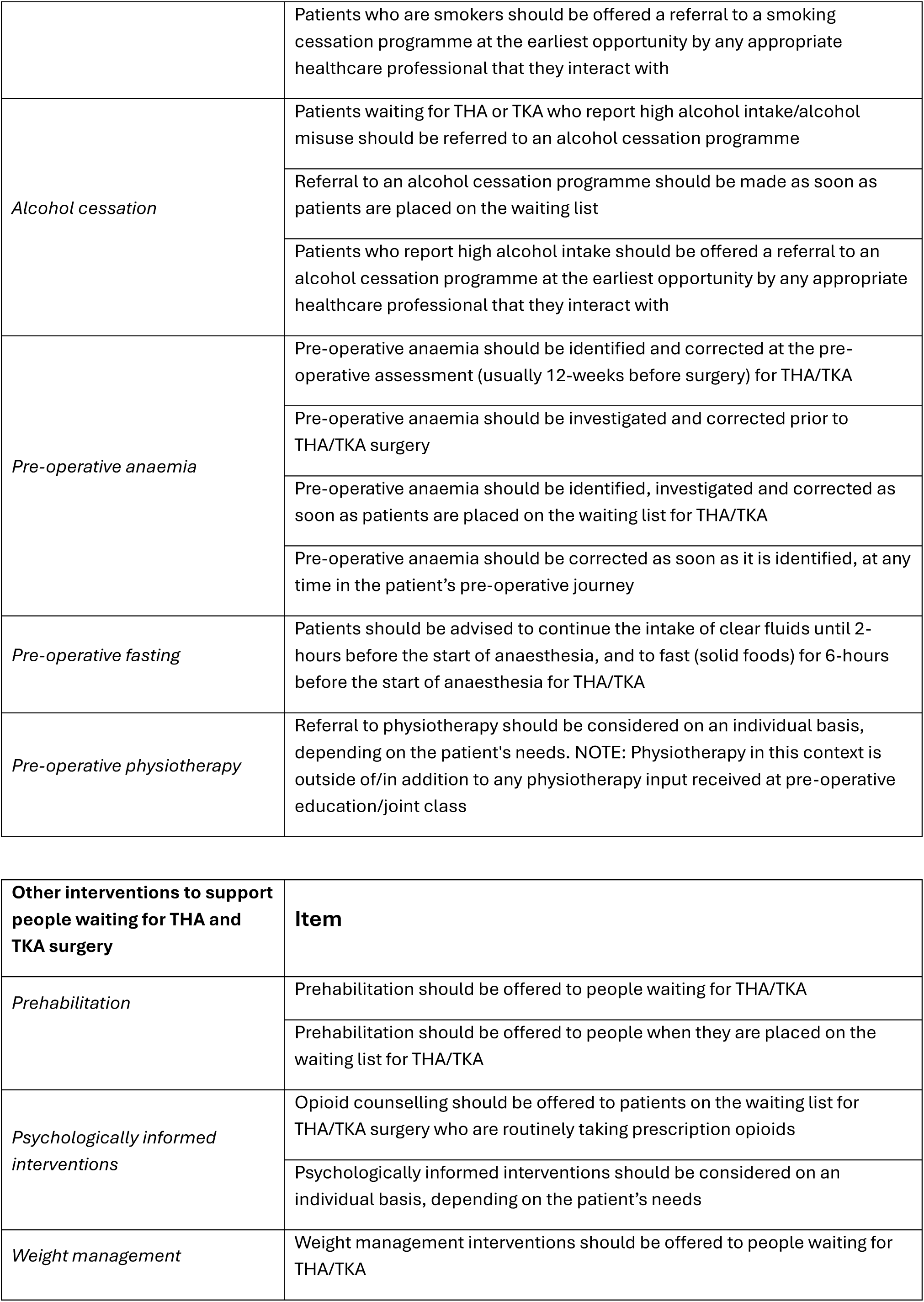
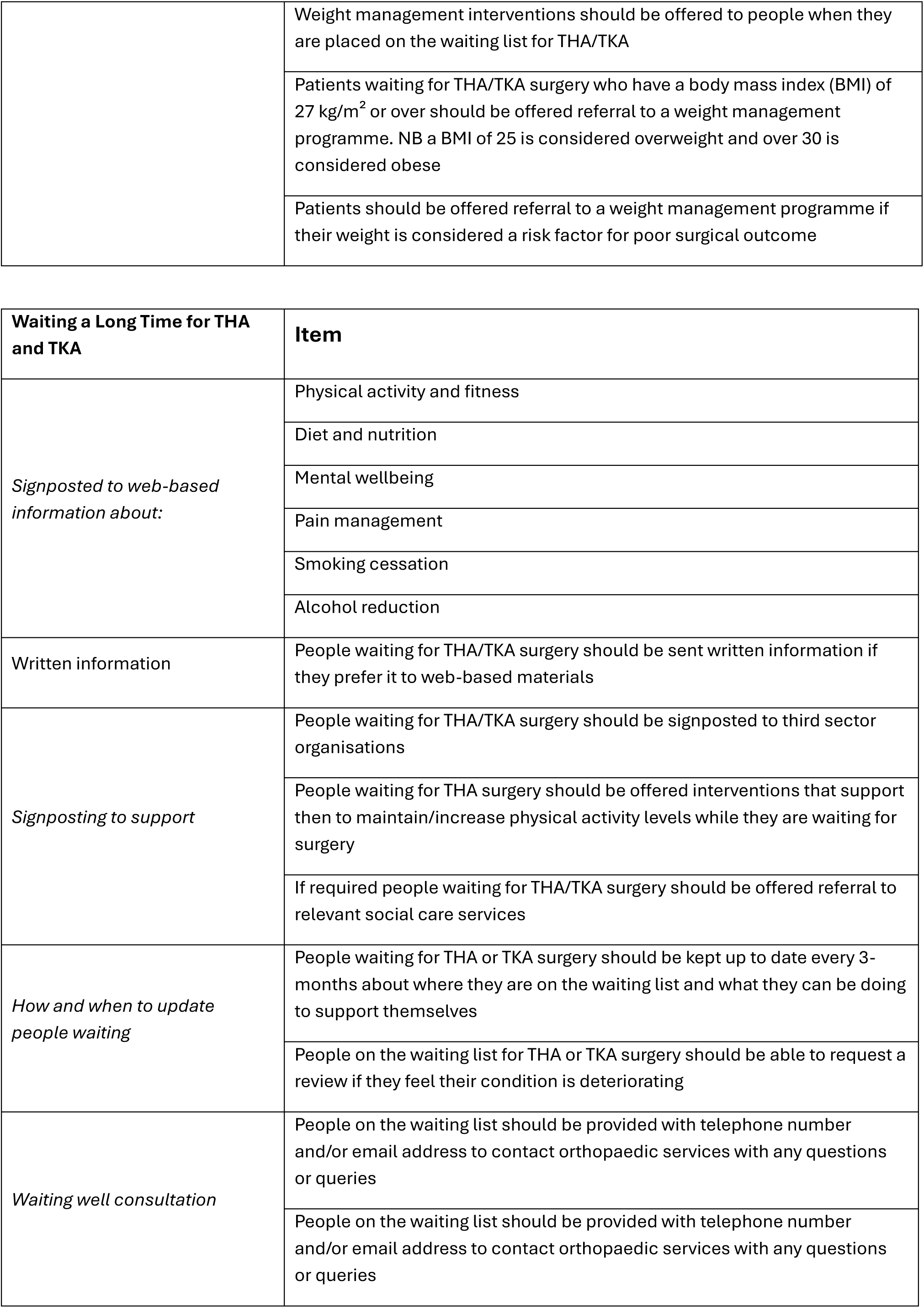
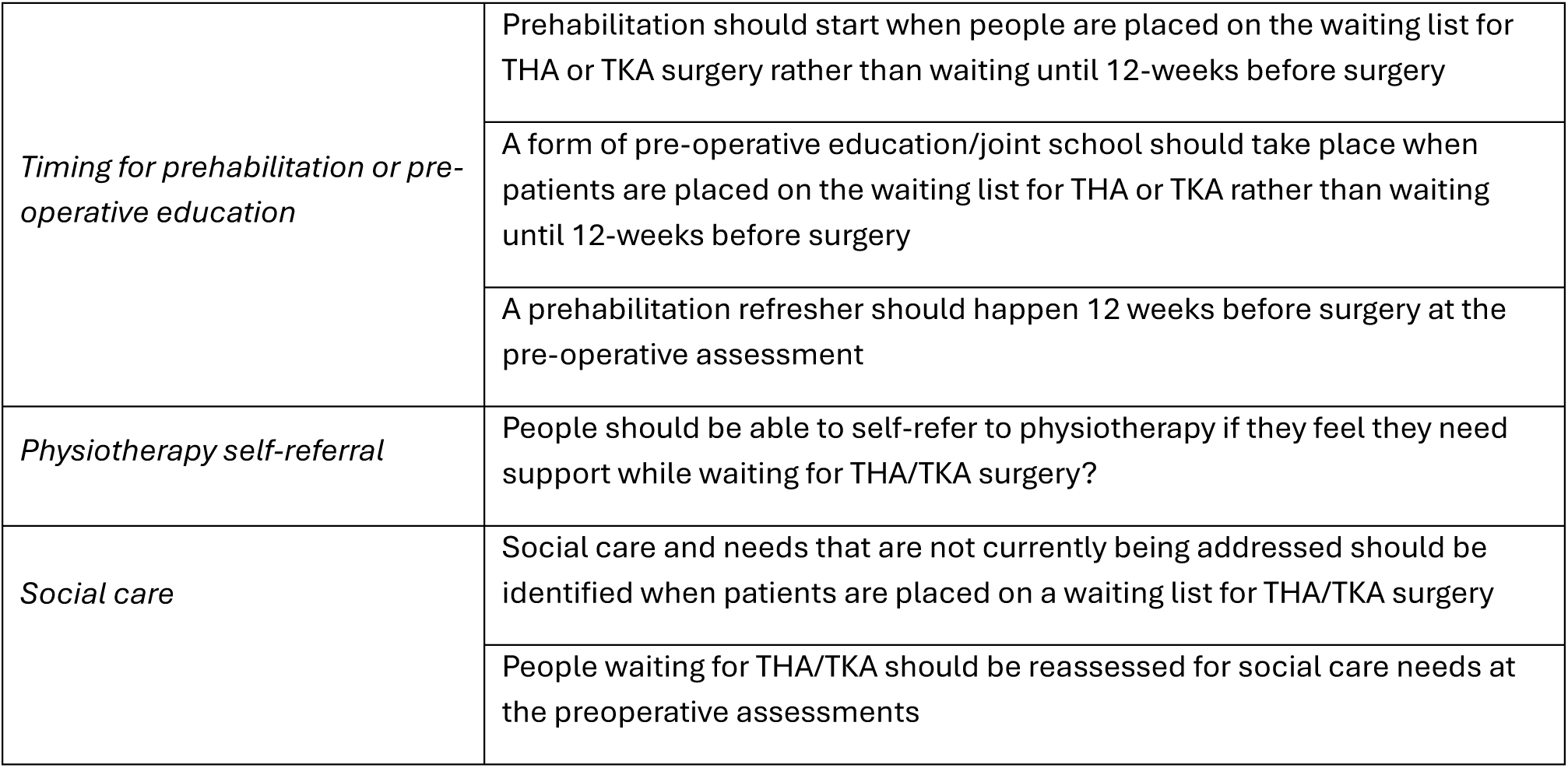

| Pre-operative Education | Item |
| --- | --- |
| *Background information should cover* | Anatomy of joint |
|  | Conditions contributing to surgery need |
|  | Alternatives to surgery |
| *Information on preparing for surgery should cover* | Surgical site care |
|  | Purpose of pre-operative rehabilitation |
|  | Patient involvement in their own management |
|  | Goal setting |
|  | Using heat and cold for pain relief |
|  | Obtaining and using walking aids and other equipment |
|  | Making home preparations |
|  | Arranging any social support |
|  | Arranging transport to and from the hospital |
|  | Optimising management of diabetes |
|  | Emotional well-being |
|  | Education for other people, such as carers |
|  | Identifying and arranging any social care provision |
| *What to expect while in hospital and at home should cover* | What to expect during the hospital stay |
|  | Packing list for hospital |
|  | In-hospital timeline |
|  | Visitor information |
|  | Day of surgery logistics |
|  | What a THA/TKA surgical procedure involves |
|  | Information on the prosthesis (replacement) |
|  | Anaesthesia options |
|  | Risks of THA/TKA surgery and how to minimise them |
|  | Common issues that may occur following THA/TKA surgery which do not need to cause alarm |
|  | Pain expectations |
|  | Swelling |
|  | Wound healing |
|  | What to expect following discharge |
|  | Recovery expectations |
| *Recovering from THA/TKA surgery should cover* | Post-operative infection prevention |
|  | Organising help if complications occur |
|  | Pain management |
|  | Nutrition |
|  | Precautions (e.g., movements/activities to avoid in early post-operative period) |
|  | Rehabilitation following THA/TKA surgery |
|  | Returning to daily activities |
|  | Returning to a normal walking pattern |
|  | Returning to driving and other types of travel |
|  | Returning to sports and leisure activities |
|  | Returning to work |
|  | Realistic timelines for recovery and activities |
|  | When to ask for help (e.g. wound care) |
| *Healthy lifestyle guidance should cover* | Physical activity |
|  | Weight management |
|  | Stopping smoking |
|  | Recommended alcohol guidelines and cutting down on drinking |
|  | Mental wellbeing |
|  | Signposting to information and sources of support for healthy lifestyle guidance |
| *Delivery of pre-operative education should* | Be informed by a multi-disciplinary team, including members of the orthopaedic surgery team, nursing, physiotherapy, and occupational therapy teams |
|  | Be informed by patients who have previously had THA/TKA surgery |
| *Pre-operative information should* | Be delivered, at least partly, by providing examples of other patients' experiences of THA/TKA surgery |
|  | Be delivered using a combination of more than one format, including face to face group sessions, a booklet or other written format and a website or other electronic format |
|  | Be delivered through a combination of providing the patient with information and giving them an opportunity to actively take part in tasks |
|  | Be provided separately for THA and TKA – i.e., patients waiting for THA should receive group education separately from patients waiting for TKA |
|  | Provide an opportunity for the patient's questions to be addressed |
|  | Provide an opportunity for a family member or friend of the patient to be involved |
|  | Be delivered, at least partly, within four weeks of the patient's surgery |
|  | Make use of digital technologies such as video, interactive websites and/or virtual meetings |
| *A pre-operative exercise programme should include* | Leg strengthening exercises |
|  | Leg flexibility exercises |
|  | Balance exercises |
|  | Functional movement exercises - i.e., sit to stand exercises |
|  | Cardiovascular exercises |
|  | Practicing post operative exercises |
|  | Delivery should be tailored according to each patient's individual needs |
|  | Delivery should be standardised across Scotland |

| **Other interventions to support people waiting for THA and TKA surgery** | **Item** |
| --- | --- |
| *Prehabilitation* | Prehabilitation should be offered to people waiting for THA/TKA |
|  | Prehabilitation should be offered to people when they are placed on the waiting list for THA/TKA |
| *Psychologically informed interventions* | Opioid counselling should be offered to patients on the waiting list for THA/TKA surgery who are routinely taking prescription opioids |
|  | Psychologically informed interventions should be considered on an individual basis, depending on the patient's needs |
| *Weight management* | Weight management interventions should be offered to people waiting for THA/TKA |
|  | Weight management interventions should be offered to people when they are placed on the waiting list for THA/TKA |
|  | Patients waiting for THA/TKA surgery who have a body mass index (BMI) of 27 kg/m<sup>2</sup> or over should be offered referral to a weight management programme. NB a BMI of 25 is considered overweight and over 30 is considered obese |
|  | Patients should be offered referral to a weight management programme if their weight is considered a risk factor for poor surgical outcome |

| **Waiting a Long Time for THA and TKA** | **Item** |
| --- | --- |
| *Signposted to web-based information about:* | Physical activity and fitness |
|  | Diet and nutrition |
|  | Mental wellbeing |
|  | Pain management |
|  | Smoking cessation |
|  | Alcohol reduction |
| Written information | People waiting for THA/TKA surgery should be sent written information if they prefer it to web-based materials |
| *Signposting to support* | People waiting for THA/TKA surgery should be signposted to third sector organisations |
|  | People waiting for THA surgery should be offered interventions that support them to maintain/increase physical activity levels while they are waiting for surgery |
|  | If required people waiting for THA/TKA surgery should be offered referral to relevant social care services |
| *How and when to update people waiting* | People waiting for THA or TKA surgery should be kept up to date every 3-months about where they are on the waiting list and what they can be doing to support themselves |
|  | People on the waiting list for THA or TKA surgery should be able to request a review if they feel their condition is deteriorating |
| *Waiting well consultation* | People on the waiting list should be provided with telephone number and/or email address to contact orthopaedic services with any questions or queries |
|  | People on the waiting list should be provided with telephone number and/or email address to contact orthopaedic services with any questions or queries |
| *Timing for prehabilitation or pre-operative education* | Prehabilitation should start when people are placed on the waiting list for THA or TKA surgery rather than waiting until 12-weeks before surgery |
|  | A form of pre-operative education/joint school should take place when patients are placed on the waiting list for THA or TKA rather than waiting until 12-weeks before surgery |
|  | A prehabilitation refresher should happen 12 weeks before surgery at the pre-operative assessment |
| *Physiotherapy self-referral* | People should be able to self-refer to physiotherapy if they feel they need support while waiting for THA/TKA surgery? |
| *Social care* | Social care and needs that are not currently being addressed should be identified when patients are placed on a waiting list for THA/TKA surgery |
|  | People waiting for THA/TKA should be reassessed for social care needs at the preoperative assessments |

## References

Ackerman, I. N., Bennell, K. L., & Osborne, R. H. (2011). Decline in Health-Related Quality of Life reported by more than half of those waiting for joint replacement surgery: a prospective cohort study. BMC Musculoskeletal Disorders, 12(1), 108. 10.1186/1471-2474-12-108

Ainslie-Garcia, M., Anderson, L. A., Bloch, B. V, Board, T. N., Chen, A. F., Craigie, S.,…Zagra, L. (2024). International Delphi Study on Wound Closure and Incision Management in Joint Arthroplasty Part 2: Total Hip Arthroplasty. The Journal of Arthroplasty, 39(6), 1524–1529. 10.1016/j.arth.2024.01.047

Anderson, A. M., Comer, C., Smith, T. O., Drew, B. T., Pandit, H., Antcliff, D.,…McHugh, G. A. (2021). Consensus on pre-operative total knee replacement education and prehabilitation recommendations: a UK-based modified Delphi study. BMC Musculoskeletal Disorders, 22(1), 352. 10.1186/s12891-021-04160-5

Baker, P., Kottam, L., Coole, C., Drummond, A., McDaid, C., & Rangan, A. (2020). Development of an occupational advice intervention for patients undergoing elective hip and knee replacement: A Delphi study. BMJ Open, 10(7). 10.1136/bmjopen-2019-036191

Beiderbeck, D., Frevel, N., von der Gracht, H. A., Schmidt, S. L., & Schweitzer, V. M. (2021). The impact of COVID-19 on the European football ecosystem – A Delphi-based scenario analysis. Technological Forecasting and Social Change, 165, 120577. 10.1016/j.techfore.2021.120577

Burns, R. J., Rothman, A. J., Fu, S. S., Lindgren, B., Vock, D. M., & Joseph, A. M. (2016). Longitudinal Care Improves Cessation in Smokers Who Do Not Initially Respond to Treatment by Increasing Cessation Self-Efficacy, Satisfaction, and Readiness to Quit: A Mediated Moderation Analysis. Annals of Behavioral Medicine, 50(1), 58–69. 10.1007/s12160-015-9732-1

de Villiers, M. R., de Villiers, P. J. T., & Kent, A. P. (2005). The Delphi technique in health sciences education research. Medical Teacher, 27(7), 639–643. 10.1080/13611260500069947

Doiron-Cadrin, P., Kairy, D., Vendittoli, P.-A., Lowry, V., Poitras, S., & Desmeules, F. (2020). Feasibility and preliminary effects of a tele-prehabilitation program and an in-person prehablitation program compared to usual care for total hip or knee arthroplasty candidates: a pilot randomized controlled trial. Disability and Rehabilitation, 42(7), 989–998. 10.1080/09638288.2018.1515992

Farrow, L., McLoughlin, J., Gaba, S., & Ashcroft, G. P. (2023). Future demand for primary hip and knee arthroplasty in Scotland. Musculoskeletal Care, 21(2), 355–361. 10.1002/msc.1701

Fearon, A. M., Grimaldi, A., Mellor, R., Nasser, A. M., Fitzpatrick, J., Ladurner, A.,…Vicenzino, B. (2024). ICON 2020—International Scientific Tendinopathy Symposium Consensus: the development of a core outcome set for gluteal tendinopathy. British Journal of Sports Medicine, 58(5), 245. 10.1136/bjsports-2023-107150

Fong, M., Kaner, E., Rowland, M., Graham, H. E., McEvoy, L., Hallsworth, K.,…Madigan, C. D. (2023). The effect of preoperative behaviour change interventions on pre- and post-surgery health behaviours, health outcomes, and health inequalities in adults: A systematic review and meta-analyses. PLOS ONE, 18(7), e0286757-. Retrieved from 10.1371/journal.pone.0286757

French, J. M. R., Deere, K., Jones, T., Pegg, D. J., Reed, M. R., Whitehouse, M. R., & Sayers, A. (2024). An analysis of the effect of the COVID-19-induced joint replacement deficit in England, Wales, and Northern Ireland suggests recovery will be protracted. The Bone & Joint Journal, 106-B(8), 834–841. doi:10.1302/0301-620X.106B8.BJJ-2024-0036.R1

Grocott, M. P. W., Plumb, J. O. M., Edwards, M., Fecher-Jones, I., & Levett, D. Z. H. (2017). Re-designing the pathway to surgery: better care and added value. Perioperative Medicine, 6(1), 9. 10.1186/s13741-017-0065-4

Jennison, T., MacGregor, A., & Goldberg, A. (2023). Hip arthroplasty practice across the Organisation for Economic Co-operation and Development (OECD) over the last decade. The Annals of The Royal College of Surgeons of England, 105(7), 645–652. 10.1308/rcsann.2022.0101

Jünger, Saskia, Payne, Sheila A, Brine, Jenny, Radbruch, Lukas, & Brearley, Sarah G. (2017). Guidance on Conducting and REporting DElphi Studies (CREDES) in palliative care: Recommendations based on a methodological systematic review. Palliative Medicine, 31(8), 684–706. 10.1177/0269216317690685

Manning, L., & Islam, Md. S. (2023). A systematic review to identify the challenges to achieving effective patient flow in public hospitals. The International Journal of Health Planning and Management, 38(3), 805–828. 10.1002/hpm.3626

Miller, W. C., Mohammadi, S., Watson, W., Crocker, M., & Westby, M. (2021). The Hip Instructional Prehabilitation Program for Enhanced Recovery (HIPPER) as an eHealth Approach to Presurgical Hip Replacement Education: Protocol for a Randomized Controlled Trial. JMIR Res Protoc, 10(7), e29322. 10.2196/29322

National Institute for Health and Care Excellence. (2020). Joint replacement (primary): hip, knee and shoulder | Guidance. Retrieved from NICE website: https://www.nice.org.uk/guidance/ng157/chapter/Context

National Institute for Health and Care Excellence. (2022). Visual summary on the management of osteoarthritis. Retrieved from NICE website: https://www.nice.org.uk/guidance/ng226/resources/visual-summary-on-the-management-of-osteoarthritis-pdf-11251842157

Niederberger, M., & Spranger, J. (2020). Delphi Technique in Health Sciences: A Map. Frontiers in Public Health, Volume 8-2020. Retrieved from https://www.frontiersin.org/journals/public-health/articles/10.3389/fpubh.2020.00457

Omar, I., Wylde, V., Fogg, J., Whitehouse, M., & Bertram, W. (2025). Pre-operative education and prehabilitation provision for patients undergoing hip and knee replacement: a national survey of current NHS practice. BMC Musculoskeletal Disorders, 26(1), 421. 10.1186/s12891-025-08637-5

Page, B., Irving, D., Amalberti, R., & Vincent, C. (2024). Health services under pressure: a scoping review and development of a taxonomy of adaptive strategies. BMJ Quality & Safety, 33(11), 738. 10.1136/bmjqs-2023-016686

Plenge, U., Nortje, M. B., Marais, L. C., Jordaan, J. D., Parker, R., van der Westhuizen, N.,…Biccard, B. M. (2018). Optimising perioperative care for hip and knee arthroplasty in South Africa: a Delphi consensus study. BMC Musculoskeletal Disorders, 19(1), 140. 10.1186/s12891-018-2062-2

Prochaska, J. O., DiClemente, C. C., & Norcross, J. C. (1992). In search of how people change. Applications to addictive behaviors. The American Psychologist, 47(9), 1102–1114. 10.1037//0003-066x.47.9.1102

Public Health Scotland. (2024). Scottish Arthroplasty Project national report 2024. Retrieved from https://www.publichealthscotland.scot/media/28353/2024-08-06-sap-report-2024.pdf.

Public Health Scotland. (2025). View hospital waiting times. Retrieved from https://waitingtimes.publichealthscotland.scot/view-waiting-times/

Shichman, I., Roof, M., Askew, N., Nherera, L., Rozell, J. C., Seyler, T. M., & Schwarzkopf, R. (2023). Projections and Epidemiology of Primary Hip and Knee Arthroplasty in Medicare Patients to 2040-2060. JBJS Open Access, 8(1). Retrieved from https://journals.lww.com/jbjsoa/fulltext/2023/03000/projections_and_epidemiology_of_primary_hip_and.22.aspx

Singh, B., Murphy, A., Maher, C., & Smith, A. E. (2024, December 1). Time to Form a Habit: A Systematic Review and Meta-Analysis of Health Behaviour Habit Formation and Its Determinants. Healthcare (Switzerland), Vol. 12. Multidisciplinary Digital Publishing Institute (MDPI). 10.3390/healthcare12232488

Singh, N., Stewart, R. A. H., & Benatar, J. R. (2019). Intensity and duration of lifestyle interventions for long-term weight loss and association with mortality: a meta-analysis of randomised trials. BMJ Open, 9(8), e029966. 10.1136/bmjopen-2019-029966

Steinmetz, J. D., Culbreth, G. T., Haile, L. M., Rafferty, Q., Lo, J., Fukutaki, K. G.,…Kopec, J. A. (2023). Global, regional, and national burden of osteoarthritis, 1990-2020 and projections to 2050: a systematic analysis for the Global Burden of Disease Study 2021. The Lancet Rheumatology, 5(9), e508–e522. 10.1016/S2665-9913(23)00163-7

Straat, A. C., Maarleveld, J. M., Smit, D. J. M., Visch, L., Hulsegge, G., Huirne, J. A. F.,…Kuijer, P. P. F. M. (2023). (Cost-)effectiveness of a personalized multidisciplinary eHealth intervention for knee arthroplasty patients to enhance return to activities of daily life, work and sports – rationale and protocol of the multicentre ACTIVE randomized controlled trial. BMC Musculoskeletal Disorders, 24(1), 162. 10.1186/s12891-023-06236-w

The Royal Australian College of General Practitioners. (2025). Guideline for the management of knee and hip osteoarthritis (2nd ed.). Retrieved from https://www.racgp.org.au/clinical-resources/clinical-guidelines/key-racgp-guidelines/view-all-racgp-guidelines/knee-and-hip-osteoarthritis

Tønnesen, H., Raffing, R., Lauridsen, S. V., Lauritzen, J. B., Elholm, A. M. H., Jensen, H. S.,…Combalia, A. (2023). Two novel prehabilitation apps to help patients stop smoking and risky drinking prior to hip and knee arthroplasty. International Orthopaedics, 47(11), 2645–2653. 10.1007/s00264-023-05890-y

Wainwright, T W, Immins, T., & Middleton, R. G. (2024). Changes in hip and knee arthroplasty practice post-COVID-19 in the English NHS: a retrospective analysis of hospital episode statistics data. The Annals of The Royal College of Surgeons of England, 107(4), 253–256. 10.1308/rcsann.2024.0100

Wainwright, Thomas W, Gill, M., Mcdonald, D. A., G Middleton, R., Reed, M., Sahota, O.,…Ljungqvist, O. (2019). Consensus statement for perioperative care in total hip replacement and total knee replacement surgery: Enhanced Recovery After Surgery (ERAS®) Society recommendations. Acta Orthopaedica, 91(1), 3–19. 10.1080/17453674.2019.1683790

Yapp, L. Z., Clarke, J. V, Moran, M., Simpson, A. H. R. W., & Scott, C. E. H. (2021). National operating volume for primary hip and knee arthroplasty in the COVID-19 era: a study utilizing the Scottish arthroplasty project dataset. Bone & Joint Open, 2(3), 203–210. doi:10.1302/2633-1462.23.BJO-2020-0193.R1

