## Supplementary File 1 for "Consensus recommendations for supporting people waiting for total hip and knee arthroplasty in Scotland: A modified Delphi study"

**Supporting people while they wait for total hip or knee replacement surgery in Scotland: A modified Delphi study**

**Supplementary File 1**

**Round 1 survey pages 2-34**

**Round 2 survey pages 35-59**

**Round 3 survey pages 60-65**

**Supporting people while they wait for total hip or knee replacement surgery in Scotland: A modified Delphi study**

### Round 1 Survey

The Online Survey was hosted on JISC online survey platform. Panellists were only able to access the survey once they had completed the consent and were provided a link to the survey via email.

Panellists were asked to rate their level of agreement using the following scale:

| | Strongly disagree | Disagree | Neither agree nor disagree | Agree | Strongly disagree |
|---|---|---|---|---|---|

#### Page 1: Introduction

Thank you for agreeing to take part in this research study.

The aim of this study is to involve service users (people who have experience waiting for hip and/or knee replacement surgery), experts (clinicians and researchers), policymakers, and funders in establishing consensus on a future Scottish care pathway for patients waiting for total hip and knee replacement surgery.

Taking part involves up to three online questionnaires over a period of approximately 3-months, followed by an optional online workshop which we anticipate will take place in October or November 2024. If we do not reach consensus then a second workshop may be required.

This is the first questionnaire, and it includes statements on interventions to support people waiting for hip or knee replacement surgery. The following topics are covered:

- Pre-operative education including what topics should be included, how it should be delivered and what exercises should be covered.
- Patient optimisation (enhancing patient health before surgery) including smoking cessation, alcohol reduction, and other health factors.
- Other interventions including prehabilitation, psychologically informed interventions, weight management interventions, and interventions for other health factors.
- Support for those waiting longer than the 18-weeks referral to treatment target. For example signposting to information and support, interventions to support physical activity, contacting services for updates "waiting well" consultations, and timing for prehabilitation and pre-operative education.

Throughout this questionnaire we will use the following abbreviations:

- THR for total HIP replacement
- TKR for total KNEE replacement

Throughout the questionnaire you will be asked how much you agree with various statements. You will rate your agreement on a 7-point scale from 1(strongly disagree) to 5 (strongly agree). You will also have the opportunity to provide comments throughout the questionnaire. You are free to write anything that you consider relevant. Your comments may be shared with the other people who are taking part in this study, but it will not be possible for them to identify who made the comments.

We anticipate that the questionnaire will take approximately 30 minutes to complete.

If required, there is an option to save your responses and return to finish by clicking "finish later".

1. Please enter your unique identifier code below (this is the code that we sent to you along with the participant information sheet. If you cannot locate it please contact

Page: 2

2. Do you have experience with THR or TKR or both THR and TKR?
   - THR
   - TKR
   - Both THR and THR

Panellist who selected Both THR and THR saw questions 3-52, Panellists who selected TKR saw questions 53-102 and Panellists who selected THR saw questions 103-152

**Page 3: Interventions to support people waiting for hip or knee replacement surgery**

This section asks for your opinions on a range of interventions that may be used to support people waiting for a hip (THR) or knee (TKR) replacement in Scotland. Sometimes the questions relate to both THR and TKR together and sometimes they are separate. Please answer according to the type of surgery that you have experience of.

**1. Pre-Operative education**

Pre-operative education is part of routine care for people waiting for THR or TKR surgery. It is often delivered as a face-to-face "joint class" when patients are within 12-weeks of their expected surgery date, and often combined with a pre-operative assessment clinic appointment.

This section asks about pre-operative education for hip/knee surgery.

The statements in this section are principally informed by recommendations for TKR pre-operative education published in 2021 (UK, Anderson et al https://doi.org/10.1186/s12891-021-04160-5), supplemented by a review of pre-operative education content at ten high-volume joint replacement centres (USA, Pitaro et al, 2023 https://doi.org/10.1111/jep.13865), 19 other research studies, and a review of publicly available information on THR and TKR pre-operative education. There are three parts to this section: (i) topics that should be included in pre-operative education; (ii) how pre-operative education should be delivered, and (iii) what exercise types should be recommended as part of pre-operative assessment. Please read each of the following statements and indicate your level of agreement, providing additional comments where you would like to.

**Part 1: topics**

3. **Topics that should be included in pre-operative education for THR or TKR surgery:**

Background information

- Anatomy of the hip/knee joint
- Health conditions that may contribute to needing THR/TKR surgery
- Alternative options to THR/TKR surgery

4. Comments: e.g., other topics that should be included or comments related to the above statements

5. Preparing for THR/TKR surgery should cover:
    - Surgical site care (e.g., pre-operative bathing)
    - Purpose of pre-operative rehabilitation
    - Patient involvement in their own management
    - Goal setting
    - Using heat and cold for pain relief
    - Obtaining and using walking aids and other equipment
    - Making home preparations
    - Arranging any social support
    - Arranging transport to and from the hospital

- Optimising management of diabetes
- Emotional well-being
- Education for other people, such as carers
- Identifying and arranging any social care provision

6. Comments: e.g., other topics that should be included

7. Understanding what to expect while in hospital and after going home should cover
    - What to expect during the hospital stay
    - Packing list for hospital
    - In-hospital timeline
    - Visitor information
    - Day of surgery logistics
    - What a THR/TKR surgical procedure involves
    - Information on the prosthesis (replacement)
    - Anaesthesia options
    - Risks of THR/TKR surgery and how to minimise them
    - Common issues that may occur following THR/TKR surgery which do not need to cause alarm
    - Pain expectations
    - Swelling
    - Wound healing
    - What to expect following discharge
    - Recovery expectations

8. Comments: e.g., other topics that should be included

9. Recovering from THR/TKR surgery should cover:
    - Post-operative infection prevention
    - Organising help if complications occur
    - Pain management
    - Nutrition
    - Precautions (e.g., movements/activities to avoid in early post-operative period)
    - Rehabilitation following THR/TKR surgery
    - Returning to daily activities
    - Returning to a normal walking pattern

- Returning to driving and other types of travel
- Returning to sports and leisure activities
- Returning to work
- Realistic timelines for recovery and activities
- When to ask for help (e.g. wound care)

10. Comments: e.g., other topics that should be included

11. Healthy lifestyle guidance should cover:
    - Physical activity
    - Weight management
    - Stopping smoking
    - Recommended alcohol guidelines and cutting down on drinking

12. Comments: e.g., other topics that should be included

**Part ii: delivery**

13. **Delivery of pre-operative education.**

Pre-operative education should:

- Be informed by a multi-disciplinary team, including members of the orthopaedic surgery team, nursing, physiotherapy, and occupational therapy teams
- Be informed by patients who have previously had THR/TKR surgery

14. Comments: e.g., other topics that should be included

15. Pre-operative information should:
    - Be delivered, at least partly, by providing examples of other patients' experiences of THR/TKR surgery
    - Be delivered 1-1 if the patient does not wish to attend a group session
    - Be delivered using a combination of more than one format, including face to face group sessions, a booklet or other written format and a website or other electronic format

- Be delivered through a combination of providing the patient with information and giving them an opportunity to actively take part in tasks
- Be provided separately for THR and TKR – i.e., patients waiting for THR should receive group education separately from patients waiting for TKR
- Provide an opportunity for the patient's questions to be addressed
- Provide an opportunity for a family member or friend of the patient to be involved
- Be delivered, at least partly, within four weeks of the patient's surgery

16. Any other comments. (including timing and mode of delivery for pre-operative education: e.g. anything else)

17. **Part iii exercise types**

**Pre-operative exercise types**

A pre-operative exercise programme should include:

- Leg strengthening exercises
- Leg flexibility exercises
- Balance exercises
- Functional movement exercises - i.e., sit to stand exercises
- Cardiovascular exercises
- Core control exercises
- Walking practice with walking aids
- Training on steps
- Practicing post operative exercises
- Delivery should be tailored according to each patient's individual needs
- Delivery should be standardised across Scotland

18. Comments about exercise types (e.g., anything that is missing):

**2. Patient Optimisation**

**Patient optimisation for THR and TKR**

Patient optimisation is the process of supporting patients to be as healthy as possible before surgery and address any health problems that may affect this (NHS England » Earlier screening, risk assessment and health optimisation in perioperative pathways: guide for providers and integrated care boards). This can include the management of long-term health conditions and support people to make changes to improve their health (The following recommendations have been informed by the ERAS Society (Enhanced recovery after Surgery®) publication (Wainwright et al [2020] https://doi.org/10.1080/17453674.2019.1683790). Please read the recommendations below and consider how much you agree with the statements related to them.

19. ***Smoking cessation*** of four-weeks or more is recommended before THR or TKR
    - Patients who identify as smokers should be referred to a smoking cessation programme while they are waiting for surgery
    - Referral to smoking cessation programmes should be made as soon as patients are placed on the waiting list for surgery
    - Referral should be made at the same time as pre-operative education/assessment (typically within 12-weeks of surgery)

20. Please make any comments here about smoking cessation interventions:

21. ***Alcohol cessation*** programmes are recommended before THR or TKR for patients who misuse alcohol or report high alcohol intake
    - Patients waiting for THR or TKR who report high alcohol intake/alcohol misuse should be referred to an alcohol cessation programme
    - Referral to an alcohol cessation programme should be made as soon as patients are placed on the waiting list
    - Referral to an alcohol cessation programme should be made at the same time as pre-operative education/assessment (typically within 12-weeks of surgery)

22. Please make any comments here about alcohol cessation interventions:

23. ***Pre-operative anaemia (lower than normal amount of healthy red blood cells)***
    - Pre-operative anaemia should be identified and corrected at the pre-operative assessment (usually 12-weeks before surgery) for THR/TKR
    - Pre-operative anaemia should be investigated and corrected prior to THR/TKR surgery
    - Pre-operative anaemia should be identified, investigated and corrected as soon as patients are placed on the waiting list for THR/TKR

24. Please make any comments here about the identification, investigation and correction of pre-operative anaemia:

25. ***Pre-operative fasting: Intake of clear fluids until 2 hours before surgery and a 6 hour fast for solid food before surgery for TKR/THR***
    - Patients should be advised to continue the intake of clear fluids until 2-hours before the start of anaesthesia, and to fast (solid foods) for 6-hours before the start of anaesthesia for THR/TKR

26. Please make any comments here about pre-operative fluid intake & fasting

27. ***Pre-operative carbohydrate loading: in THR/TKR carbohydrate loading may improve patient well-being and metabolism, but has not been shown to accelerate the achievement of discharge criteria or reduce complications***
    - Pre-operative carbohydrate loading should not be recommended prior to THR/TKR

28. Please make any comments here about carbohydrate loading:

29. ***Pre-operative physiotherapy: current evidence does not support preoperative physiotherapy as an essential intervention***
    - Pre-operative physiotherapy is not essential for patients waiting for THR/TKR

30. Please make any comments here about pre-operative physiotherapy:

**3. Other interventions to support people waiting for THR and TKR surgery**

The following questions have been informed by a review of evidence (research and reports) conducted by the research team and include more than 400 sources of evidence

31. "Prehabilitation can be defined as a formal pre-operative programme, aimed at enabling the patient to withstand the stress of surgery. Prehabilitation for THR/TKR may combine exercise with other interventions (e.g., education, information provision)."

Prehabilitation should be offered to people waiting for THR/TKR

Prehabilitation should be offered within 12-weeks of THR/TKR surgery

Prehabilitation should be offered to people when they are placed on the waiting list for THR/TKR

32. Please provide any comments on the timing of prehabilitation and comments to support your answer

33. **Psychologically informed interventions**

- Psychological informed interventions should be offered to people waiting for THR/TKR
- Psychological informed interventions should be offered to people waiting for THR/TKR
- Psychologically informed Interventions should be offered within 12-weeks of THR/TKR surgery
- Patients waiting for THR/TKR surgery who have been formally diagnosed with anxiety or depression should be offered referral to cognitive behavioural therapy (CBT) based therapy
- Mindfulness should be considered for people waiting for THR/TKR to help with post-operative pain and opioid use (Mindfulness is about living more in the present moment, appreciating the here and now, and not dwelling too much on the past or future)
- Opioid counselling should be offered to patients on the waiting list for THR/TKR surgery who are routinely taking prescription opioids

34. Please make any comments about psychological interventions (opioid counselling, mindfulness and CBT) here:

35. **Weight management**

Weight management interventions should be offered to people waiting for THR/TKR

Weight management interventions should be offered to people when they are placed on the waiting list for THR/TKR

Weight management interventions should be offered within 12-weeks of THR/TKR surgery

Patients waiting for THR/TKR surgery who have a body mass index (BMI) of 27 $kg/m^2$ or over should be offered referral to a weight management programme. NB a BMI of 25 is considered overweight and over 30 is considered obese

36. Please make any comments about weight-loss programmes for people waiting for THR/TKR here:

### 4. Waiting a long time for Surgery

#### Waiting a long time for THR and TKR

In Scotland, waiting times for THR and TKR are at a record high. Interventions such as pre-operative education and prehabilitation are typically offered close to the expected date of surgery, meaning that other interventions may be required to support people while they wait. The following statements relate to patients waiting for longer than the 18-weeks referral to treatment target.

37. ***Signposting to web-based information***
    - People waiting for THR/TKR surgery should be signposted to web-based information about physical activity and fitness
    - People waiting for THR/TKR surgery should be signposted to web-based information about diet and nutrition
    - People waiting for THR/TKR surgery should be signposted to web-based information about mental wellbeing
    - People waiting for THR/TKR surgery should be signposted to web-based information about pain management
    - People waiting for THR/TKR surgery should be signposted to web-based information about smoking cessation
    - People waiting for THR/TKR surgery should be signposted to web-based information about alcohol reduction

38. Please make any comments about web-based information for people waiting for THR/TKR here:

39. ***Written information***
    - People waiting for THR/TKR surgery should be sent written information if they prefer it to web-based materials

40. **Signposting to support**

- People waiting for THR/TKR surgery should be signposted to third sector organisations
- People waiting for THR surgery should be offered interventions that support then to maintain/increase physical activity levels while they are waiting for surgery
- If required people waiting for THR/TKR surgery should be offered referral to relevant social care services

41. Please make any comments about signposting to social care and third sector organisations for people waiting for THR/TKR here:

42. ***How and when to update people waiting***
    - People waiting for THR or TKR surgery should be kept up to date every 3-months about where they are on the waiting list and what they can be doing to support themselves
    - People on the waiting list for THR or TKR surgery should be able to request a review if they feel their condition is deteriorating

43. Please make comments here, e.g., who should keep people up to date and how this could be managed

44. Please comment on patient-initiated review, e.g., how this should be triggered

45. ***Waiting well consultation***
    - People waiting for THR and TKR surgery should be offered at least one 'waiting well' telephone consultation to provide wellbeing advice, signposting, encourage vaccination & screening uptake, and referral to relevant health and social care services
    - People on the waiting list should be provided with telephone number and/or email address to contact orthopaedic services with any questions or queries

46. Please make comments here, e.g., who should provide a 'waiting well' telephone consultation

47. ***Timing for prehabilitation or pre-operative education***
    - Prehabilitation should start when people are placed on the waiting list for THR or TKR surgery rather than waiting until 12-weeks before surgery
    - A form of pre-operative education/joint school should take place when patients are placed on the waiting list for THR or TKR rather than waiting until 12-weeks before surgery
    - A prehabilitation refresher should happen 12 weeks before surgery at the pre-operative assessment

48. Please comment on the timing of prehabilitation

49. **Physiotherapy self-referral**
    - People should be able to self-refer to physiotherapy if they feel they need support while waiting for THR/TKR surgery?

50. **Social Care**
    - Social care and needs that are not currently being addressed should be identified when patients are placed on a waiting list for THR/TKR surgery
    - People waiting for THR/TKR should be reassessed for social care needs at the preoperative assessments

51. Please comment on social care here:

52. Please tell us anything else you think should be considered for people waiting for THR and TKR surgery

**Page 4: Interventions to support people waiting for hip replacement surgery**

This section asks for your opinions on a range of interventions that may be used to support people waiting for a hip (THR) replacement in Scotland. Some of the topics and questions are relevant to hip (THR) and knee (TKR) surgery, therefore you will see both terms in some questions. Please answer from your knowledge of waiting for THR.

**1. Pre-Operative education**

Pre-operative education is part of routine care for people waiting for THR or TKR surgery. It is often delivered as a face-to-face "joint class" when patients are within 12-weeks of their expected surgery date, and often combined with a pre-operative assessment clinic appointment.

This section asks about pre-operative education for hip/knee surgery.

The statements in this section are principally informed by recommendations for TKR pre-operative education published in 2021 (UK, Anderson et al https://doi.org/10.1186/s12891-021-04160-5), supplemented by a review of pre-operative education content at ten high-volume joint replacement centres (USA, Pitaro et al, 2023 https://doi.org/10.1111/jep.13865), 19 other research studies, and a review of publicly available information on THR and TKR pre-operative education. There are three parts to this section: (i) topics that should be included in pre-operative education; (ii) how pre-operative education should be delivered, and (iii) what exercise types should be recommended as part of pre-operative assessment. Please read each of the following statements and indicate your level of agreement, providing additional comments where you would like to.

**Part 1: topics**

53. **Topics that should be included in pre-operative education for THR or TKR surgery:**

Background information

- Anatomy of the hip/knee joint
- Health conditions that may contribute to needing THR/TKR surgery
- Alternative options to THR/TKR surgery

54. Comments: e.g., other topics that should be included or comments related to the above statements

55. Preparing for THR/TKR surgery should cover:

- Surgical site care (e.g., pre-operative bathing)
- Purpose of pre-operative rehabilitation
- Patient involvement in their own management
- Goal setting
- Using heat and cold for pain relief

- Obtaining and using walking aids and other equipment
- Making home preparations
- Arranging any social support
- Arranging transport to and from the hospital
- Optimising management of diabetes
- Emotional well-being
- Education for other people, such as carers
- Identifying and arranging any social care provision

56. Comments: e.g., other topics that should be included

57. Understanding what to expect while in hospital and after going home should cover
    - What to expect during the hospital stay
    - Packing list for hospital
    - In-hospital timeline
    - Visitor information
    - Day of surgery logistics
    - What a THR/TKR surgical procedure involves
    - Information on the prosthesis (replacement)
    - Anaesthesia options
    - Risks of THR/TKR surgery and how to minimise them
    - Common issues that may occur following THR/TKR surgery which do not need to cause alarm
    - Pain expectations
    - Swelling
    - Wound healing
    - What to expect following discharge
    - Recovery expectations

58. Comments: e.g., other topics that should be included

59. Recovering from THR/TKR surgery should cover:
    - Post-operative infection prevention
    - Organising help if complications occur
    - Pain management
    - Nutrition

- Precautions (e.g., movements/activities to avoid in early post-operative period)
- Rehabilitation following THR/TKR surgery
- Returning to daily activities
- Returning to a normal walking pattern
- Returning to driving and other types of travel
- Returning to sports and leisure activities
- Returning to work
- Realistic timelines for recovery and activities
- When to ask for help (e.g. wound care)

60. Comments: e.g., other topics that should be included

61. Healthy lifestyle guidance should cover:
    - Physical activity
    - Weight management
    - Stopping smoking
    - Recommended alcohol guidelines and cutting down on drinking

62. Comments: e.g., other topics that should be included

**Part ii: delivery**

63. **Delivery of pre-operative education.**

Pre-operative education should:

- Be informed by a multi-disciplinary team, including members of the orthopaedic surgery team, nursing, physiotherapy, and occupational therapy teams
- Be informed by patients who have previously had THR/TKR surgery

64. Comments: e.g., other topics that should be included

65. Pre-operative information should:
    - Be delivered, at least partly, by providing examples of other patients' experiences of THR/TKR surgery
    - Be delivered 1-1 if the patient does not wish to attend a group session

- Be delivered using a combination of more than one format, including face to face group sessions, a booklet or other written format and a website or other electronic format
- Be delivered through a combination of providing the patient with information and giving them an opportunity to actively take part in tasks
- Be provided separately for THR and TKR – i.e., patients waiting for THR should receive group education separately from patients waiting for TKR
- Provide an opportunity for the patient’s questions to be addressed
- Provide an opportunity for a family member or friend of the patient to be involved
- Be delivered, at least partly, within four weeks of the patient's surgery

66. Any other comments. (including timing and mode of delivery for pre-operative education: e.g. anything else)

67. **Part iii exercise types**

**Pre-operative exercise types**

A pre-operative exercise programme should include:

- Leg strengthening exercises
- Leg flexibility exercises
- Balance exercises
- Functional movement exercises - i.e., sit to stand exercises
- Cardiovascular exercises
- Core control exercises
- Walking practice with walking aids
- Training on steps
- Practicing post operative exercises
- Delivery should be tailored according to each patient’s individual needs
- Delivery should be standardised across Scotland

68. Comments about exercise types (e.g., anything that is missing):

**2. Patient Optimisation**

**Patient optimisation for THR**

Patient optimisation is the process of supporting patients to be as healthy as possible before surgery and address any health problems that may affect this (NHS England » Earlier screening, risk assessment and health optimisation in perioperative pathways: guide for providers and integrated care boards). This can include the management of long-term health conditions and support people to make changes to improve their health (The following recommendations have been informed by the ERAS Society (Enhanced recovery after Surgery®) publication (Wainwright et al [2020] https://doi.org/10.1080/17453674.2019.1683790). Please read the recommendations below and consider how much you agree with the statements related to them.

69. ***Smoking cessation*** of four-weeks or more is recommended before THR
- Patients who identify as smokers should be referred to a smoking cessation programme while they are waiting for surgery
- Referral to smoking cessation programmes should be made as soon as patients are placed on the waiting list for surgery
- Referral should be made at the same time as pre-operative education/assessment (typically within 12-weeks of surgery)

70. Please make any comments here about smoking cessation interventions:

71. ***Alcohol cessation*** programmes are recommended before THR for patients who misuse alcohol or report high alcohol intake
- Patients waiting for THR who report high alcohol intake/alcohol misuse should be referred to an alcohol cessation programme
- Referral to an alcohol cessation programme should be made as soon as patients are placed on the waiting list
- Referral to an alcohol cessation programme should be made at the same time as pre-operative education/assessment (typically within 12-weeks of surgery)

72. Please make any comments here about alcohol cessation interventions:

73. ***Pre-operative anaemia (lower than normal amount of healthy red blood cells)***
- Pre-operative anaemia should be identified and corrected at the pre-operative assessment (usually 12-weeks before surgery) for THR
- Pre-operative anaemia should be investigated and corrected prior to THR surgery
- Pre-operative anaemia should be identified, investigated and corrected as soon as patients are placed on the waiting list for THR

74. Please make any comments here about the identification, investigation and correction of pre-operative anaemia:

75. ***Pre-operative fasting: Intake of clear fluids until 2 hours before surgery and a 6 hour fast for solid food before surgery for TKR***
    - Patients should be advised to continue the intake of clear fluids until 2-hours before the start of anaesthesia, and to fast (solid foods) for 6-hours before the start of anaesthesia for THR
76. Please make any comments here about pre-operative fluid intake & fasting

77. ***Pre-operative carbohydrate loading: in THR carbohydrate loading may improve patient well-being and metabolism, but has not been shown to accelerate the achievement of discharge criteria or reduce complications***
    - Pre-operative carbohydrate loading should not be recommended prior to THR

78. Please make any comments here about carbohydrate loading:

79. ***Pre-operative physiotherapy: current evidence does not support preoperative physiotherapy as an essential intervention***
    - Pre-operative physiotherapy is not essential for patients waiting for THR

80. Please make any comments here about pre-operative physiotherapy:

**3. Other Interventions**

**Other interventions to support people waiting for THR surgery**

The following questions have been informed by a review of evidence (research and reports) conducted by the research team and include more than 400 sources of evidence

81. "Prehabilitation can be defined as a formal pre-operative programme, aimed at enabling the patient to withstand the stress of surgery. Prehabilitation for THR may combine exercise with other interventions (e.g., education, information provision)."
    - Prehabilitation should be offered to people waiting for THR
    - Prehabilitation should be offered within 12-weeks of THR surgery
    - Prehabilitation should be offered to people when they are placed on the waiting list for THR
82. Please provide any comments on the timing of prehabilitation and comments to support your answer

83. **Psychologically informed interventions**
    - Psychological informed interventions should be offered to people waiting for THR
    - Psychological informed interventions should be offered to people waiting for THR
    - Psychologically informed Interventions should be offered within 12-weeks of THR surgery
    - Patients waiting for THR surgery who have been formally diagnosed with anxiety or depression should be offered referral to cognitive behavioural therapy (CBT) based therapy
    - Mindfulness should be considered for people waiting for THR to help with post-operative pain and opioid use (Mindfulness is about living more in the present moment, appreciating the here and now, and not dwelling too much on the past or future)
    - Opioid counselling should be offered to patients on the waiting list for THR surgery who are routinely taking prescription opioids

84. Please make any comments about psychological interventions (opioid counselling, mindfulness and CBT) here:

85. **Weight management**
    - Weight management interventions should be offered to people waiting for THR
    - Weight management interventions should be offered to people when they are placed on the waiting list for THR
    - Weight management interventions should be offered within 12-weeks of THR surgery

- Patients waiting for THR surgery who have a body mass index (BMI) of 27 kg/$m^2$ or over should be offered referral to a weight management programme. NB a BMI of 25 is considered overweight and over 30 is considered obese

86. Please make any comments about weight-loss programmes for people waiting for THR here:

**4. Waiting a long time for Surgery**

**Waiting a long time for THR**

In Scotland, waiting times for THR are at a record high. Interventions such as pre-operative education and prehabilitation are typically offered close to the expected date of surgery, meaning that other interventions may be required to support people while they wait. The following statements relate to patients waiting for longer than the 18-weeks referral to treatment target.

87. ***Signposting to web-based information***
    - People waiting for THR surgery should be signposted to web-based information about physical activity and fitness
    - People waiting for THR surgery should be signposted to web-based information about diet and nutrition
    - People waiting for THR surgery should be signposted to web-based information about mental wellbeing
    - People waiting for THR surgery should be signposted to web-based information about pain management
    - People waiting for THR surgery should be signposted to web-based information about smoking cessation
    - People waiting for THR surgery should be signposted to web-based information about alcohol reduction

88. Please make any comments about web-based information for people waiting for THR here:

89. ***Written information***
    - People waiting for THR surgery should be sent written information if they prefer it to web-based materials

90. **Signposting to support**

- People waiting for THR surgery should be signposted to third sector organisations
- People waiting for THR surgery should be offered interventions that support then to maintain/increase physical activity levels while they are waiting for surgery
- If required people waiting for THR surgery should be offered referral to relevant social care services

91. Please make any comments about signposting to social care and third sector organisations for people waiting for THR here:

92. ***How and when to update people waiting***
    - People waiting for THR surgery should be kept up to date every 3-months about where they are on the waiting list and what they can be doing to support themselves
    - People on the waiting list for THR surgery should be able to request a review if they feel their condition is deteriorating

93. Please make comments here, e.g., who should keep people up to date and how this could be managed

94. Please comment on patient-initiated review, e.g., how this should be triggered

95. ***Waiting well consultation***
    - People waiting for THR surgery should be offered at least one 'waiting well' telephone consultation to provide wellbeing advice, signposting, encourage vaccination & screening uptake, and referral to relevant health and social care services
    - People on the waiting list should be provided with telephone number and/or email address to contact orthopaedic services with any questions or queries

96. Please make comments here, e.g., who should provide a 'waiting well' telephone consultation

97. ***Timing for prehabilitation or pre-operative education***

- Prehabilitation should start when people are placed on the waiting list for THR or surgery rather than waiting until 12-weeks before surgery
- A form of pre-operative education/joint school should take place when patients are placed on the waiting list for THR rather than waiting until 12-weeks before surgery
- A prehabilitation refresher should happen 12 weeks before surgery at the pre-operative assessment

98. Please comment on the timing of prehabilitation

99. **Physiotherapy self-referral**

- People should be able to self-refer to physiotherapy if they feel they need support while waiting for THR surgery?

100. **Social Care**

- Social care and needs that are not currently being addressed should be identified when patients are placed on a waiting list for THR surgery
- People waiting for THR should be reassessed for social care needs at the preoperative assessments

101. Please comment on social care here:

102. Please tell us anything else you think should be considered for people waiting for THR surgery

**Page 5: Interventions to support people waiting for knee replacement surgery**

This section asks for your opinions on a range of interventions that may be used to support people waiting for a knee (TKR) replacement in Scotland. Some of the topics and questions are relevant to hip (THR) and knee (TKR) surgery, therefore you will see both terms in some questions. Please answer from your knowledge of waiting for TKR.

**1. Pre-Operative education**

Pre-operative education is part of routine care for people waiting for THR or TKR surgery. It is often delivered as a face-to-face “joint class” when patients are within 12-weeks of their expected surgery date and often combined with a pre-operative assessment clinic appointment.

This section asks about pre-operative education for hip/knee surgery.

The statements in this section are principally informed by recommendations for TKR pre-operative education published in 2021 (UK, Anderson et al https://doi.org/10.1186/s12891-021-04160-5), supplemented by a review of pre-operative education content at ten high-volume joint replacement centres (USA, Pitaro et al, 2023 https://doi.org/10.1111/jep.13865), 19 other research studies, and a review of publicly available information on THR and TKR pre-operative education. There are three parts to this section: (i) topics that should be included in pre-operative education; (ii) how pre-operative education should be delivered, and (iii) what exercise types should be recommended as part of pre-operative assessment. Please read each of the following statements and indicate your level of agreement, providing additional comments where you would like to.

**Part 1: topics**

103. **Topics that should be included in pre-operative education for THR or TKR surgery:**

Background information

- Anatomy of the hip/knee joint
- Health conditions that may contribute to needing THR/TKR surgery
- Alternative options to THR/TKR surgery

104. Comments: e.g., other topics that should be included or comments related to the above statements

105. Preparing for THR/TKR surgery should cover:
- Surgical site care (e.g., pre-operative bathing)
- Purpose of pre-operative rehabilitation
- Patient involvement in their own management
- Goal setting
- Using heat and cold for pain relief
- Obtaining and using walking aids and other equipment
- Making home preparations

- Arranging any social support
- Arranging transport to and from the hospital
- Optimising management of diabetes
- Emotional well-being
- Education for other people, such as carers
- Identifying and arranging any social care provision

106. Comments: e.g., other topics that should be included

107. Understanding what to expect while in hospital and after going home should cover
    - What to expect during the hospital stay
    - Packing list for hospital
    - In-hospital timeline
    - Visitor information
    - Day of surgery logistics
    - What a THR/TKR surgical procedure involves
    - Information on the prosthesis (replacement)
    - Anaesthesia options
    - Risks of THR/TKR surgery and how to minimise them
    - Common issues that may occur following THR/TKR surgery which do not need to cause alarm
    - Pain expectations
    - Swelling
    - Wound healing
    - What to expect following discharge
    - Recovery expectations

108. Comments: e.g., other topics that should be included

109. Recovering from THR/TKR surgery should cover:
    - Post-operative infection prevention
    - Organising help if complications occur
    - Pain management
    - Nutrition
    - Precautions (e.g., movements/activities to avoid in early post-operative period)

- Rehabilitation following THR/TKR surgery
- Returning to daily activities
- Returning to a normal walking pattern
- Returning to driving and other types of travel
- Returning to sports and leisure activities
- Returning to work
- Realistic timelines for recovery and activities
- When to ask for help (e.g. wound care)

110. Comments: e.g., other topics that should be included

111. Healthy lifestyle guidance should cover:
- Physical activity
- Weight management
- Stopping smoking
- Recommended alcohol guidelines and cutting down on drinking

112. Comments: e.g., other topics that should be included

**Part ii: delivery**

113. **Delivery of pre-operative education.**

Pre-operative education should:

- Be informed by a multi-disciplinary team, including members of the orthopaedic surgery team, nursing, physiotherapy, and occupational therapy teams
- Be informed by patients who have previously had THR/TKR surgery

114. Comments: e.g., other topics that should be included

115. Pre-operative information should:
- Be delivered, at least partly, by providing examples of other patients' experiences of THR/TKR surgery
- Be delivered 1-1 if the patient does not wish to attend a group session

- Be delivered using a combination of more than one format, including face to face group sessions, a booklet or other written format and a website or other electronic format
- Be delivered through a combination of providing the patient with information and giving them an opportunity to actively take part in tasks
- Be provided separately for THR and TKR – i.e., patients waiting for THR should receive group education separately from patients waiting for TKR
- Provide an opportunity for the patient’s questions to be addressed
- Provide an opportunity for a family member or friend of the patient to be involved
- Be delivered, at least partly, within four weeks of the patient's surgery

116. Any other comments. (including timing and mode of delivery for pre-operative education: e.g. anything else)

117. **Part iii exercise types**

**Pre-operative exercise types**

A pre-operative exercise programme should include:

- Leg strengthening exercises
- Leg flexibility exercises
- Balance exercises
- Functional movement exercises - i.e., sit to stand exercises
- Cardiovascular exercises
- Core control exercises
- Walking practice with walking aids
- Training on steps
- Practicing post operative exercises
- Delivery should be tailored according to each patient’s individual needs
- Delivery should be standardised across Scotland

118. Comments about exercise types (e.g., anything that is missing):

**2. Patient Optimisation**

**Patient optimisation for TKR**

Patient optimisation is the process of supporting patients to be as healthy as possible before surgery and address any health problems that may affect this (NHS England » Earlier screening, risk assessment and health optimisation in perioperative pathways: guide for providers and integrated care boards). This can include the management of long-term health conditions and support people to make changes to improve their health (The following recommendations have been informed by the ERAS Society (Enhanced recovery after Surgery®) publication (Wainwright et al [2020] https://doi.org/10.1080/17453674.2019.1683790). Please read the recommendations below and consider how much you agree with the statements related to them.

119. ***Smoking cessation*** of four-weeks or more is recommended before TKR
- Patients who identify as smokers should be referred to a smoking cessation programme while they are waiting for surgery
- Referral to smoking cessation programmes should be made as soon as patients are placed on the waiting list for surgery
- Referral should be made at the same time as pre-operative education/assessment (typically within 12-weeks of surgery)

120. Please make any comments here about smoking cessation interventions:

121. ***Alcohol cessation*** programmes are recommended before TKR for patients who misuse alcohol or report high alcohol intake
- Patients waiting for TKR who report high alcohol intake/alcohol misuse should be referred to an alcohol cessation programme
- Referral to an alcohol cessation programme should be made as soon as patients are placed on the waiting list
- Referral to an alcohol cessation programme should be made at the same time as pre-operative education/assessment (typically within 12-weeks of surgery)

122. Please make any comments here about alcohol cessation interventions:

123. ***Pre-operative anaemia (lower than normal amount of healthy red blood cells)***
- Pre-operative anaemia should be identified and corrected at the pre-operative assessment (usually 12-weeks before surgery) for TKR
- Pre-operative anaemia should be investigated and corrected prior to TKR surgery

- Pre-operative anaemia should be identified, investigated and corrected as soon as patients are placed on the waiting list for TKR

124. Please make any comments here about the identification, investigation and correction of pre-operative anaemia:

125. ***Pre-operative fasting: Intake of clear fluids until 2 hours before surgery and a 6 hour fast for solid food before surgery for TKR***
   - Patients should be advised to continue the intake of clear fluids until 2-hours before the start of anaesthesia, and to fast (solid foods) for 6-hours before the start of anaesthesia for TKR

126. Please make any comments here about pre-operative fluid intake & fasting

127. ***Pre-operative carbohydrate loading: in TKR carbohydrate loading may improve patient well-being and metabolism, but has not been shown to accelerate the achievement of discharge criteria or reduce complications***
   - Pre-operative carbohydrate loading should not be recommended prior to TKR

128. Please make any comments here about carbohydrate loading:

129. ***Pre-operative physiotherapy: current evidence does not support preoperative physiotherapy as an essential intervention***
   - Pre-operative physiotherapy is not essential for patients waiting for TKR

130. Please make any comments here about pre-operative physiotherapy:

**3. Other Interventions**

131. "Prehabilitation can be defined as a formal pre-operative programme, aimed at enabling the patient to withstand the stress of surgery. Prehabilitation for TKR may combine exercise with other interventions (e.g., education, information provision)."

- Prehabilitation should be offered to people waiting for TKR
- Prehabilitation should be offered within 12-weeks of TKR surgery
- Prehabilitation should be offered to people when they are placed on the waiting list for TKR

132. Please provide any comments on the timing of prehabilitation and comments to support your answer

133. **Psychologically informed interventions**

- Psychological informed interventions should be offered to people waiting for TKR
- Psychological informed interventions should be offered to people waiting for TKR
- Psychologically informed Interventions should be offered within 12-weeks of TKR surgery
- Patients waiting for TKR surgery who have been formally diagnosed with anxiety or depression should be offered referral to cognitive behavioural therapy (CBT) based therapy
- Mindfulness should be considered for people waiting for TKR to help with post-operative pain and opioid use (Mindfulness is about living more in the present moment, appreciating the here and now, and not dwelling too much on the past or future)
- Opioid counselling should be offered to patients on the waiting list for TKR surgery who are routinely taking prescription opioids

134. Please make any comments about psychological interventions (opioid counselling, mindfulness and CBT) here:

135. **Weight management**

- Weight management interventions should be offered to people waiting for TKR
- Weight management interventions should be offered to people when they are placed on the waiting list for TKR
- Weight management interventions should be offered within 12-weeks of TKR surgery
- Patients waiting for TKR surgery who have a body mass index (BMI) of 27 $kg/m^2$ or over should be offered referral to a weight management programme. NB a BMI of 25 is considered overweight and over 30 is considered obese

136. Please make any comments about weight-loss programmes for people waiting for TKR here:

**4. Waiting a long time for Surgery**

**Waiting a long time for TKR**

In Scotland, waiting times for TKR are at a record high. Interventions such as pre-operative education and prehabilitation are typically offered close to the expected date of surgery, meaning that other interventions may be required to support people while they wait. The following statements relate to patients waiting for longer than the 18-weeks referral to treatment target.

137. ***Signposting to web-based information***

- People waiting for TKR surgery should be signposted to web-based information about physical activity and fitness
- People waiting for TKR surgery should be signposted to web-based information about diet and nutrition
- People waiting for TKR surgery should be signposted to web-based information about mental wellbeing
- People waiting for TKR surgery should be signposted to web-based information about pain management
- People waiting for TKR surgery should be signposted to web-based information about smoking cessation
- People waiting for TKR surgery should be signposted to web-based information about alcohol reduction

138. Please make any comments about web-based information for people waiting for TKR here:

139. ***Written information***

- People waiting for TKR surgery should be sent written information if they prefer it to web-based materials

140. **Signposting to support**

- People waiting for TKR surgery should be signposted to third sector organisations

- People waiting for TKR surgery should be offered interventions that support then to maintain/increase physical activity levels while they are waiting for surgery
- If required people waiting for TKR surgery should be offered referral to relevant social care services

141. Please make any comments about signposting to social care and third sector organisations for people waiting for TKR here:

142. ***How and when to update people waiting***

- People waiting for TKR surgery should be kept up to date every 3-months about where they are on the waiting list and what they can be doing to support themselves
- People on the waiting list for TKR surgery should be able to request a review if they feel their condition is deteriorating

143. Please make comments here, e.g., who should keep people up to date and how this could be managed

144. Please comment on patient-initiated review, e.g., how this should be triggered

145. ***Waiting well consultation***

- People waiting for TKR surgery should be offered at least one 'waiting well' telephone consultation to provide wellbeing advice, signposting, encourage vaccination & screening uptake, and referral to relevant health and social care services
- People on the waiting list should be provided with telephone number and/or email address to contact orthopaedic services with any questions or queries

146. Please make comments here, e.g., who should provide a 'waiting well' telephone consultation

147. ***Timing for prehabilitation or pre-operative education***

- Prehabilitation should start when people are placed on the waiting list for TKR surgery rather than waiting until 12-weeks before surgery
- A form of pre-operative education/joint school should take place when patients are placed on the waiting list for TKR rather than waiting until 12-weeks before surgery
- A prehabilitation refresher should happen 12 weeks before surgery at the pre-operative assessment

148. Please comment on the timing of prehabilitation

149. **Physiotherapy self-referral**

- People should be able to self-refer to physiotherapy if they feel they need support while waiting for TKR surgery?

150. **Social Care**

- Social care and needs that are not currently being addressed should be identified when patients are placed on a waiting list for TKR surgery
- People waiting for TKR should be reassessed for social care needs at the preoperative assessments

151. Please comment on social care here:

152. Please tell us anything else you think should be considered for people waiting for TKR surgery

**Page 6**

Thank you for taking part in this survey. We appreciate the time you have taken to do this and the information you have provided will be very useful.

We will collect all the responses and analyse the findings together. We will consider statements that more than 70% of participants agree or strongly agree on as having reached consensus. Items rated as disagree or strongly disagree by 50% or more of participants will be excluded from the recommendations on pre-operative interventions. Items that do not reach consensus or exclusion will be included in the next survey that we send to you.

If you have any questions or comments, or do not want to take part in the next round please contact

**Round 2 Survey**

The Online Survey was hosted on JISC online survey platform. Panellists who completed the first survey were able to access the survey via an email link

Panelists were asked to rate their level of agreement using the following scale:

| | Strongly disagree | Disagree | Neither agree nor disagree | Agree | Strongly disagree |
|---|---|---|---|---|---|

**Page 1 – Introduction**

Thank you for taking part in Round 1 of this research study.

As a reminder, the aim of this study is to involve service users (called patients in this survey), as well as experts, policymakers, and funders (together called professionals in this survey) in establishing consensus on a future Scottish care pathway for patients waiting for total hip and knee replacement surgery (THR/TKR throughout this survey).

Taking part involves two online surveys, followed by an optional online workshop, which will take place in February 2025. This is later than we originally told you it would be. It took us longer than anticipated for Round 1 to be completed. We hope you will still be willing to take part in this second survey and consider taking part in a workshop.

***Purpose of Round 2***

Many of the statements we asked you about in Round 1 reached consensus, defined as at least 70% of each group (patients and professionals) agreeing with the statement. These statements will be included in the recommendations for the future Scottish care pathway.

In this survey we will present the levels of agreement for all the statements from Round 1. You do not have to do anything further with the statements that reached consensus (70% agreement or more).

For the statements that **did not reach consensus in one or both groups**, we will present the levels of agreement for each group (patients and professionals) along with a graph showing more detail. For these statements, we will ask you to rate your agreement on a 5-point scale from strongly disagree to strongly agree, as you did in Round 1. Your opinion might stay the same or might change from the previous round, there are no right or wrong responses. We have modified the wording of some statements, based on feedback received in Round 1. For these statements, we will present the original wording, levels of agreement and graphs, and the modified statement. We will ask you to rate your agreement with the modified statement.

Statements that do not reach consensus following Round 2 will go on to be discussed in the online workshops to decide if they should be included or excluded from the recommendations for a future Scottish care pathway. You will also have the opportunity to leave comments for any of the statements and are free to write anything that you consider relevant. These statements will be analysed and will also inform the online workshops. Finally, there are some questions in this Round that were not asked in Round 1. These are identified throughout the survey and are based on some of the comments received in Round 1. We will ask you to rate your agreement with these new statements in this round.

1. Please enter your unique identifier code below. If you cannot locate it please contact

**Page 2 - Section 1: Pre-operative education**

Pre-operative education is part of routine care for people waiting for THR or TKR surgery. It is often delivered as a face-to-face "joint class" when patients are within 12-weeks of their expected surgery date, and often combined with a pre-operative assessment clinic appointment.

This section asks about pre-operative education for hip/knee replacement surgery.

The statements in this section are principally informed by recommendations for TKR pre-operative education published in 2021 (UK, Anderson et al https://doi.org/10.1186/s12891-021-04160-5), supplemented by a review of pre-operative education content at ten high-volume joint replacement centres (USA, Pitaro et al, 2023 https://doi.org/10.1111/jep.13865), 19 other research studies, and a review of publicly available information on THR and TKR pre-operative education. There are three parts to this section: (i) topics that should be included in pre-operative education; (ii) how pre-operative education should be delivered, and (iii) what exercise types should be recommended as part of pre-operative education.

The statements that reached agreement in the survey are provided below in **purple**. The statements that **did not** reach agreement are provided in **green**. The amended statements, and new statements are also provided in **green**. We are asking you to rate your agreement with all of these **green** statements in this survey. Box graphs (histograms) are also included to provide a breakdown of responses for each statement that **did not** reach agreement. You can find these underneath each statement, as appropriate.

Please read each of the statements below to see which achieved consensus, and to indicate your level of agreement for those that did not reach consensus, and for amended or new statements. Please provide additional comments where you would like to.

**i) Topics that should be included in pre-operative education for THR or TKR surgery**

**The following statements reached at least 70% agreement, and will be included in the recommendations for a future Scottish care pathway:**

**Background Information:**

- Anatomy of the hip/knee joint
- Health conditions that may contribute to needing THR/TKR surgery
- Alternative options to THR/TKR surgery

**Preparing for THR/TKR surgery:**

- Surgical site care (e.g., pre-operative bathing)
- Purpose of pre-operative rehabilitation
- Patient involvement in their own management
- Goal setting
- Using heat and cold for pain relief
- Obtaining and using walking aids and other equipment
- Making home preparations
- Arranging any social support
- Arranging transport to and from the hospital
- Emotional well-being

**Understanding what to expect while in hospital after going home:**

- Education for other people, such as carers
- What to expect during the hospital stay
- Packing list for hospital
- In-hospital timeline
- Visitor information
- Day of surgery logistics
- What a THR/TKR surgical procedure involves
- Information on the prosthesis (replacement)
- Anaesthesia options
- Risks of THR/TKR surgery and how to minimise them
- Common issues that may occur following THR/TKR surgery which do not need to cause alarm
- Pain expectations
- Swelling
- Wound healing
- What to expect following discharge

- Recovery expectations

**Recovering from THR/TKR surgery:**

- Post-operative infection prevention
- Organising help if complications occur
- Pain management
- Nutrition
- Precautions (e.g., movements/activities to avoid in early post-operative period)
- Rehabilitation following THR/TKR
- Returning to daily activities
- Returning to a normal walking pattern
- Returning to driving and other types of travel
- Returning to sports and leisure activities
- Returning to work
- Realistic timelines for recovery and activities
- When to ask for help

**Healthy lifestyle guidance:**

- Physical activity
- Weight management

**The following statements did not reach consensus. Please indicate your level of agreement with these statements.**

Please note: The percentage agreements indicated after each statement represent the sum of people who responded with 'Agree' or 'Strongly Agree.' The full range of responses is shown in the graphs below the statements.

**Preparing for THR/TKR surgery should cover:**

- Optimising management of diabetes (Round 1: Patients 43% agreement; Professionals 81% agreement)
- Identifying and arranging any social care provision (e.g., care & support at home) (Patients 67% agreement; Professionals 94% agreement)

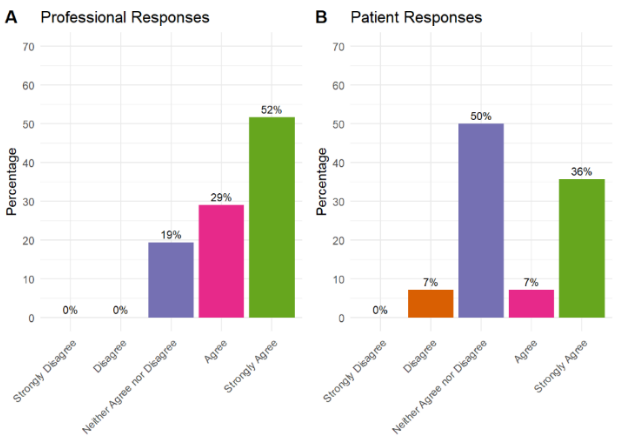

**Histogram: Optimising management of diabetes**

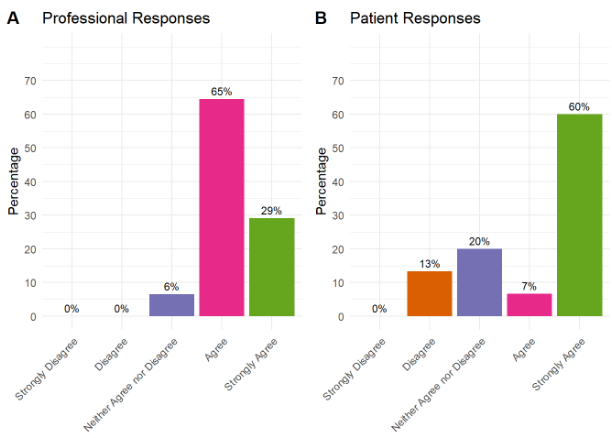

**Histogram: Identifying and arranging any social care provision**

### 2. Healthy lifestyle guidance should cover:

- Stopping smoking (Patients 67%; Professionals 100%)
- Recommended alcohol guidelines and cutting down on drinking (patients 67%; Professionals 94%)

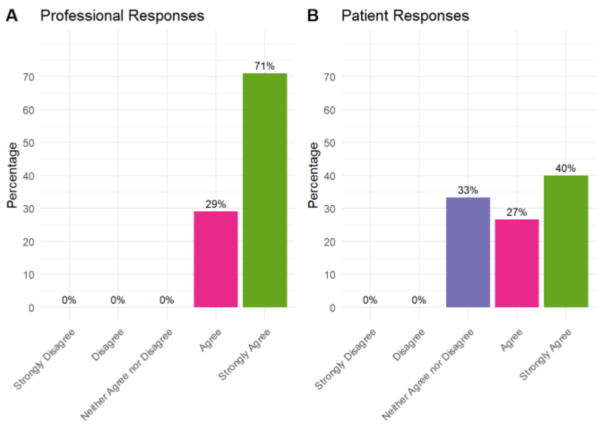

#### Histogram: Stopping smoking

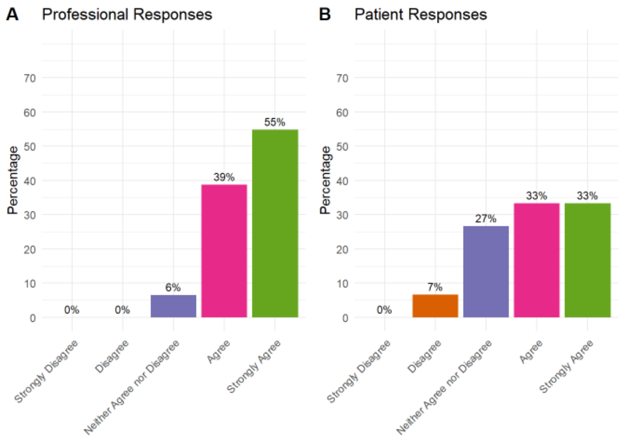

#### Histogram: Recommended alcohol guidelines and cutting down on drinking

**New statement: Based on responses from Round 1 we have developed three new statements. Please indicate your level of agreement with the following statements.**

3. Healthy lifestyle guidance should cover:
   - Mental wellbeing
   - Sleep hygiene
   - Signposting to information and sources of support for healthy lifestyle guidance

#### ii) Delivery of pre-operative education

**The following statements reached at least 70% agreement, and will be included in the recommendations for a future Scottish care pathway:**

**Pre-operative education should:**

- Be informed by a multi-disciplinary team, including members of the orthopaedic surgery team, nursing, physiotherapy, and occupational therapy teams

**Pre-operative information should:**

- Be delivered using a combination of more than one format, including face to face group sessions, a booklet or other written format and a website or other electronic format
- Be delivered through a combination of providing the patient with information and giving them an opportunity to actively take part in tasks
- Provide an opportunity for the patient’s questions to be addressed
- Provide an opportunity for a family member or friend of the patient to be involved
- Be delivered, at least partly, within four weeks of the patient's surgery

**The following statements did not reach consensus. Please indicate your level of agreement with these statements.**

**Please note: The percentage agreements indicated after each statement represent the sum of people who responded with 'Agree' or 'Strongly Agree.' The full range of responses is shown in the graphs below the statements.**

4. **Pre-operative education should:**
   - Be informed by patients who have previously had THR/TKR surgery (Patients 53%; Professionals 61%)

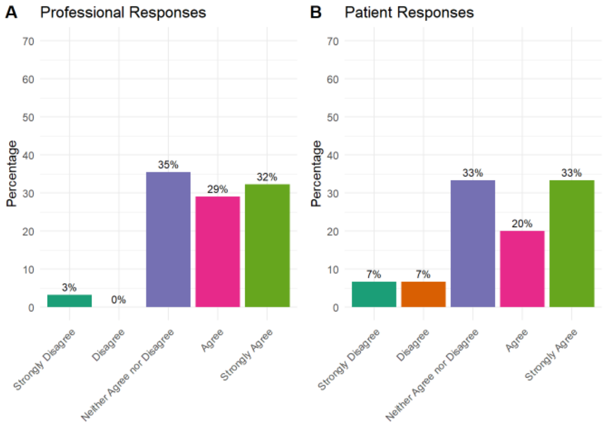

**Histogram: Be informed by patients who have previously had THR/TKR surgery**

5. **Pre-operative information should:**
   - Be delivered, at least partly, by providing examples of other patients' experiences of THR/TKR surgery (Patients 73%; Professionals 65%)
   - Be delivered 1-1 if the patient does not wish to attend a group session (Patients 61%; Professionals 61%)
   - Be provided separately for THR and TKR – i.e., patients waiting for THR should receive group education separately from patients waiting for TKR (Patients 93%; Professionals 48%)

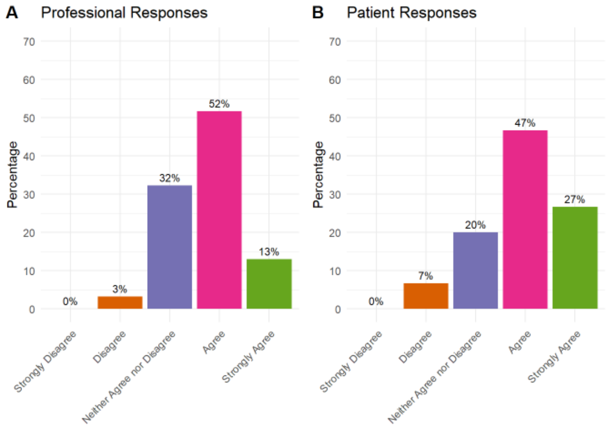

**Histogram: Be delivered, at least partly, by providing examples of other patients' experiences of THR/TKR surgery**

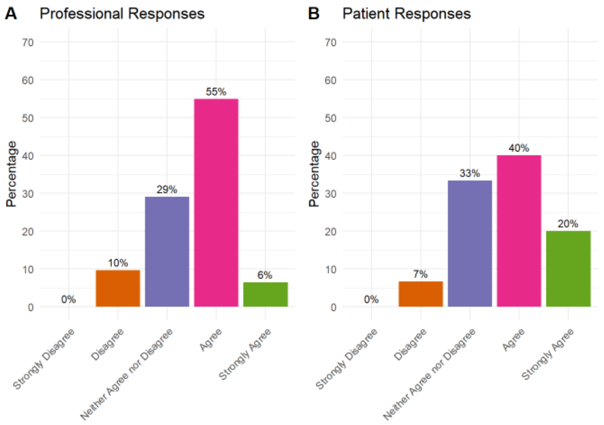

**Histogram: Be delivered 1-1 if the patient does not wish to attend a group session**

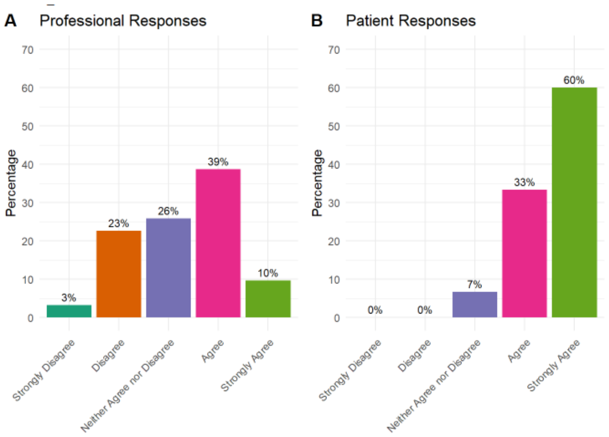

**Histogram: Be provided separately for THR and TKR**

**New statement: Based on responses from Round 1 we have developed a new statement. Please indicate your level of agreement with the following statement.**

**6. Pre-operative information should:**

- Make use of digital technologies such as video, interactive websites and/or virtual meetings

**iii) Pre-operative exercise types**

The following statements reached at least 70% agreement, and will be included in the recommendations for a future Scottish care pathway

A pre-operative exercise programme should include:

- Leg strengthening exercises
- Leg flexibility exercises
- Balance exercises

- Functional movement exercises - i.e., sit to stand exercises
- Cardiovascular exercises
- Practicing post operative exercises
- Delivery should be tailored according to each patient’s individual needs
- Delivery should be standardised across Scotland

**The following statements did not reach consensus. Please indicate your level of agreement with these statements.**

Please note: The percentage agreements indicated after each statement represent the sum of people who responded with 'Agree' or 'Strongly Agree.' The full range of responses is shown in the graphs below the statements.

7. **A pre-operative exercise programme should include:**
    - Core control exercises Patients 87%; Professionals 62%)
    - Walking practice with walking aids (Patients 67%; Professionals 67%)
    - Training on steps (patients 67%; Professionals 68%)

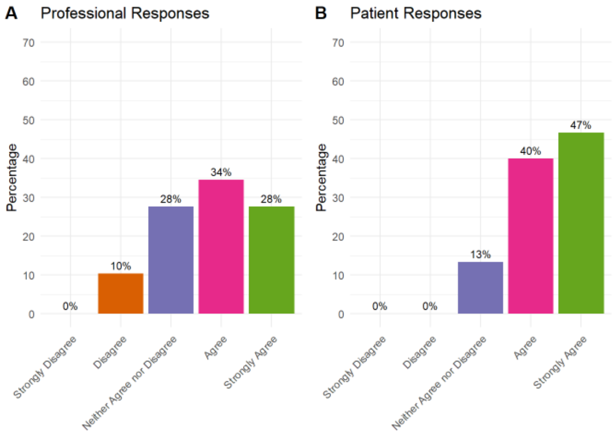

**Histogram: Core control exercises**

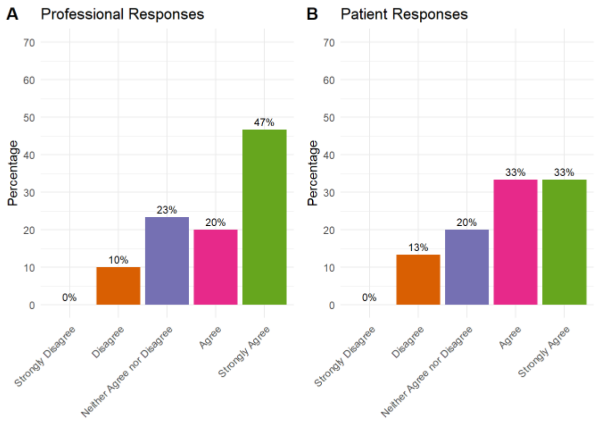

**Histogram: Walking practice with walking aids**

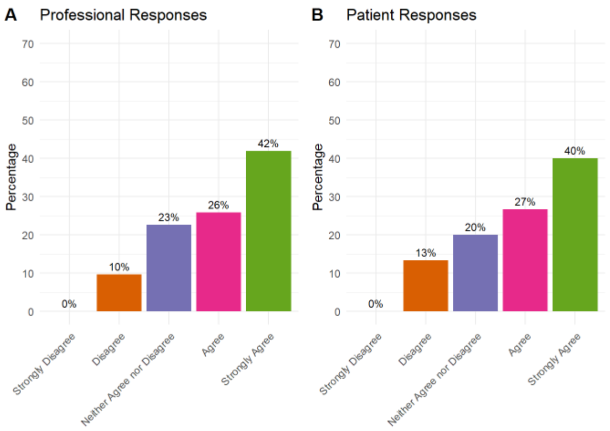

**Histogram: Training on steps**

**8. Please make any comments about pre-operative education here:**

**Page 2: Patient optimisation**

**Section 2: Patient optimisation**

Patient optimisation is the process of supporting patients to be as healthy as possible before surgery and address any health problems that may affect this (NHS England » Earlier screening, risk assessment and health optimisation in perioperative pathways: guide for providers and integrated care boards). This can include the management of long-term health conditions and support people to make changes to improve their health (The following recommendations have been informed by the ERAS Society (Enhanced recovery after Surgery®) publication (Wainwright et al [2020] https://doi.org/10.1080/17453674.2019.1683790).

Please read each of the statements below to see which achieved consensus, and to indicate your level of agreement for those that did not reach consensus, and for amended or new statements. Please provide additional comments where you would like to.

**The following statements reached at least 70% agreement, and will be included in the recommendations for a future Scottish care pathway:**

**Smoking cessation:**

- Patients who identify as smokers should be referred to a smoking cessation programme while they are waiting for surgery

- Referral to smoking cessation programmes should be made as soon as patients are placed on the waiting list for surgery
- Referral should be made at the same time as pre-operative education/assessment (typically within12-weeks of surgery)

**Alcohol cessation:**

- Patients waiting for THR or TKR who report high alcohol intake/alcohol misuse should be referred to an alcohol cessation programme

**Pre-operative anaemia:**

- Pre-operative anaemia should be investigated and corrected prior to THR/TKR surgery

**Pre-operative fasting:**

- Patients should be advised to continue the intake of clear fluids until 2-hours before the start of anaesthesia, and to fast (solid foods) for 6-hours before the start of anaesthesia for THR/TKR

**The following statements about smoking cessation did not reach consensus. Please indicate your level of agreement with these statements.**

Please note: The percentage agreements indicated after each statement represent the sum of people who responded with 'Agree' or 'Strongly Agree.' The full range of responses is shown in the graphs below the statements.

9. **Smoking cessation:**
   - Referral should be made at the same time as pre-operative education/assessment (typically within 12-weeks of surgery) (Patients 54%; Professionals 33%)

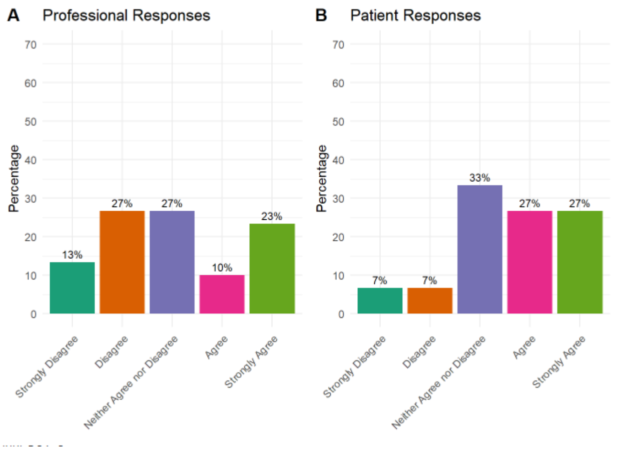

**Histogram: Referral should be made at the same time as pre-operative education/assessment**

**The following statement has been modified based on comments received from Round 1.**

**The original statement was:**

- Patients who **identify as smokers** should be **referred** to a smoking cessation programme while they are waiting for surgery

**This statement did reach consensus, but we have modified the wording based on Round 1 comments. Please read the statement below and indicate your level of agreement.**

***10. Smoking cessation (modified version)***

- Patients who are smokers should be offered a referral to a smoking cessation programme at the earliest opportunity by any appropriate healthcare professional that they interact with

The following statements about alcohol cessation did not reach consensus. Please indicate your level of agreement with these statements.

Please note: The percentage agreements indicated after each statement represent the sum of people who responded with 'Agree' or 'Strongly Agree.' The full range of responses is shown in the graphs below the statements.

11. ***Alcohol cessation* programmes are recommended before THR or TKR for patients who misuse alcohol or report high alcohol intake:**
    - Referral to an alcohol cessation programme should be made as soon as patients are placed on the waiting list (patients 67%; Professionals 68%)
    - Referral to an alcohol cessation programme should be made at the same time as pre-operative education/assessment (typically within 12-weeks of surgery) (Patients 53%; Professionals 23%)

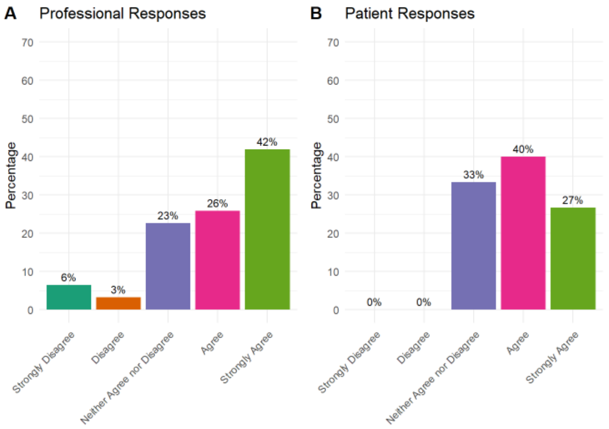

**Histogram: Referral to an alcohol cessation programme should be made as soon as patients are placed on the waiting list**

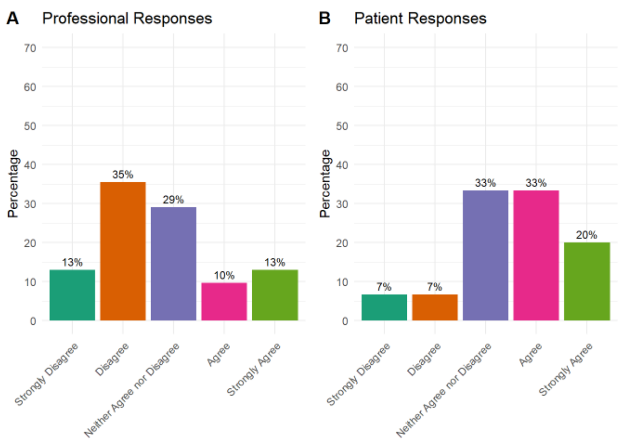

**Histogram: Referral to an alcohol cessation programme should be made at the same time as pre-operative education/assessment (typically within 12-weeks of surgery)**

**The following statement has been modified based on comments received from Round 1.**

The original statements were:

- Referral to an alcohol cessation programme should be made as soon as patients are placed on the waiting list (patients 67%; Professionals 68%)
- Referral to an alcohol cessation programme should be made at the same time as pre-operative education/assessment (typically within 12-weeks of surgery) (Patients 53%; Professionals 23%)

**Please read the new statement below and indicate your level of agreement.**

**12. *Alcohol cessation (modified version):***

- Patients who report high alcohol intake should be offered a referral to an alcohol cessation programme at the earliest opportunity by any appropriate healthcare professional that they interact with

**The following statements about pre-operative anaemia did not reach consensus. Please indicate your level of agreement with these statements.**

Please note: The percentage agreements indicated after each statement represent the sum of people who responded with 'Agree' or 'Strongly Agree.' The full range of responses is shown in the graphs below the statements.

13. ***Pre-operative anaemia (lower than normal amount of healthy red blood cells)***

- Pre-operative anaemia should be identified and corrected at the pre-operative assessment (usually 12-weeks before surgery) for THR/TKR (Patients 73%; Professionals 64%)

- Pre-operative anaemia should be identified, investigated and corrected as soon as patients are placed on the waiting list for THR/TKR (Patients 74%; Professionals 67%)

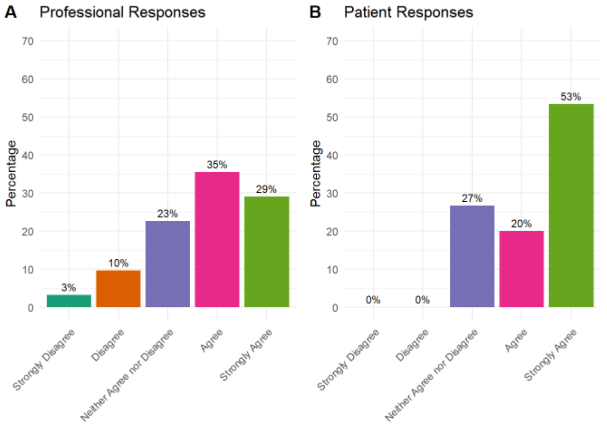

**Histogram: Pre-operative anaemia should be identified and corrected at the pre-operative assessment**

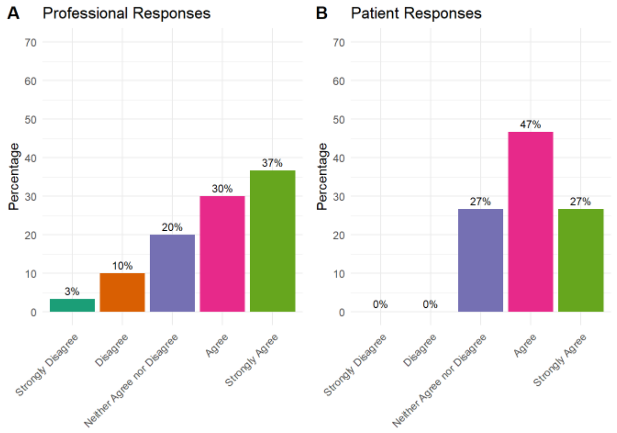

**Histogram: Pre-operative anaemia should be identified, investigated and corrected as soon as patients are placed on the waiting list for THR/TKR**

**The following Statement has been developed based on comments from Round 1.**

**The original statements were:**

- Pre-operative anaemia should be identified and corrected at the pre-operative assessment (usually 12-weeks before surgery) for THR/TKR
- Pre-operative anaemia should be investigated and corrected prior to THR/TKR surgery
- Pre-operative anaemia should be identified, investigated and corrected as soon as patients are placed on the waiting list for THR/TKR

**Please read the new statement below and indicate your level of agreement.**

**14. *Pre-operative anaemia (lower than normal amount of healthy red blood cells):***

- Pre-operative anaemia should be corrected as soon as it is identified, at any time in the patient's pre-operative journey

**The following statements about patient optimisation did not reach consensus. Please indicate your level of agreement with these statements.**

Please note: The percentage agreements indicated after each statement represent the sum of people who responded with 'Agree' or 'Strongly Agree.' The full range of responses is shown in the graphs below the statements.

15. ***Pre-operative fasting: Intake of clear fluids until 2 hours before surgery and a 6 hour fast for solid food before surgery for TKR/THR:***

- Patients should be advised to continue the intake of clear fluids until 2-hours before the start of anesthesia, and to fast (solid foods) for 6-hours before the start of anesthesia for THR/TKR (Patients 87%; Professionals 58%)

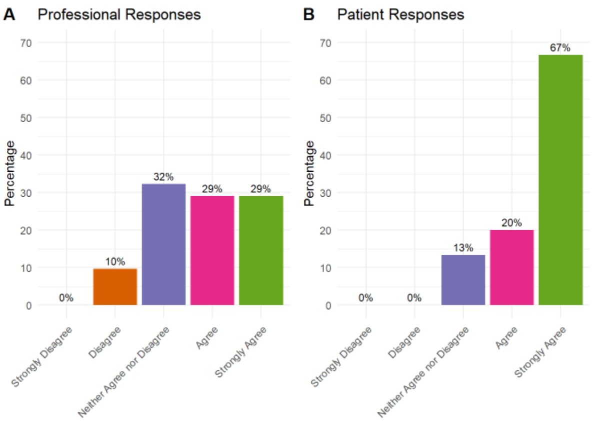

**Histogram: Patients should be advised to continue the intake of clear fluids until 2-hours before the start of anaesthesia, and to fast (solid foods) for 6-hours before the start of anaesthesia for THR/TKR**

**16. *Pre-operative carbohydrate loading: in THR/TKR carbohydrate loading may improve patient well-being and metabolism, but has not been shown to accelerate the achievement of discharge criteria or reduce complications:***

- Pre-operative carbohydrate loading should not be recommended prior to THR/TKR (Patients 7%; Professionals 16%)

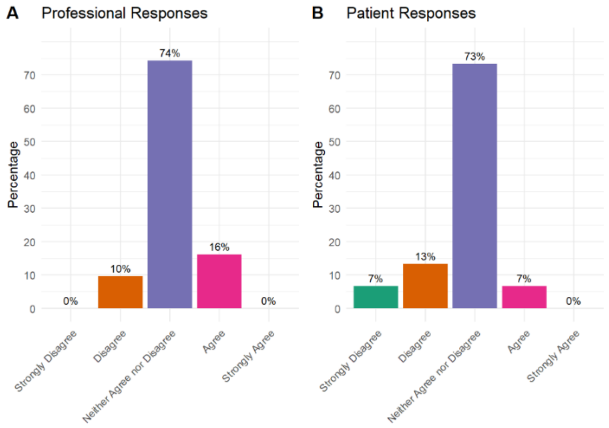

**Histogram: Pre-operative carbohydrate loading should not be recommended prior to THR/TKR**

17. ***Pre-operative physiotherapy: current evidence does not support preoperative physiotherapy as an essential intervention:***
   - Pre-operative physiotherapy is not essential for patients waiting for THR/TKR (Patients 13%; Professional 26%)

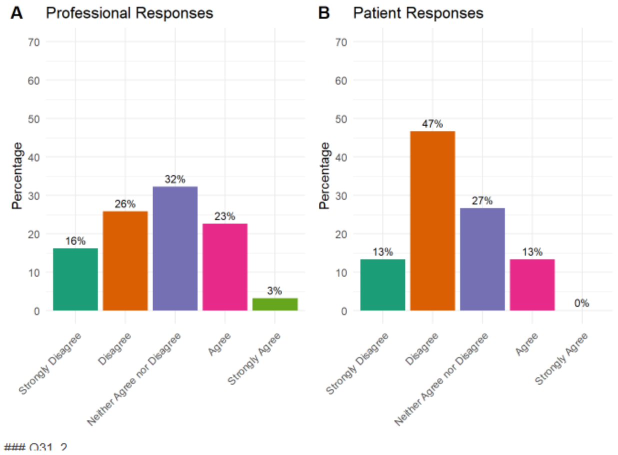

**Histogram: Pre-operative physiotherapy is not essential for patients waiting for THR/TKR**

### Pre-operative physiotherapy

The following question has been developed based on Round 1 comments.

**Please read the new statement below and indicate your level of agreement.**

18. ***Pre-operative physiotherapy:***
   - Referral to physiotherapy should be considered on an individual basis, depending on the patient's needs. NOTE: Physiotherapy in this context is outside of/in addition to any physiotherapy input received at pre-operative education/joint class

**19. Please make any comments on patient optimisation here:**

**Page 4: Section 3: Other interventions to support people waiting for THR and TKR surgery**

**The following statements reached at least 70% agreement, and will be included in the recommendations for a future Scottish care pathway:**

**Prehabilitation:**

- Prehabilitation should be offered to people waiting for THR/TKR
- Prehabilitation should be offered to people when they are placed on the waiting list for THR/TKR

**Psychologically informed interventions**

- Opioid counselling should be offered to patients on the waiting list for THR/TKR surgery who are routinely taking prescription opioids

**Weight management**

- Weight management interventions should be offered to people waiting for THR/TKR
- Weight management interventions should be offered to people when they are placed on the waiting list for THR/TKR

**The following statements did not reach consensus. Please indicate your level of agreement with these statements.**

Please note: The percentage agreements indicated after each statement represent the sum of people who responded with 'Agree' or 'Strongly Agree.' The full range of responses is shown in the graphs below the statements.

20. **Prehabilitation** can be defined as a formal pre-operative programme, aimed at enabling the patient to withstand the stress of surgery. Prehabilitation for THR/TKR may combine exercise with other interventions (e.g., education, information provision).

- Prehabilitation should be offered within 12-weeks of THR/TKR surgery (Patients 93%; Professionals 58%)

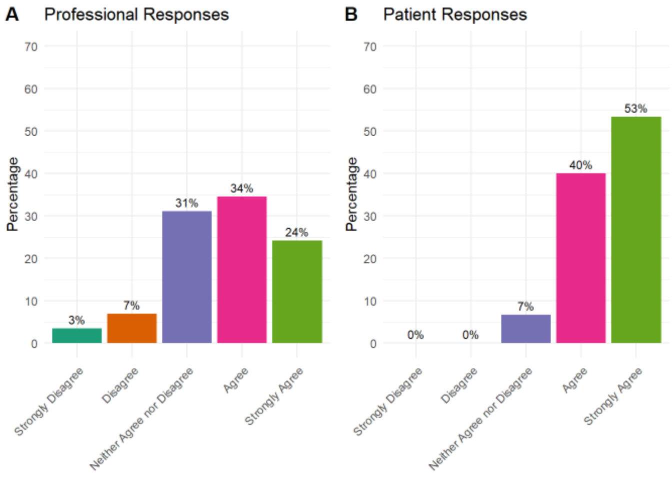

**Histogram: Prehabilitation should be offered within 12-weeks of THR/TKR surgery**

### 21. Psychologically-informed interventions:

- Psychologically informed interventions should be offered to people waiting for THR/TKR (Patients 40%; Professionals 42%)
- Psychologically informed Interventions should be offered within 12-weeks of THR/TKR surgery (Patients 46%; Professionals 36%)
- Patients waiting for THR/TKR surgery who have been formally diagnosed with anxiety or depression should be offered referral to cognitive behavioural therapy (CBT) based therapy (Patients 60%; Professionals 42%)
- Mindfulness should be considered for people waiting for THR/TKR to help with post-operative pain and opioid use (Mindfulness is about living more in the present moment, appreciating the here and now, and not dwelling too much on the past or future) (Patients 60%; Professionals 51%)

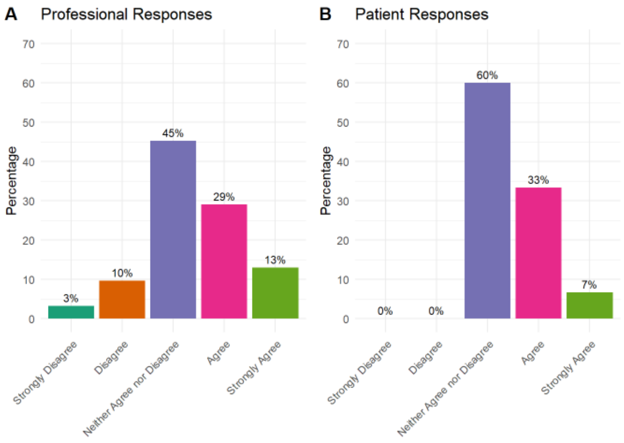

**Histogram: Psychological informed interventions should be offered to people waiting for THR/TKR**

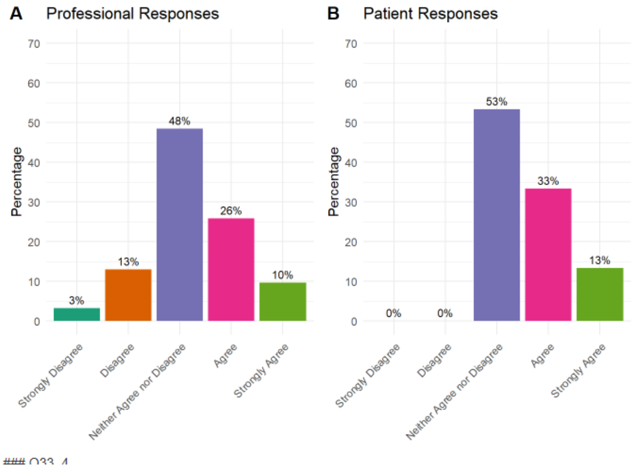

**Histogram: Psychologically informed Interventions should be offered within 12-weeks of THR/TKR surgery**

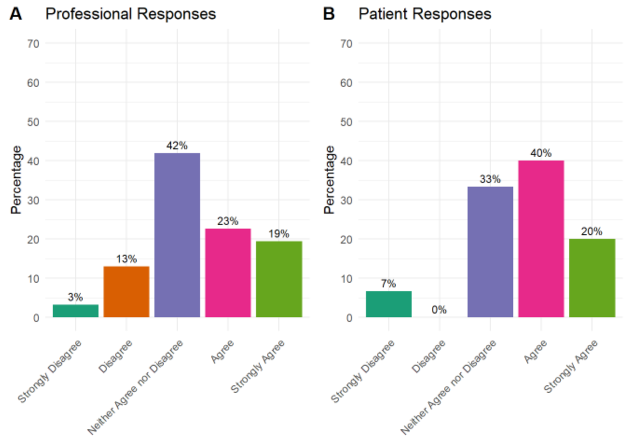

**Histogram: Patients waiting for THR/TKR surgery who have been formally diagnosed with anxiety or depression should be offered referral to cognitive behavioural therapy (CBT) based therapy**

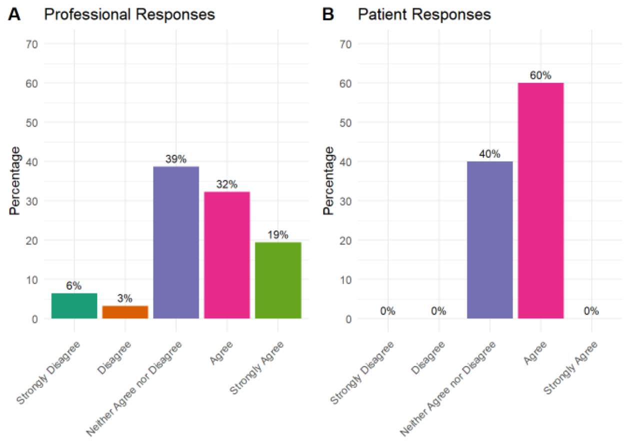

**Histogram: Mindfulness should be considered for people waiting for THR/TKR to help with post-operative pain and opioid use**

**The following question has been developed based on Round 1 comments. Please read the question and indicate your level of agreement.**

**22. Psychologically-informed interventions:**

- Psychologically informed interventions should be considered on an individual basis, depending on the patient's needs

**The following statements did not reach consensus. Please indicate your level of agreement with these statements.**

Please note: The percentage agreements indicated after each statement represent the sum of people who responded with 'Agree' or 'Strongly Agree.' The full range of responses is shown in the graphs below the statements.

**23. Weight management**

- Weight management interventions should be offered within 12-weeks of THR/TKR surgery (Patients 40%; Professionals 46%)
- Patients waiting for THR/TKR surgery who have a body mass index (BMI) of 27 $kg/m^2$ or over should be offered referral to a weight management programme. NB a BMI of 25 is considered overweight and over 30 is considered obese (Patients 73%; Professionals 49%)

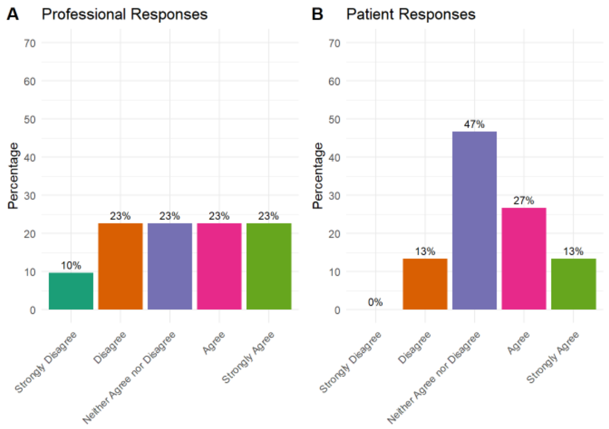

**Histogram: Weight management interventions should be offered within 12-weeks of THR/TKR surgery**

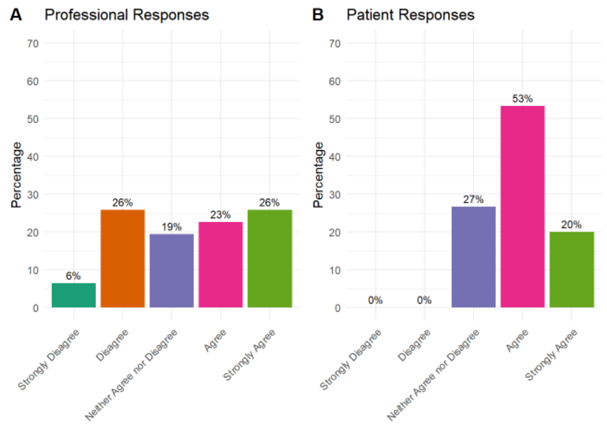

**Histogram: Patients waiting for THR/TKR surgery who have a body mass index (BMI) of 27 kg/m$^2$ or over should be offered referral to a weight management programme.**

**The following question has been developed based on Round 1 comments. Please read the question and indicate your level of agreement.**

**24. Weight management:**

- Patients should be offered referral to a weight management programme if their weight is considered a risk factor for poor surgical outcome

**25. Please make any comments on other interventions to support people waiting for THR/TKR here:**

**Page 4: Waiting a long time for surgery**

In Scotland, waiting times for THR and TKR are at a record high. Interventions such as pre-operative education and prehabilitation are typically offered close to the expected date of surgery, meaning that other interventions may be required to support people while they wait. The following statements relate to patients waiting for longer than the 18-weeks referral to treatment target.

**The following statements reached at least 70% agreement, and will be included in the recommendations for a future Scottish care pathway:**

**Signposting to web-based information:**

- People waiting for THR/TKR surgery should be signposted to web-based information about physical activity and fitness
- People waiting for THR/TKR surgery should be signposted to web-based information about diet and nutrition
- People waiting for THR/TKR surgery should be signposted to web-based information about mental wellbeing

- People waiting for THR/TKR surgery should be signposted to web-based information about pain management
- People waiting for THR/TKR surgery should be signposted to web-based information about smoking cessation
- People waiting for THR/TKR surgery should be signposted to web-based information about alcohol reduction

**Written information:**

- People waiting for THR/TKR surgery should be sent written information if they prefer it to web-based materials

**Signposting to support:**

- People waiting for THR/TKR surgery should be offered interventions that support them to maintain/increase physical activity levels while they are waiting for surgery
- If required people waiting for THR/TKR surgery should be offered referral to relevant social care services

**How and when to update people waiting:**

- People waiting for THR or TKR surgery should be kept up to date every 3-months about where they are on the waiting list and what they can be doing to support themselves
- People on the waiting list for THR or TKR surgery should be able to request a review if they feel their condition is deteriorating

**Waiting well consultation:**

- People waiting for THR and TKR surgery should be offered at least one 'waiting well' telephone consultation to provide wellbeing advice, signposting, encourage vaccination & screening uptake, and referral to relevant health and social care

**Timing for prehabilitation or pre-operative education:**

- A prehabilitation refresher should happen 12 weeks before surgery at the pre-operative assessment services

**Social care:**

- Social care and needs that are not currently being addressed should be identified when patients are placed on a waiting list for THR/TKR surgery

**The following statements did not reach consensus. Please indicate your level of agreement with these statements.**

Please note: The percentage agreements indicated after each statement represent the sum of people who responded with 'Agree' or 'Strongly Agree.' The full range of responses is shown in the graphs below the statements.

26. **Signposting to support:**

- People waiting for THR/TKR surgery should be signposted to third sector organisations (Patients 64%; Professionals 77%)

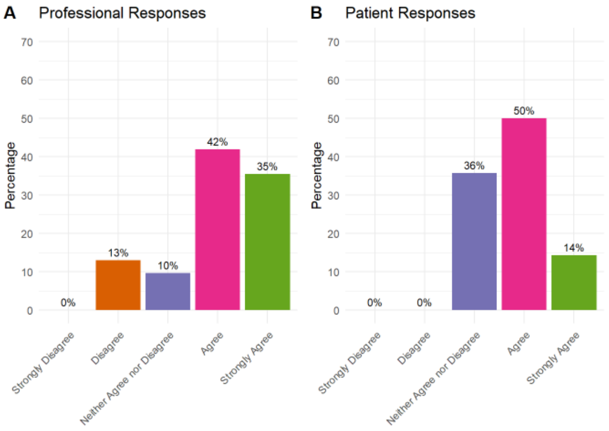

**Histogram: People waiting for THR/TKR surgery should be signposted to third sector organisations**

27. **'Waiting well' consultation:**

- People waiting for THR and TKR surgery should be offered at least one 'waiting well' telephone consultation to provide wellbeing advice, signposting, encourage vaccination & screening uptake, and referral to relevant health and social care services (Patients 73%; Professionals 67%)

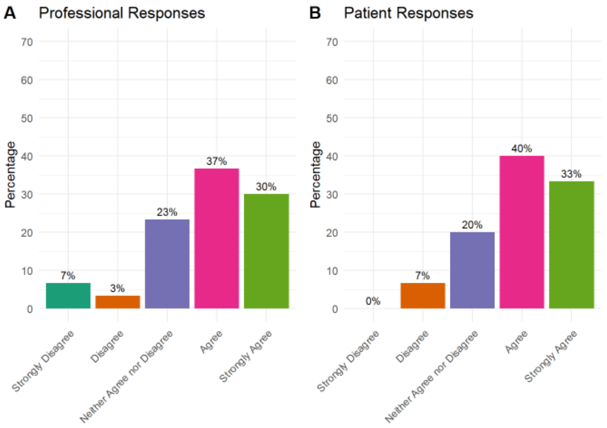

**Histogram: People waiting for THR and TKR surgery should be offered at least one 'waiting well' telephone consultation to provide wellbeing advice, signposting, encourage vaccination & screening uptake, and referral to relevant health and social care services**

28. **Timing for prehabilitation or pre-operative education:**

- Prehabilitation should start when people are placed on the waiting list for THR or TKR surgery rather than waiting until 12-weeks before surgery (Patients 60%; Professionals 74%)
- A form of pre-operative education/joint school should take place when patients are placed on the waiting list for THR or TKR rather than waiting until 12-weeks before surgery (Patients 53%; Professionals 66%)

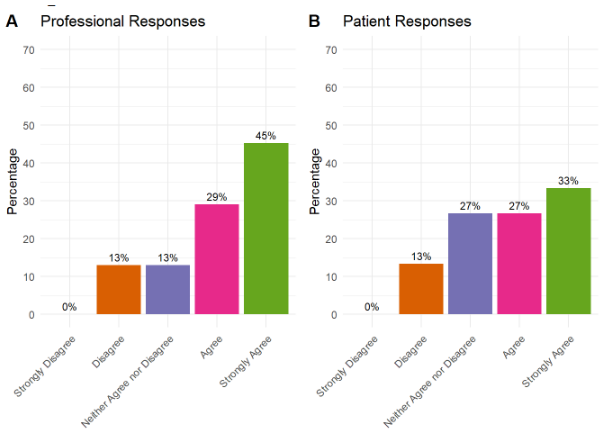

**Histogram: Prehabilitation should start when people are placed on the waiting list for THR or TKR surgery rather than waiting until 12-weeks before surgery**

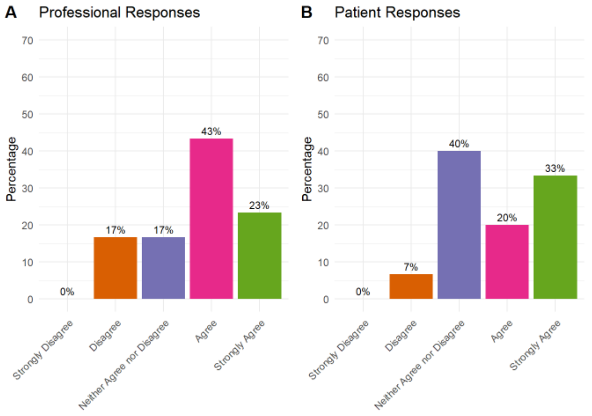

**Histogram: A form of pre-operative education/joint school should take place when patients are placed on the waiting list for THR or TKR rather than waiting until 12-weeks before surgery**

#### 29. Social care:

- People waiting for THR/TKR should be reassessed for social care needs at the preoperative assessment (Patients 87%; Professionals 69%)

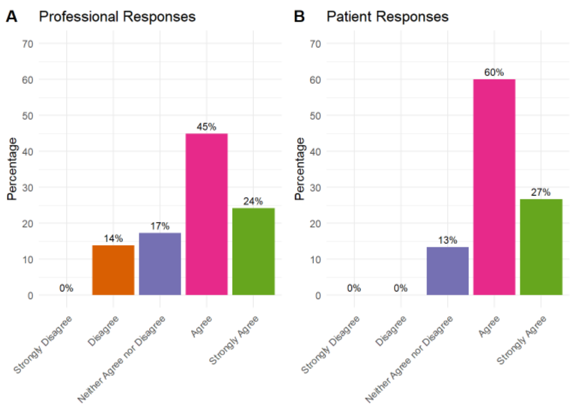

**Histogram: People waiting for THR/TKR should be reassessed for social care needs at the preoperative assessments**

**30. Please make any comments about waiting a long time for THR/TKR surgery here:**

**Page 6: Workshops**

We would like to invite you to a workshop on Microsoft Teams to establish the final consensus for the Scottish care pathway. There will be one workshop for experts and a separate workshop for people with lived experience of THR/TKR.

**Please select your availability from the options below (tick all that apply)**

- Tuesday 4th February - 12pm - 1.30pm
- Tuesday 4th February - 7pm - 8.30pm
- Thursday 20th February - 12pm - 1.30pm
- Thursday 20th February - 7pm - 8.30pm
- I am unable to attend/do not wish to attend

**Final Page**

Thank you for taking part in this survey. We appreciate the time you have taken to do this and the information you have provided will be very useful.

We will collect all the responses and analyse the findings together. Statements that do not reach agreement following this round will be discussed in the workshops planned for early 2025

If you have any questions or comments, or do not want to take part in the next round please contact

### Round 3: Workshop discussions

Survey results from Rounds 1 and 2 were presented to participants in the round 3 Workshop. The following plots were presented to participants on items that did not reach consensus and required ongoing discussion

#### Section 1: Pre-operative education

| | |
|---|---|
| Optimising management of diabetes: | 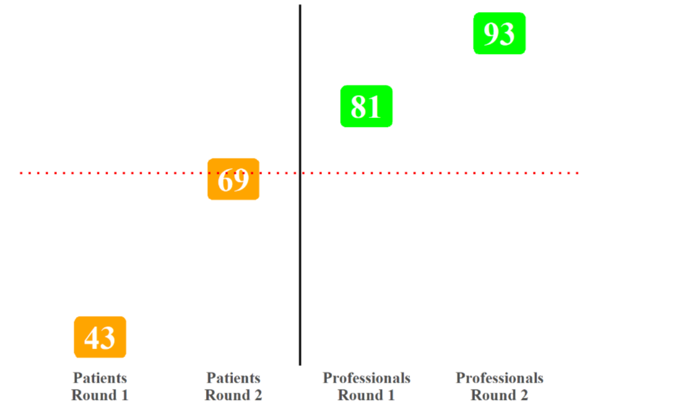 |
| Healthy lifestyle guidance should cover sleep hygiene: | 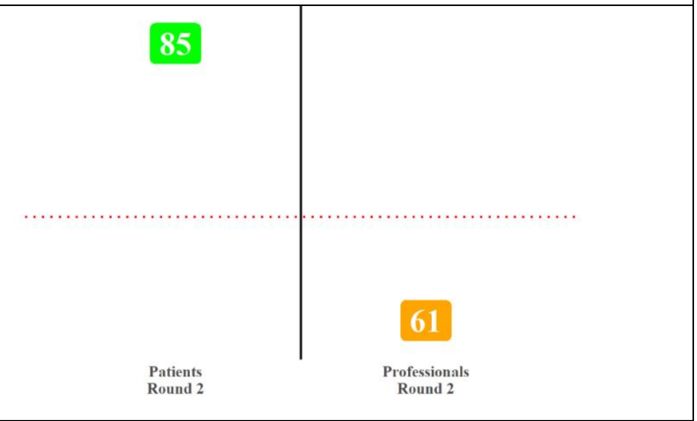 |
| Pre-operative education should be informed by patients who have previously had surgery: | 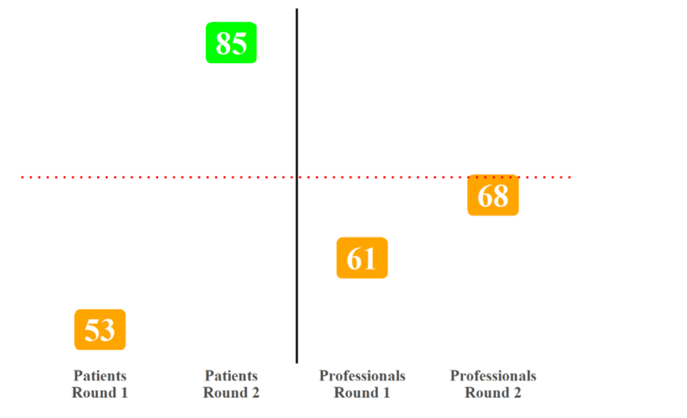 |

Preoperative education should Be delivered 1-1 if the patient does not wish to attend a group session:

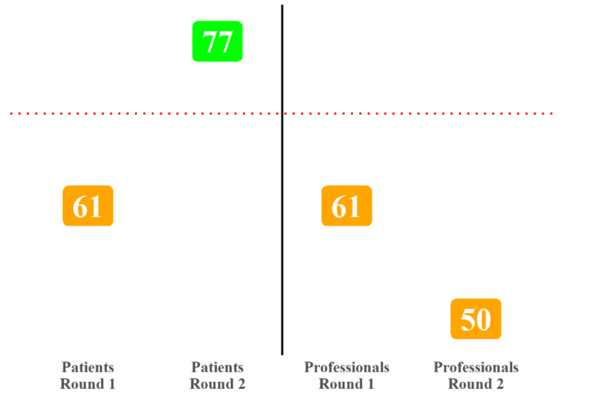

Pre-operative education should be provided separately for THR and TKR

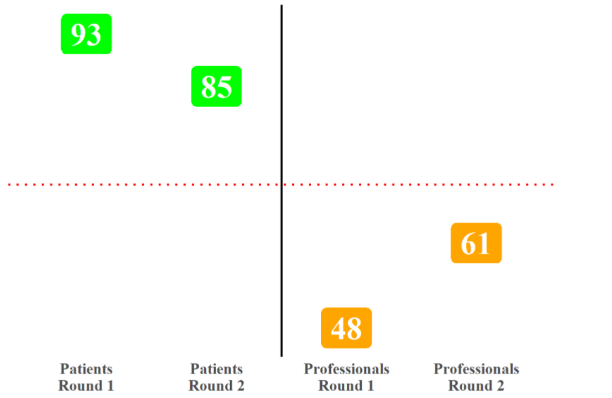

A pre-operative exercise programme should include core control exercises

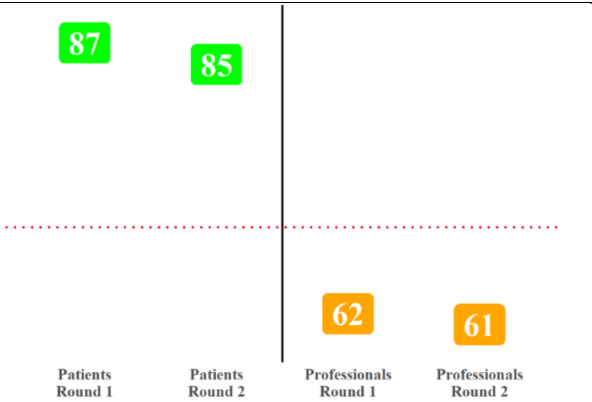

A pre-operative exercise programme should include walking practice with walking aids:

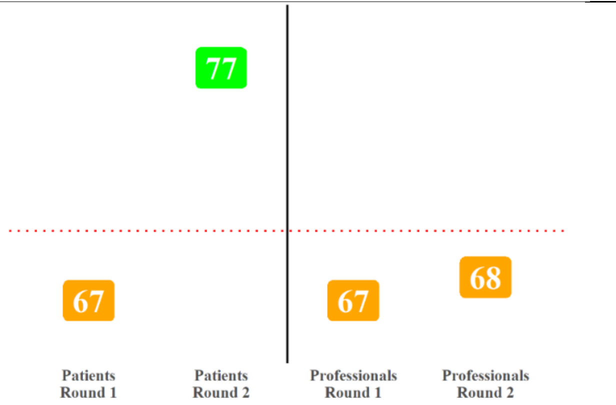

A pre-operative exercise programme should include training on steps:

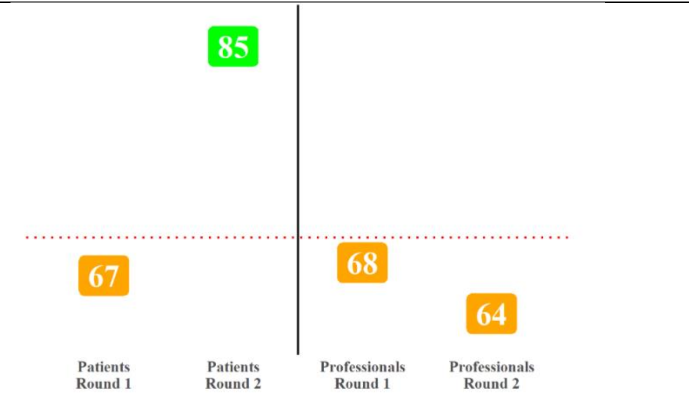

### Section 2: Patient Optimisation

Referral should be made at the same time as pre-operative education/assessment (typically within 12-weeks of surgery)

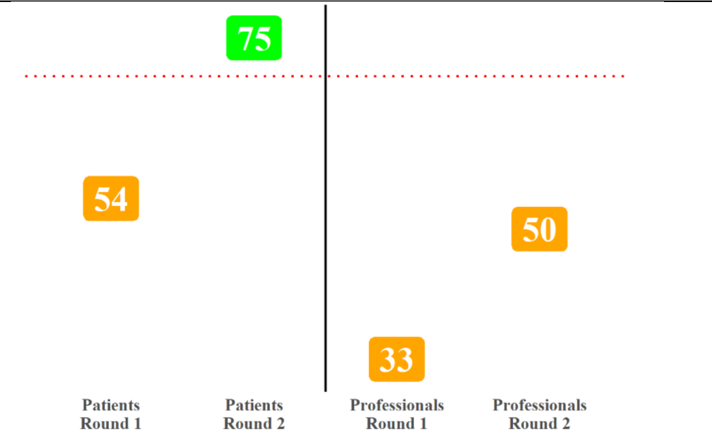

Referral to an alcohol cessation programme should be made at the same time as pre-operative education/assessment (typically within 12-weeks of surgery)

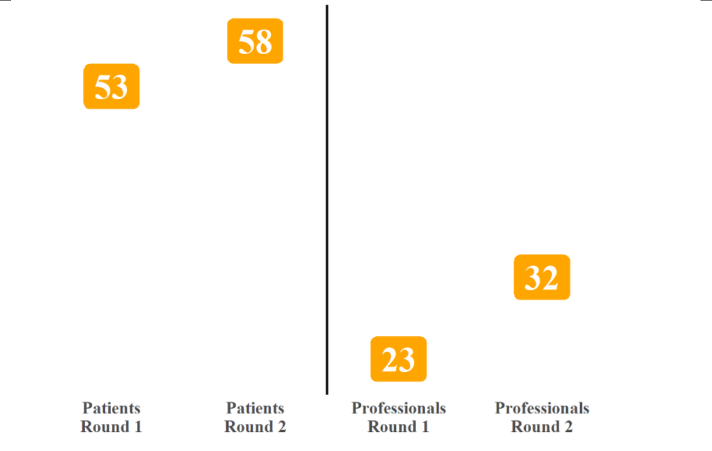

<table>
<tr><td>Advice to continue intake of clear fluids until 2-hours before anaesthesia and to fast for 6-hours before anaesthesia</td><td>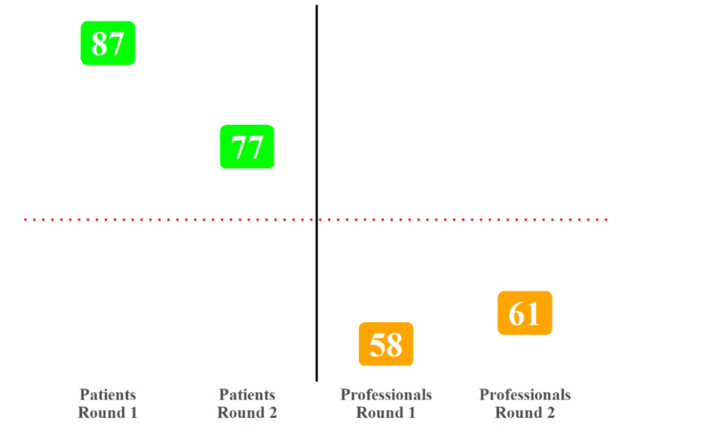
</td></tr>
<tr><td>Pre-operative carbohydrate loading should not be recommended prior to surgery</td><td>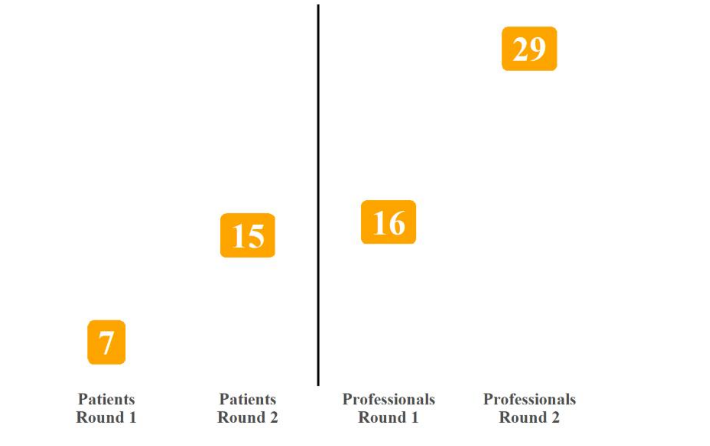
</td></tr>
</table>

**Section 3: Other interventions to support people waiting for THR and TKR Surgery**

<table>
<tr><td>Prehabilitation should be offered within 12-weeks of surgery</td><td>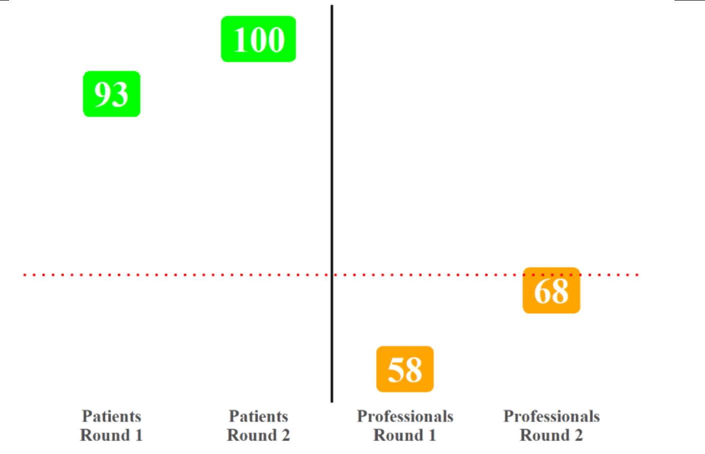
</td></tr>
</table>

| | |
|---|---|
| Psychologically informed interventions Should be offered | 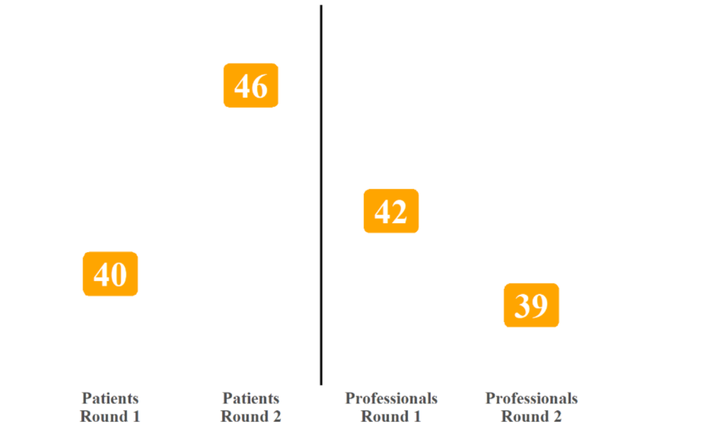  |
| Psychologically informed interventions should be offered within 12-weeks of surgery | 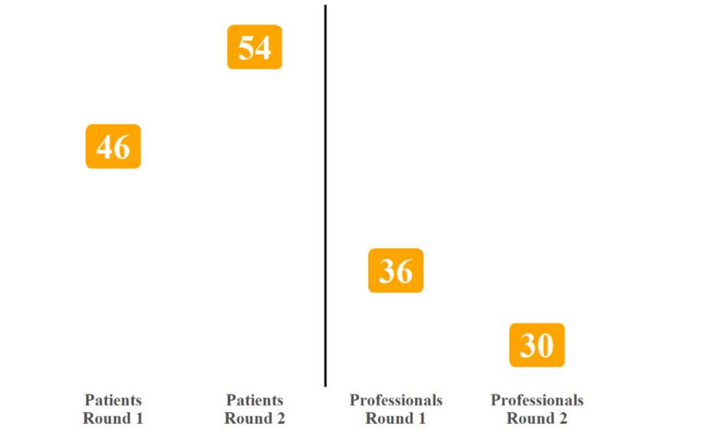  |
| Patients diagnosed with anxiety or depression should be offered CBT | 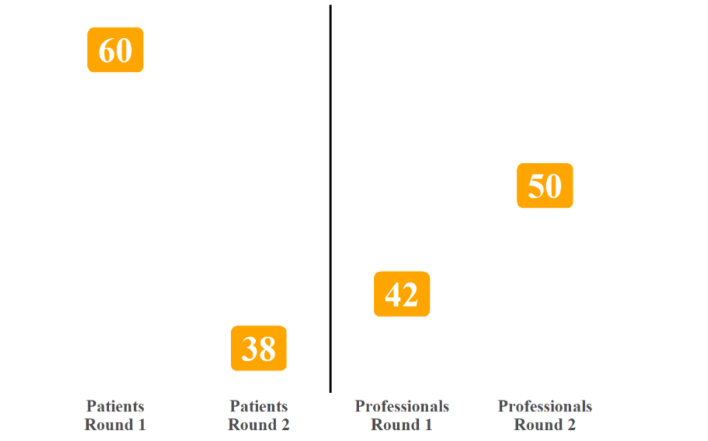  |
| Mindfulness should be considered to help with post-operative pain and opioid use | 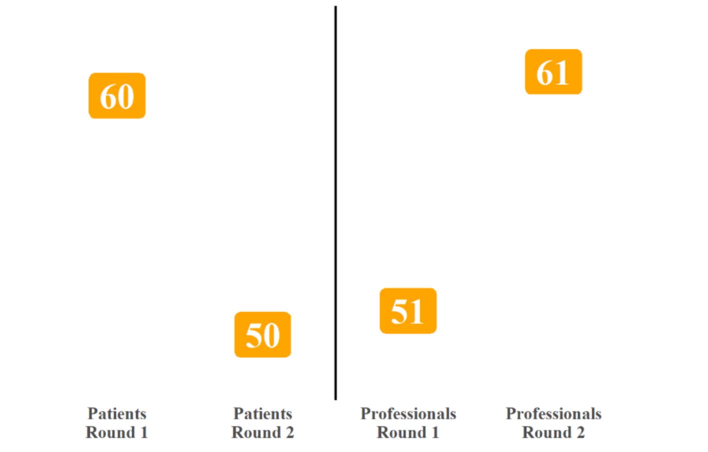  |

| | |
|---|---|
| Psychologically informed interventions should be considered on an individual basis and the patient's needs | 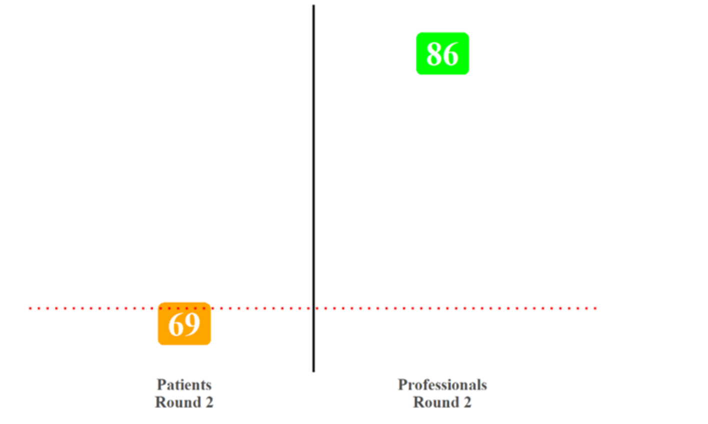 |
| Weight management interventions should be offered within 12-weeks of surgery | 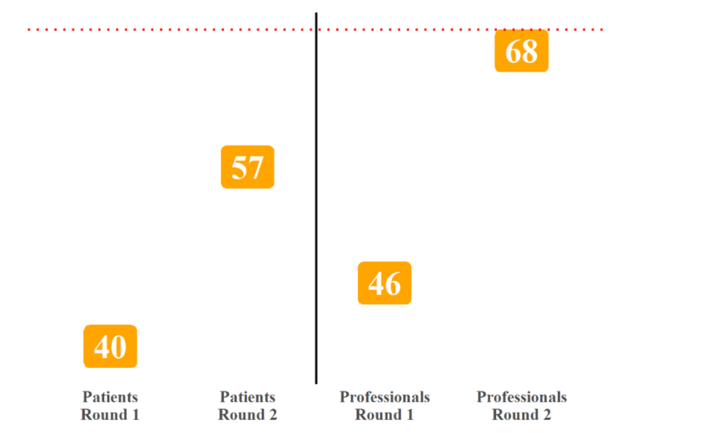 |

**Section 4: Waiting a long time for Surgery**

| | |
|---|---|
| People waiting for surgery should be signposted to third sector organisations | 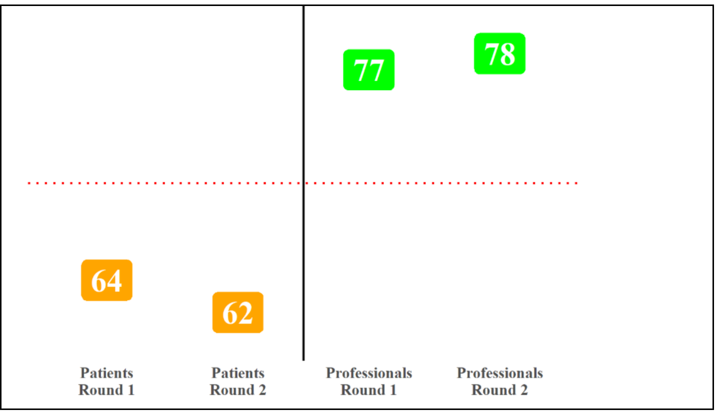 |
