## Supplementary File 2 for "Consensus recommendations for supporting people waiting for total hip and knee arthroplasty in Scotland: A modified Delphi study"

2026

### Sections

#### Pre-operative education

##### Round

Round 1

### Background information

Background information: Anatomy of joint

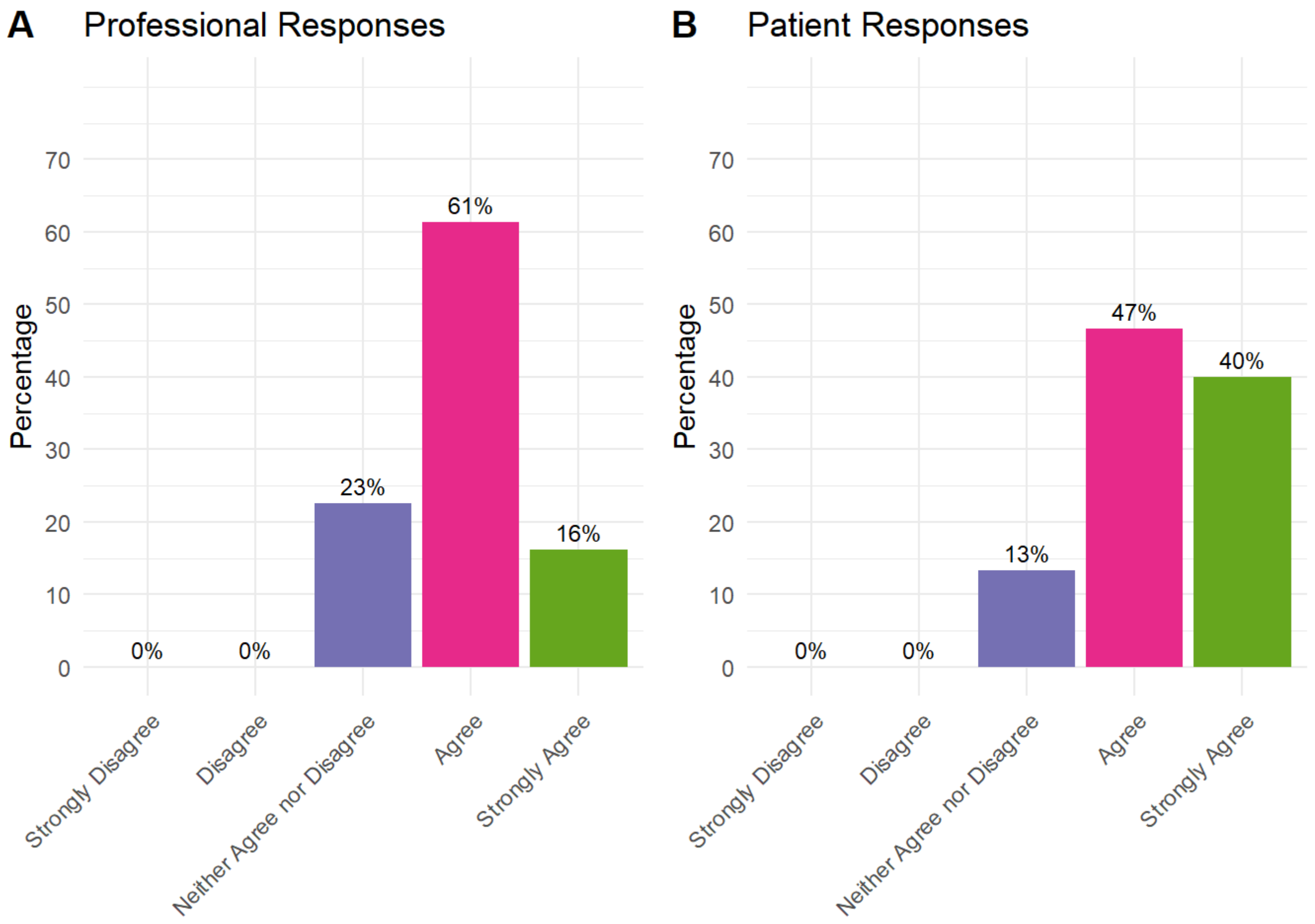


Background information: Conditions contributing to surgery need

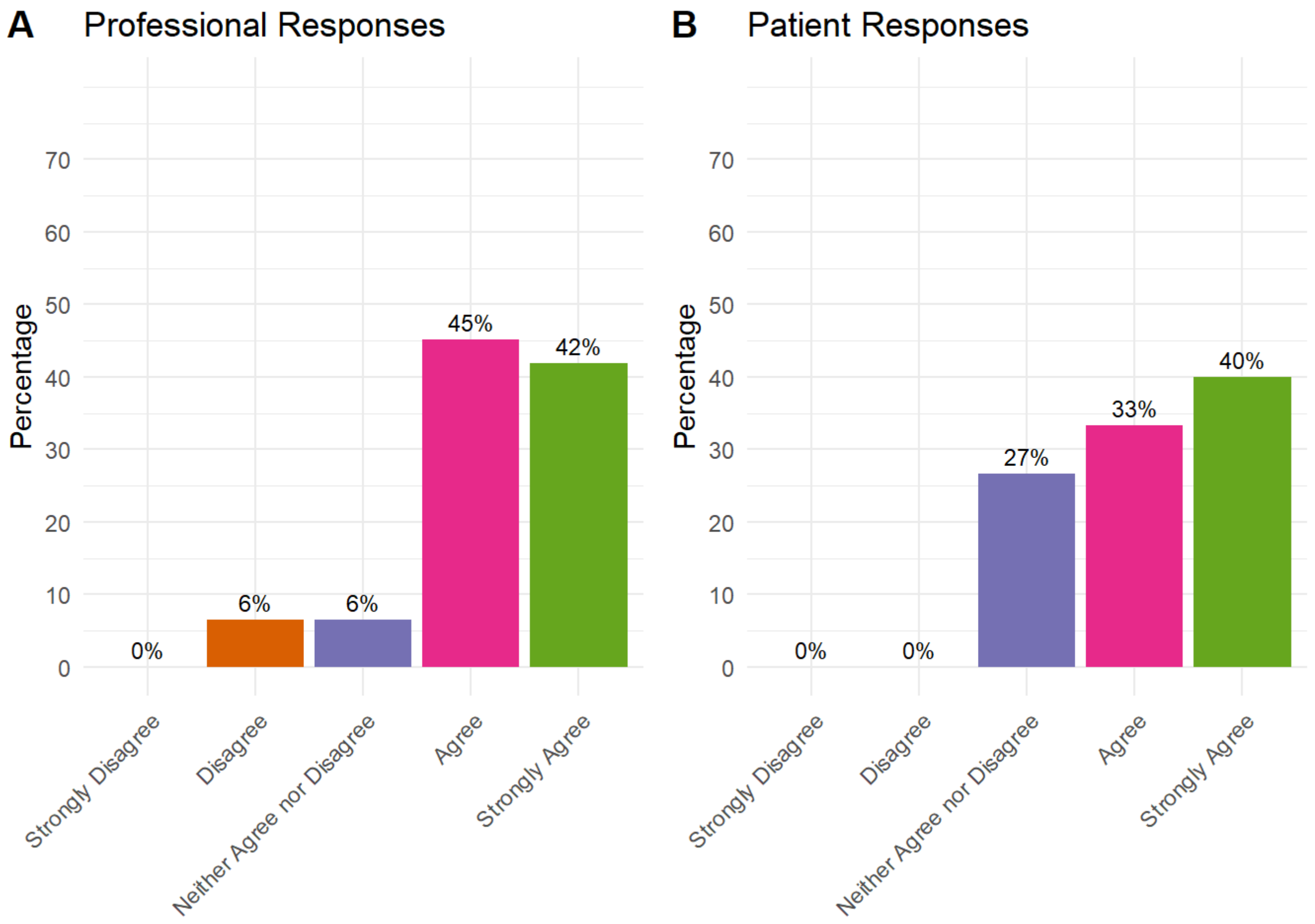


Background information: Alternatives to surgery

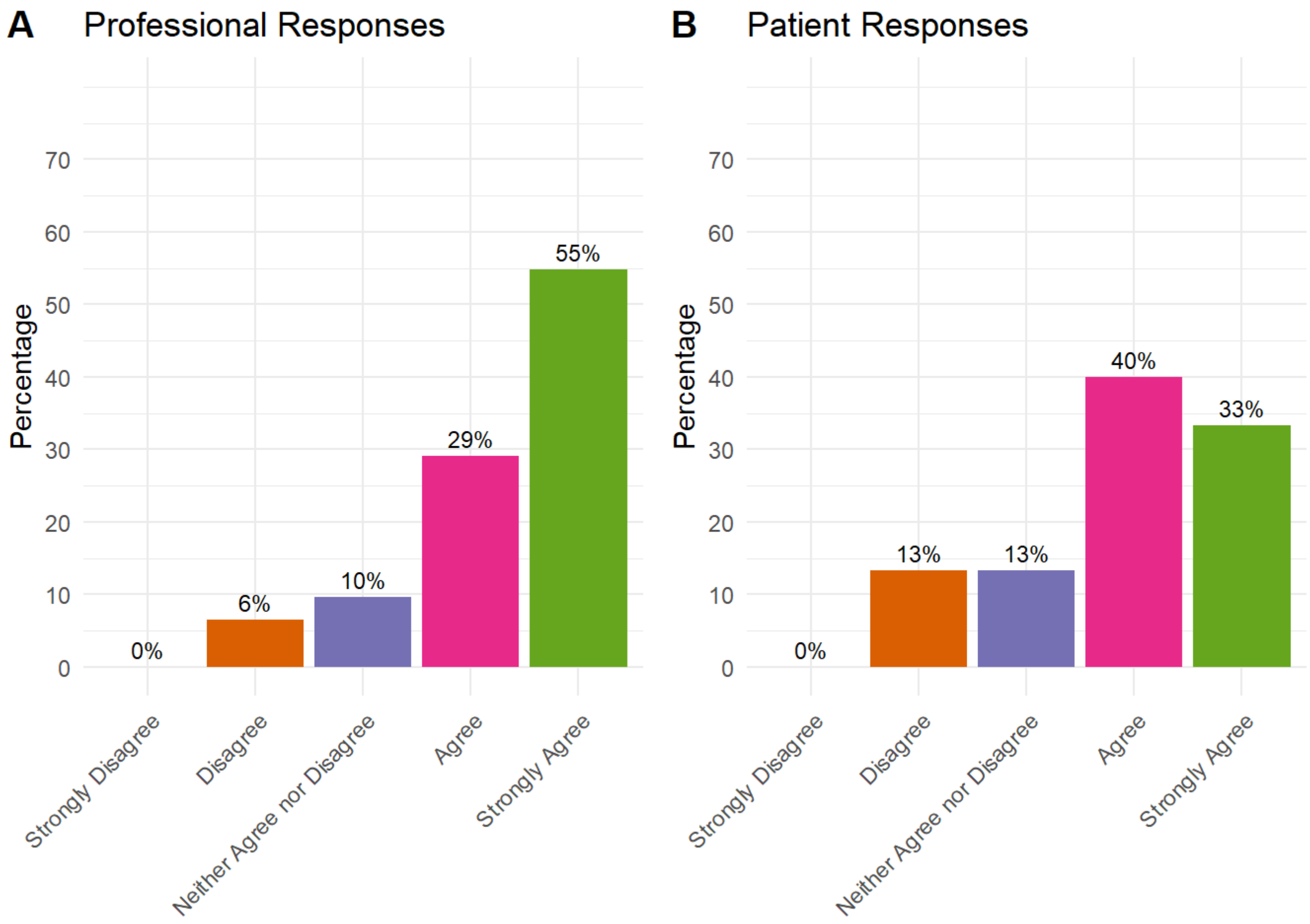


### Information on preparing for surgery should cover

Information on preparing for surgery should cover: Surgical site care

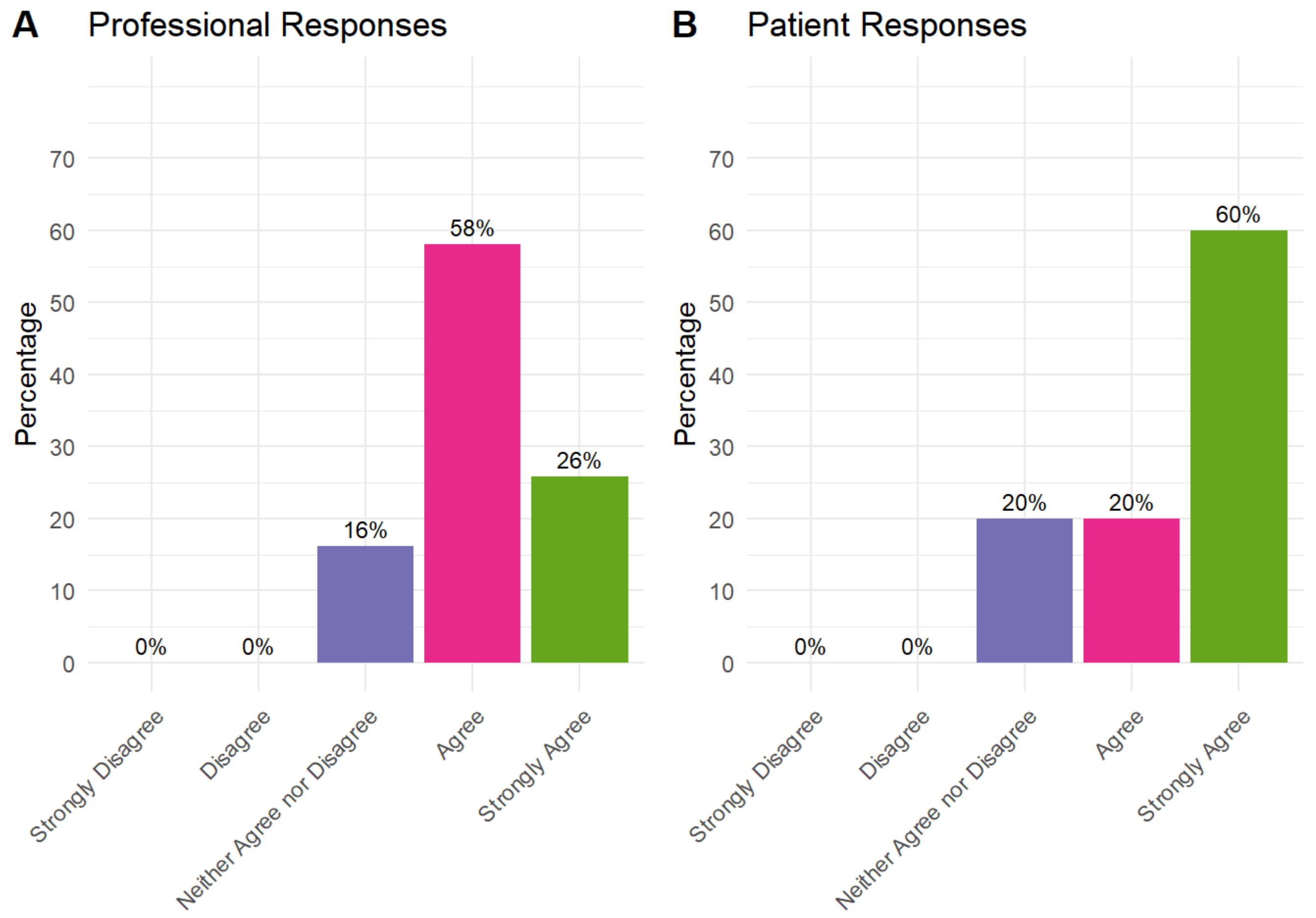


Information on preparing for surgery should cover: Purpose of pre-operative rehabilitation

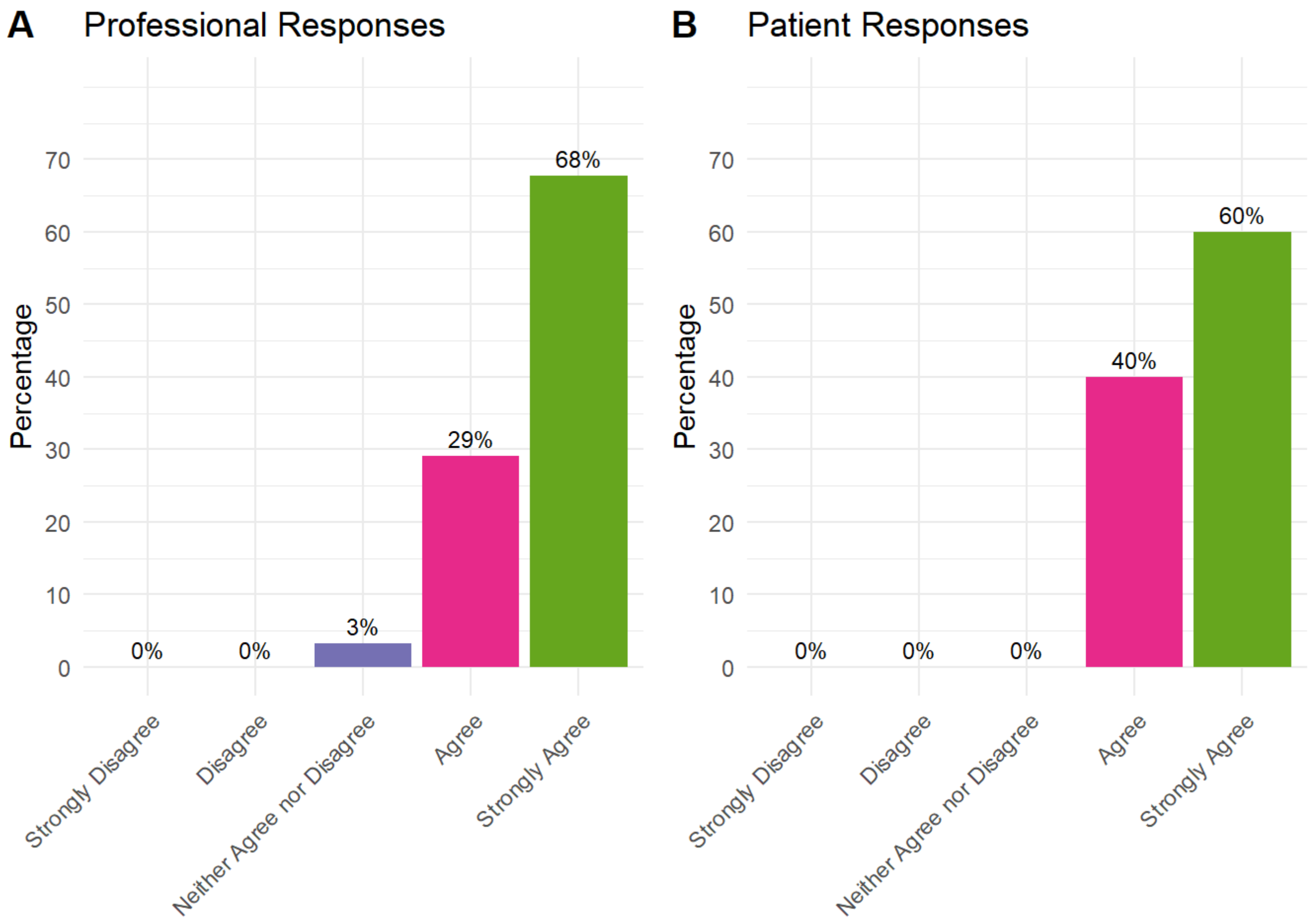


Information on preparing for surgery should cover: Patient involvement in their own management

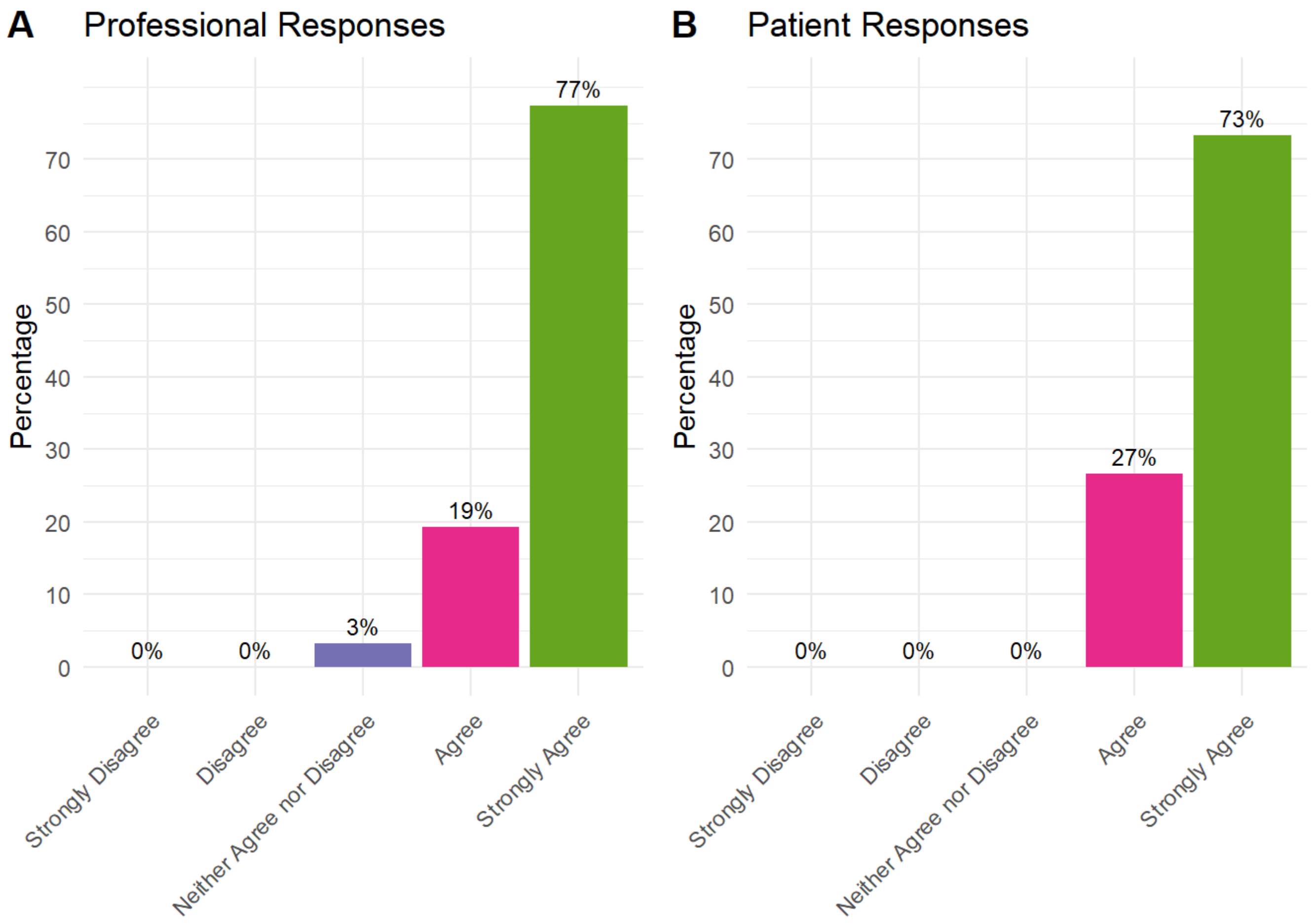


Information on preparing for surgery should cover: Goal setting

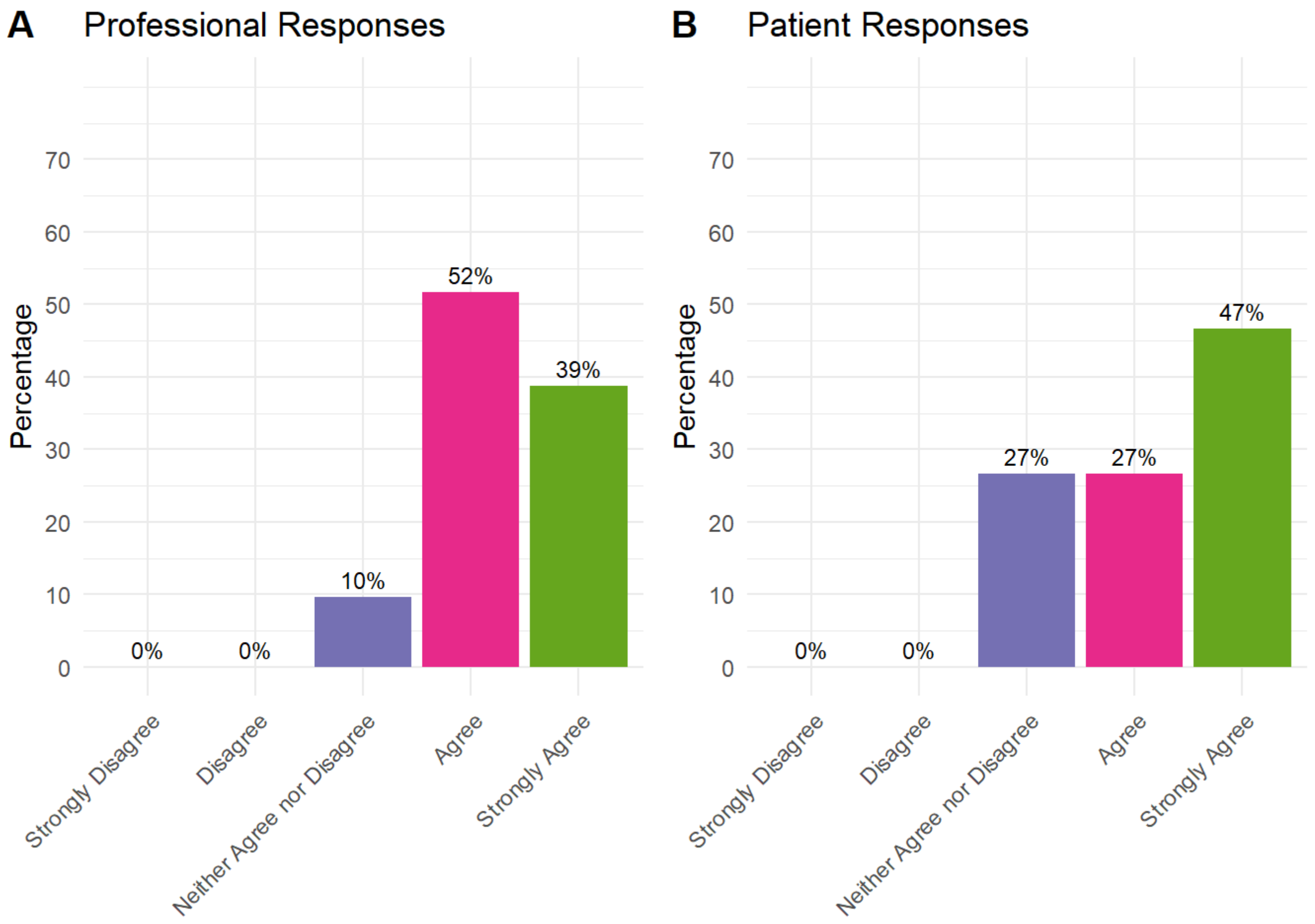


Information on preparing for surgery should cover: Using heat and cold for pain relief

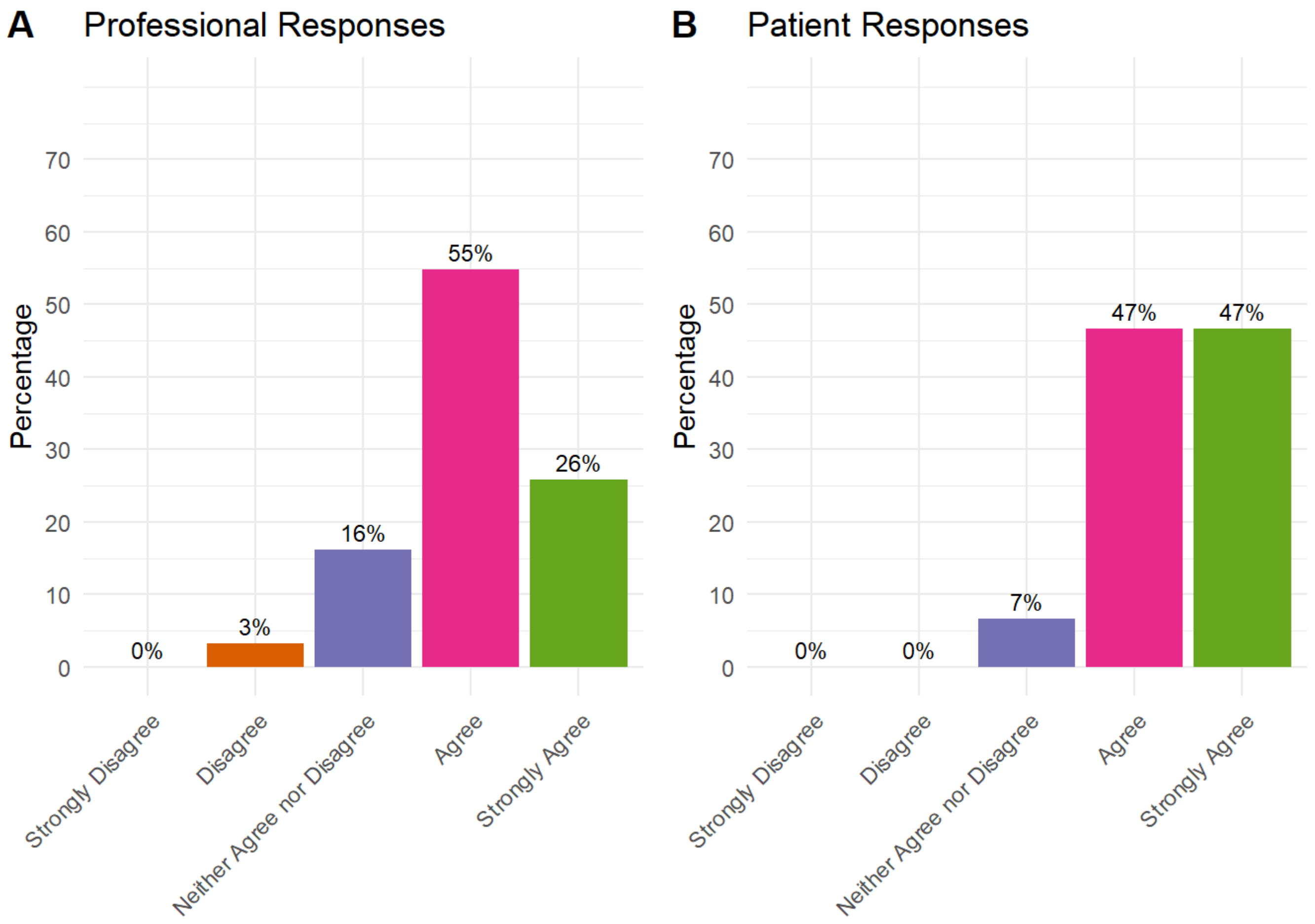


Information on preparing for surgery should cover: Obtaining and using walking aids and other equipment

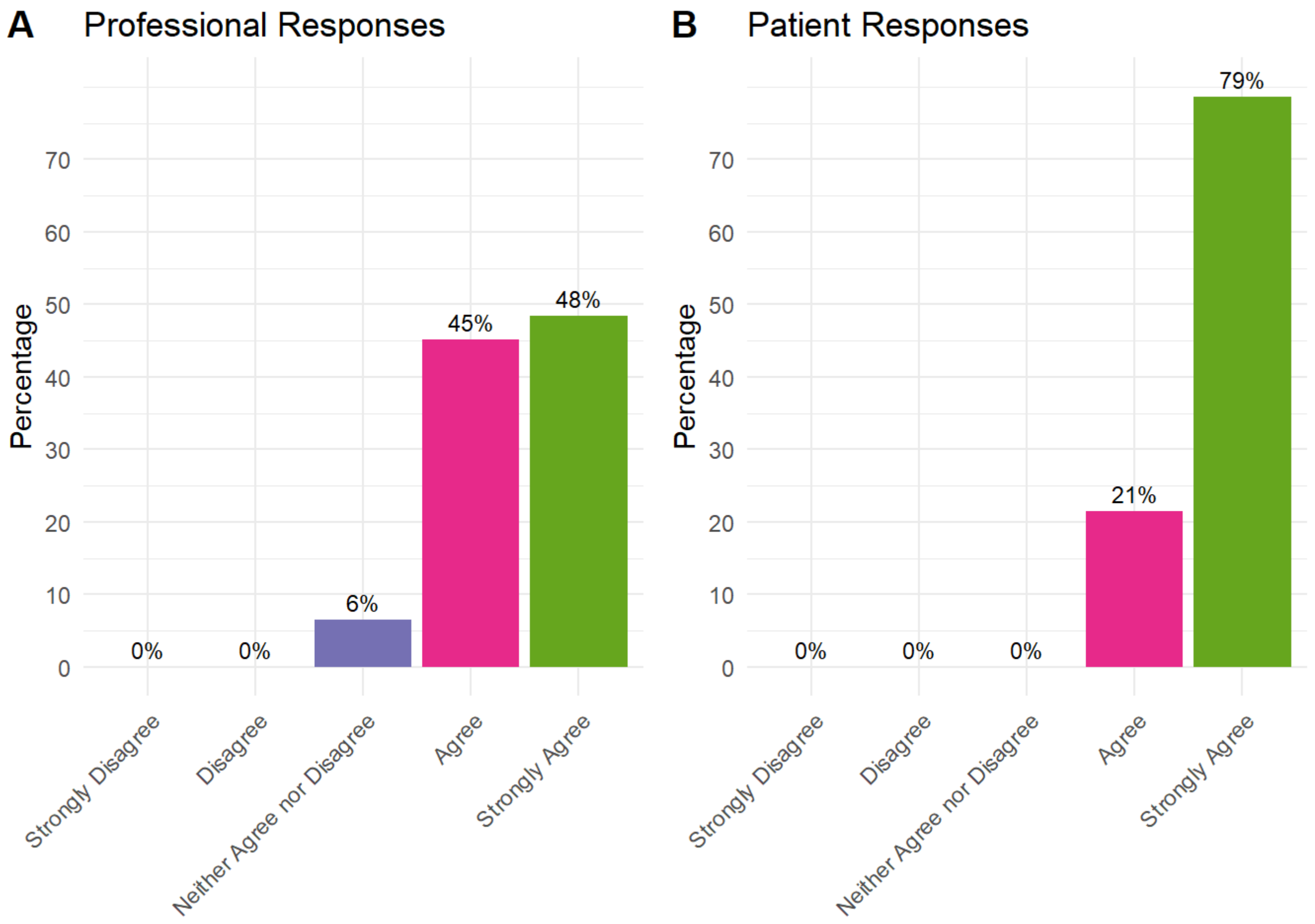


Information on preparing for surgery should cover: Making home preparations

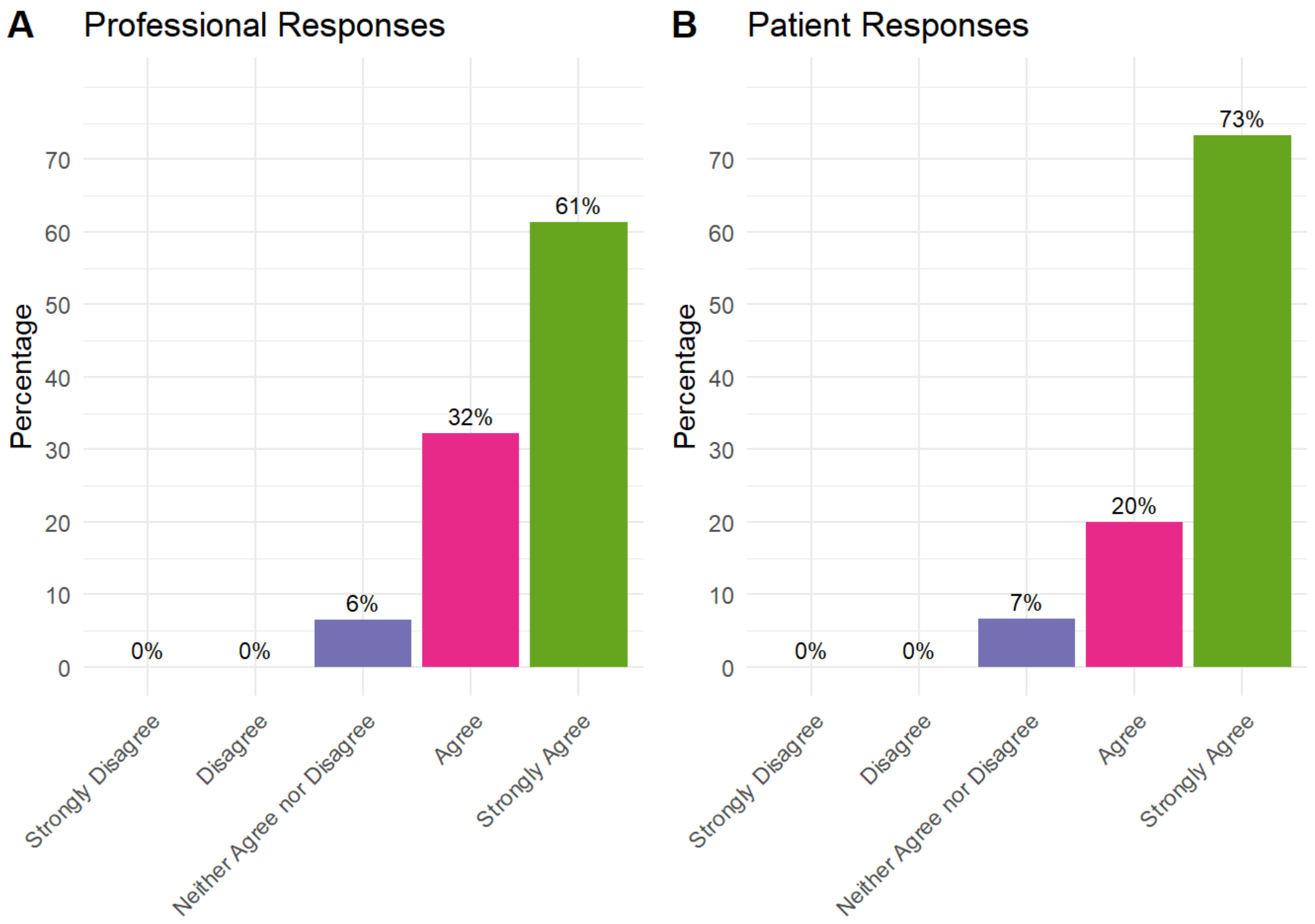


Information on preparing for surgery should cover: Arranging any social support

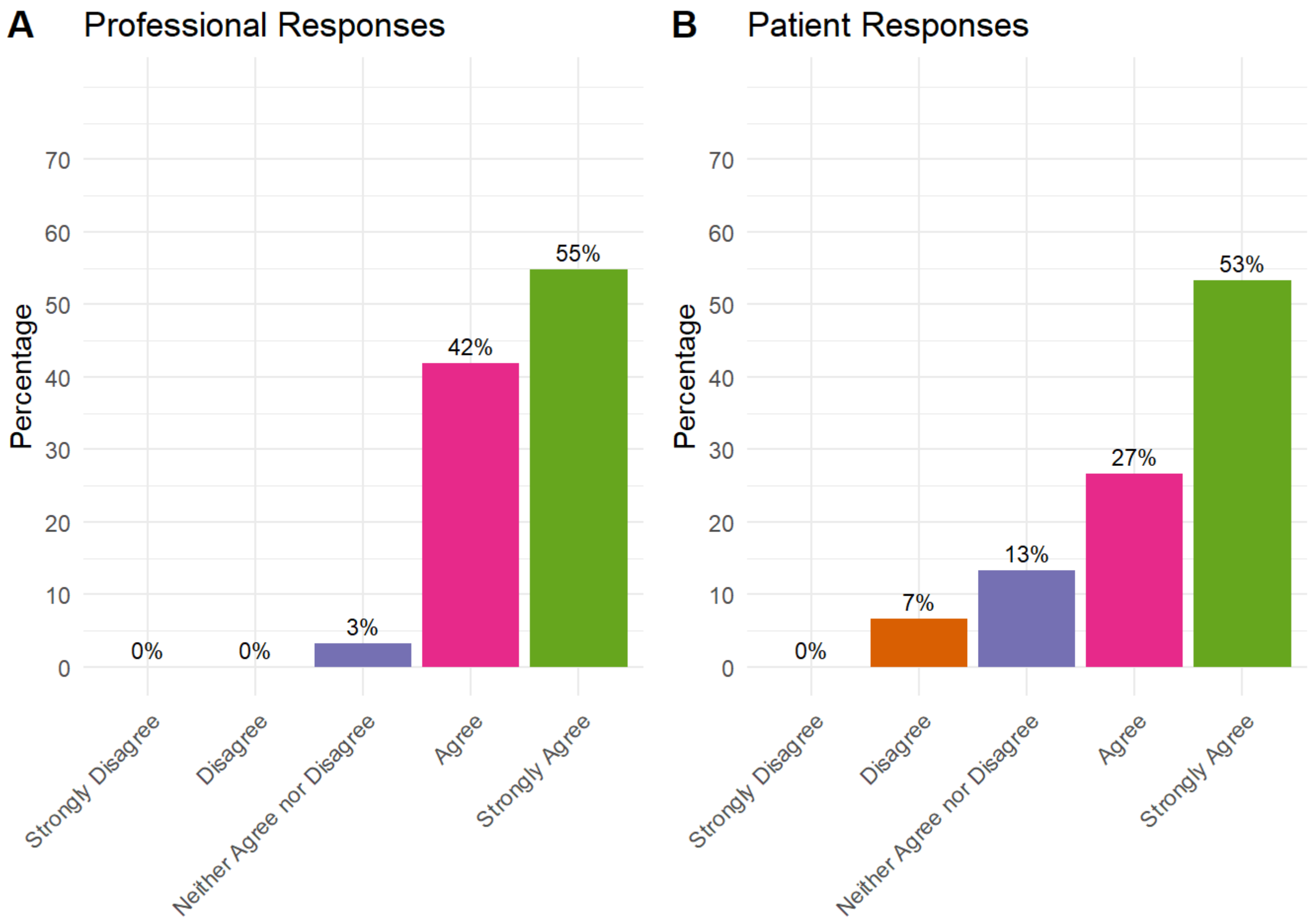


Information on preparing for surgery should cover: Arranging transport to and from the hospital

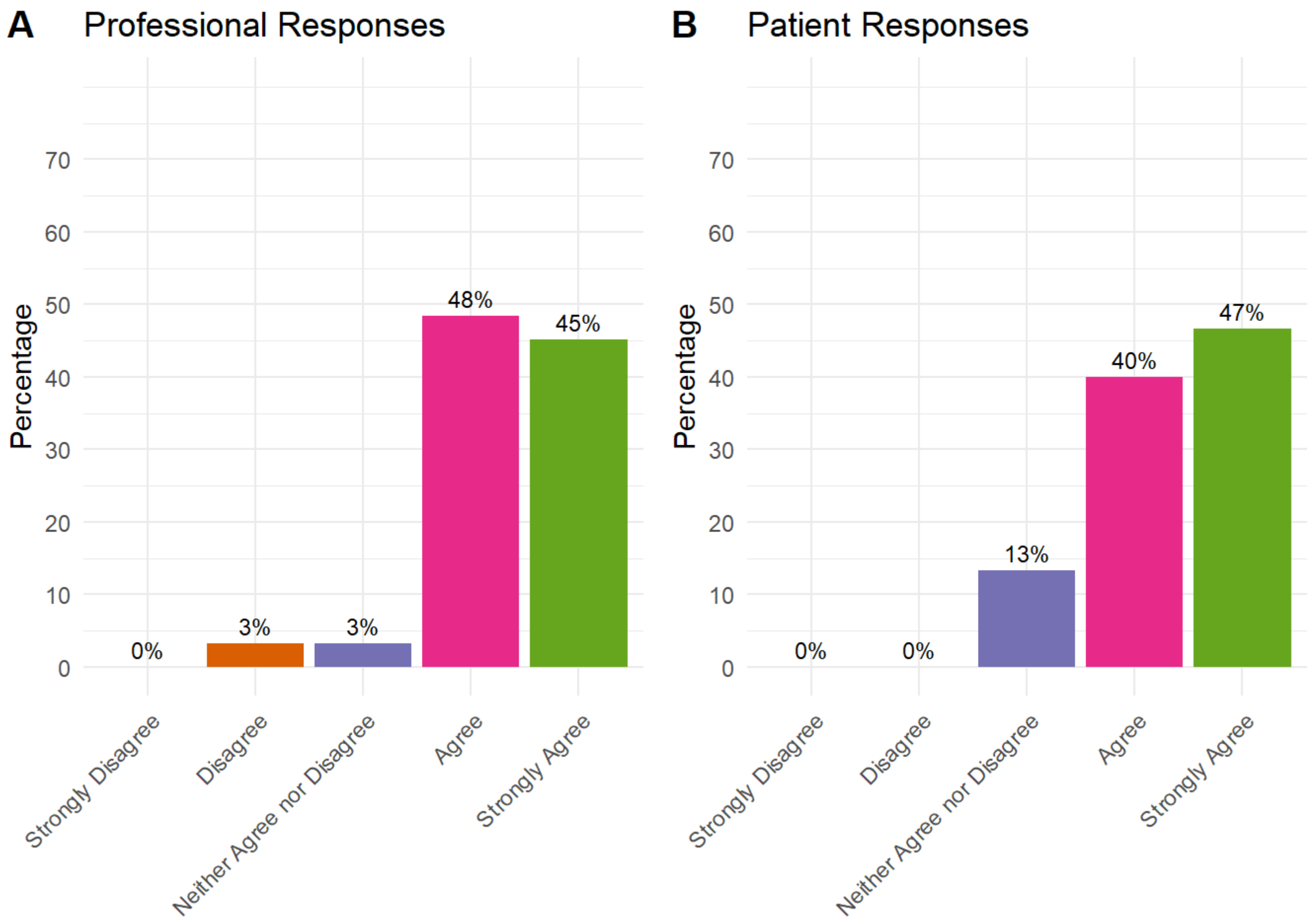


Information on preparing for surgery should cover: Optimising management of diabetes

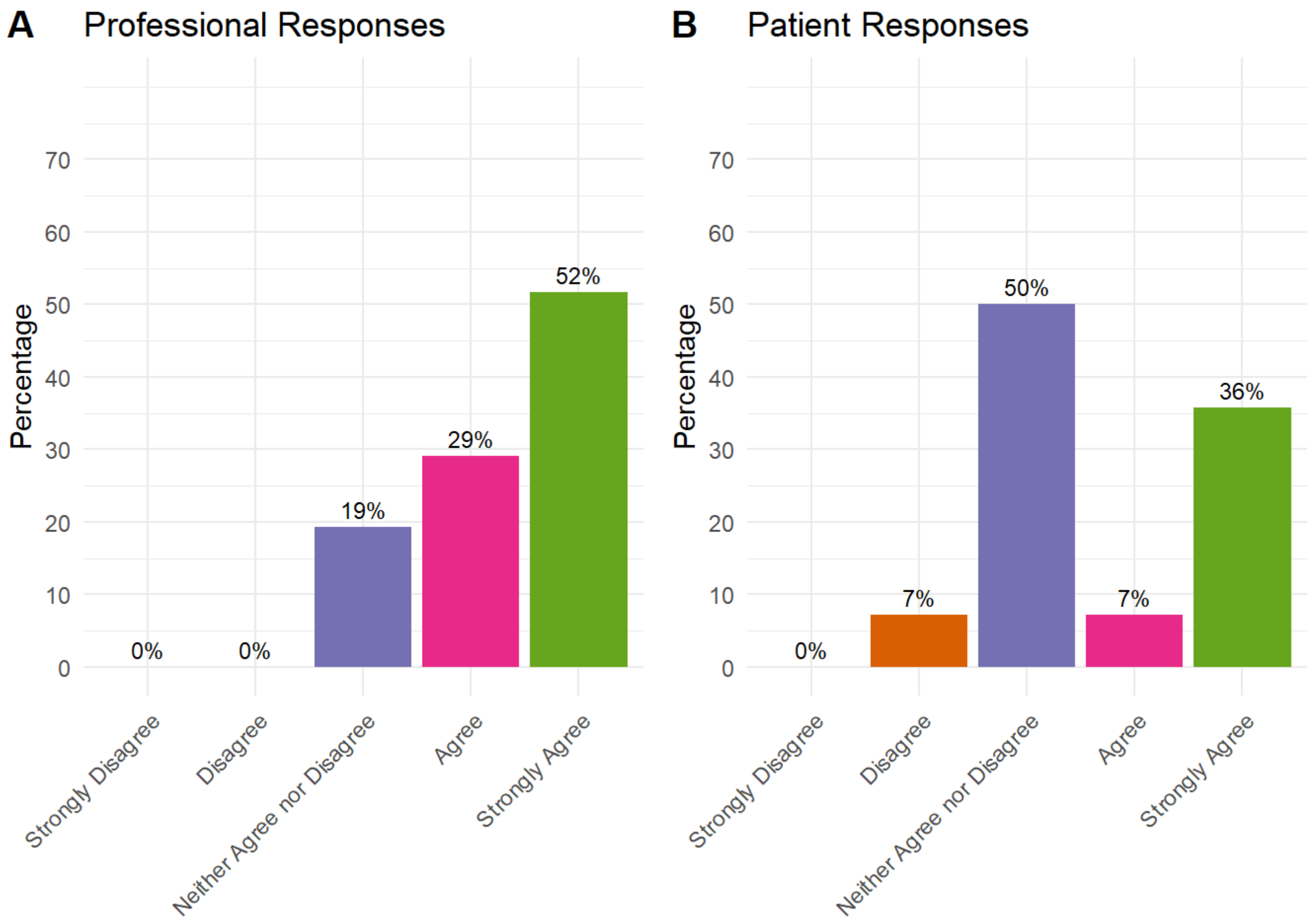


Information on preparing for surgery should cover: Emotional well-being

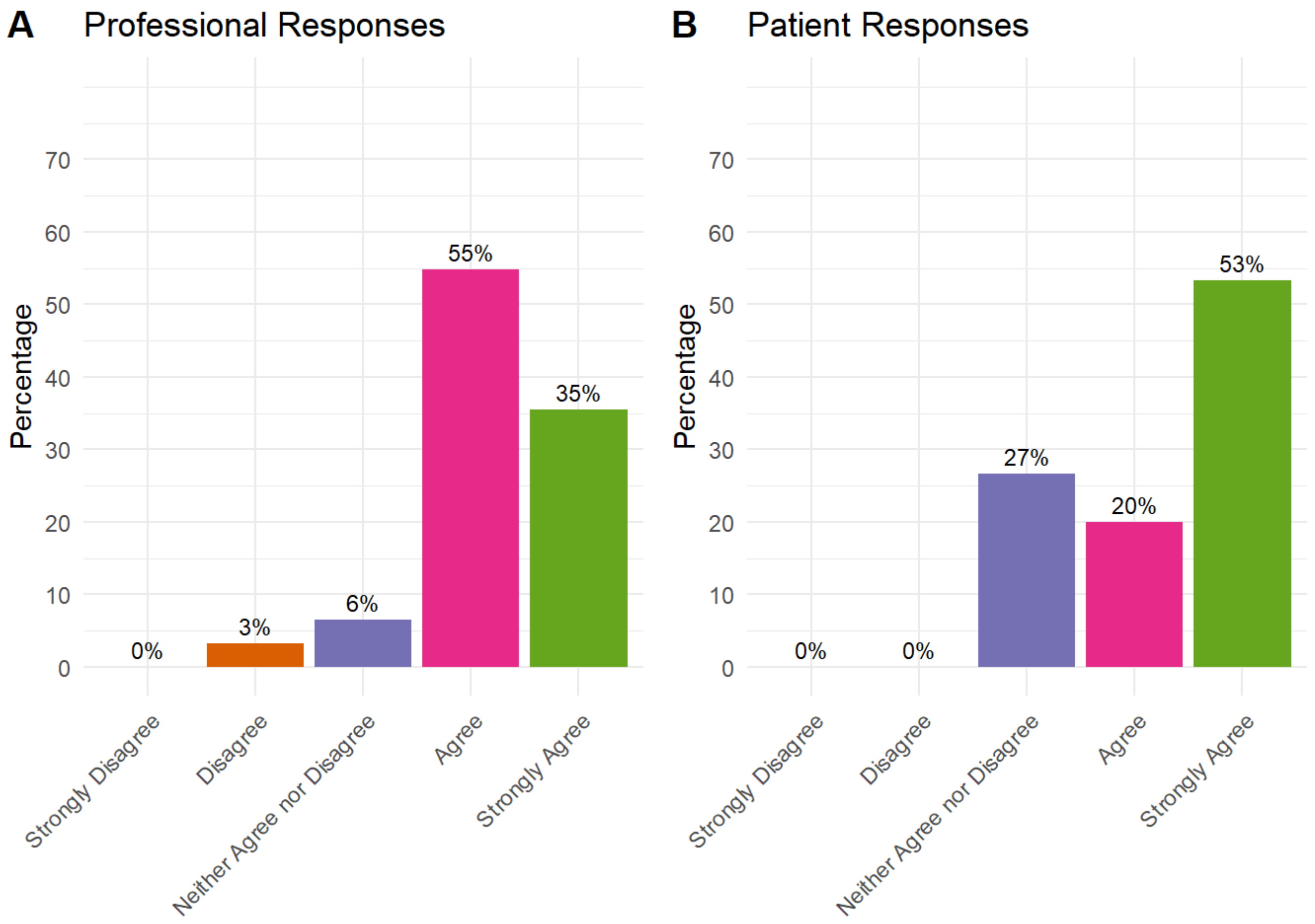


Information on preparing for surgery should cover: Education for other people, such as carers

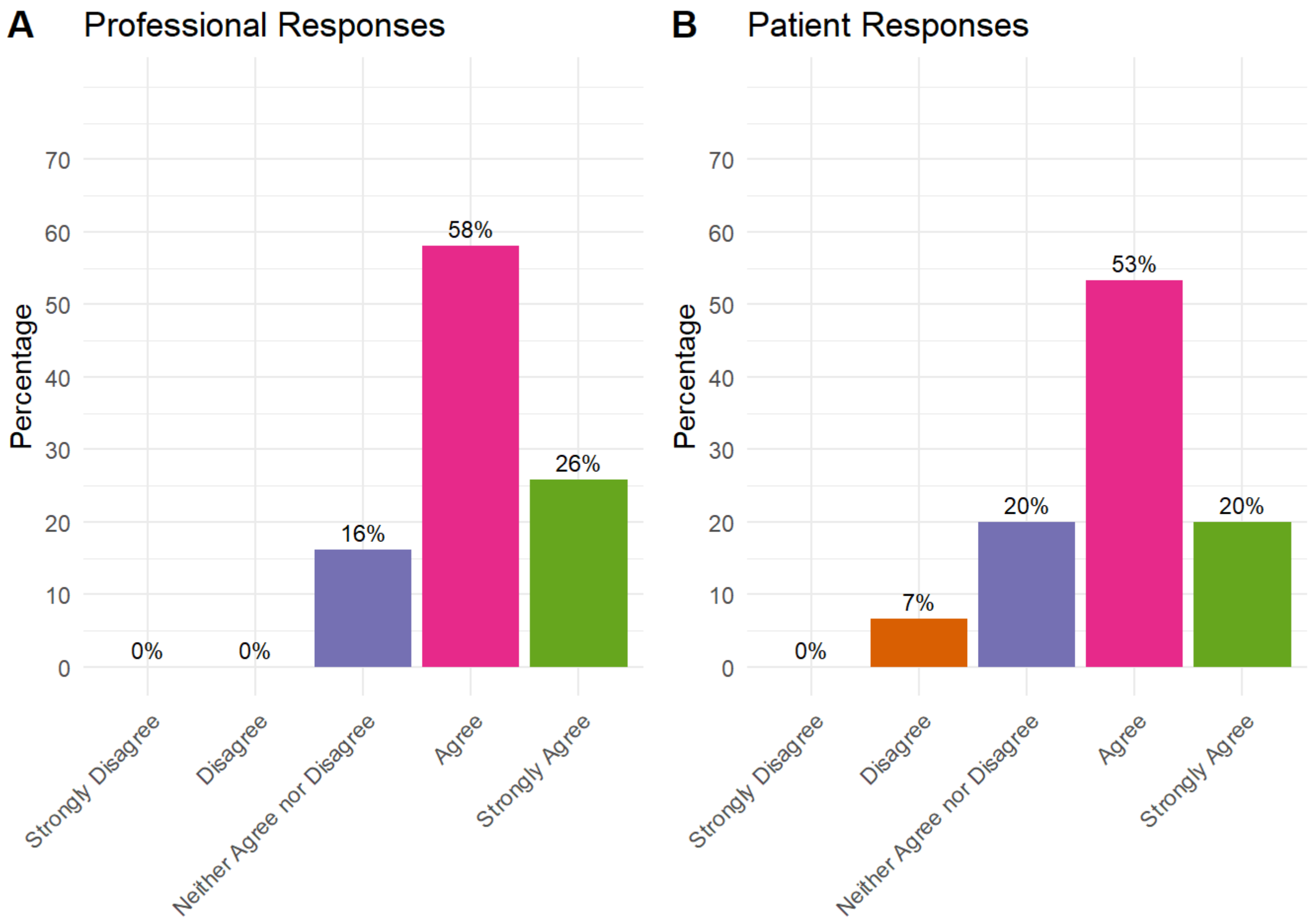


Information on preparing for surgery should cover: Identifying and arranging any social care provision

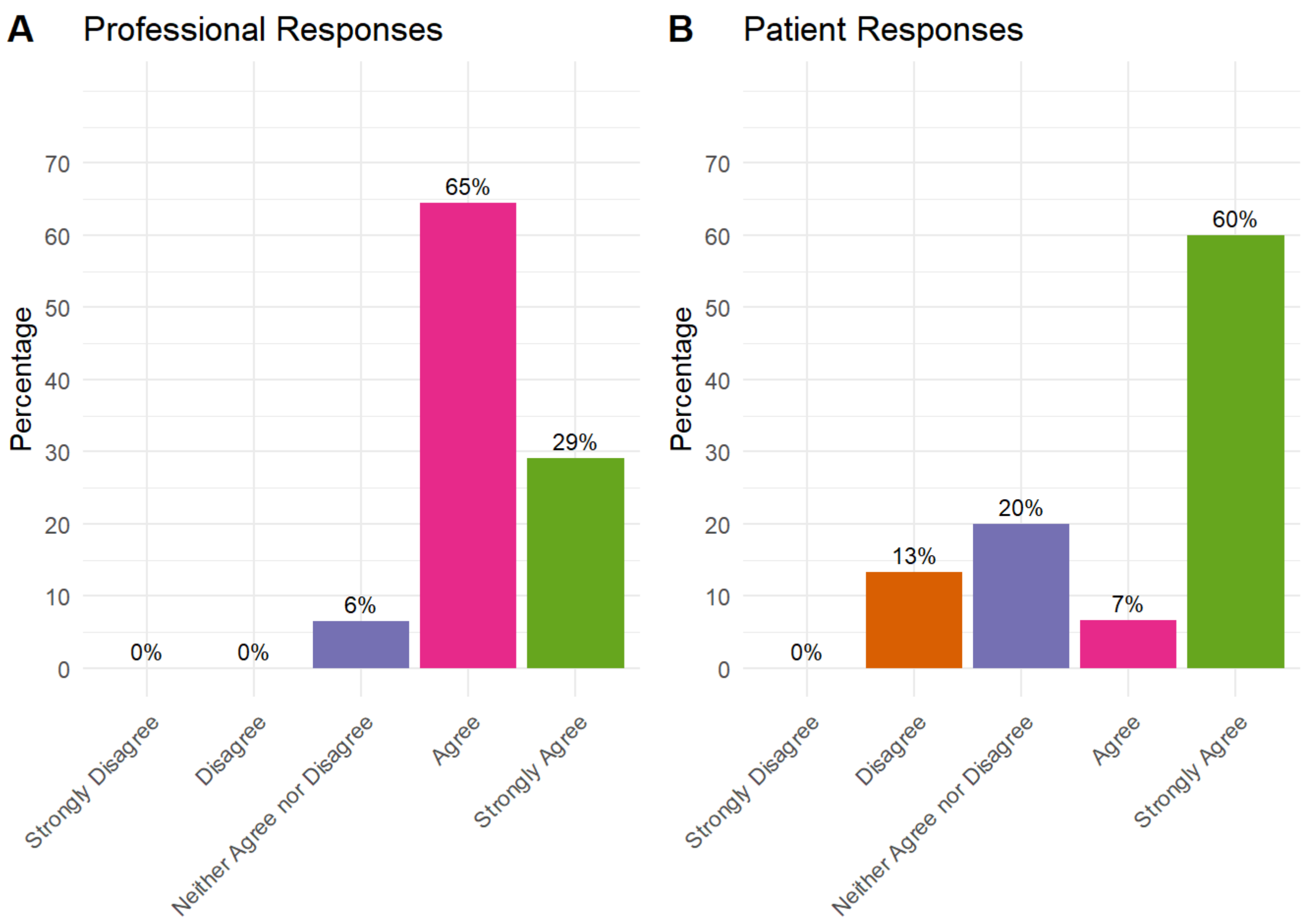


### What to expect while in hospital and at home should cover

What to expect while in hospital and at home should cover: What to expect during the hospital stay

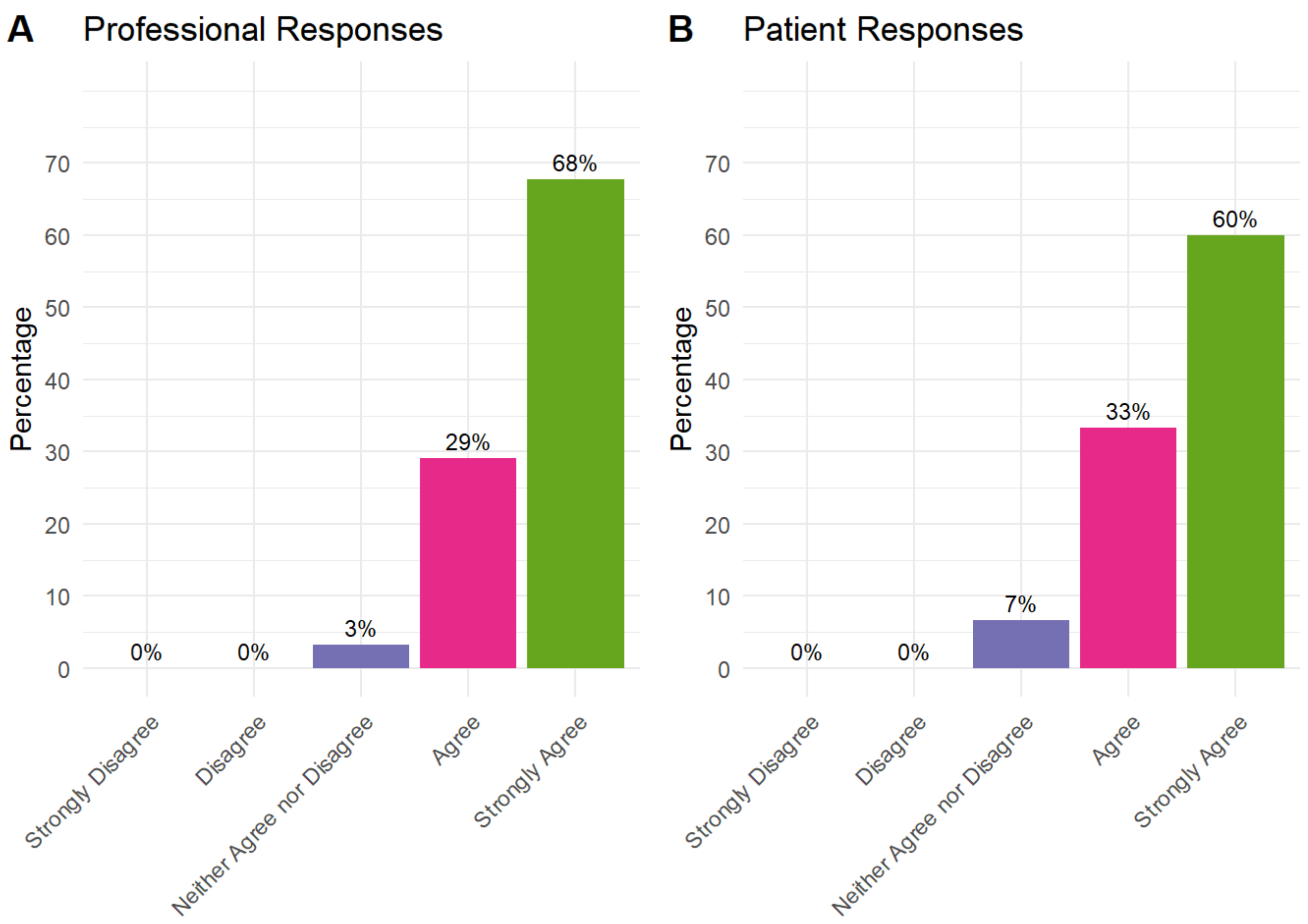


What to expect while in hospital and at home should cover: Packing list for hospital

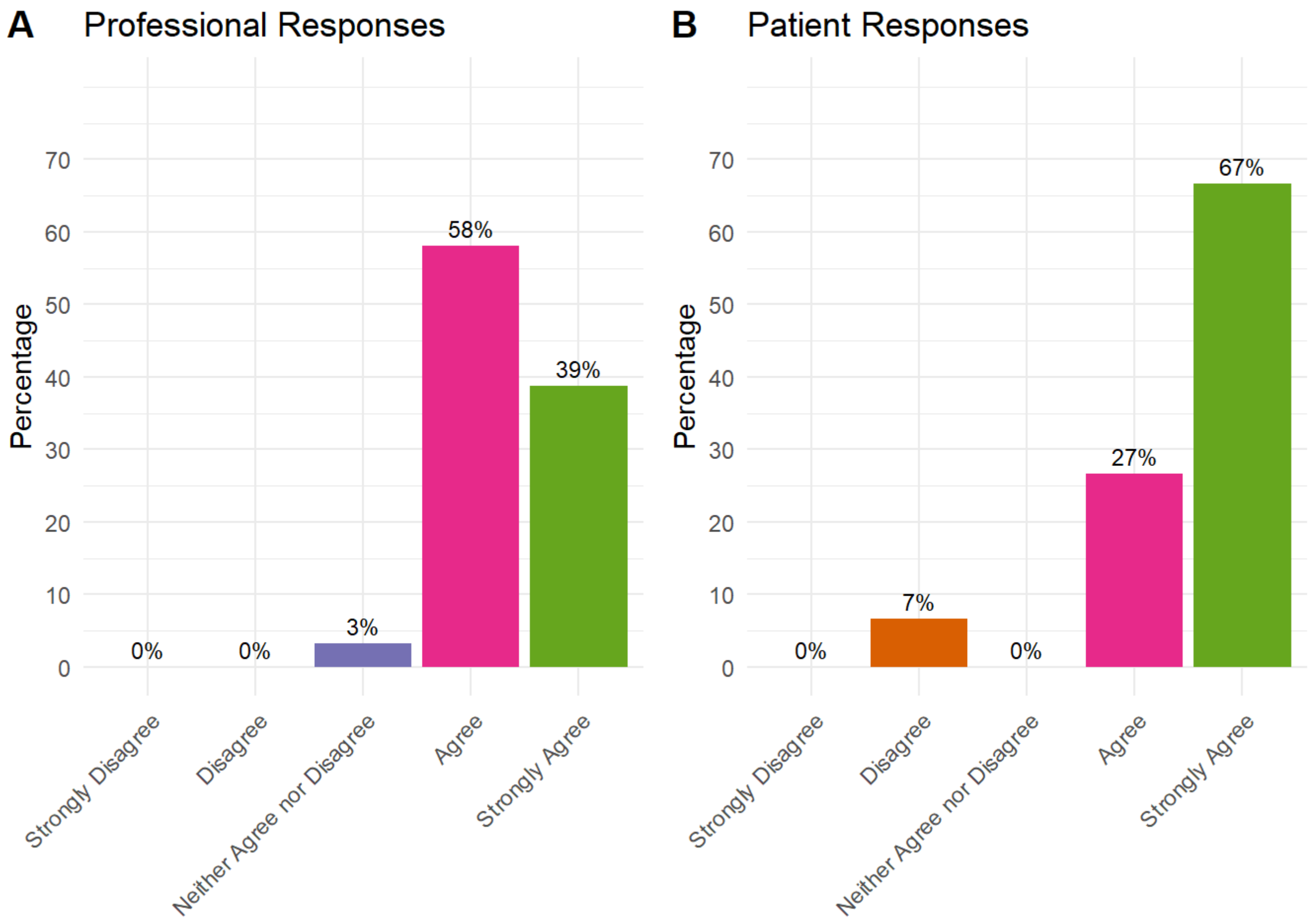


What to expect while in hospital and at home should cover: In-hospital timeline

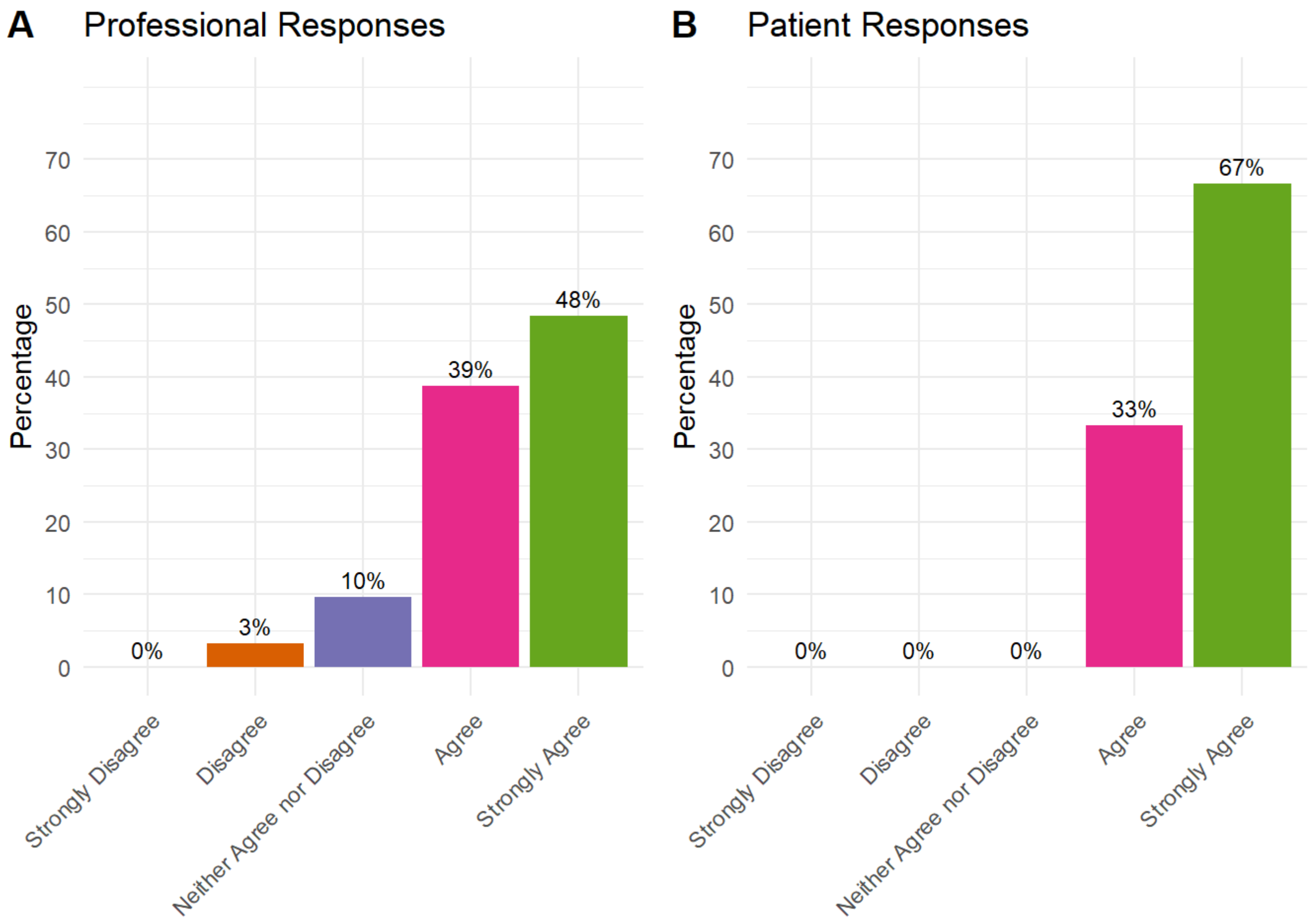


What to expect while in hospital and at home should cover: Visitor information

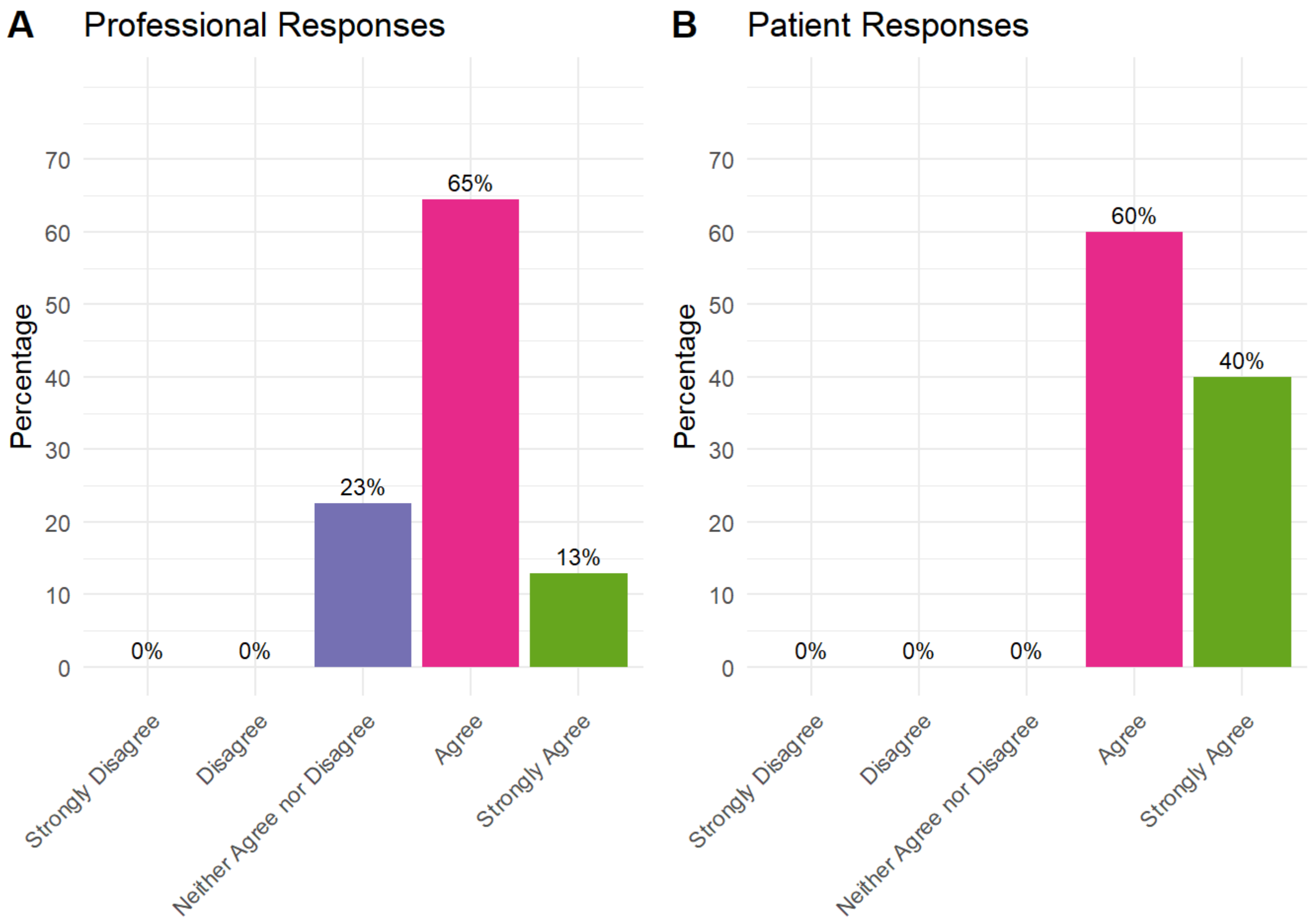


What to expect while in hospital and at home should cover: Day of surgery logistics

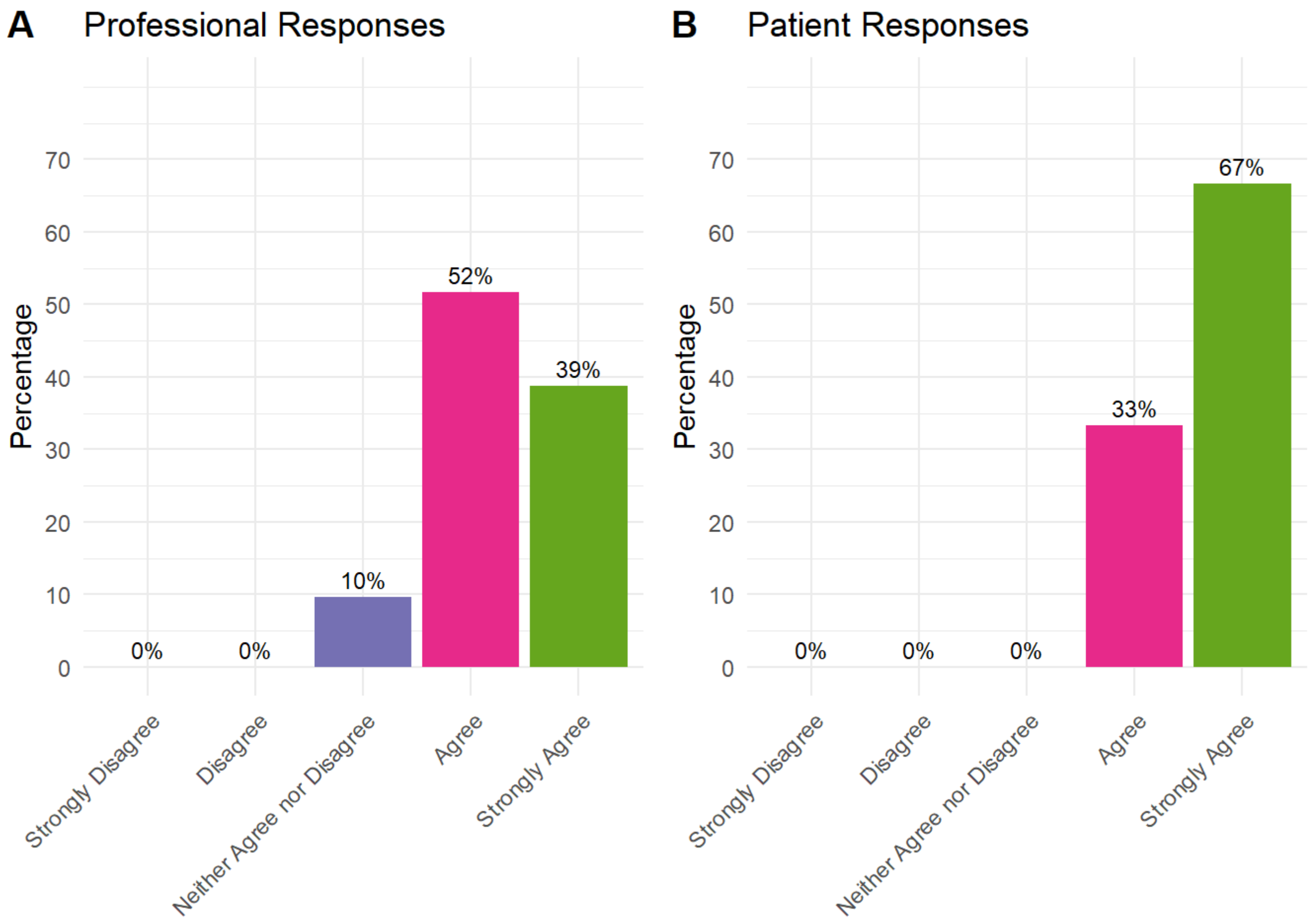


What to expect while in hospital and at home should cover: What a THR/TKR surgical procedure involves

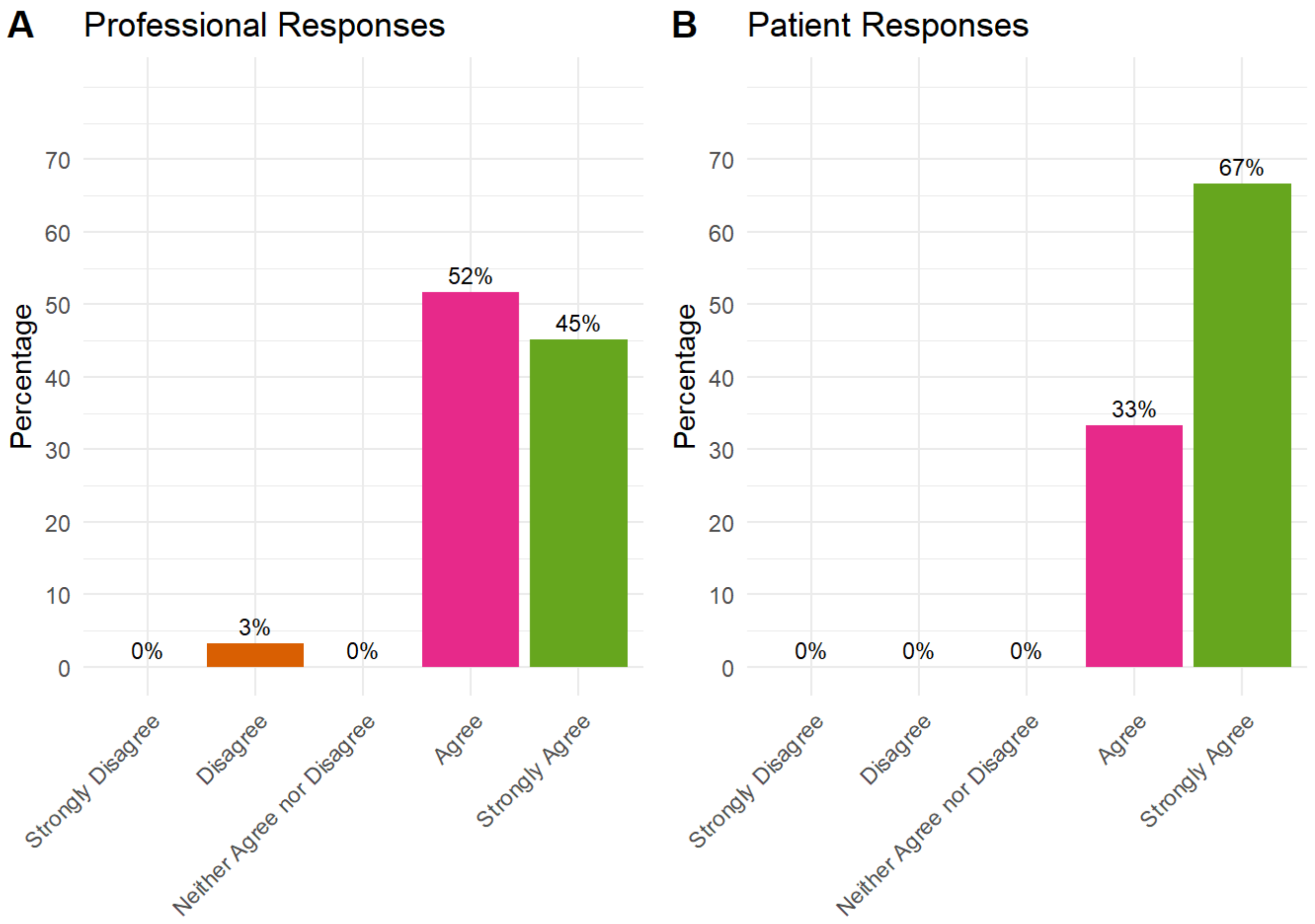


What to expect while in hospital and at home should cover: Information on the prosthesis (replacement)

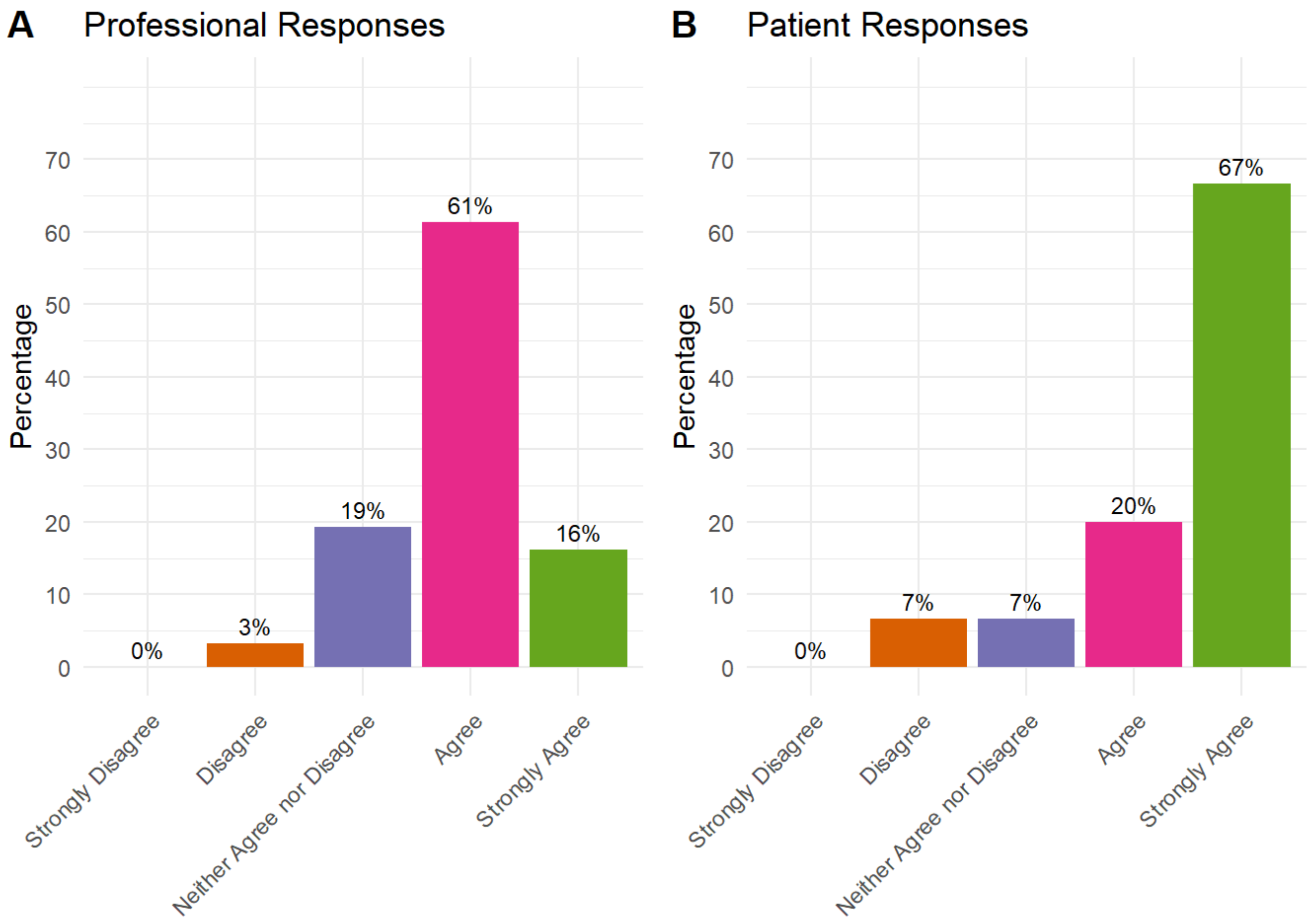


What to expect while in hospital and at home should cover: Anaesthesia options

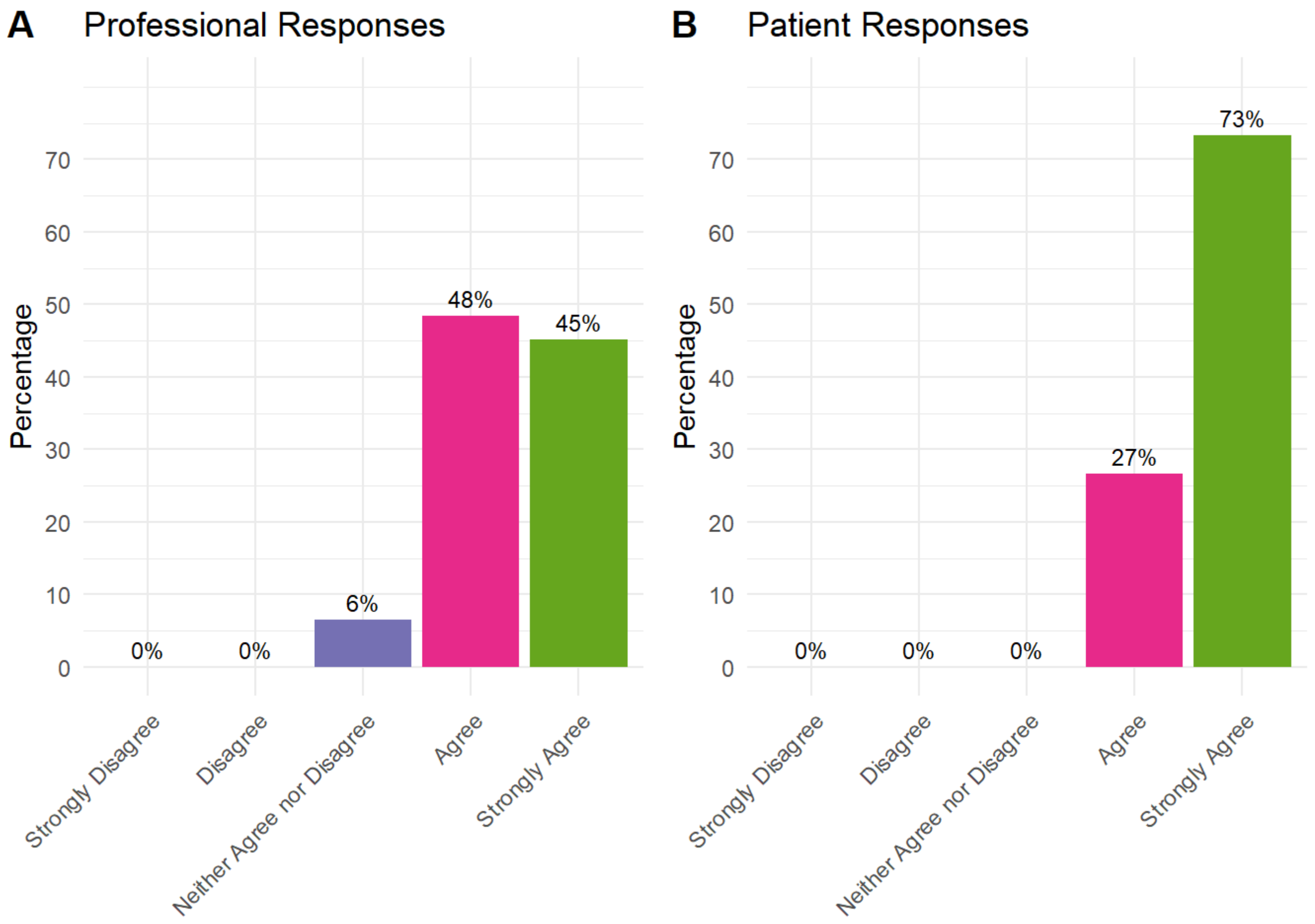


What to expect while in hospital and at home should cover: Risks of THR/TKR surgery and how to minimise them

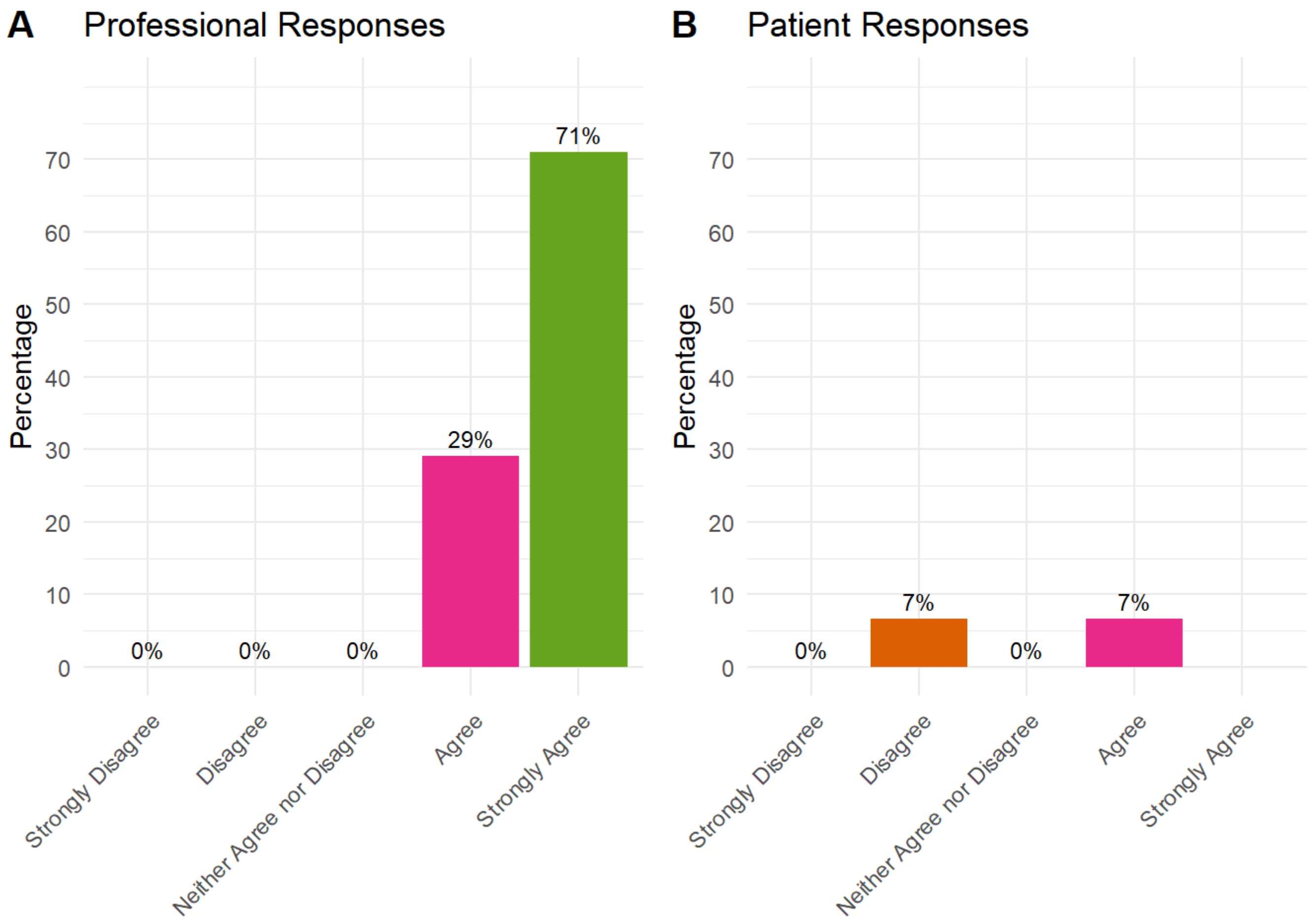


What to expect while in hospital and at home should cover: Common issues that may occur following THR/TKR surgery which do not need to cause alarm

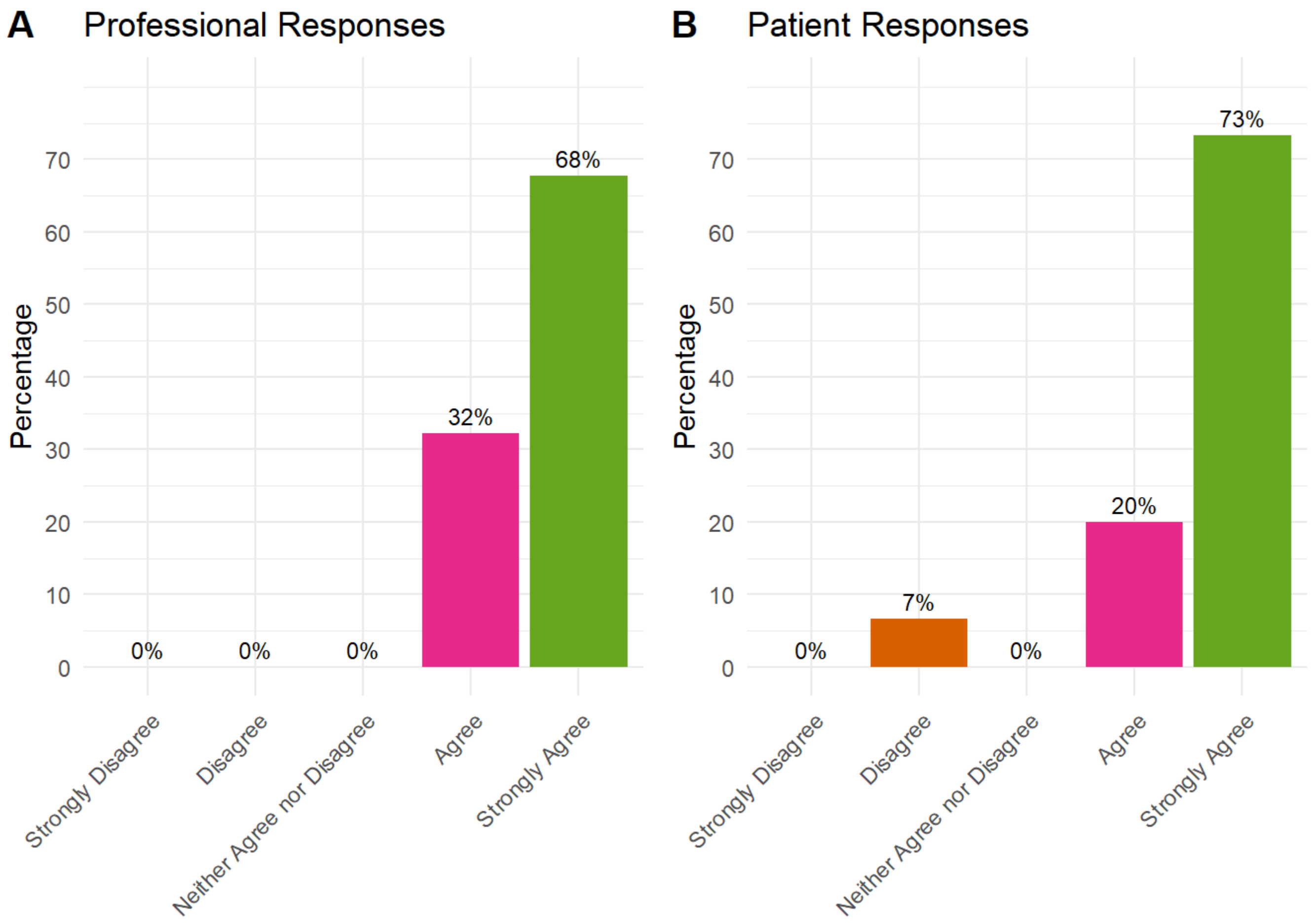


What to expect while in hospital and at home should cover: Pain expectations

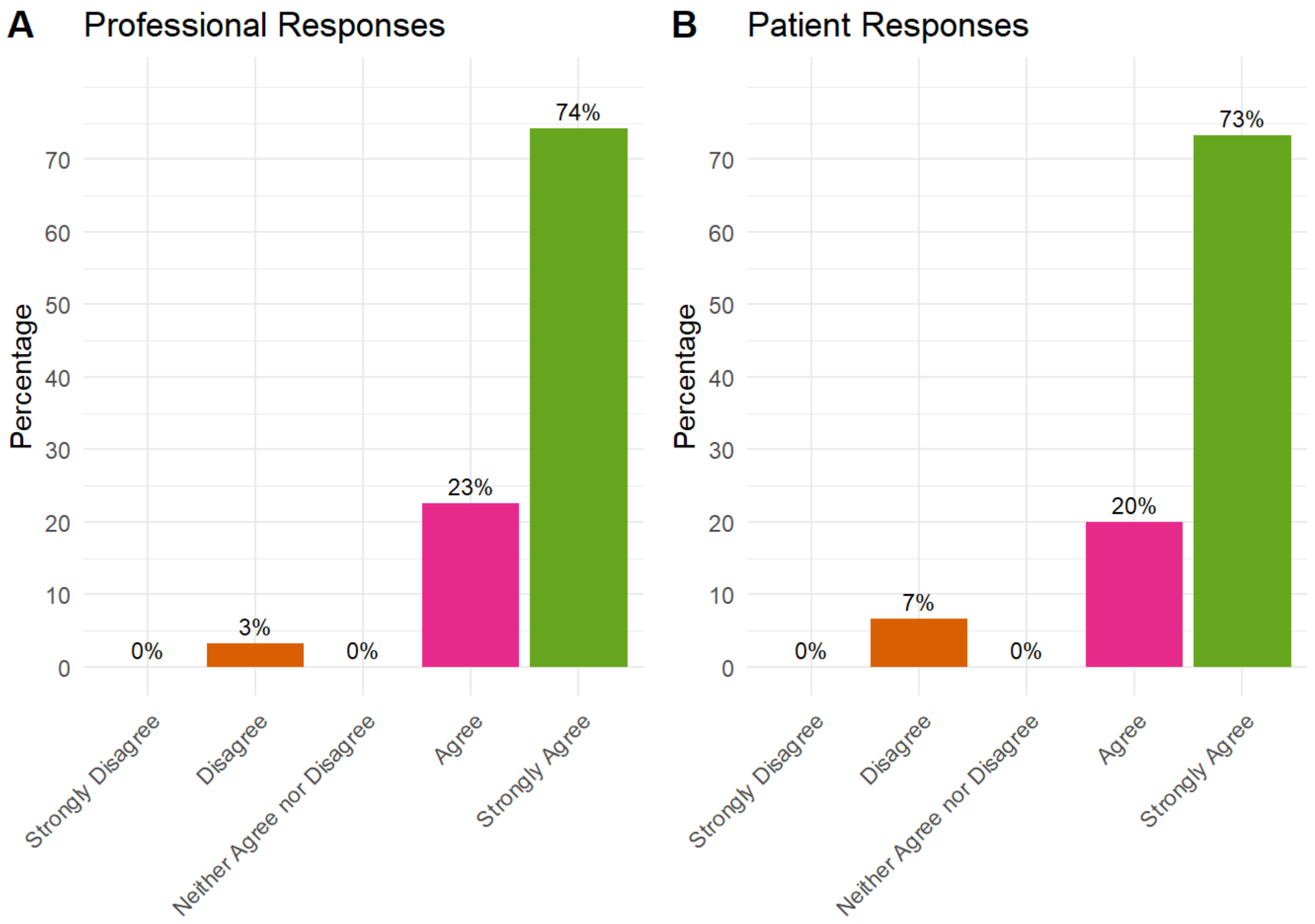


What to expect while in hospital and at home should cover: Swelling

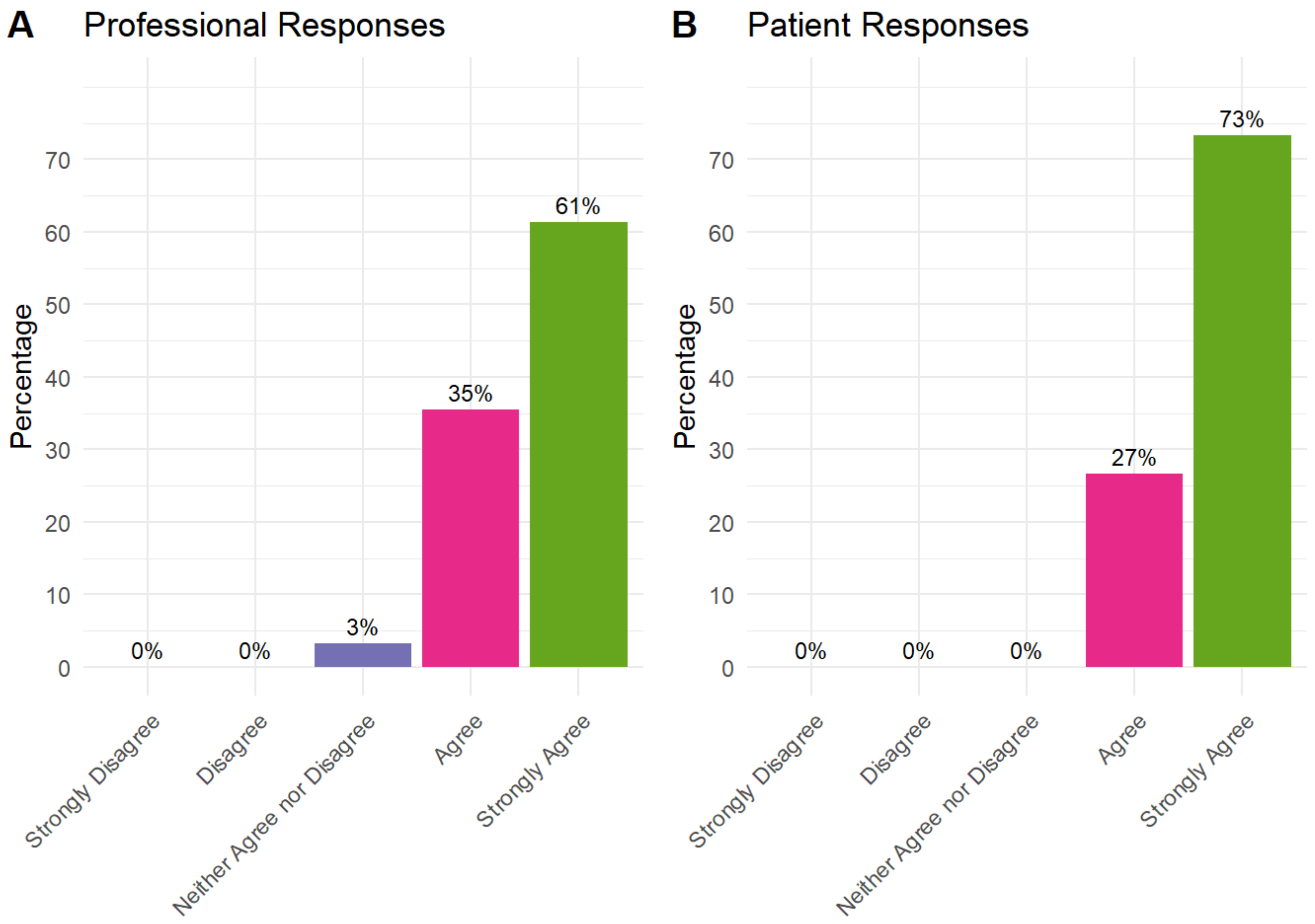


What to expect while in hospital and at home should cover: Wound healing

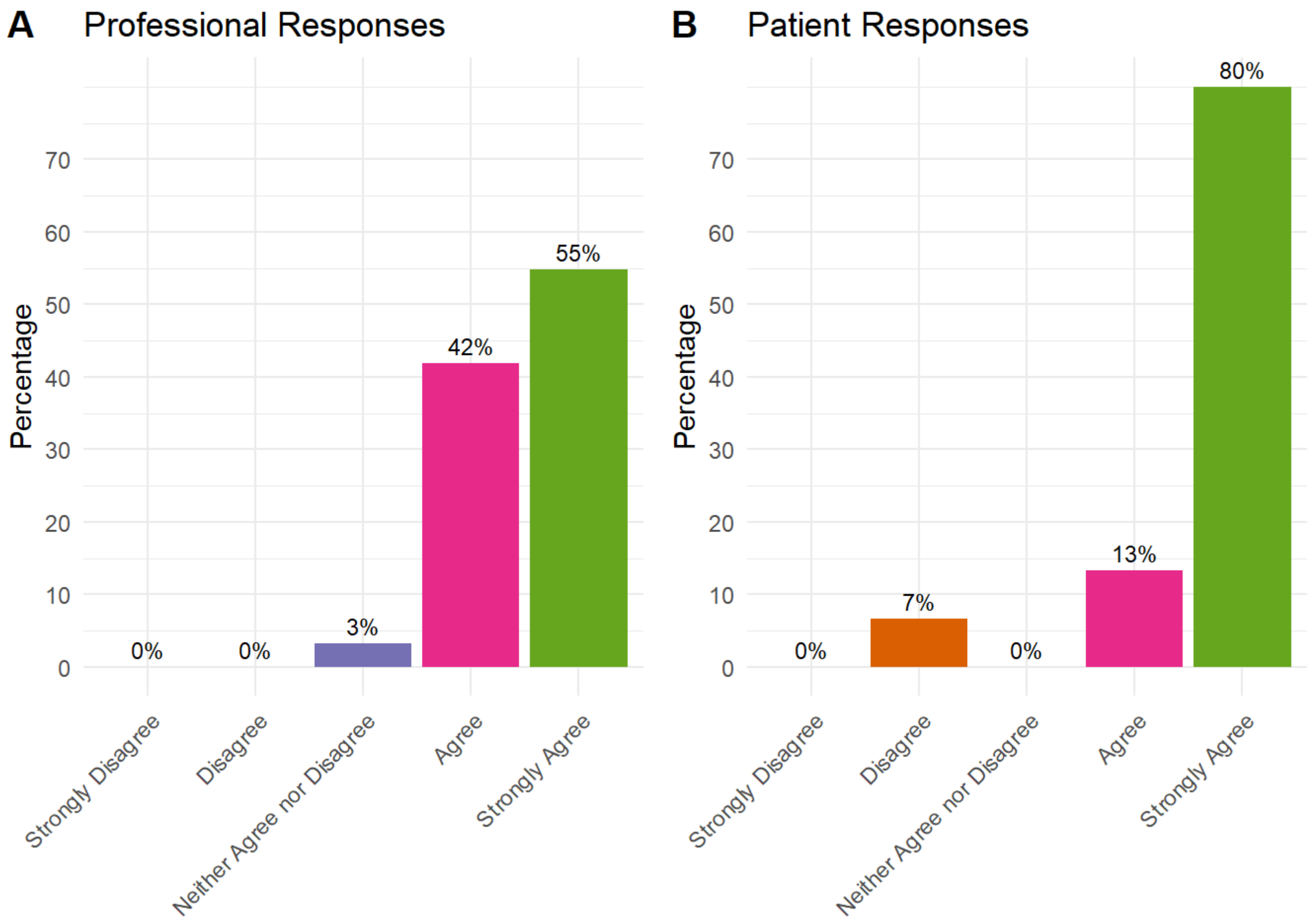


What to expect while in hospital and at home should cover: What to expect following discharge

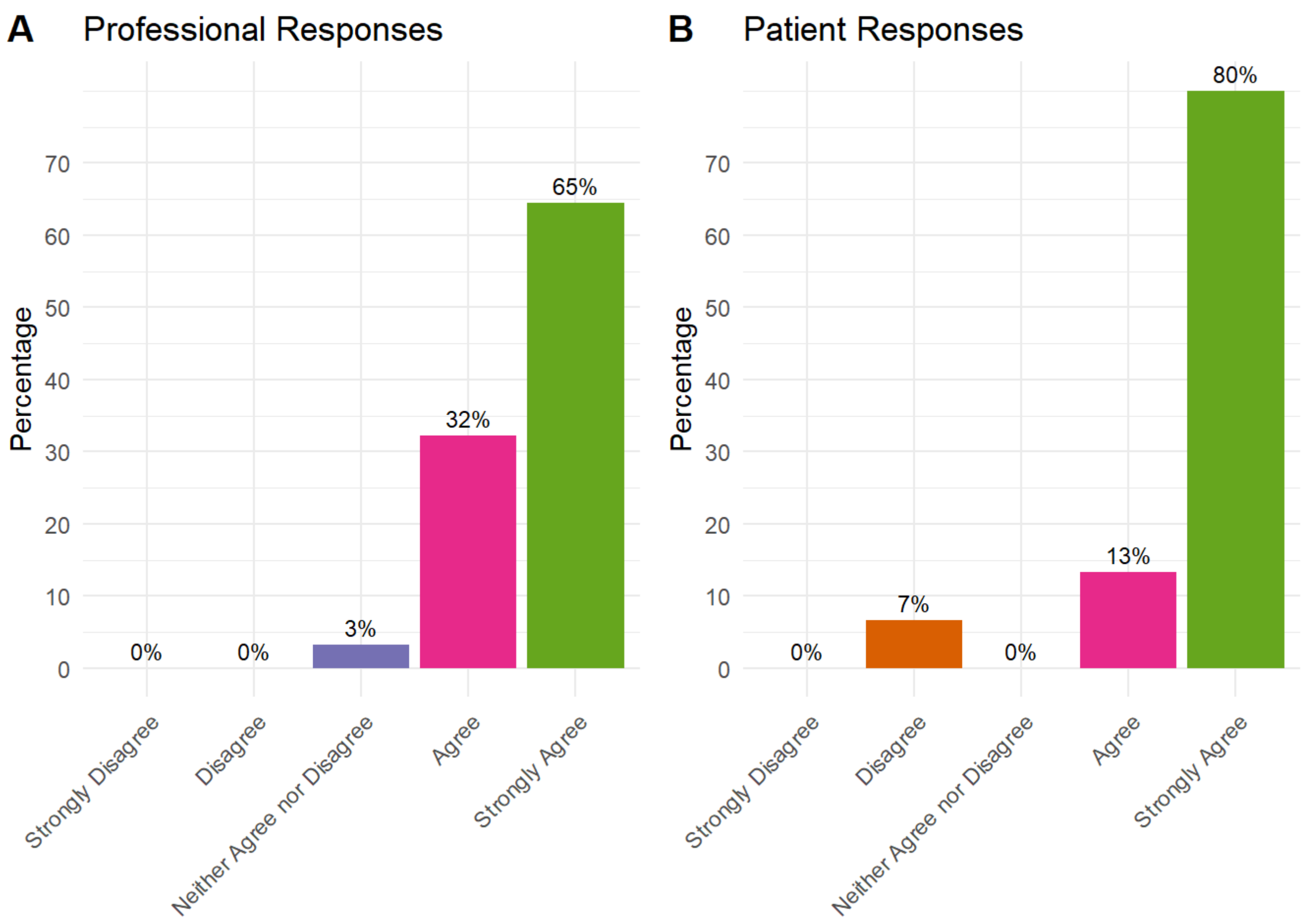


What to expect while in hospital and at home should cover: Recovery expectations

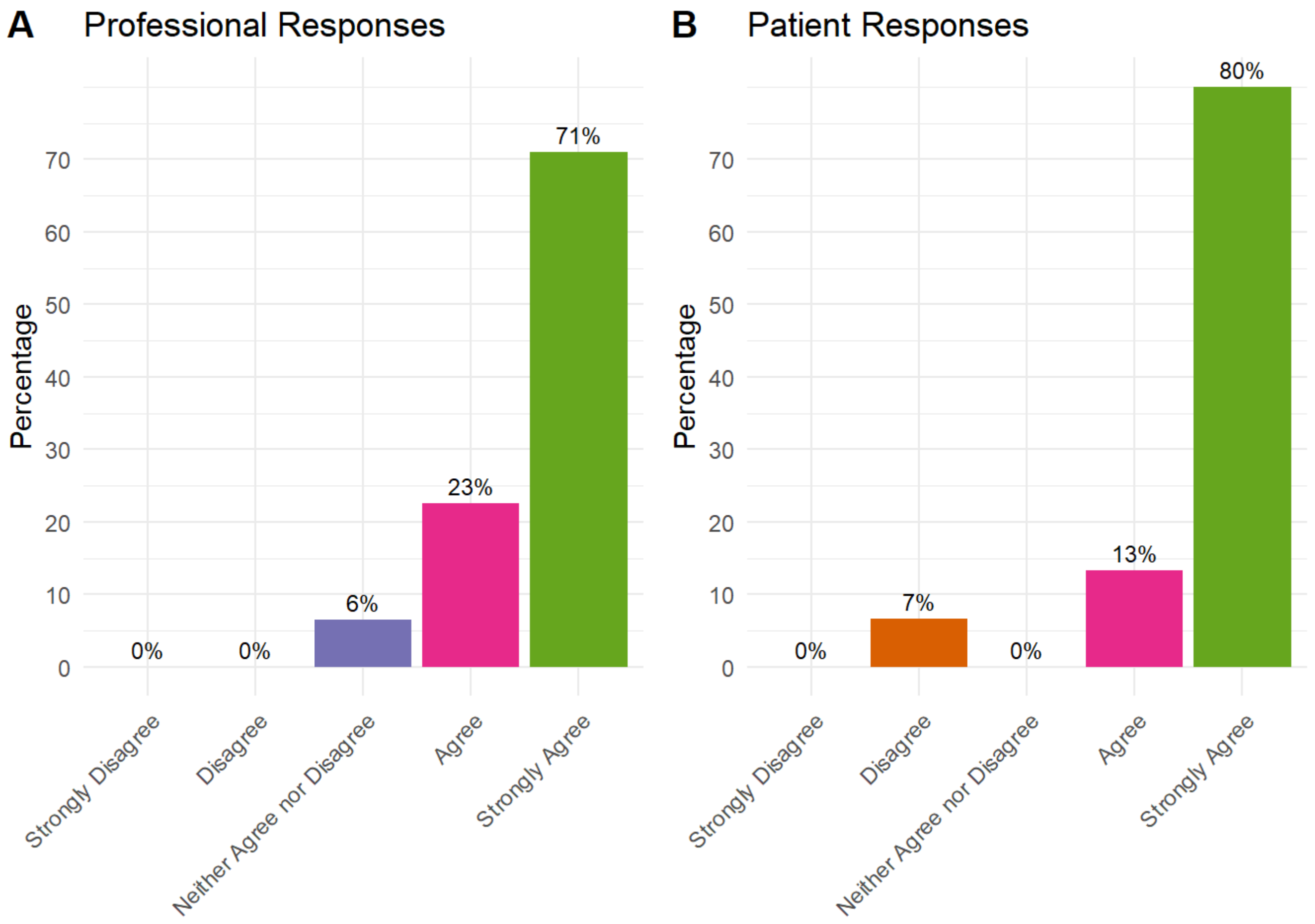


### Recovering from THR/TKR surgery should cover

Recovering from THR/TKR surgery should cover: Post-operative infection prevention

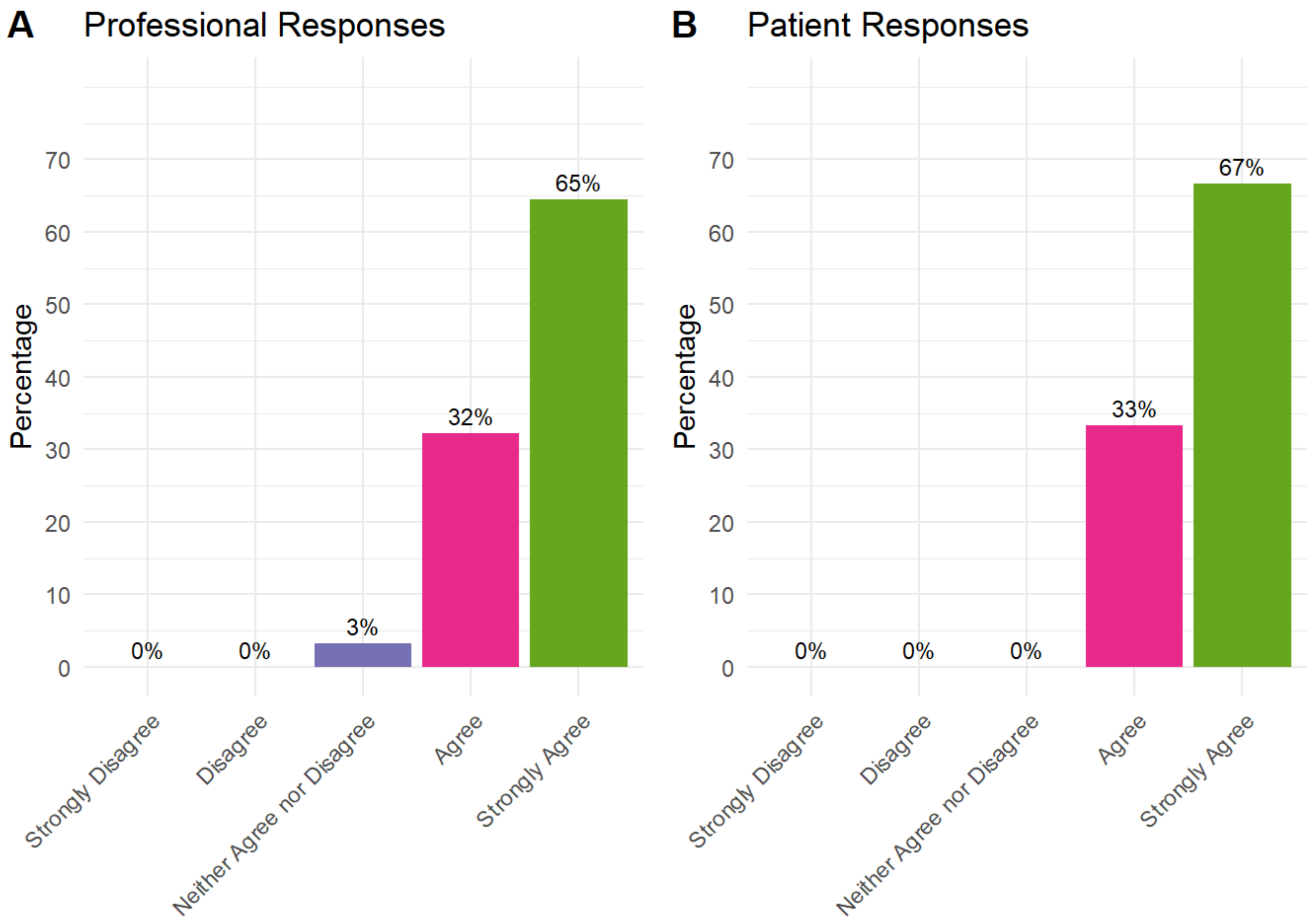


Recovering from THR/TKR surgery should cover: Organising help if complications occur

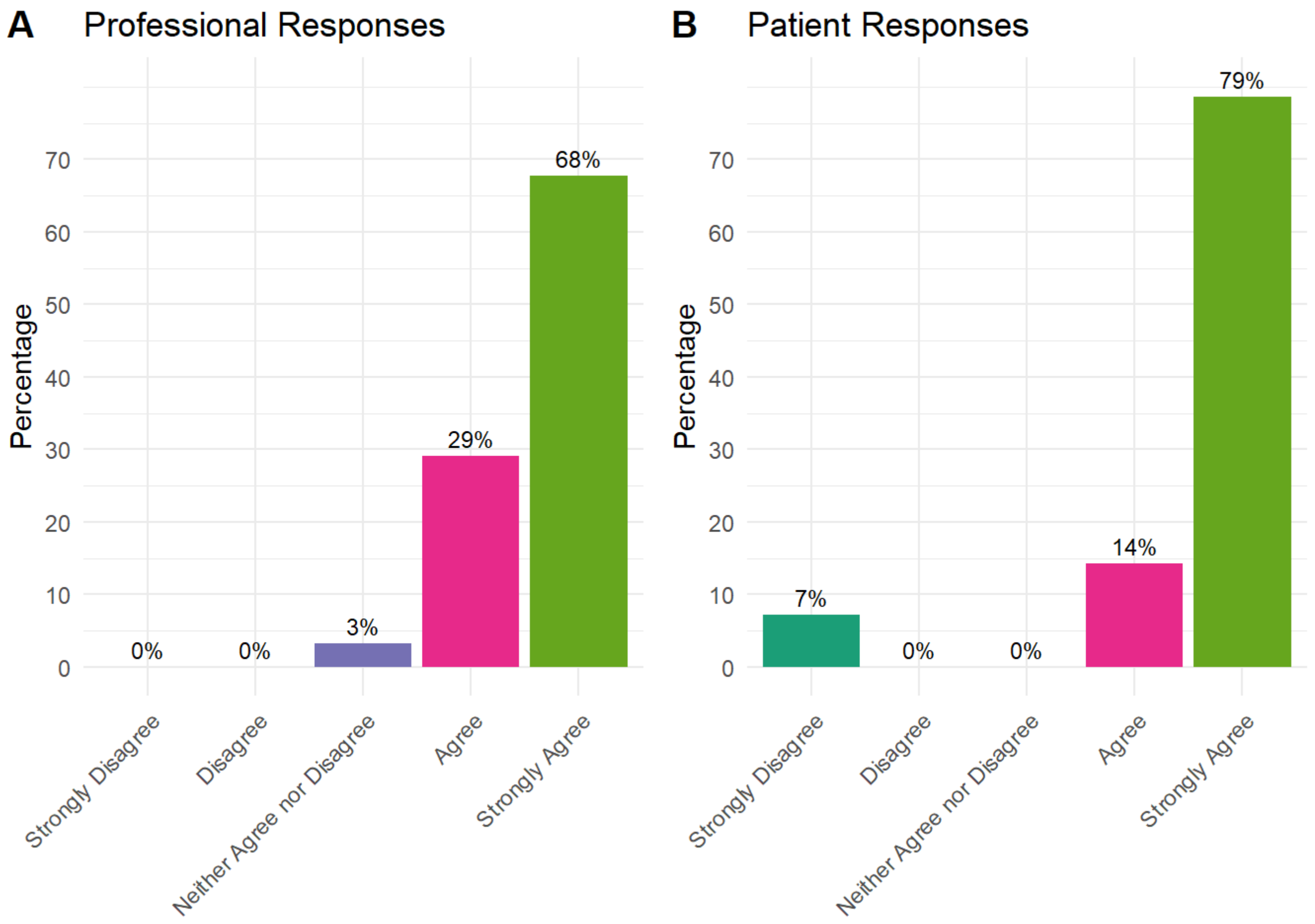


Recovering from THR/TKR surgery should cover: Pain management

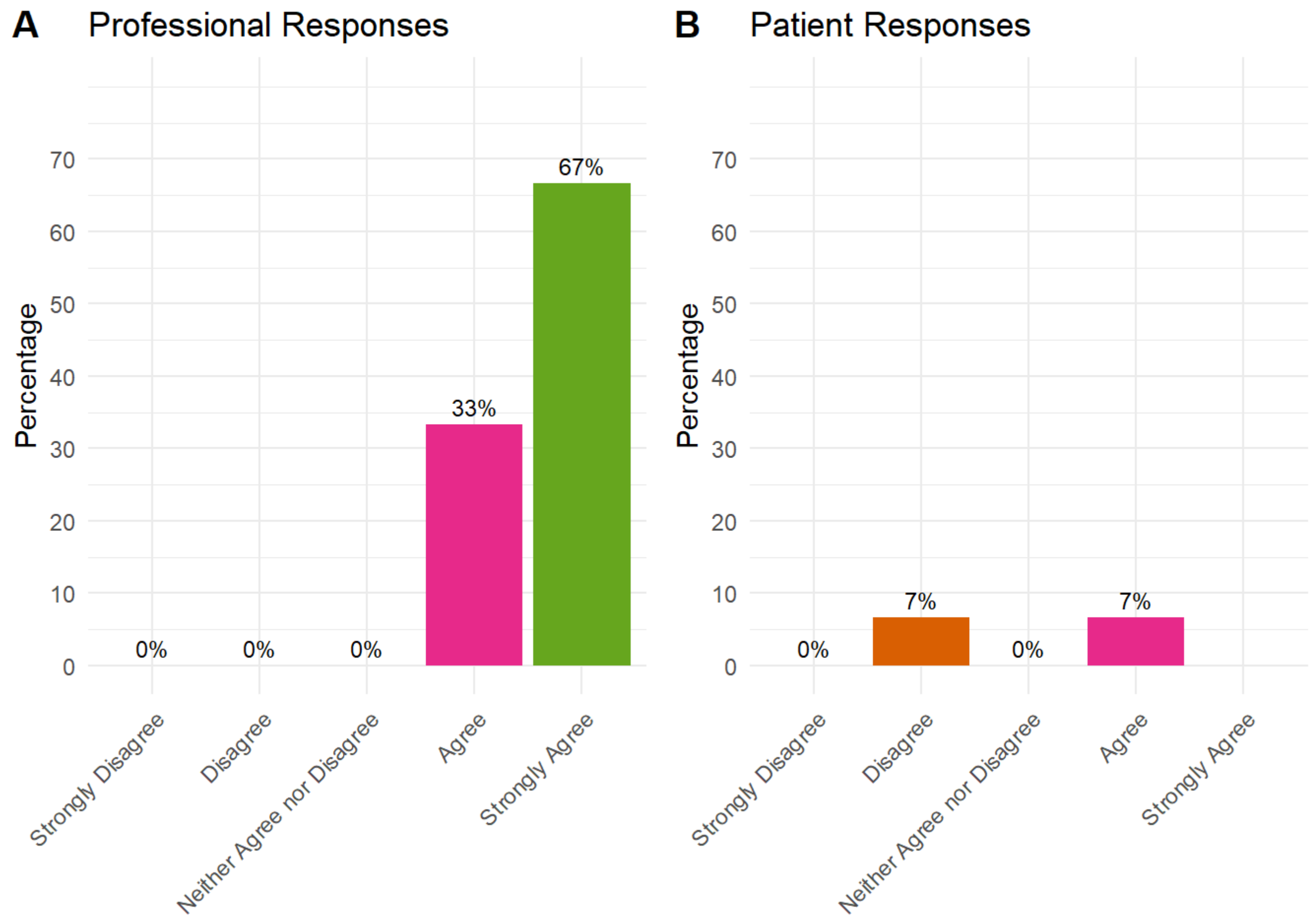


Recovering from THR/TKR surgery should cover: Nutrition

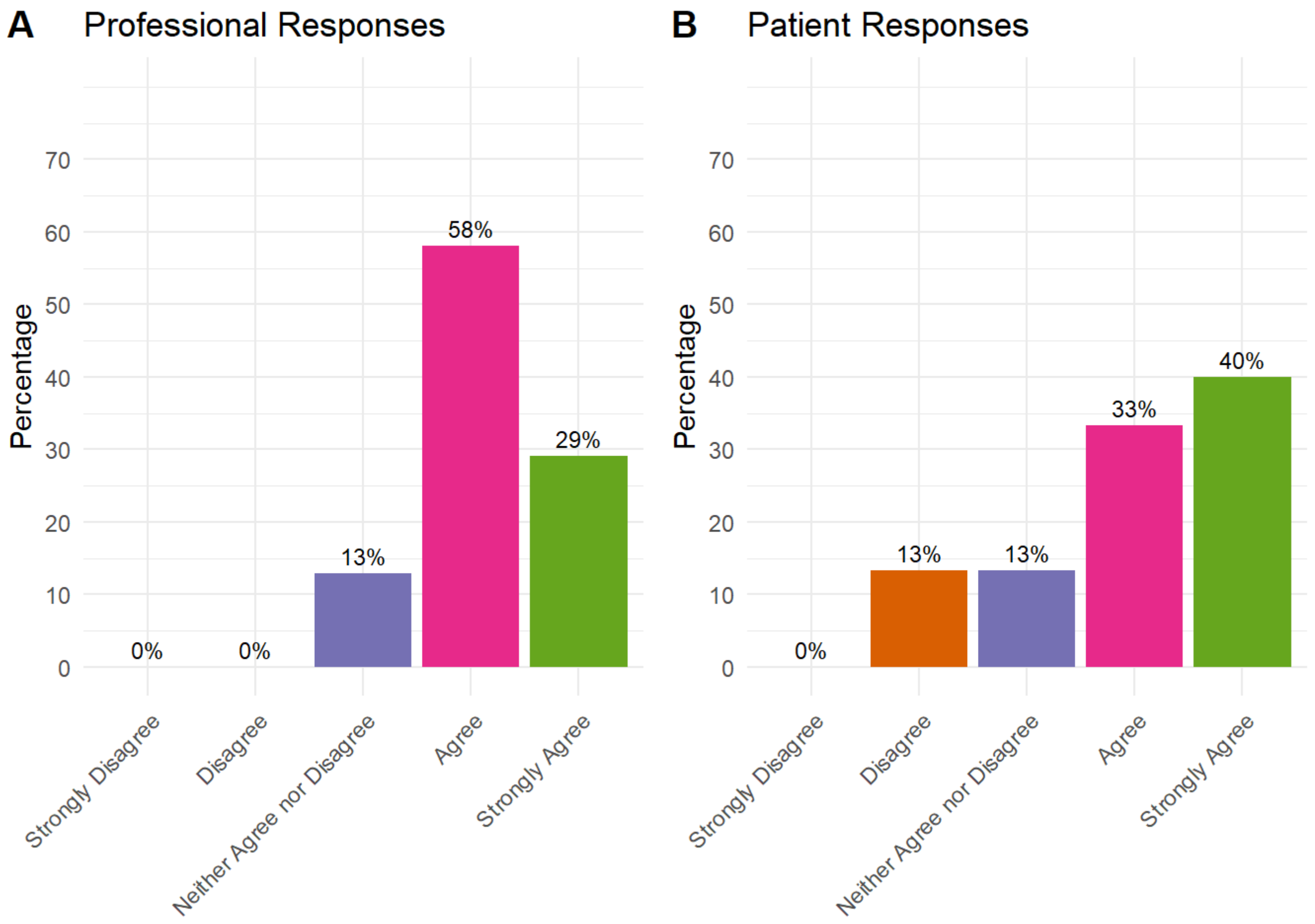


Recovering from THR/TKR surgery should cover: Precautions (e.g., movements/activities to avoid in early post-operative period)

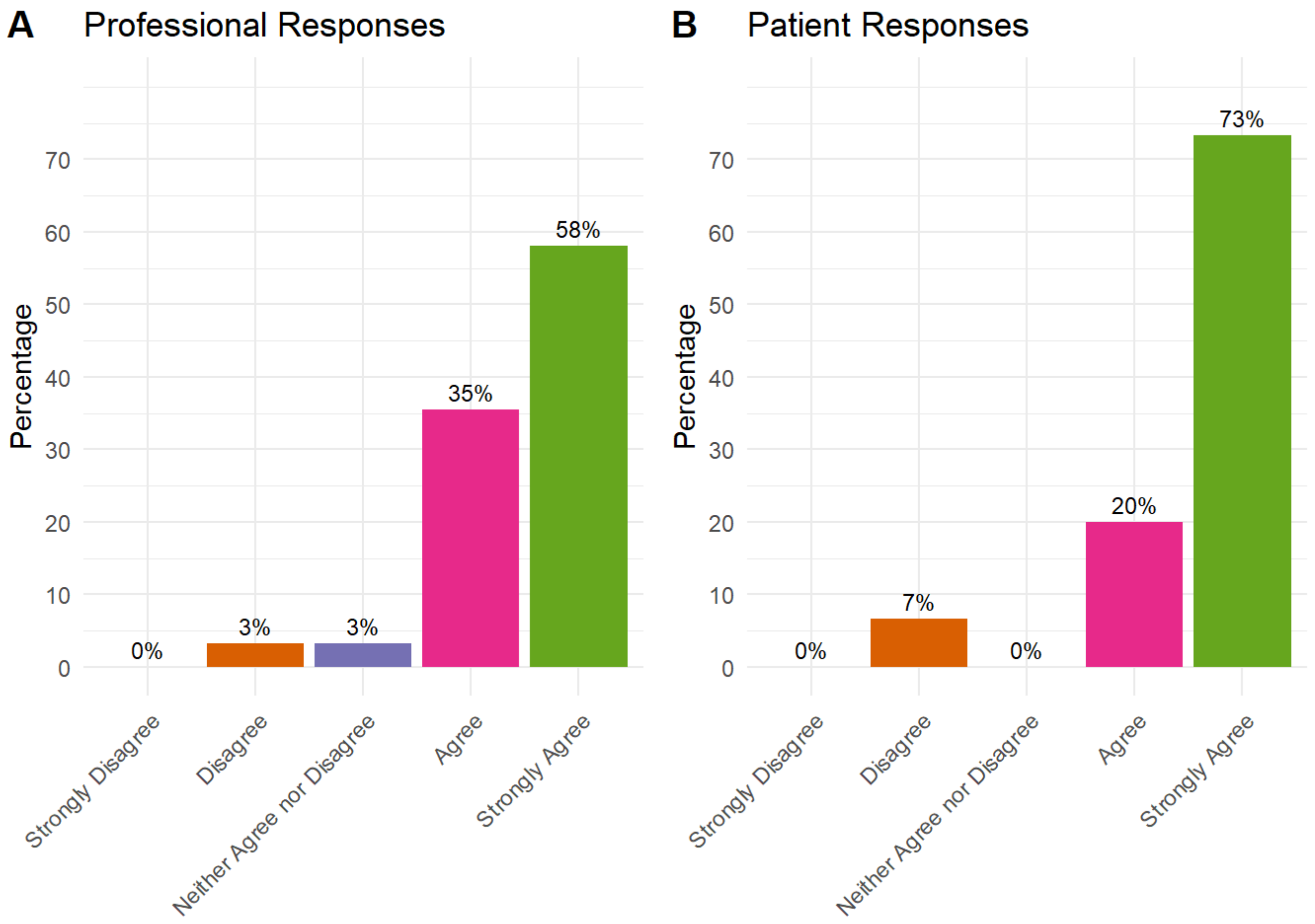


Recovering from THR/TKR surgery should cover: Rehabilitation following THR/TKR surgery

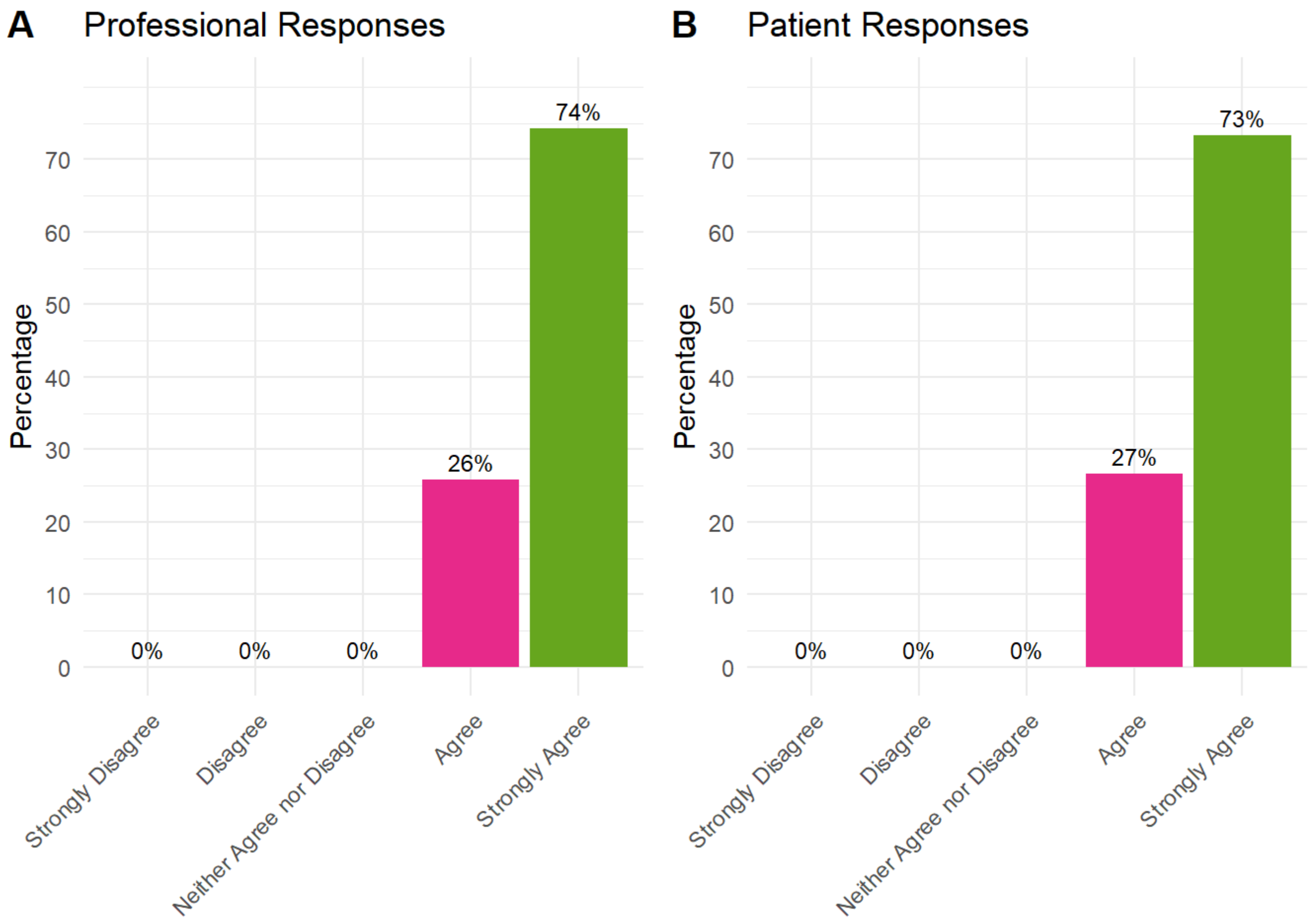


Recovering from THR/TKR surgery should cover: Returning to daily activities

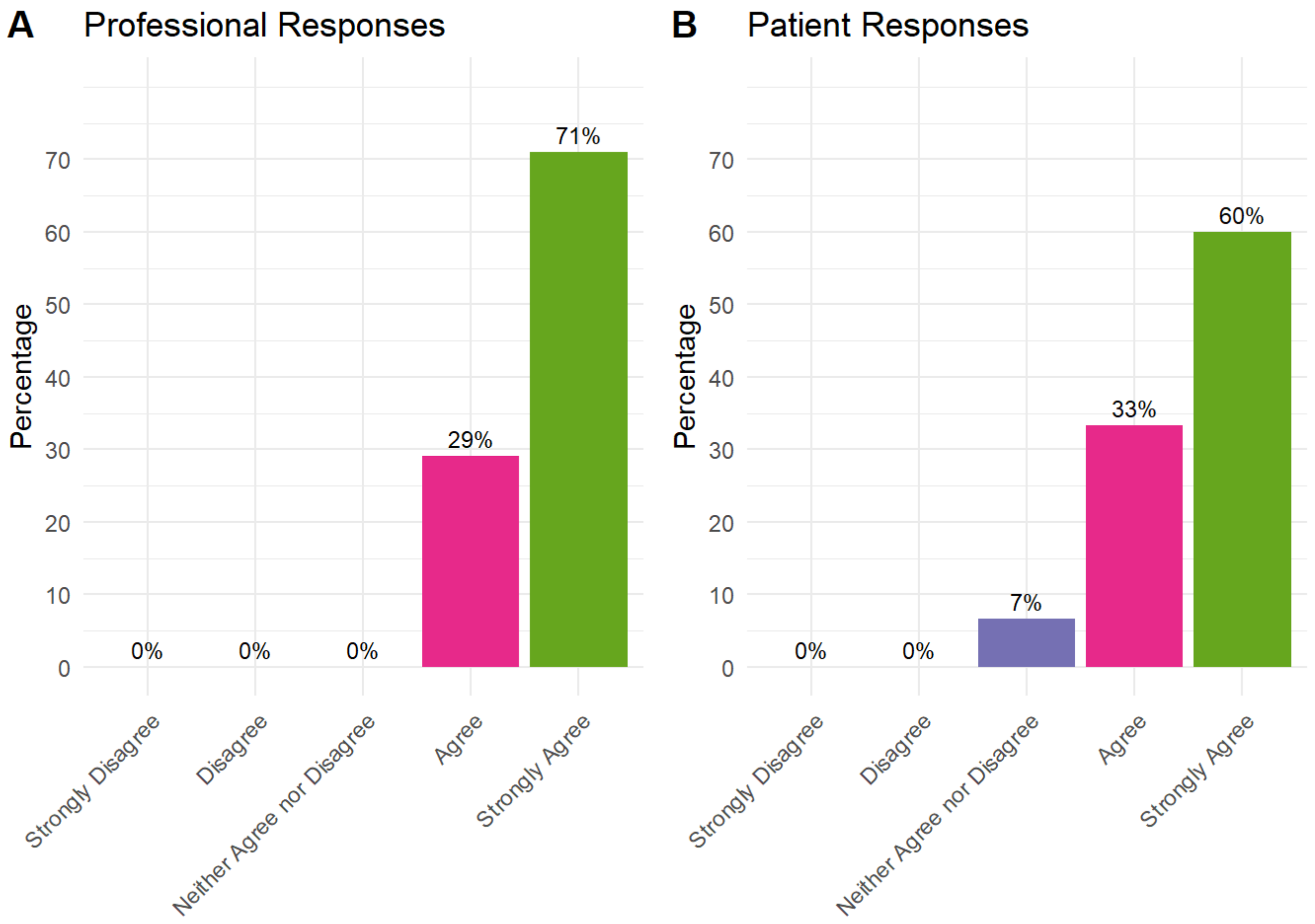


Recovering from THR/TKR surgery should cover: Returning to a normal walking pattern

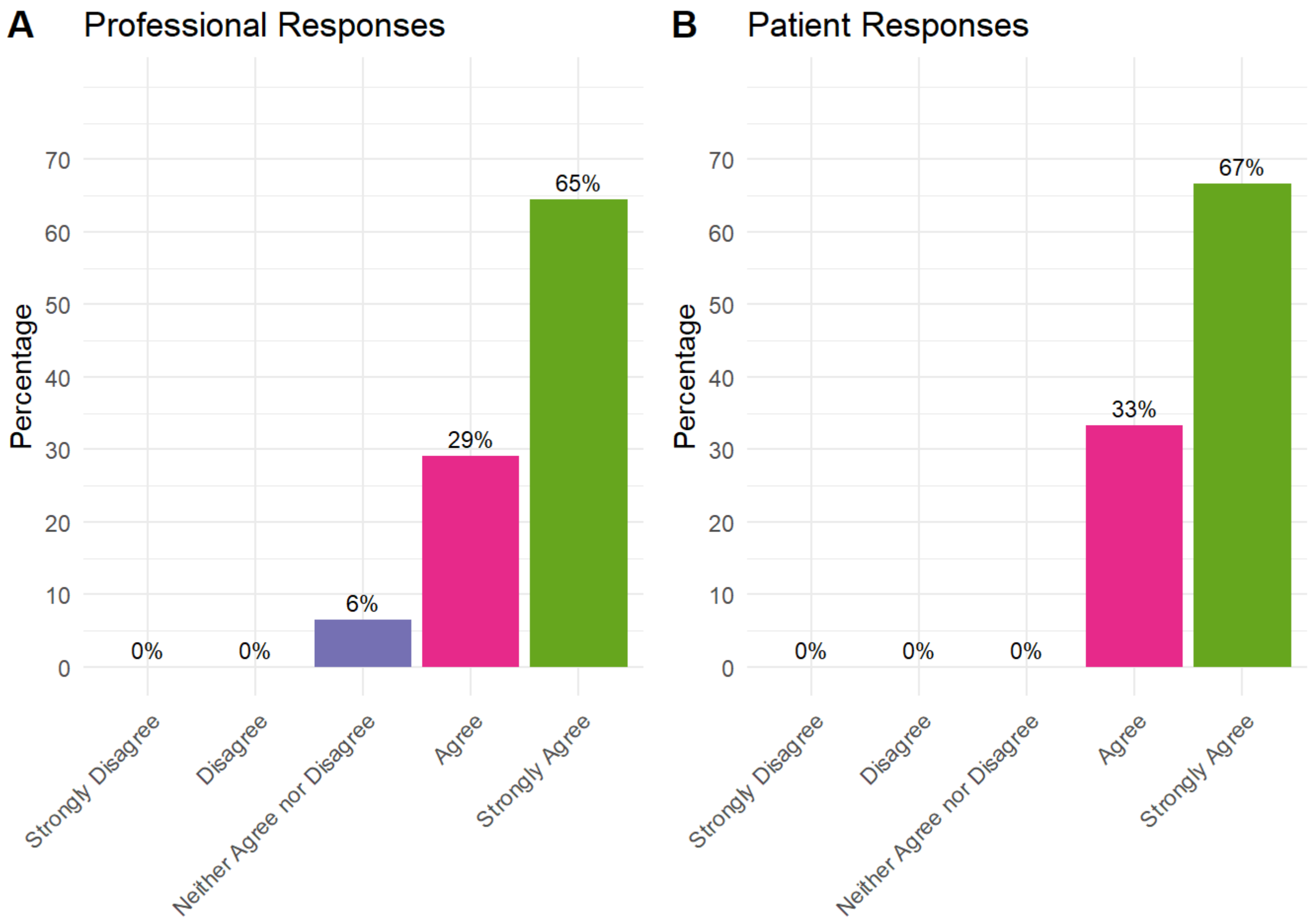


Recovering from THR/TKR surgery should cover: Returning to driving and other types of travel

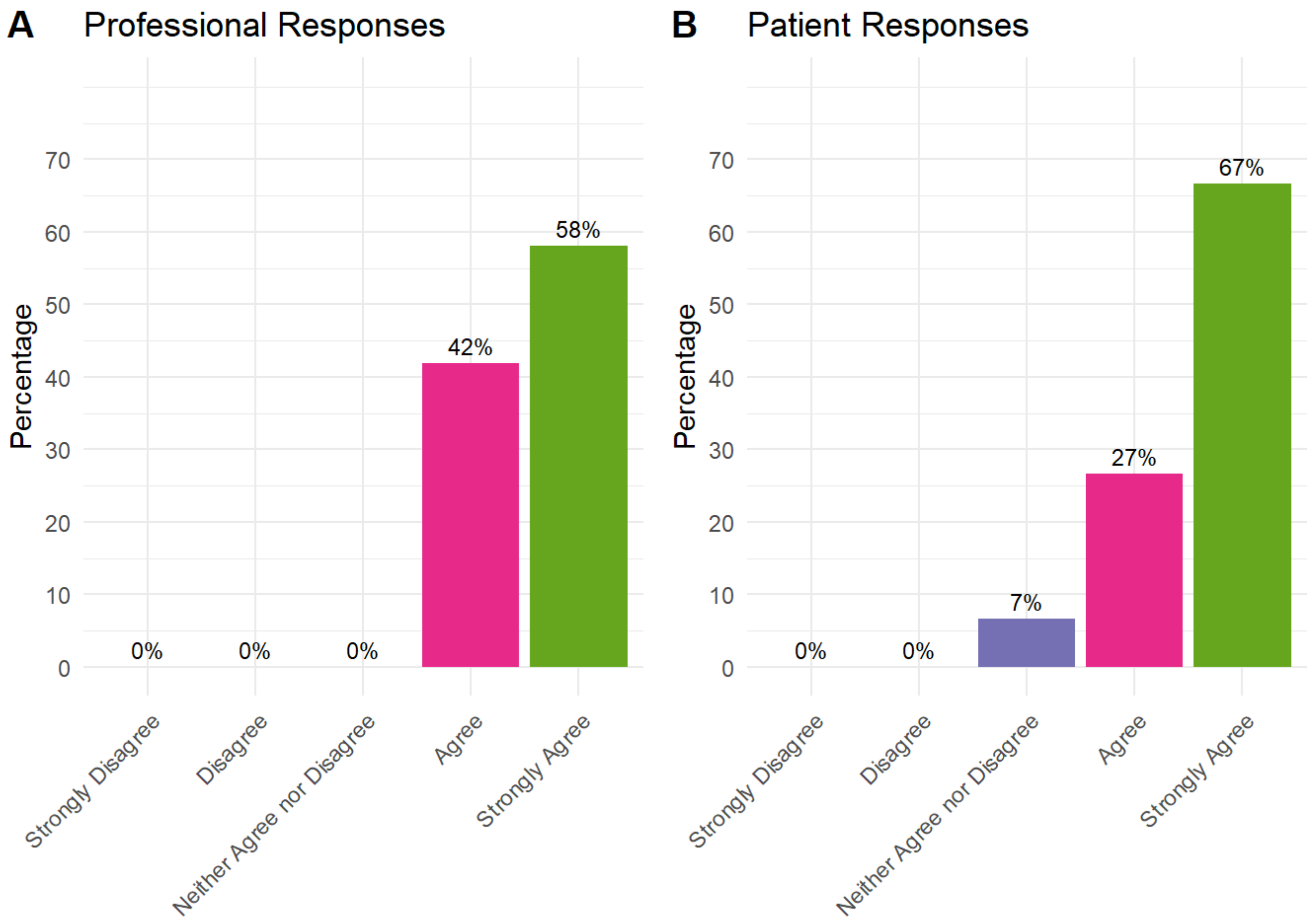


Recovering from THR/TKR surgery should cover: Returning to sports and leisure activities

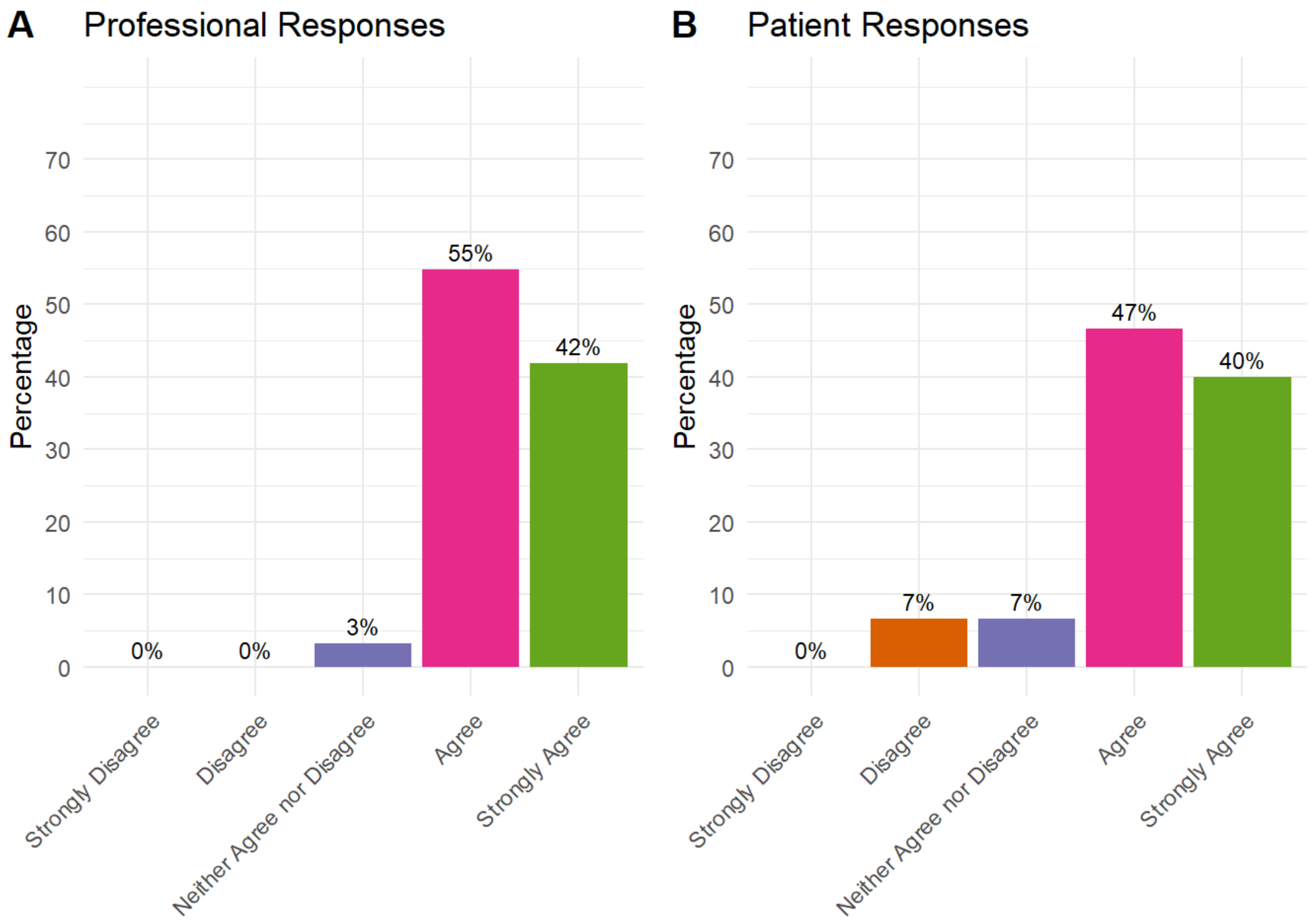


Recovering from THR/TKR surgery should cover: Returning to work

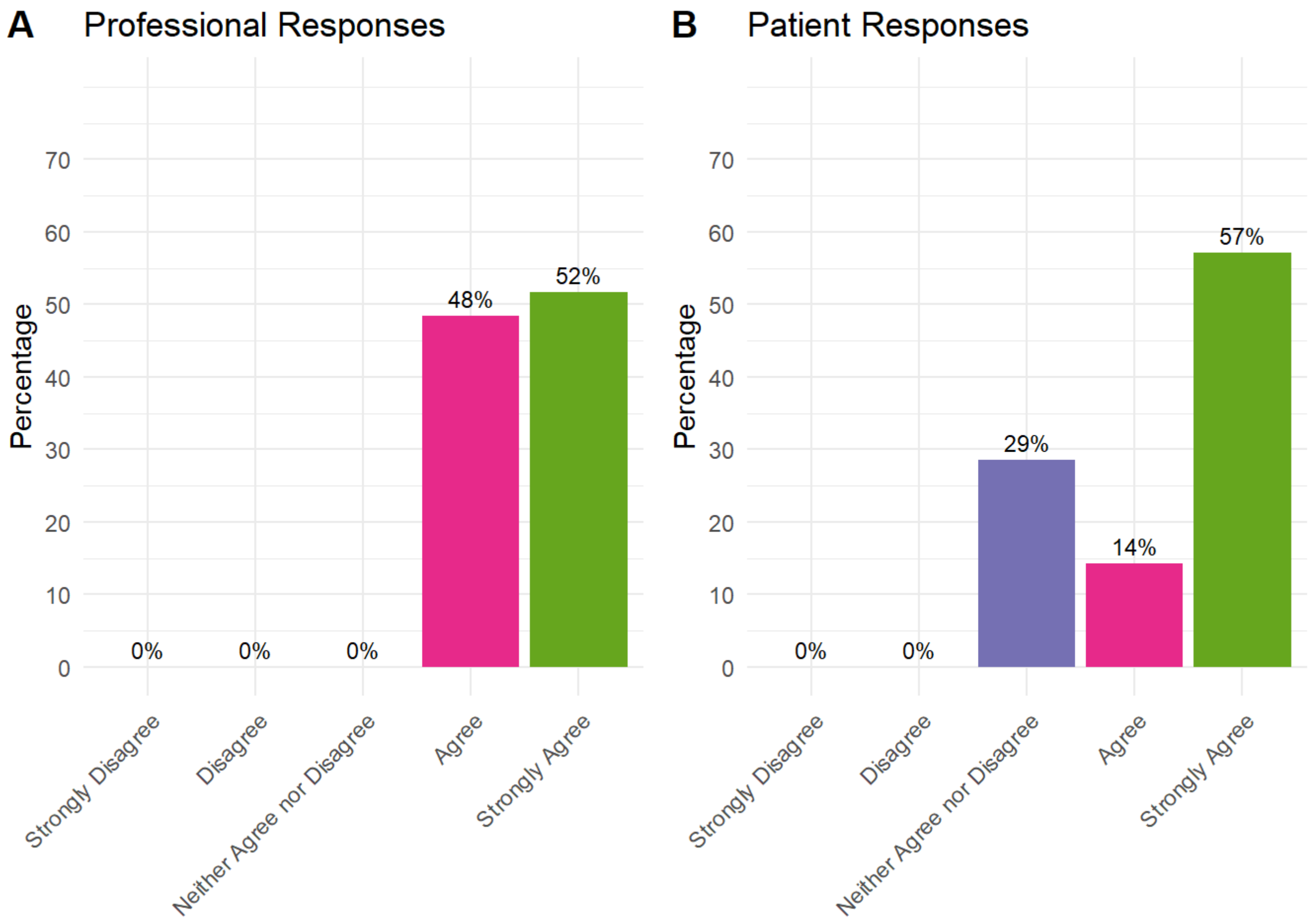


Recovering from THR/TKR surgery should cover: Realistic timelines for recovery and activities

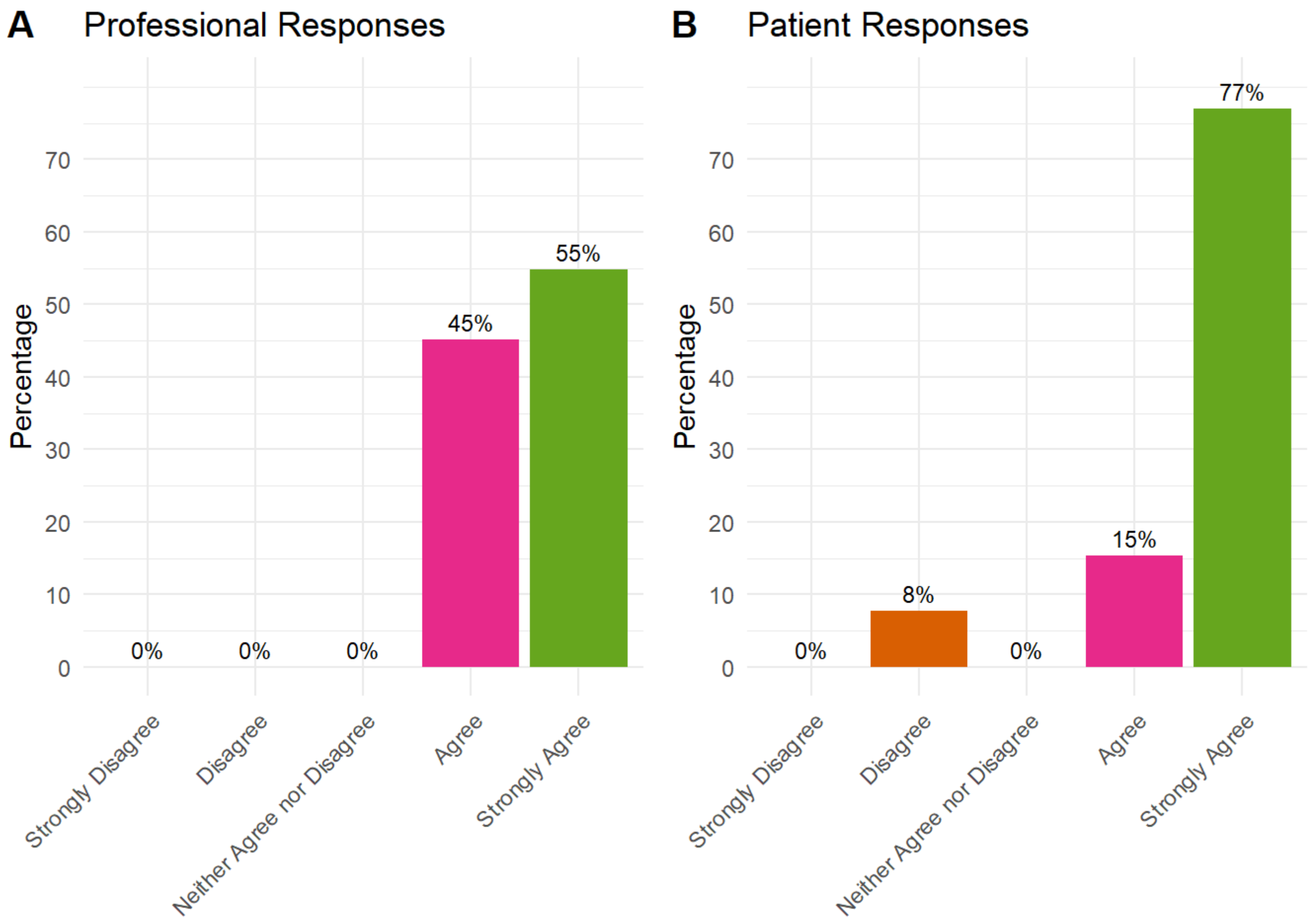


Recovering from THR/TKR surgery should cover: When to ask for help (e.g. wound care)

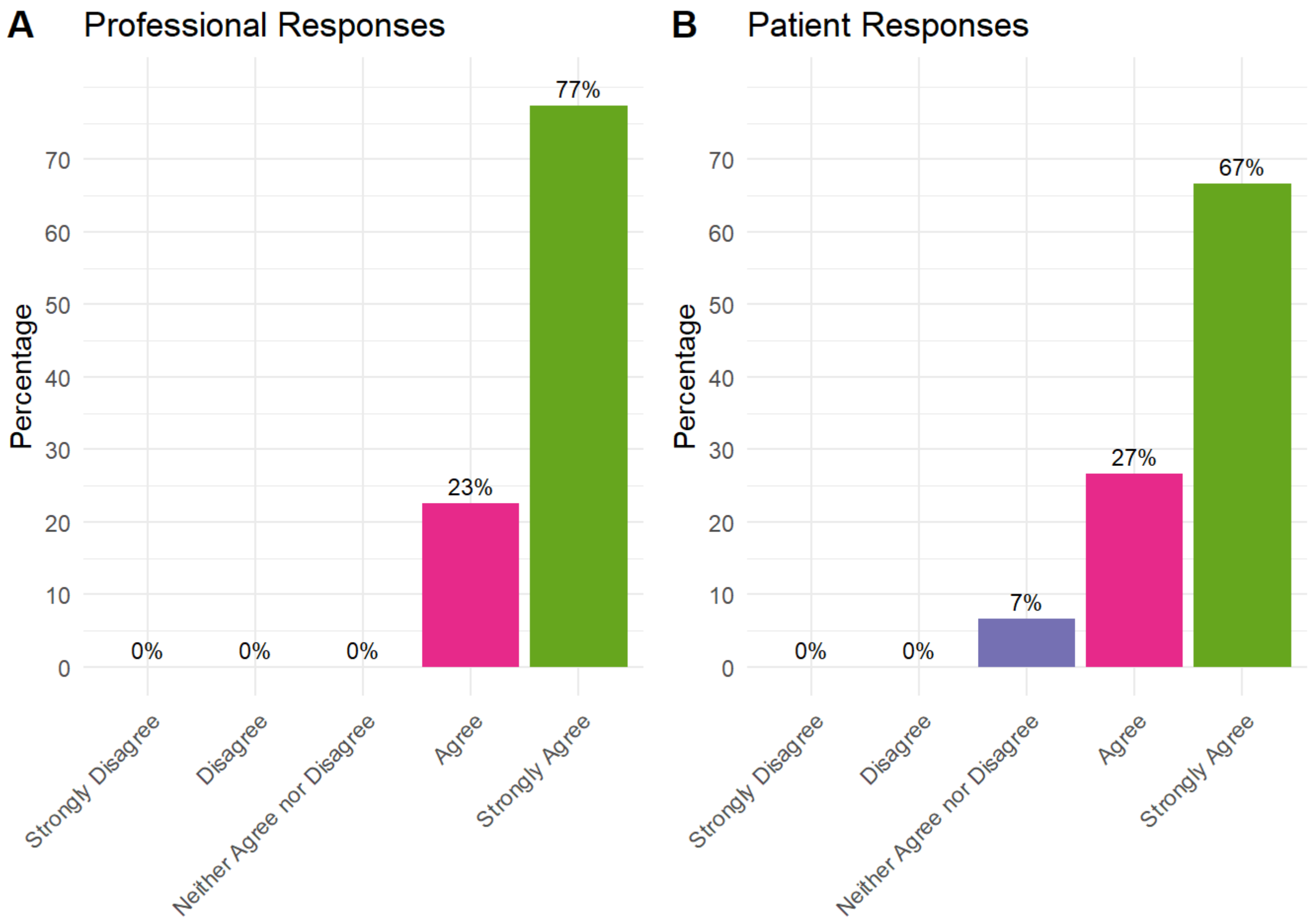


### Healthy lifestyle guidance should cover

Healthy lifestyle guidance should cover: Physical activity

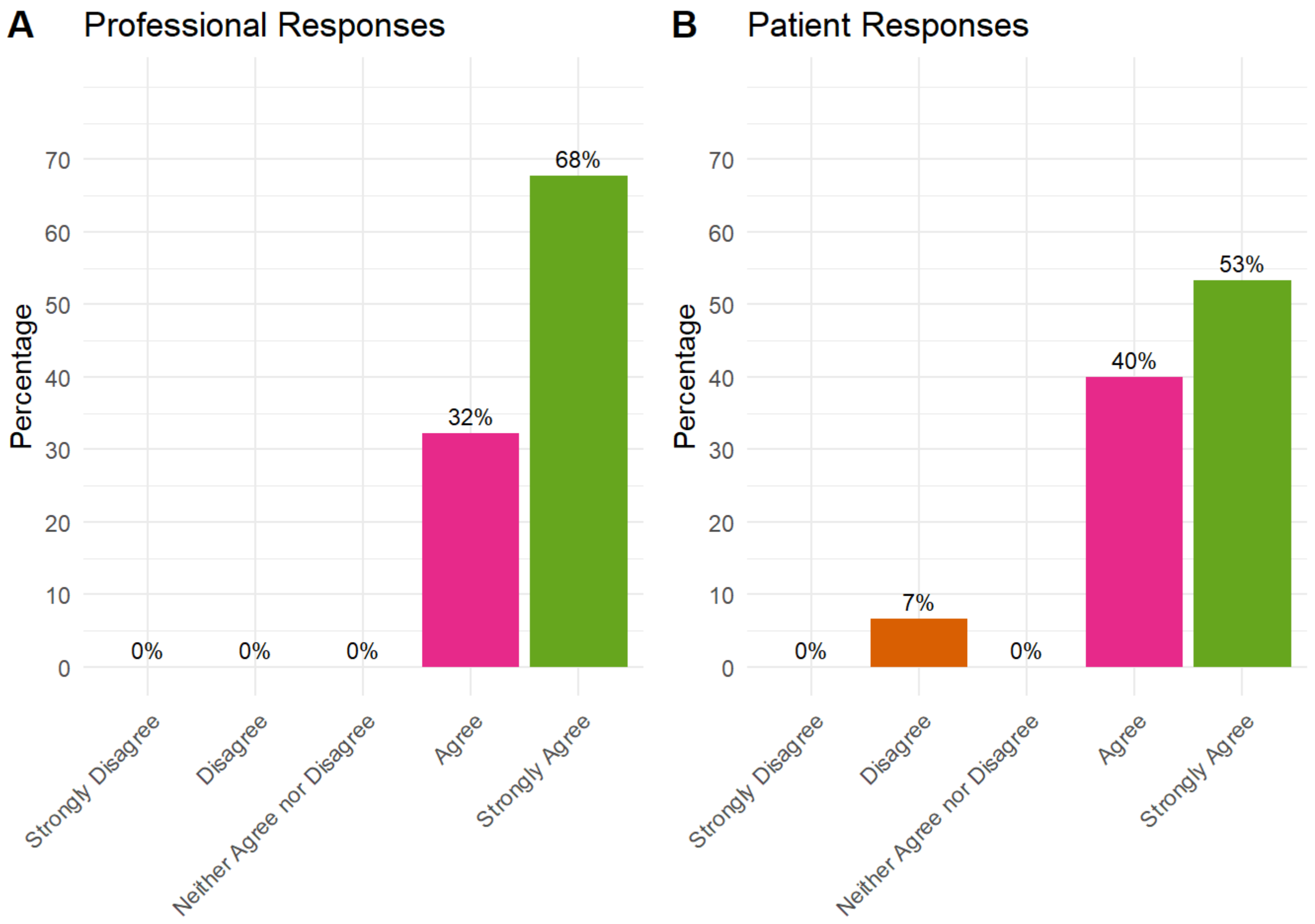


Healthy lifestyle guidance should cover: Weight management

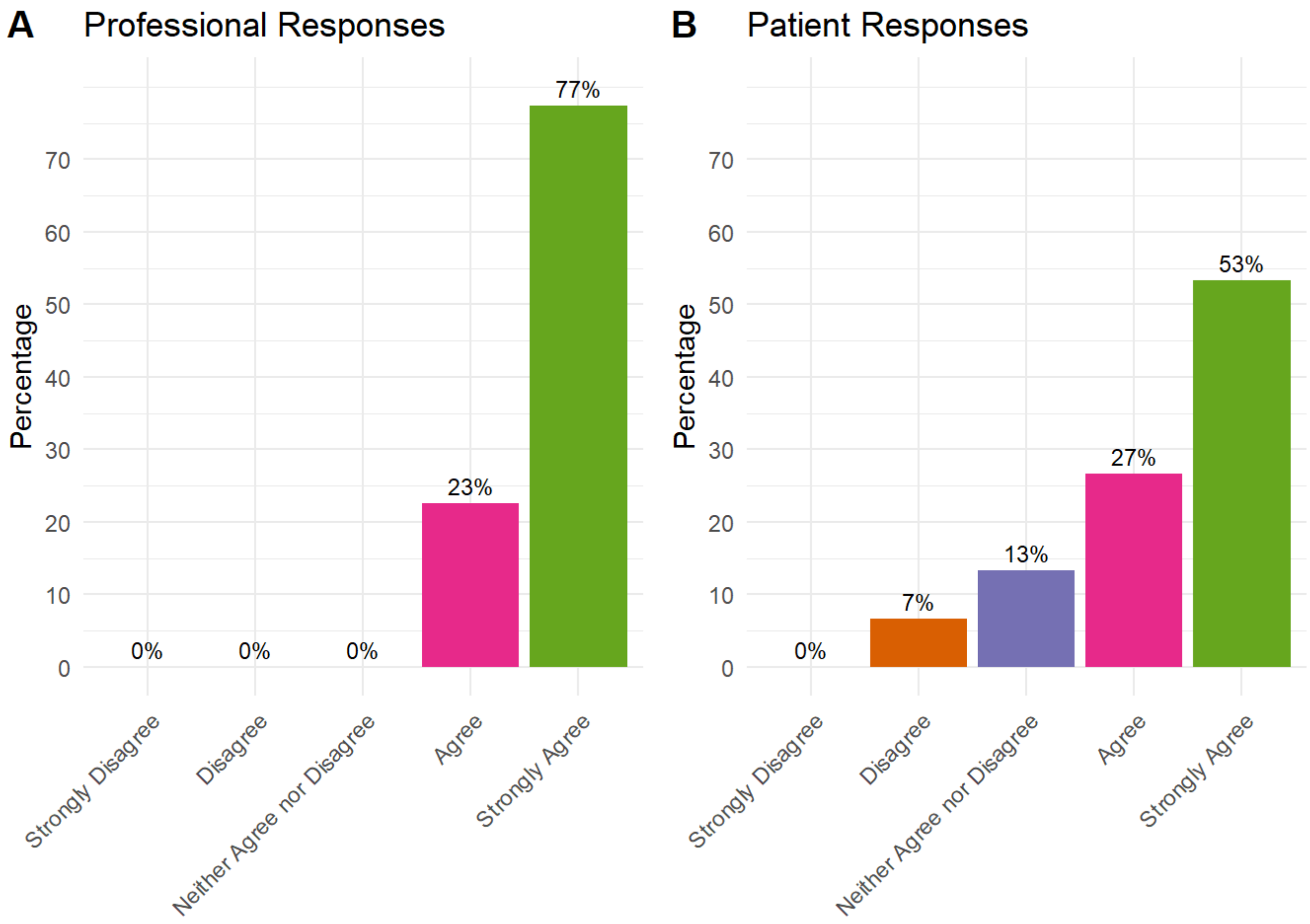


Healthy lifestyle guidance should cover: Stopping smoking

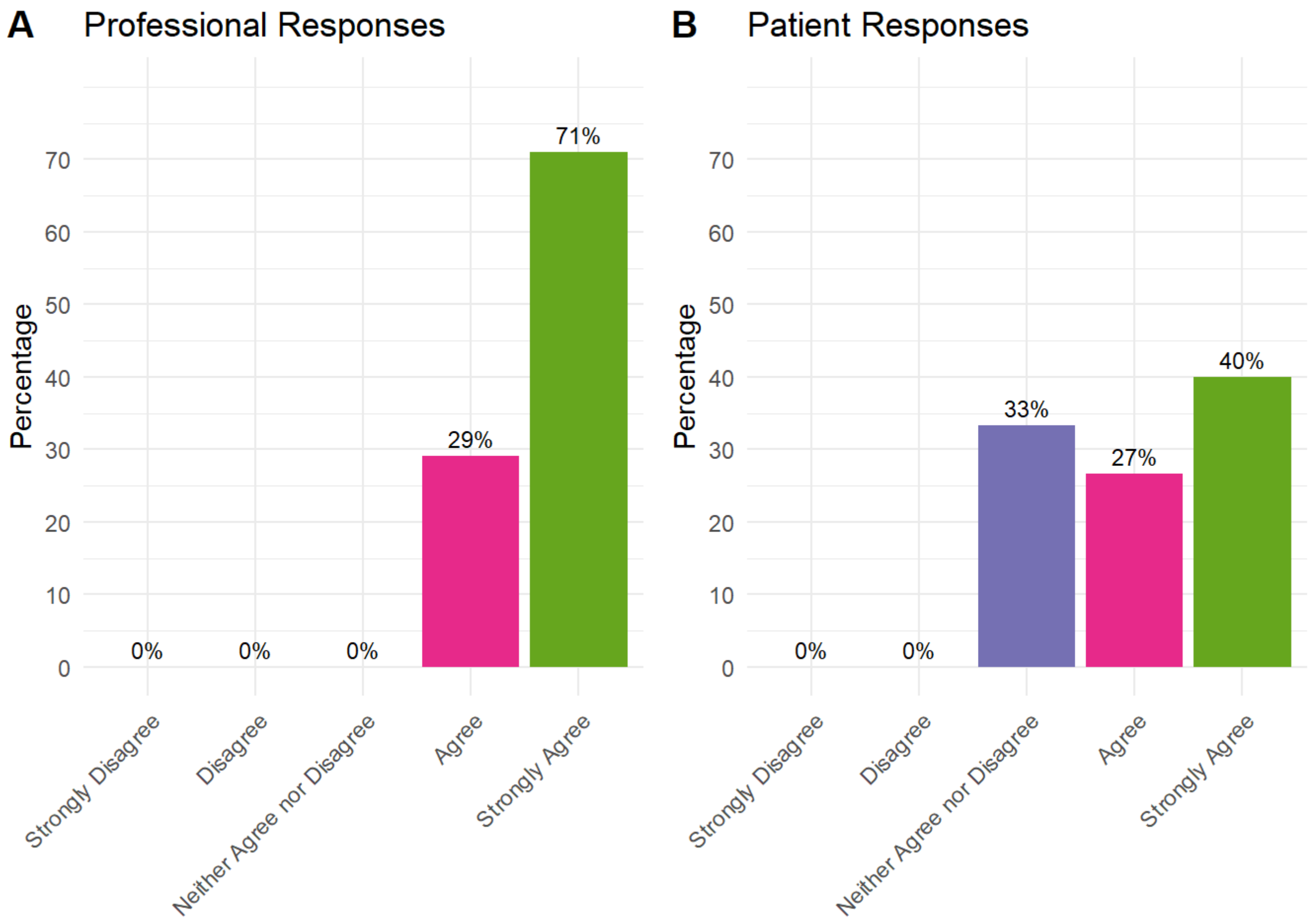


Healthy lifestyle guidance should cover: Recommended alcohol guidelines and cutting down on drinking

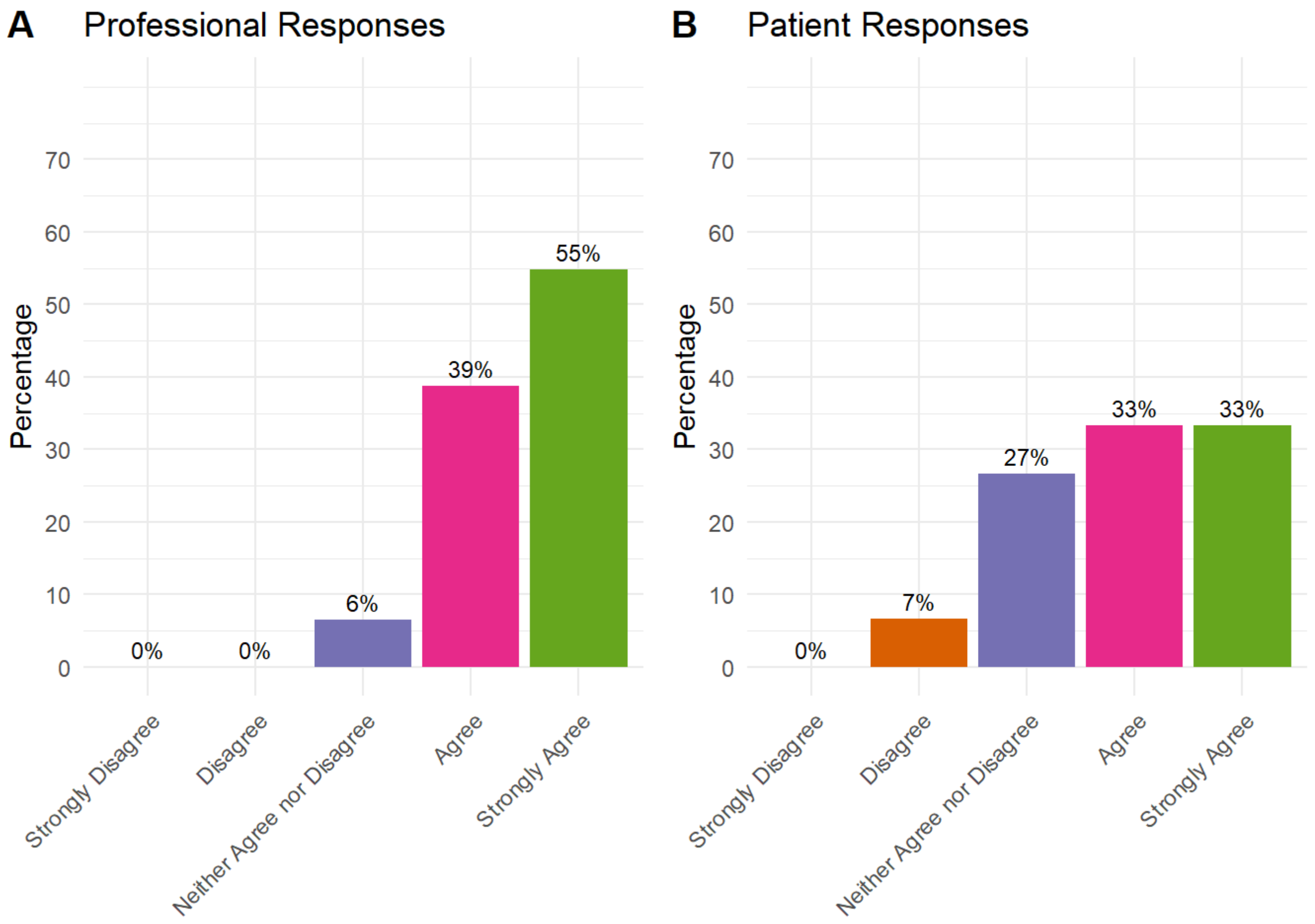


### Delivery of pre-operative education

Delivery of pre-operative education: Be informed by a multi-disciplinary team, including members of the orthopaedic surgery team, nursing, physiotherapy, and occupational therapy teams

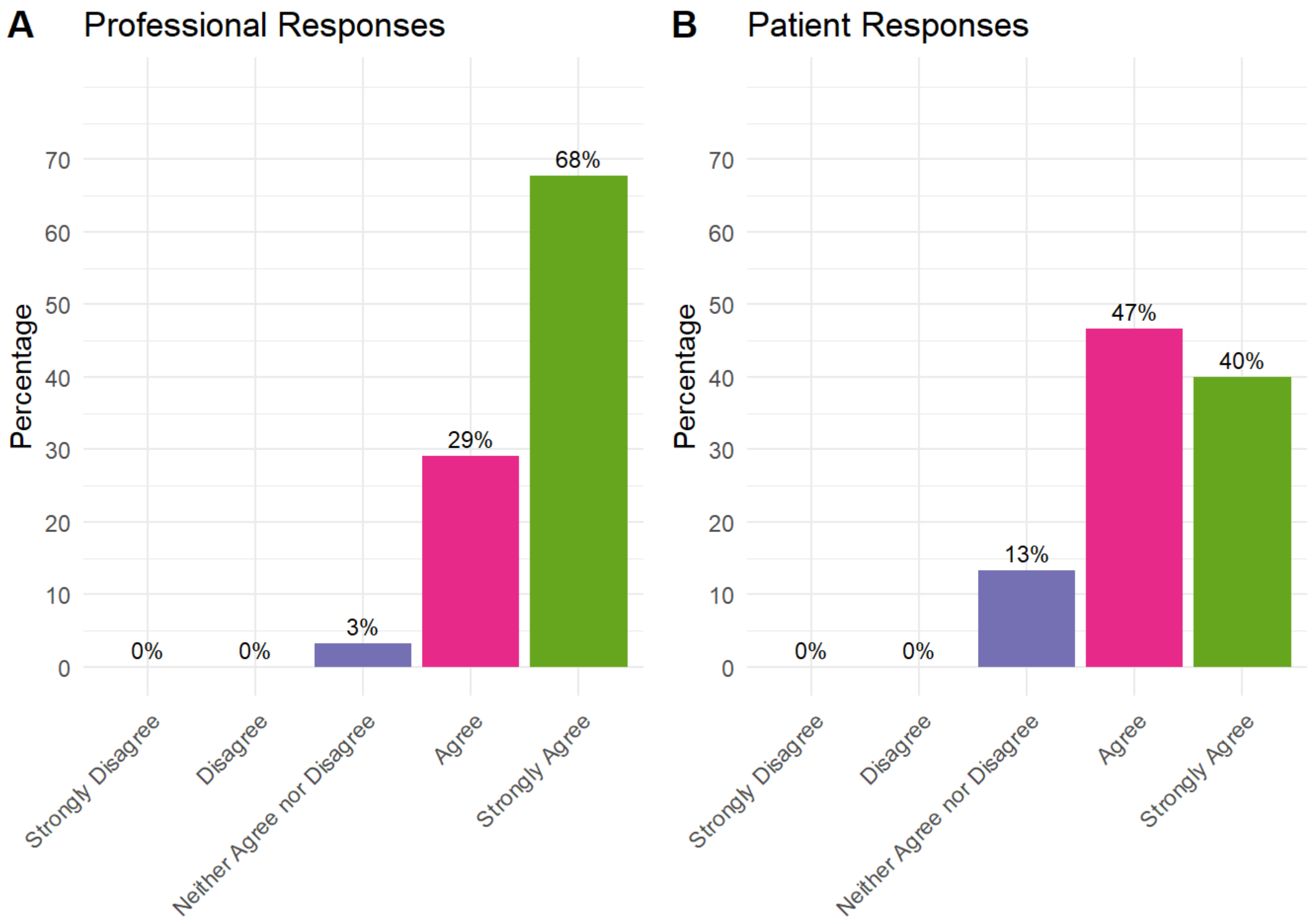


Delivery of pre-operative education: Be informed by patients who have previously had THR/TKR surgery

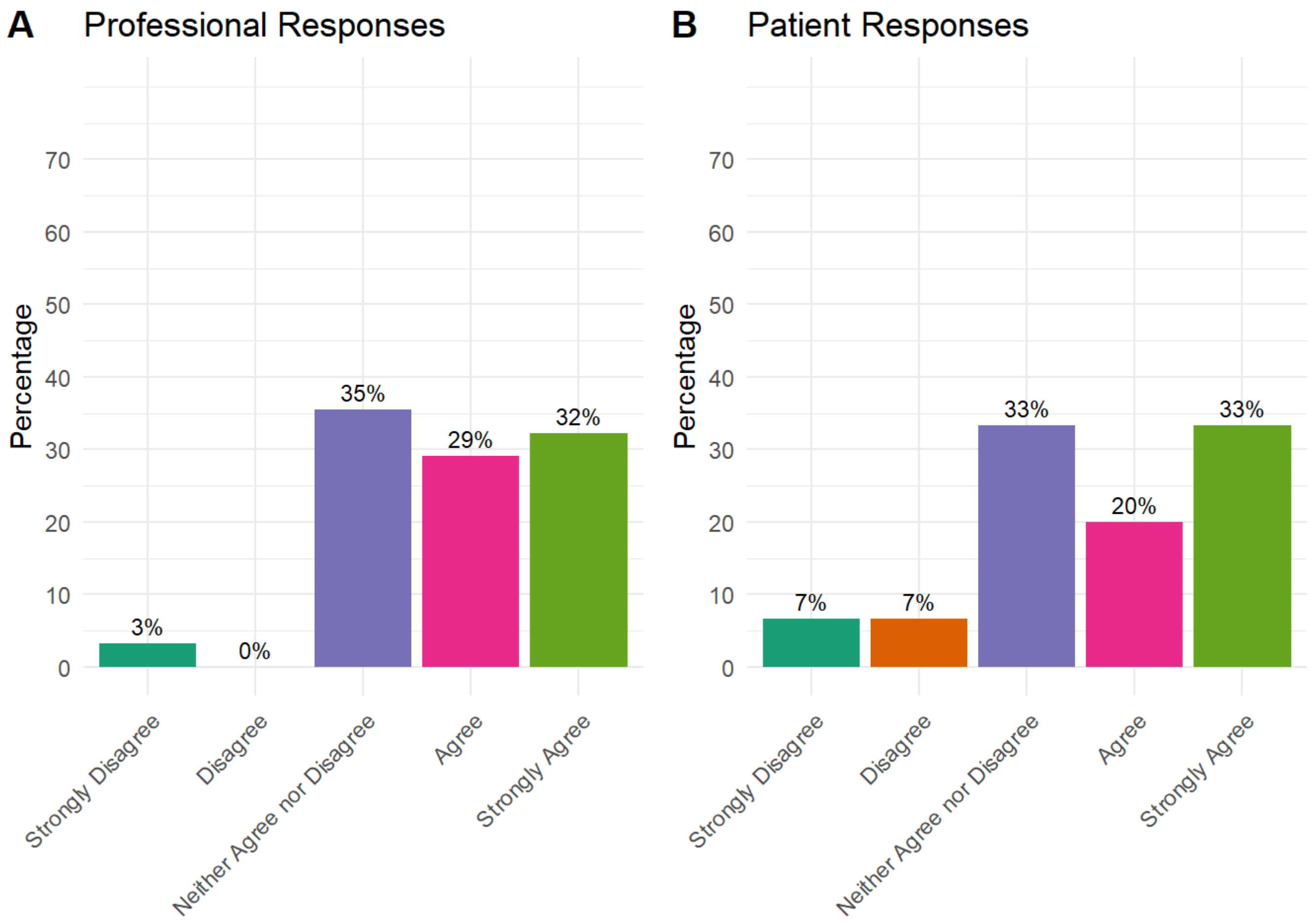


### Pre-operative information should

Pre-operative information should: Be delivered, at least partly, by providing examples of other patients' experiences of THR/TKR surgery

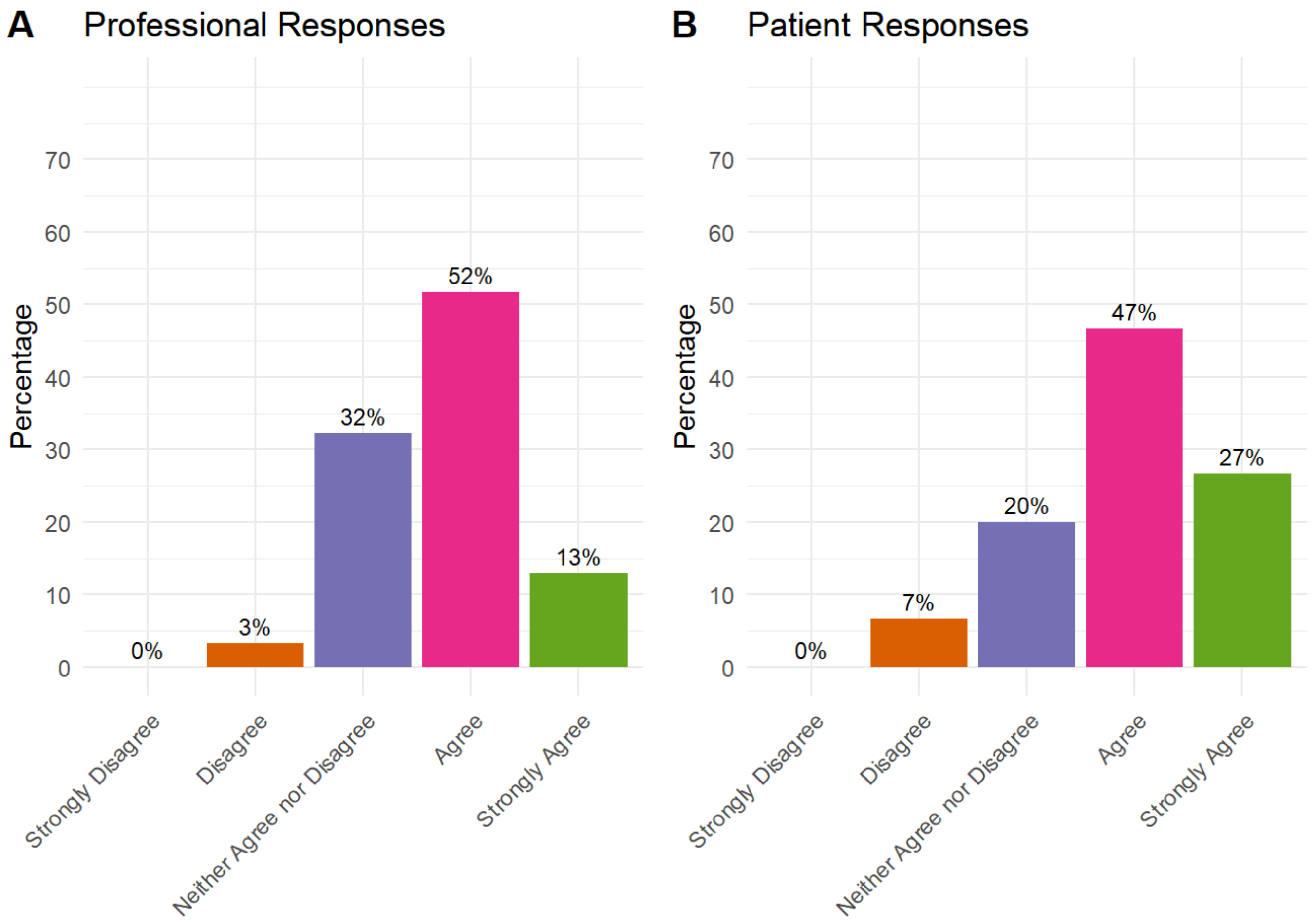


Pre-operative information should: Be delivered 1-1 if the patient does not wish to attend a group session

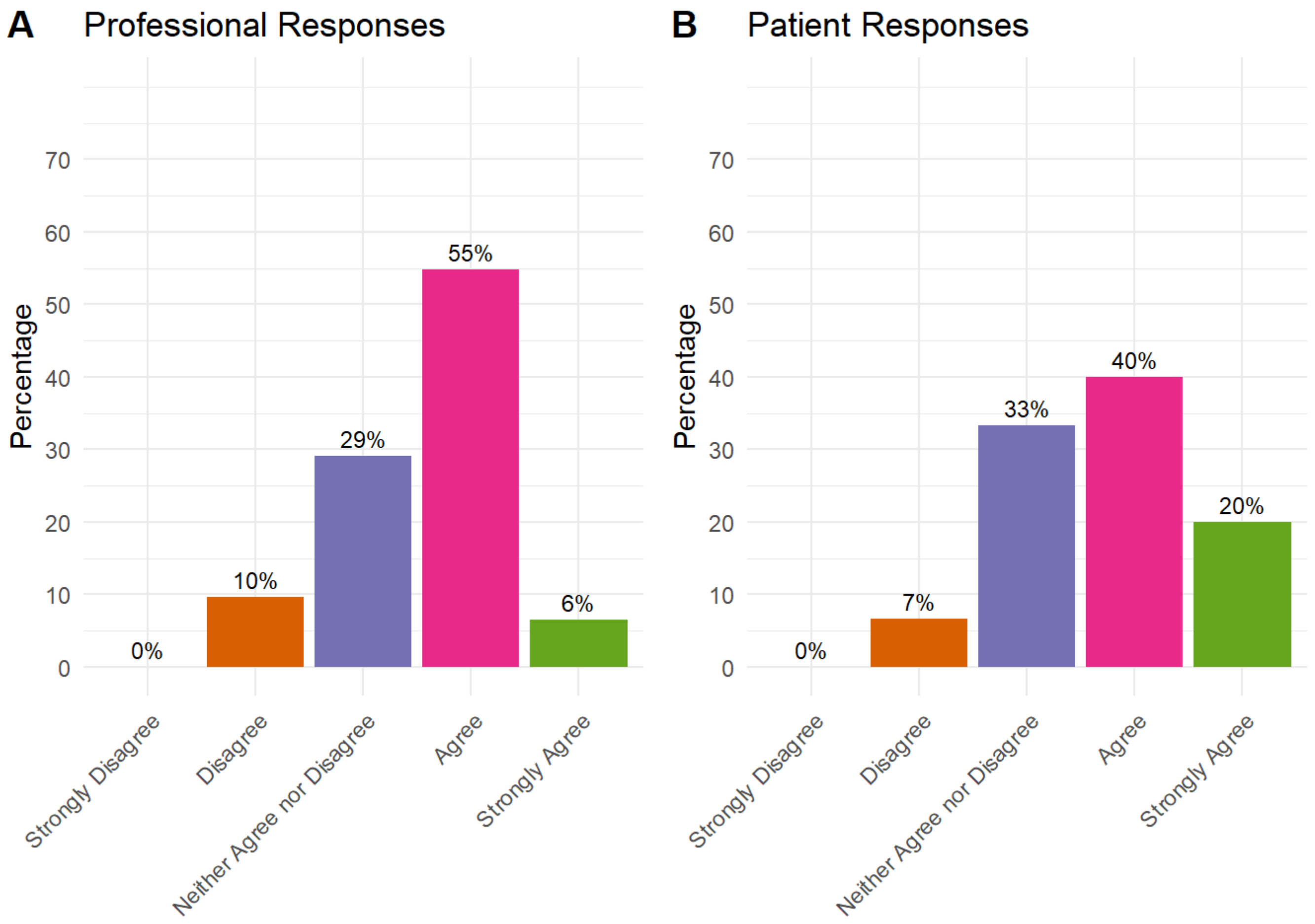


Pre-operative information should: Be delivered using a combination of more than one format, including face to face group sessions, a booklet or other written format and a website or other electronic format

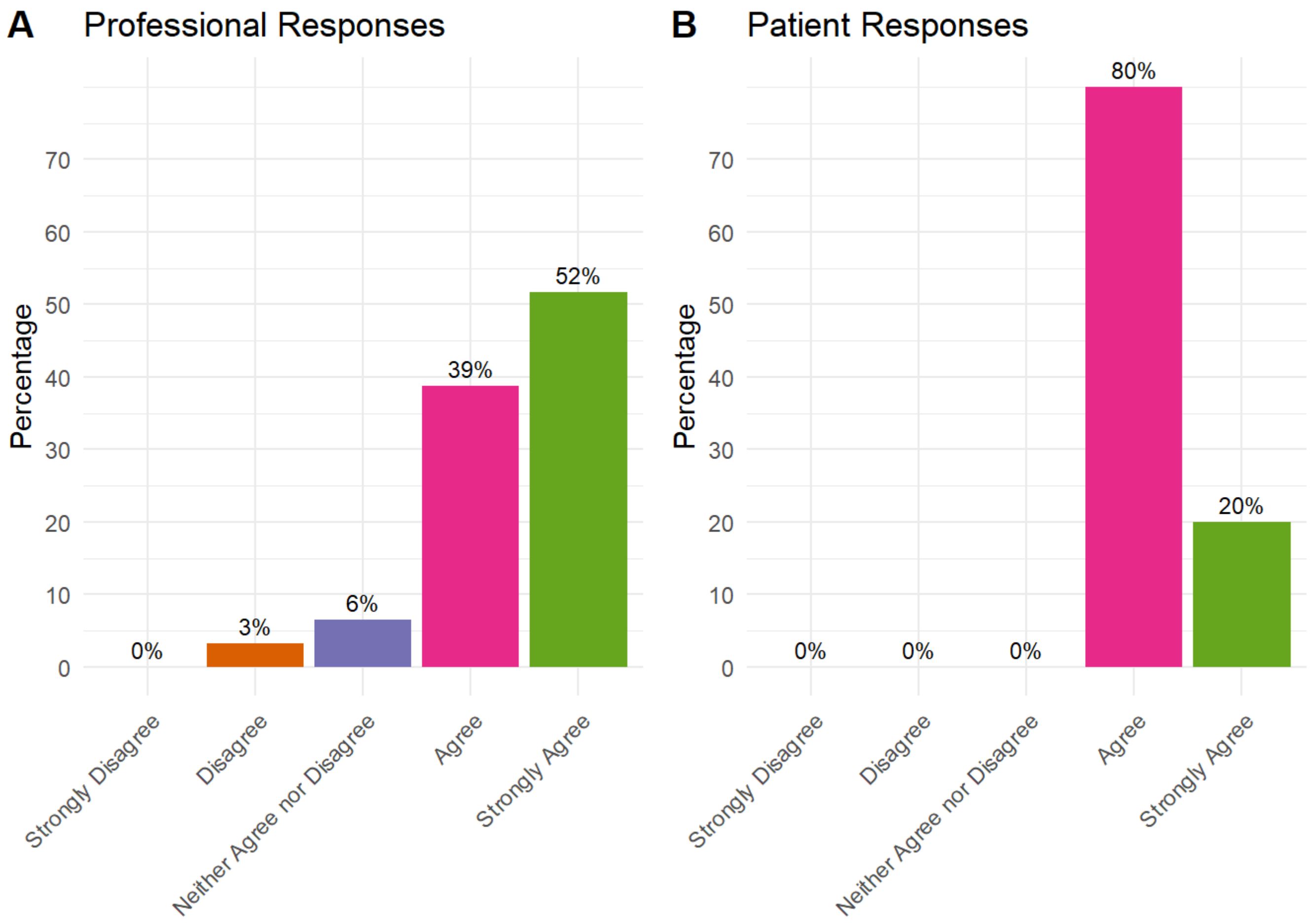


Pre-operative information should: Be delivered through a combination of providing the patient with information and giving them an opportunity to actively take part in tasks

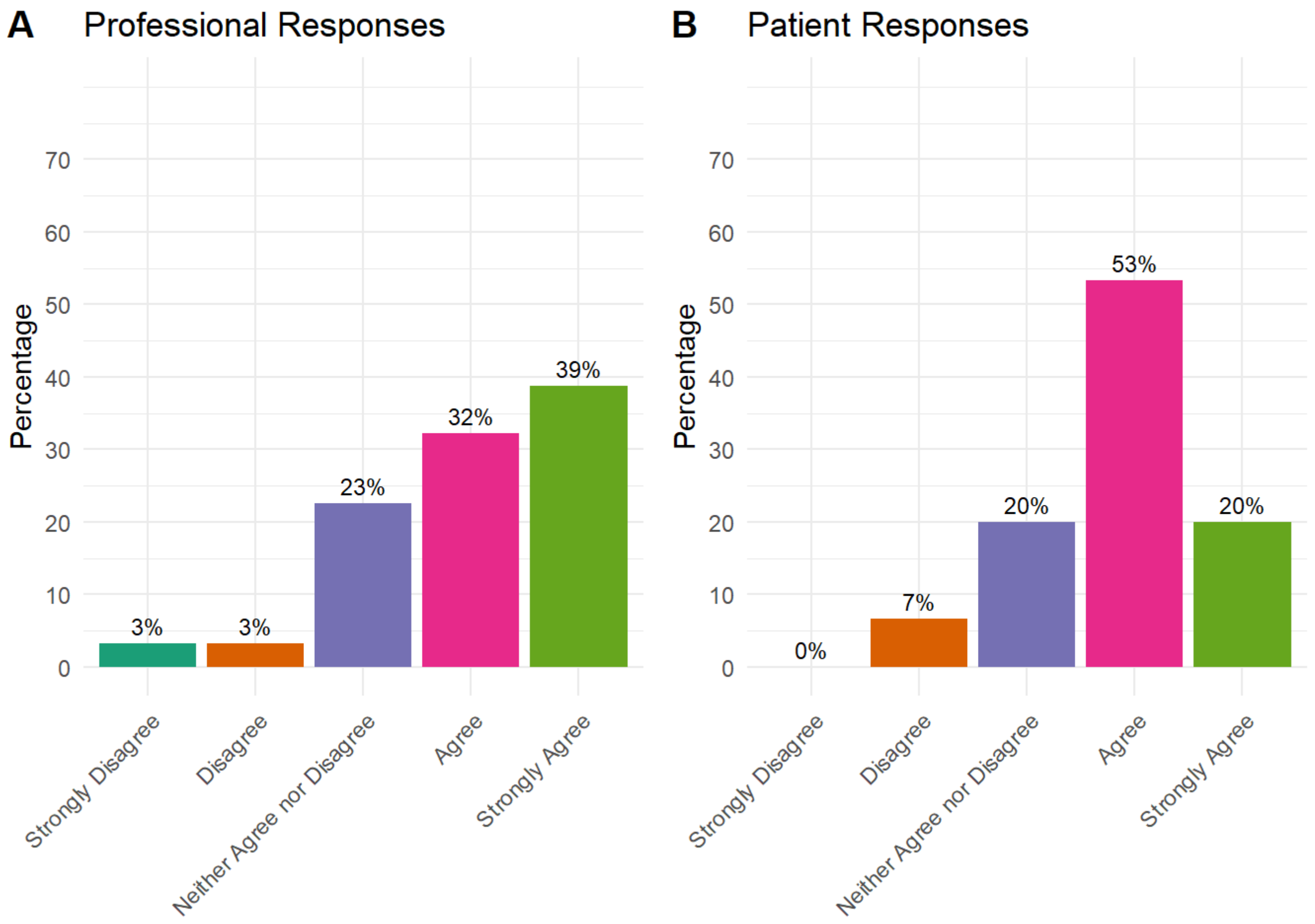


Pre-operative information should: Be provided separately for THR and TKR – i.e., patients waiting for THR should receive group education separately from patients waiting for TKR

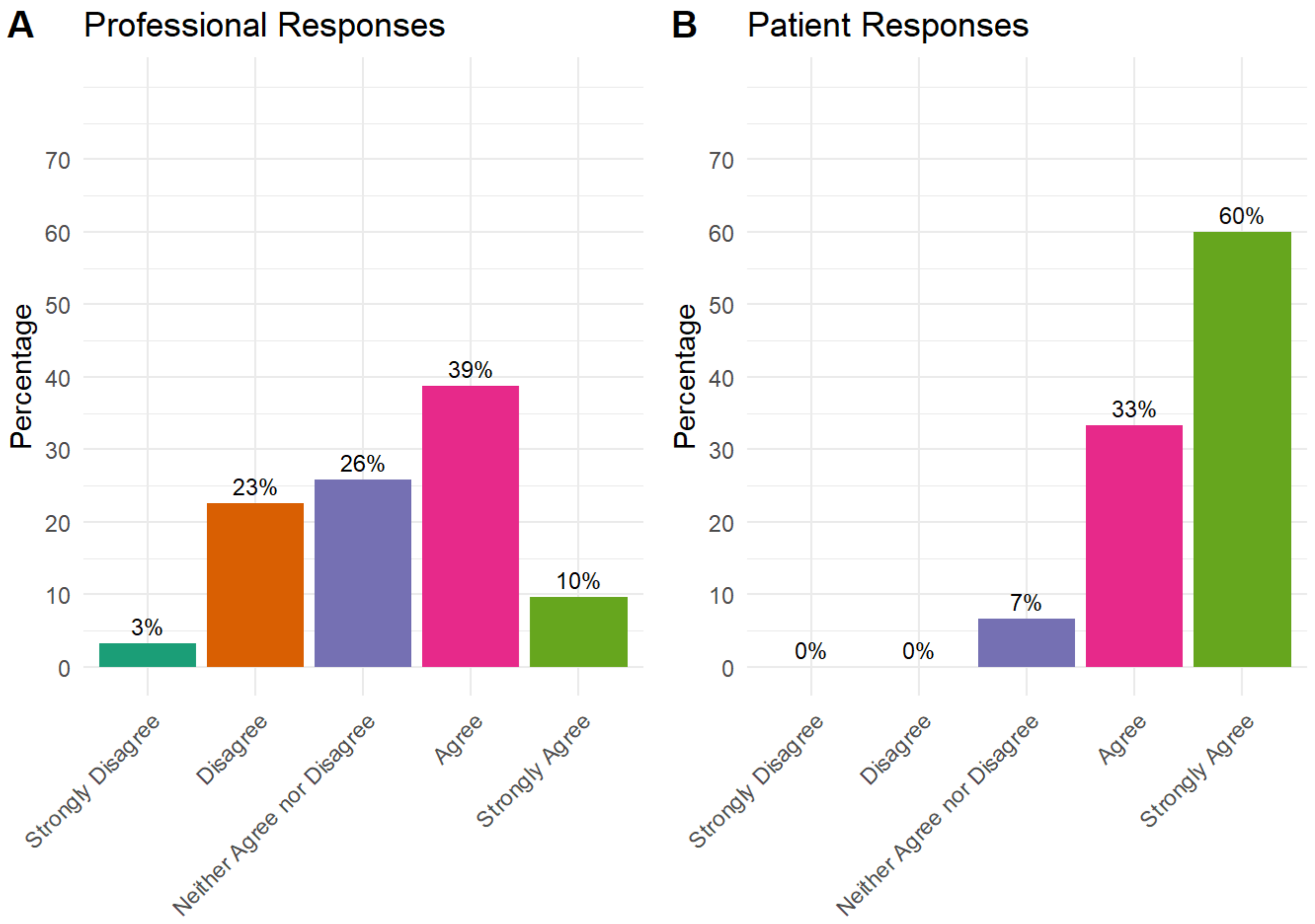


Pre-operative information should: Provide an opportunity for the patient's questions to be addressed

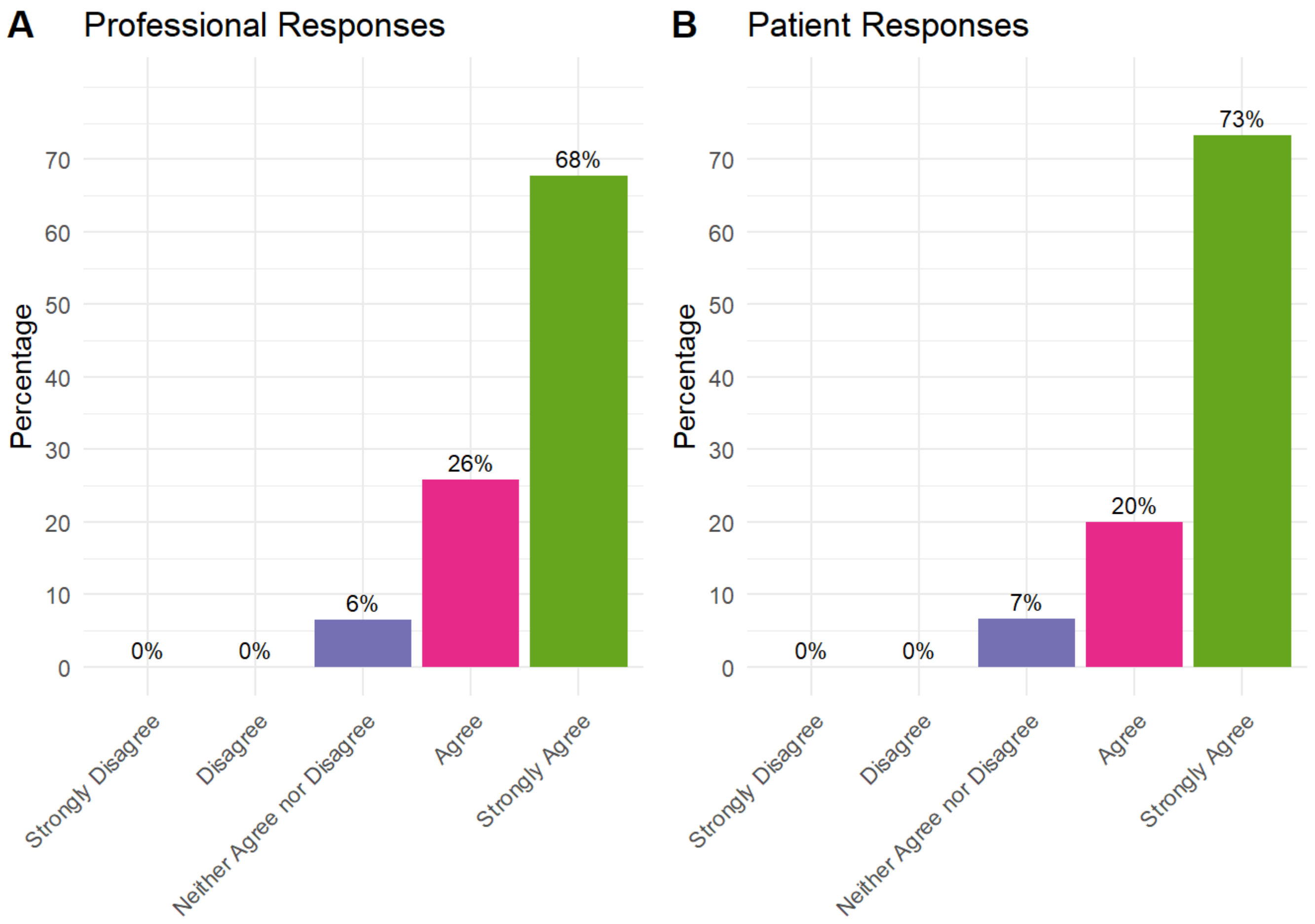


Pre-operative information should: Provide an opportunity for a family member or friend of the patient to be involved

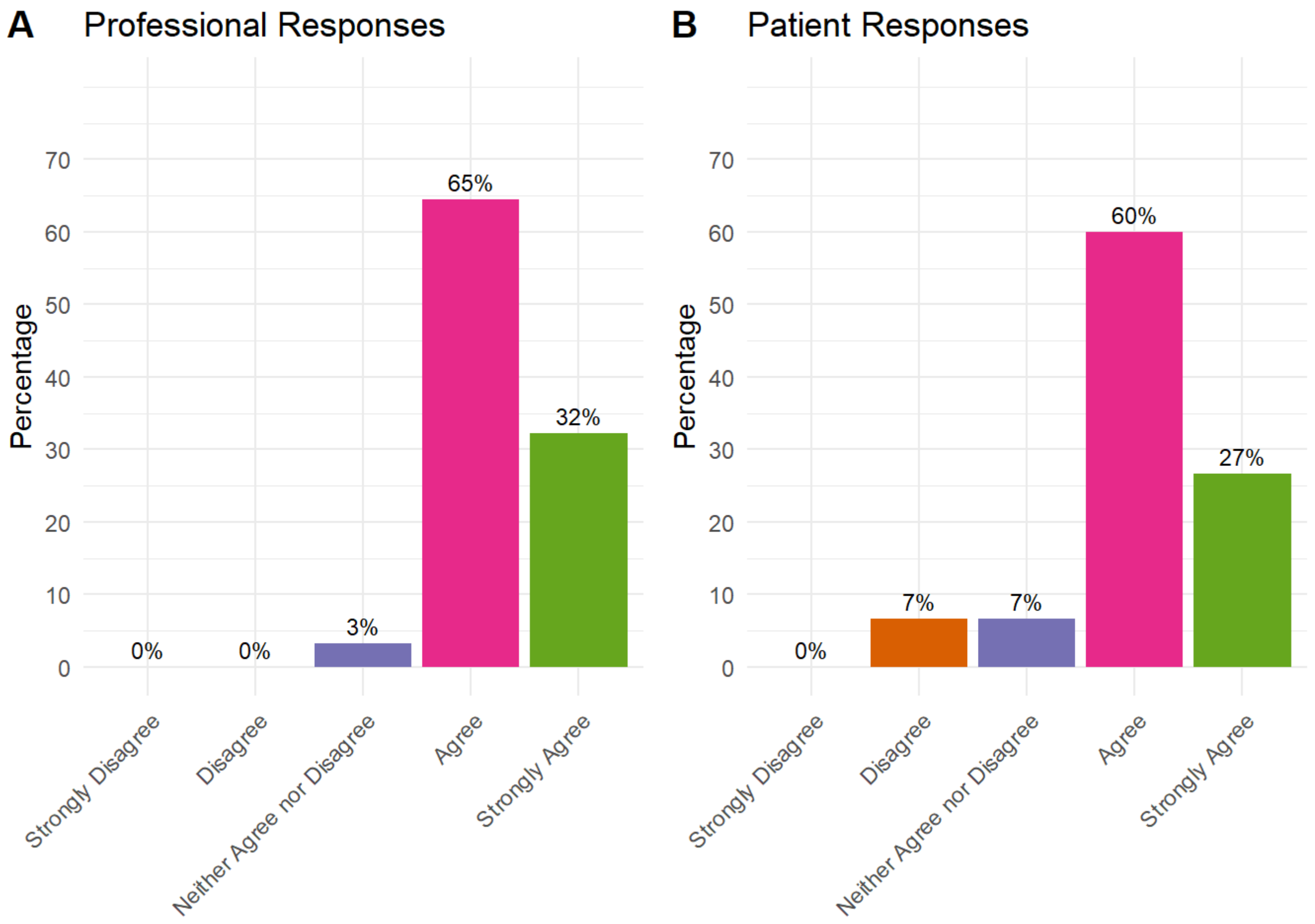


Pre-operative information should: Be delivered, at least partly, within four weeks of the patient's surgery

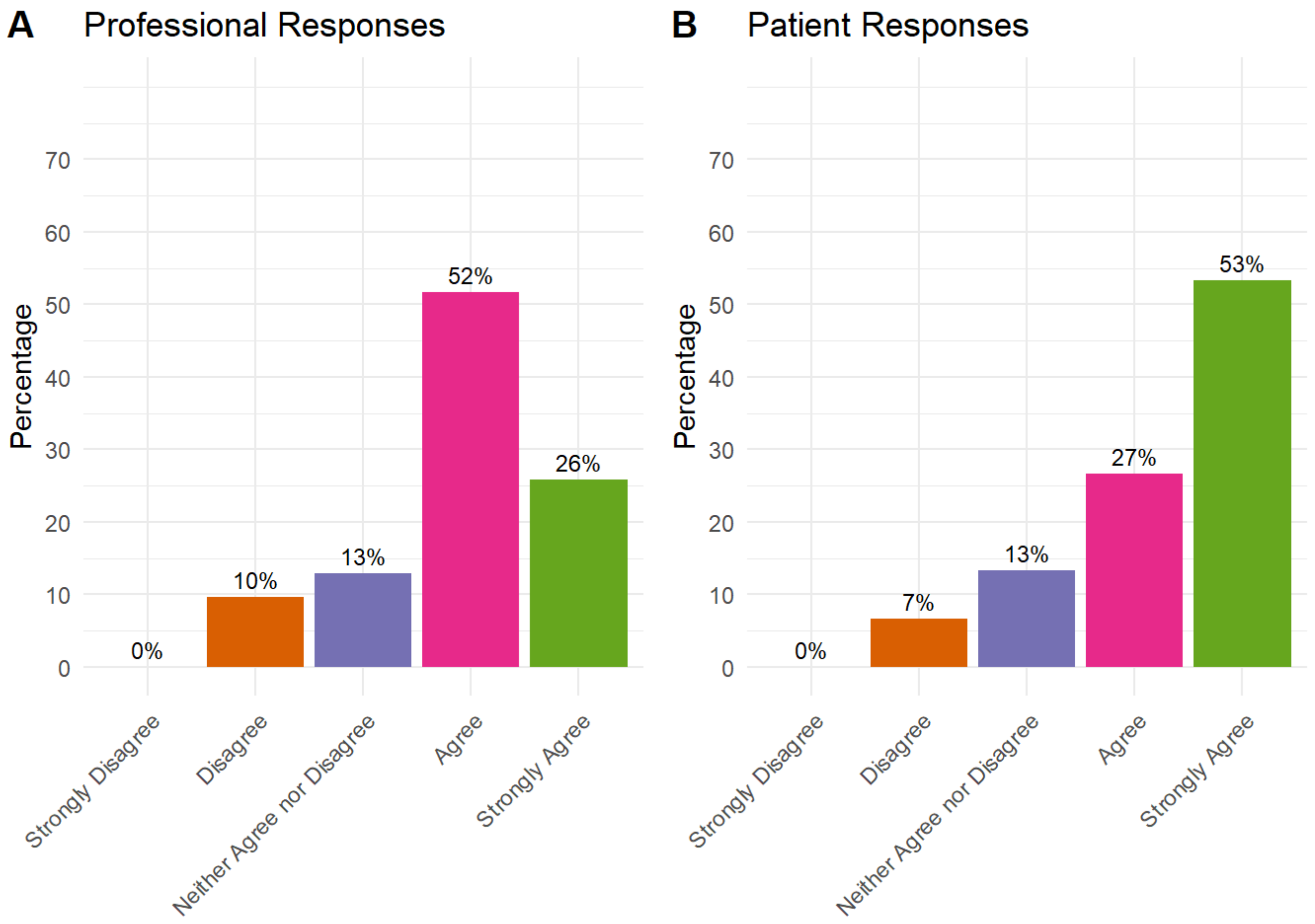


### A pre-operative exercise programme should include

A pre-operative exercise programme should include: Leg strengthening exercises

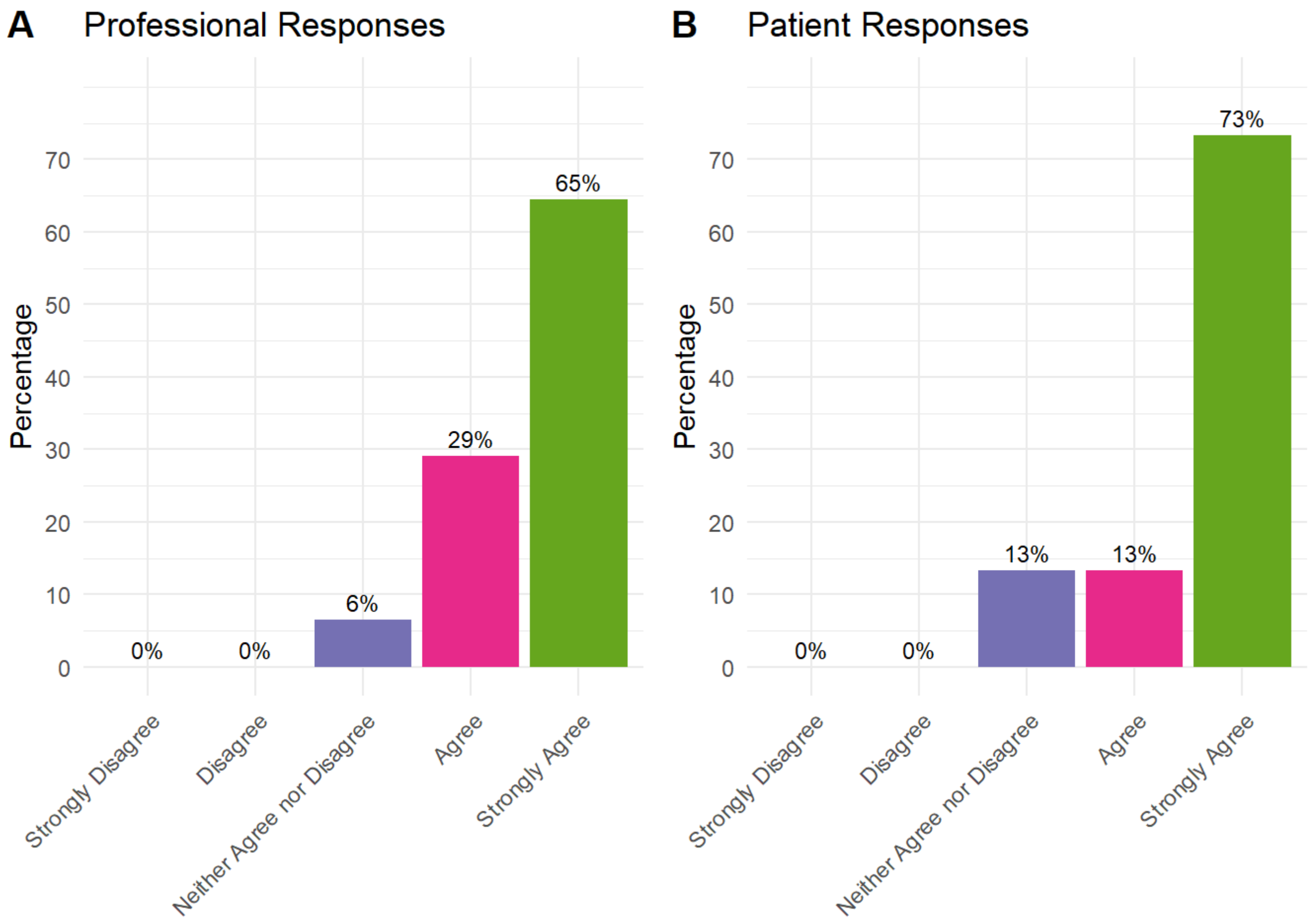


A pre-operative exercise programme should include: Balance exercises

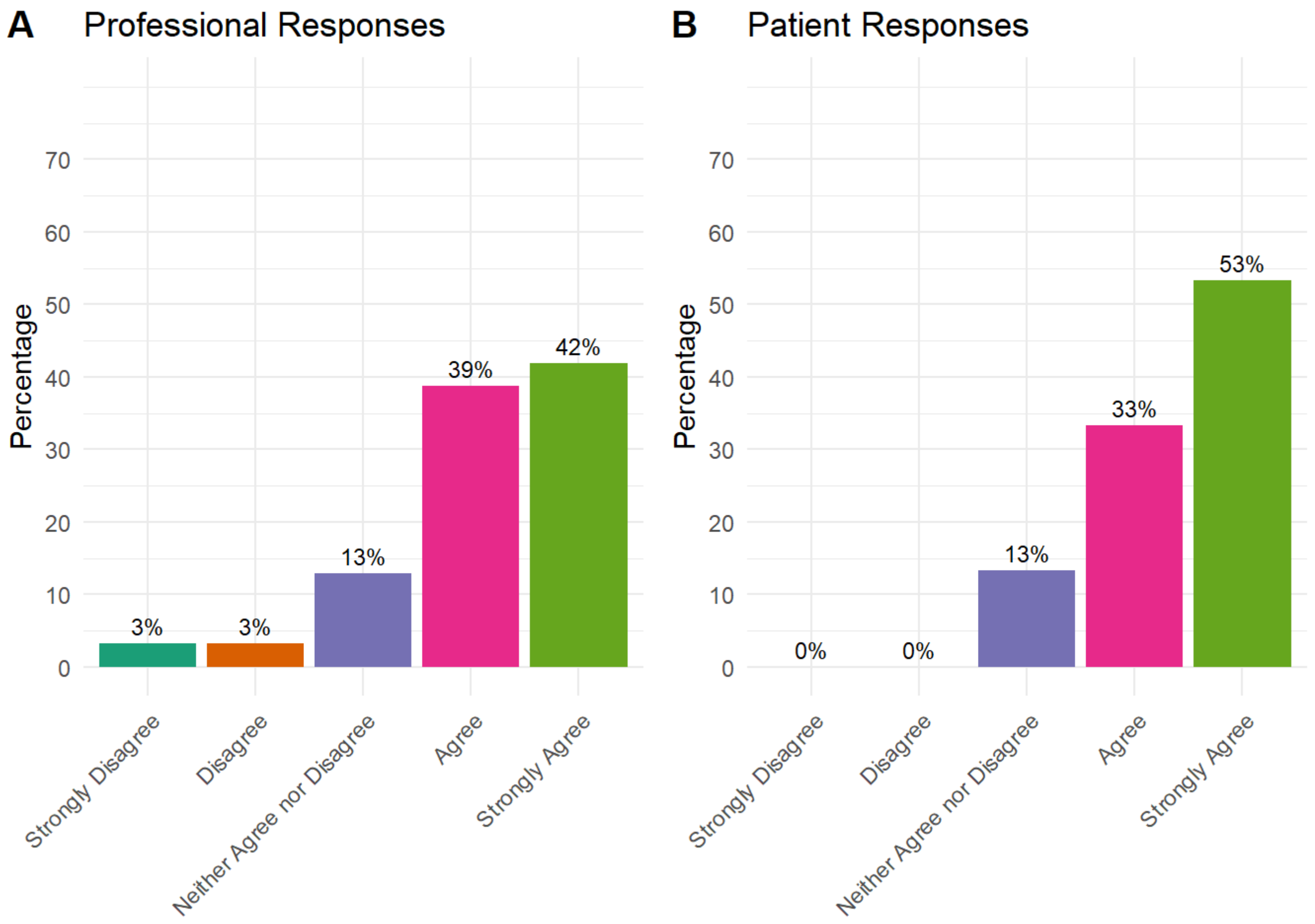


A pre-operative exercise programme should include: Functional movement exercises - i.e., sit to stand exercises

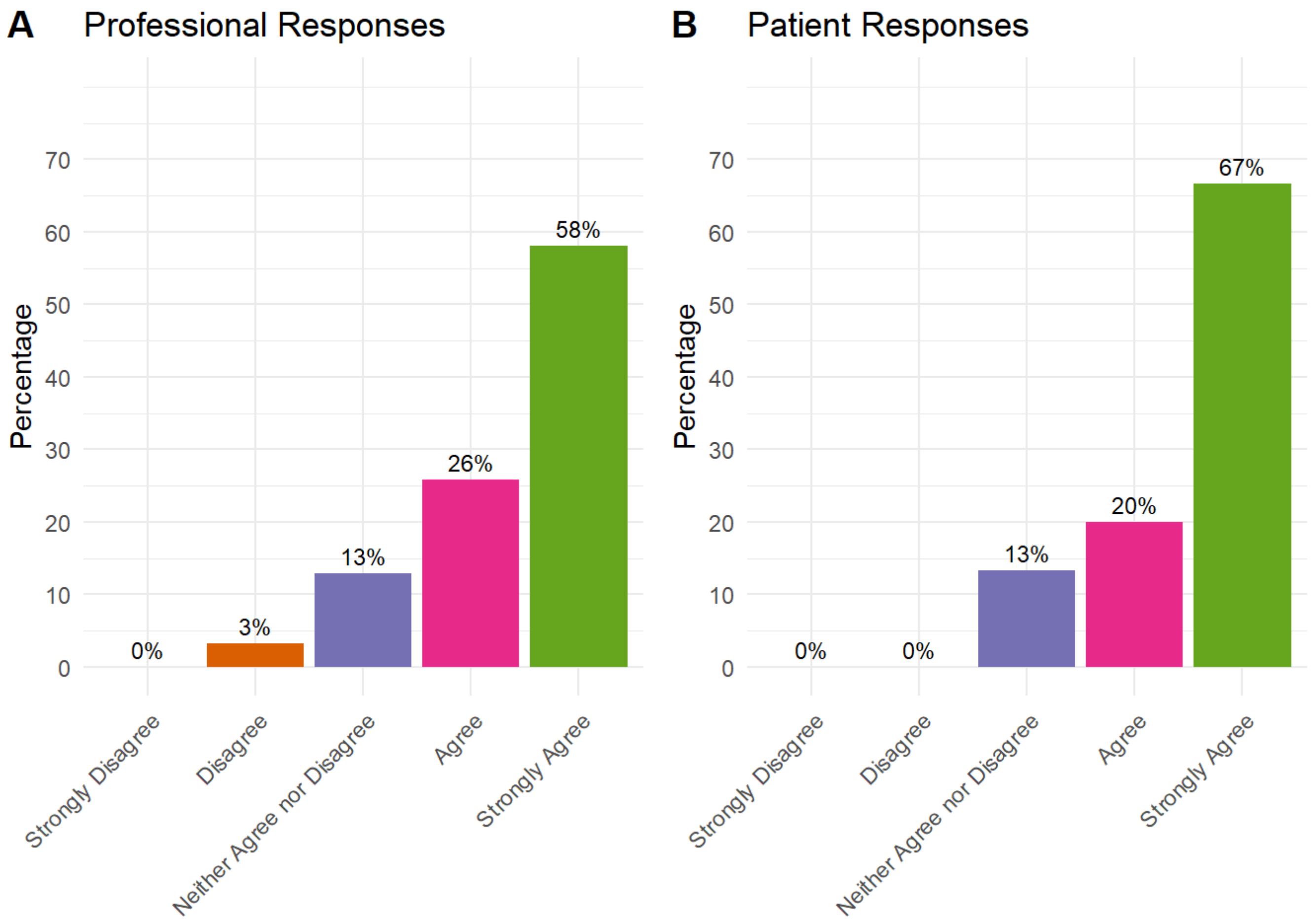


A pre-operative exercise programme should include: Cardiovascular exercises

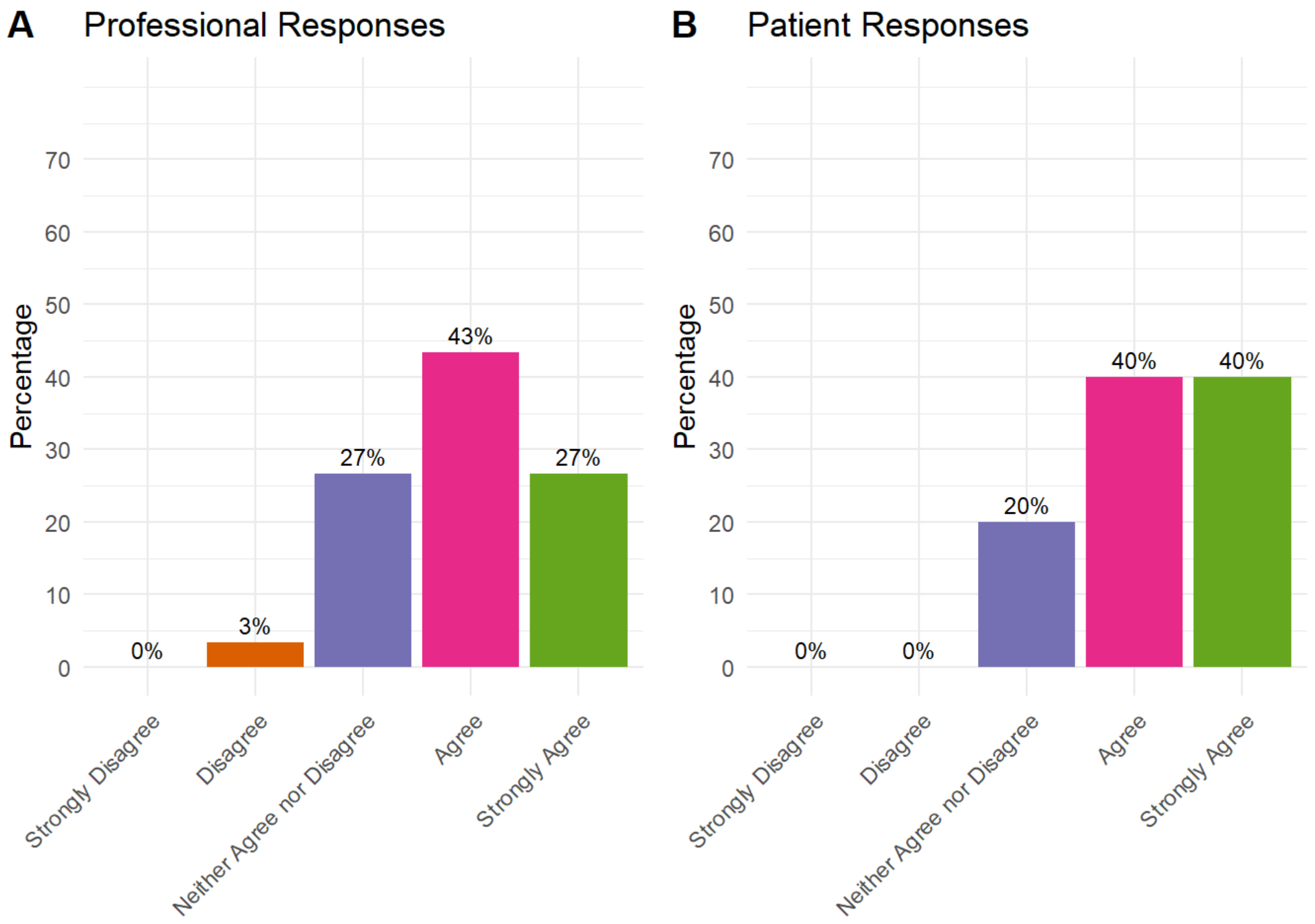


A pre-operative exercise programme should include: Core control exercises

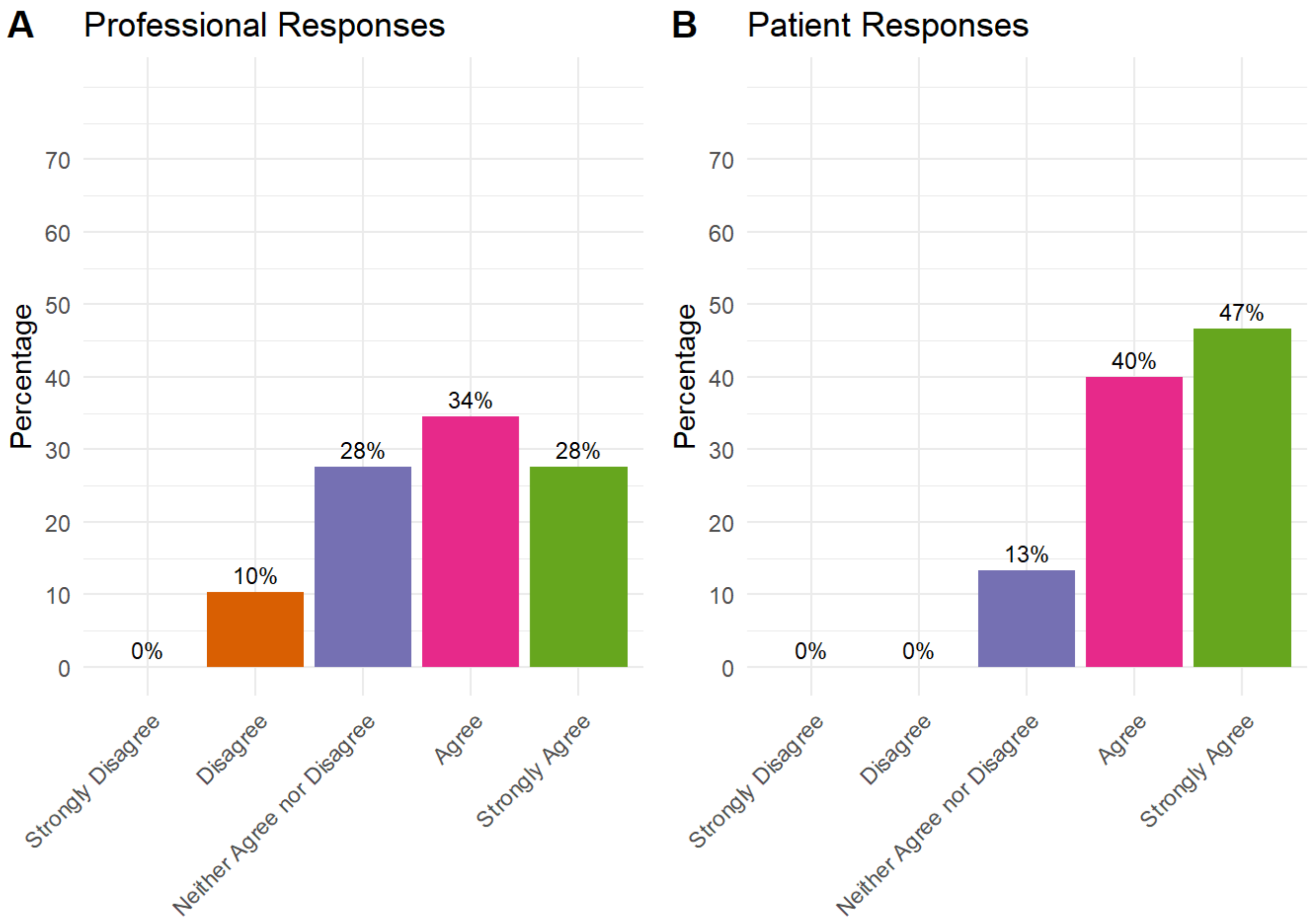


A pre-operative exercise programme should include: Walking practice with walking aids

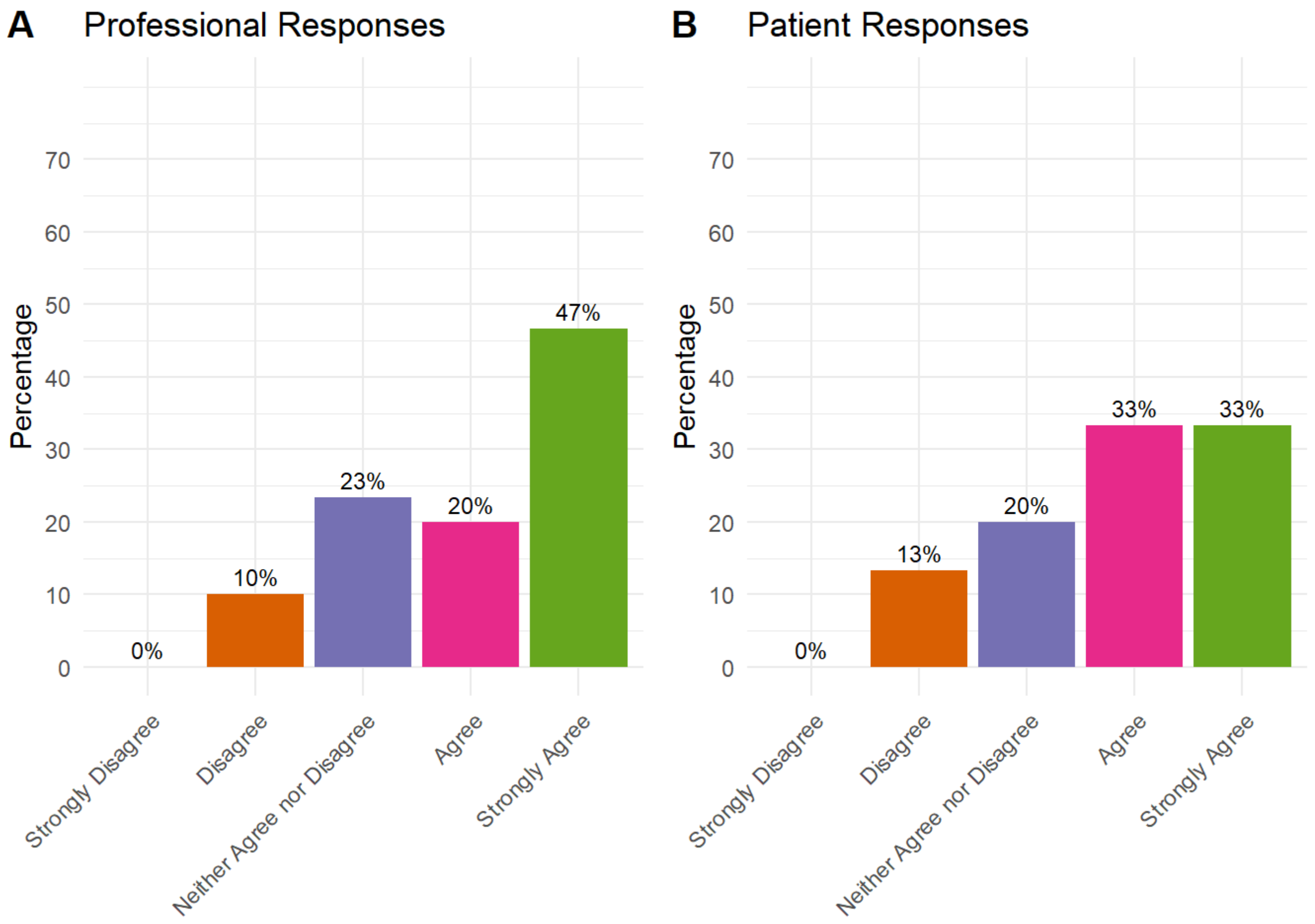


A pre-operative exercise programme should include: Training on steps

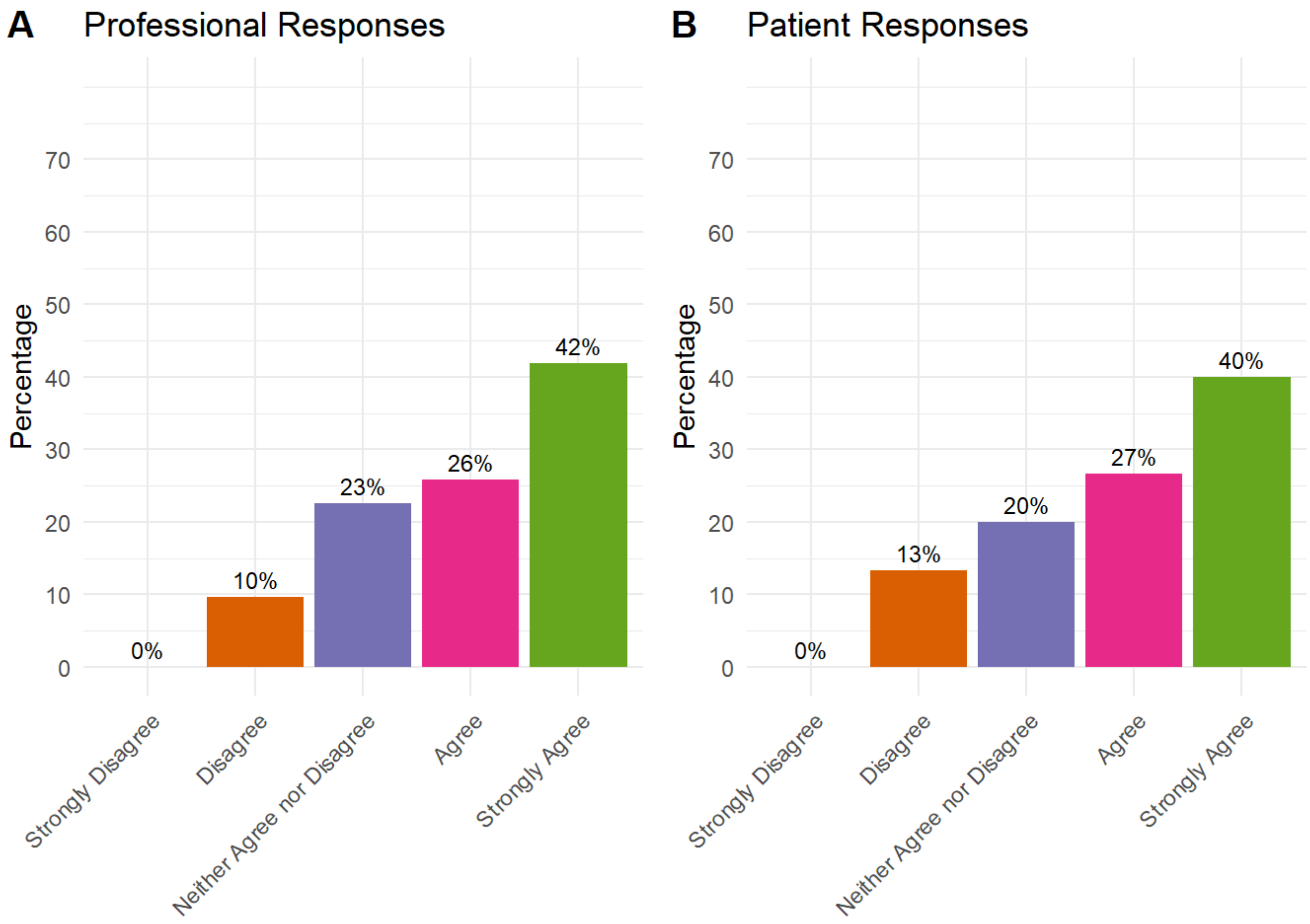


A pre-operative exercise programme should include: Practicing post operative exercises

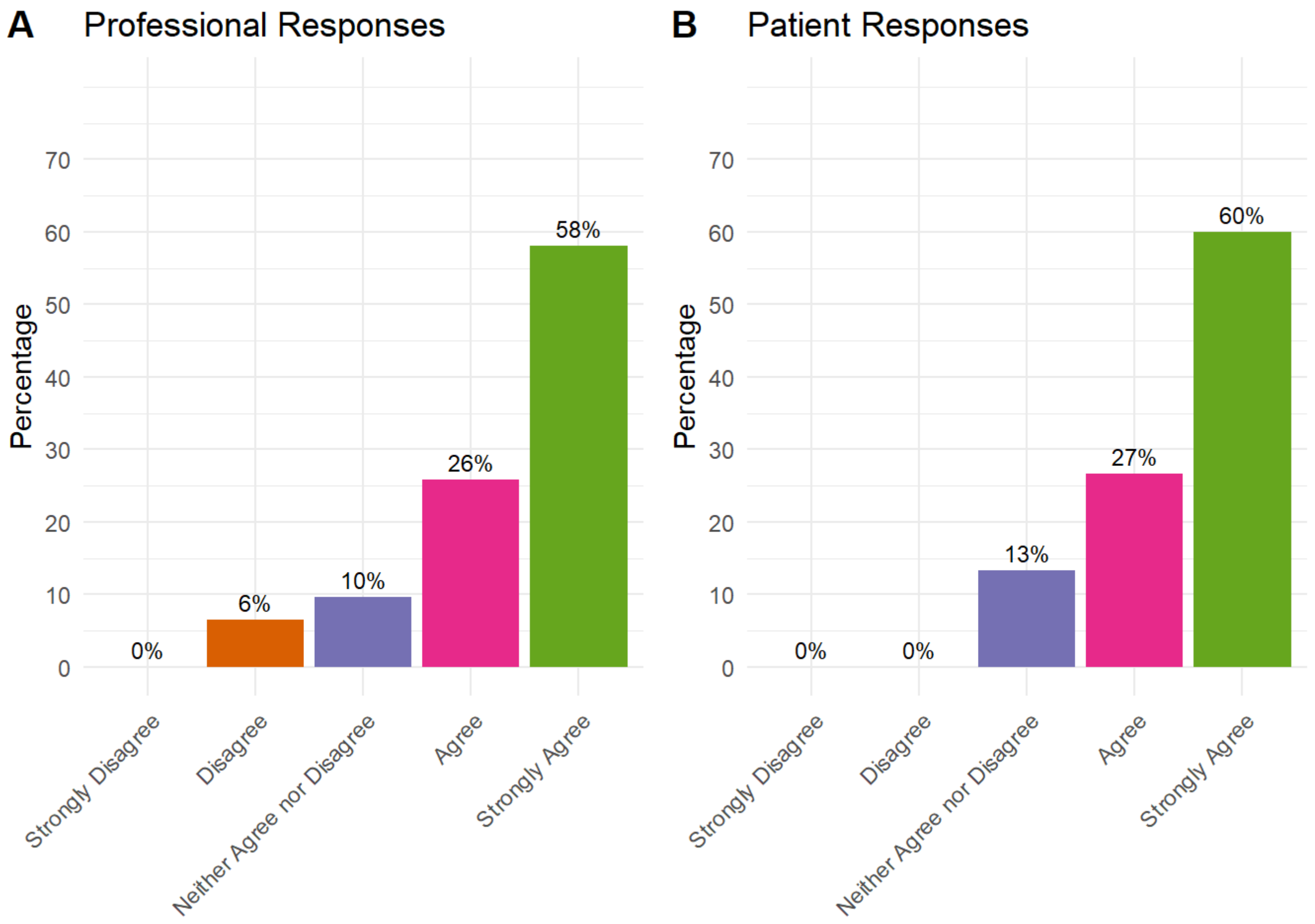


A pre-operative exercise programme should include: Delivery should be tailored according to each patient's individual needs

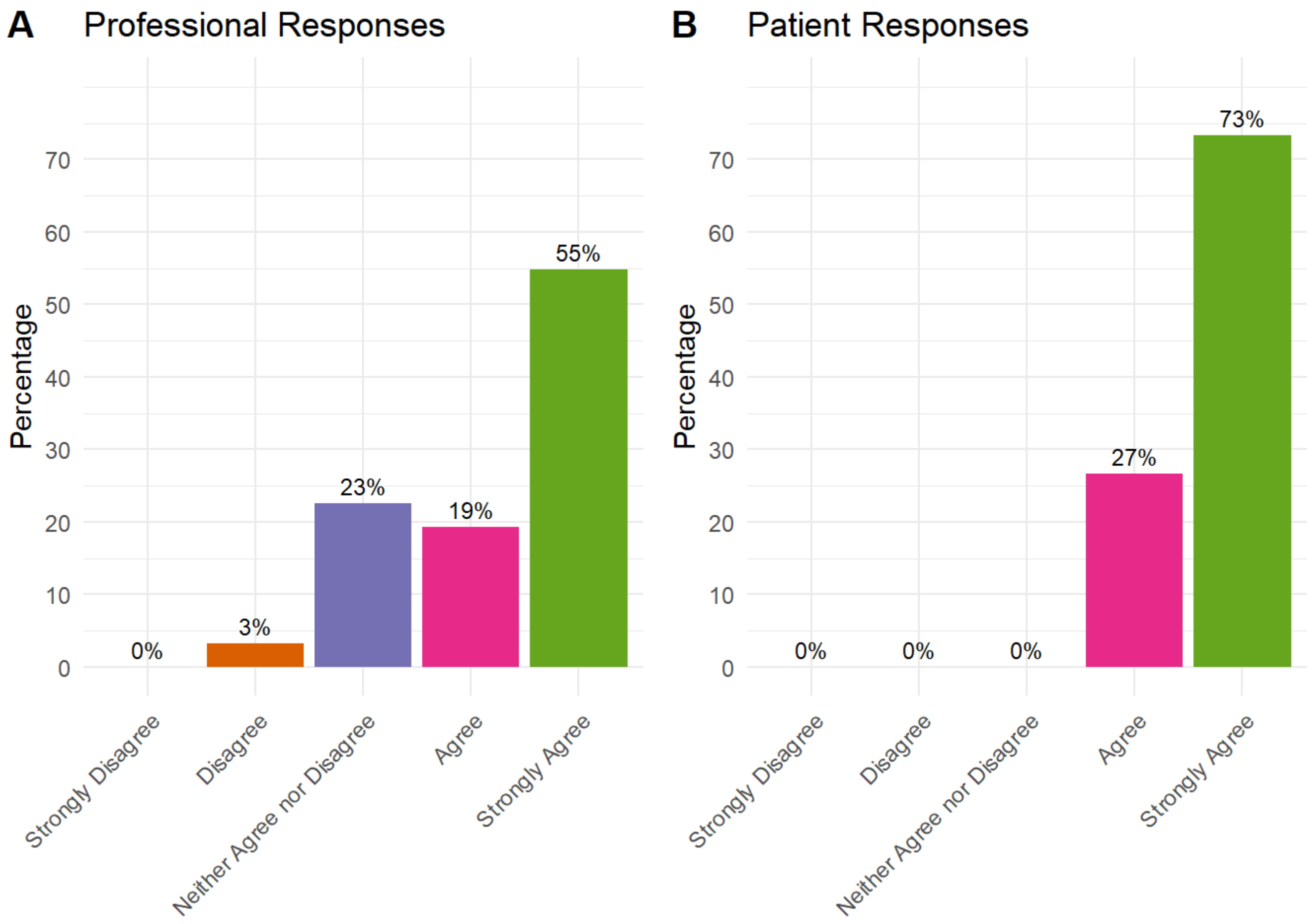


A pre-operative exercise programme should include: Leg strengthening exercises

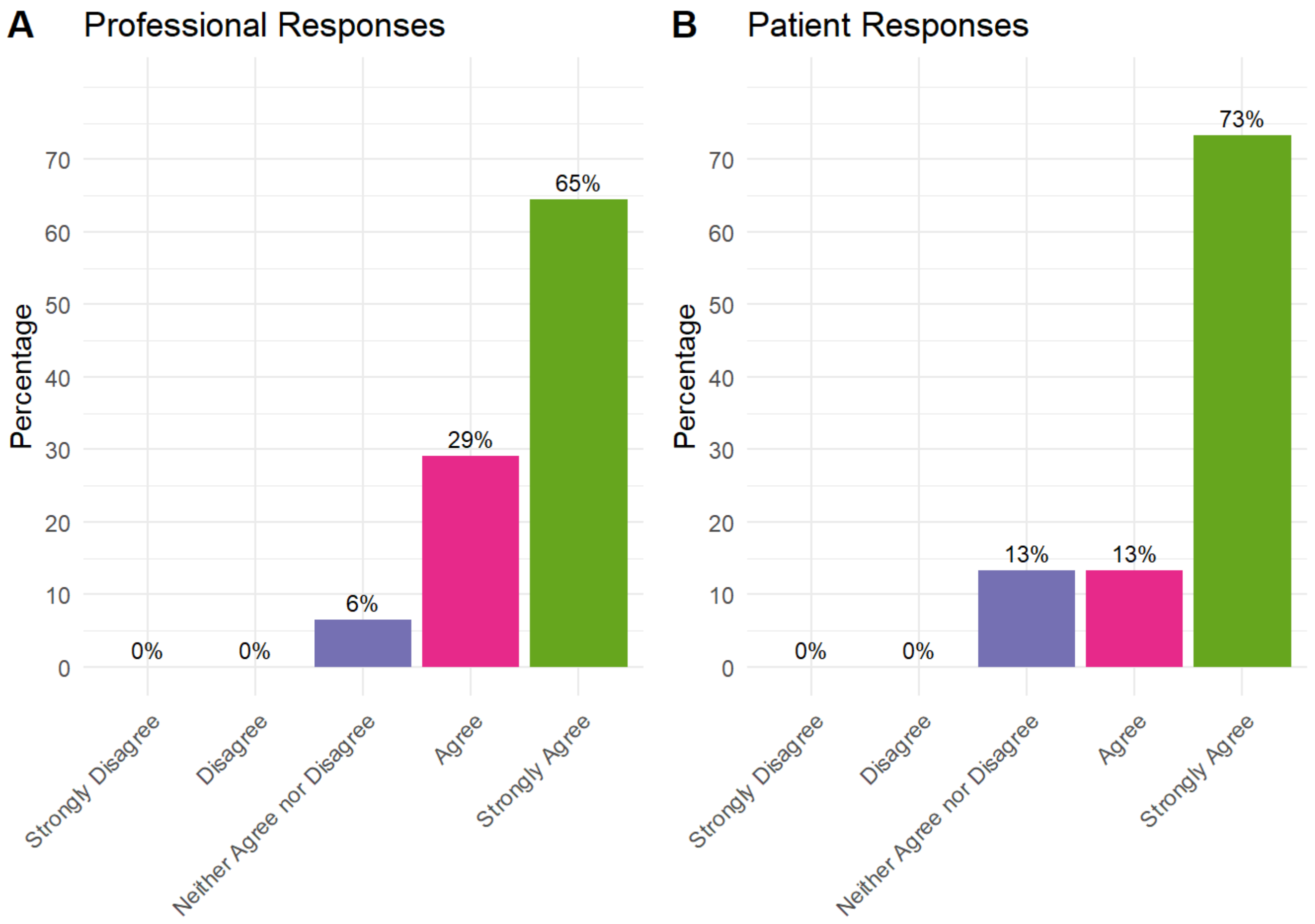


A pre-operative exercise programme should include: Delivery should be standardised across Scotland

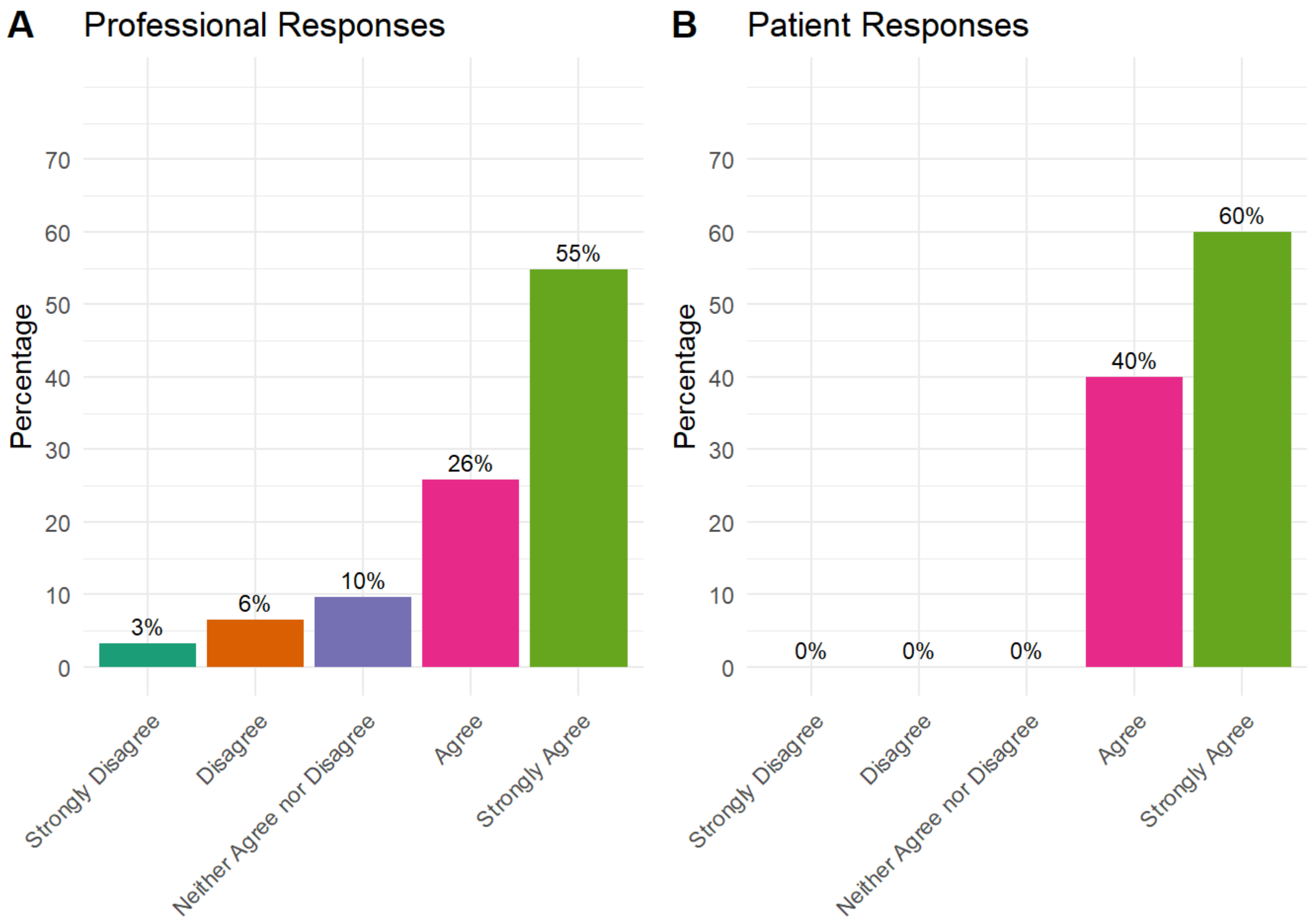


Round 2

### Information on preparing for surgery should cover

Information on preparing for surgery should cover: Optimising management of diabetes (Round 1: Patients 43% agreement; Professionals 81% agreement)

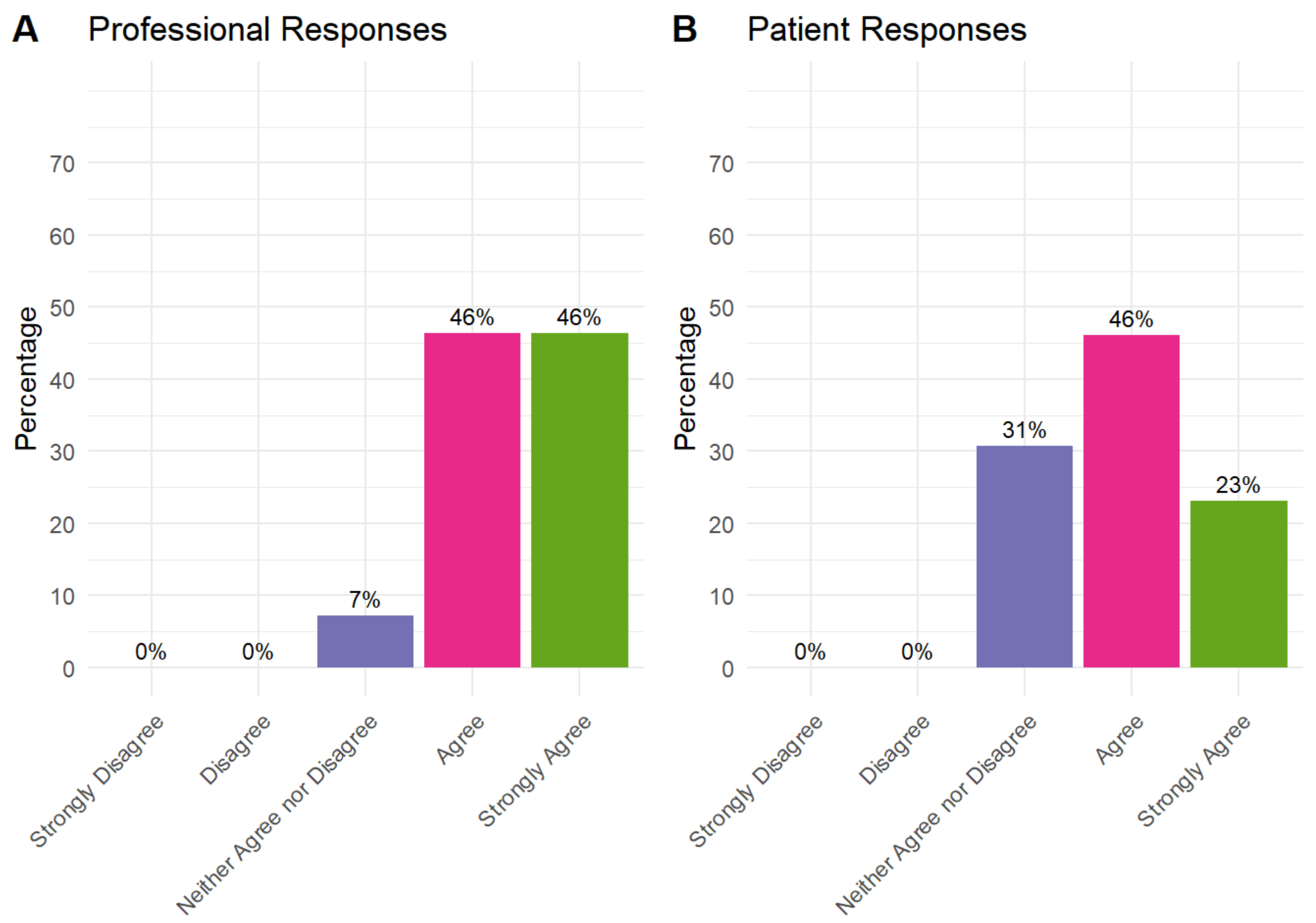


Information on preparing for surgery should cover: Identifying and arranging any social care provision (e.g., care & support at home) (Patients 67% agreement; Professionals 94% agreement)

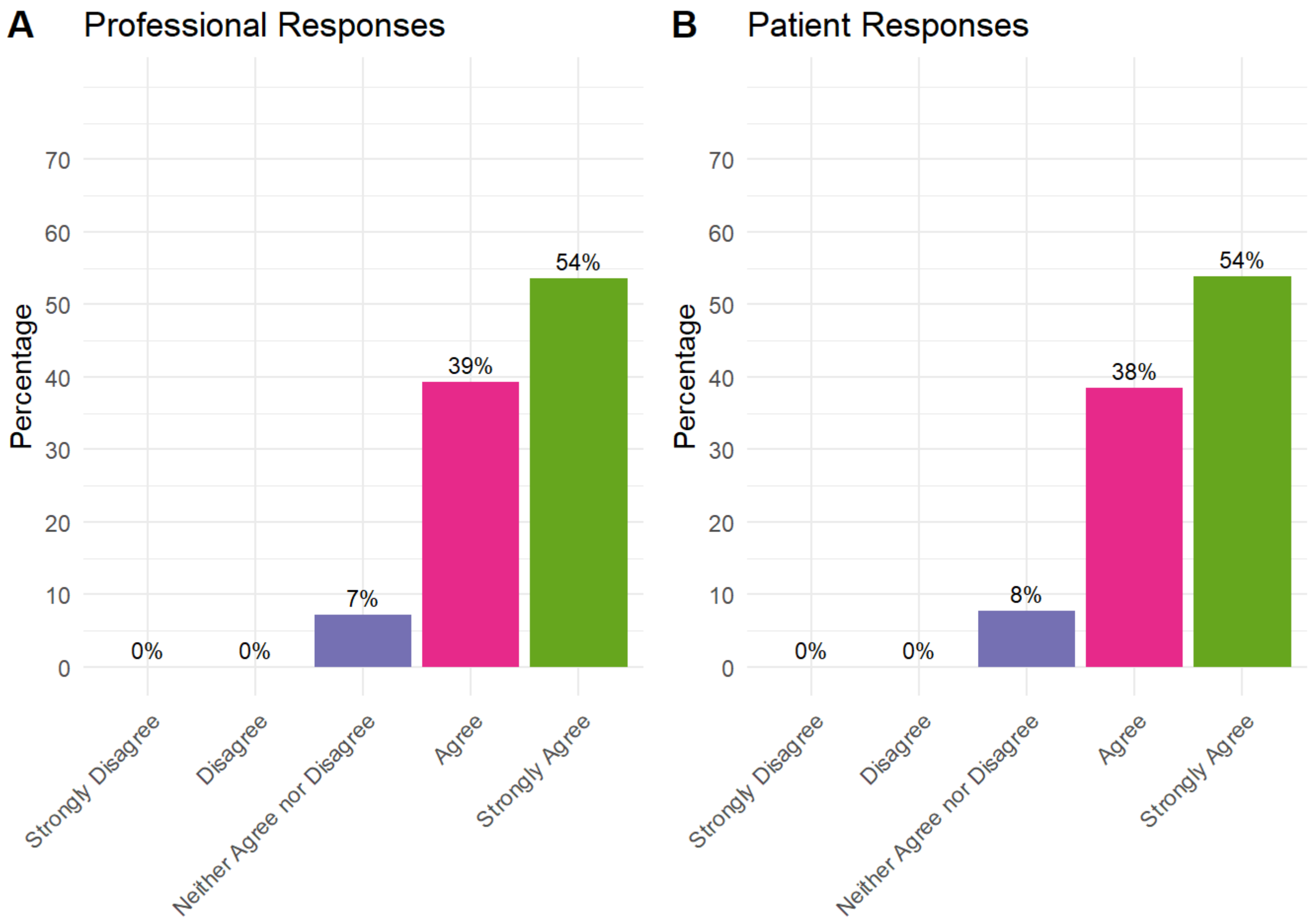


### Healthy lifestyle guidance should cover

Healthy lifestyle guidance should cover: Stopping smoking (Patients 67%; Professionals 100%)

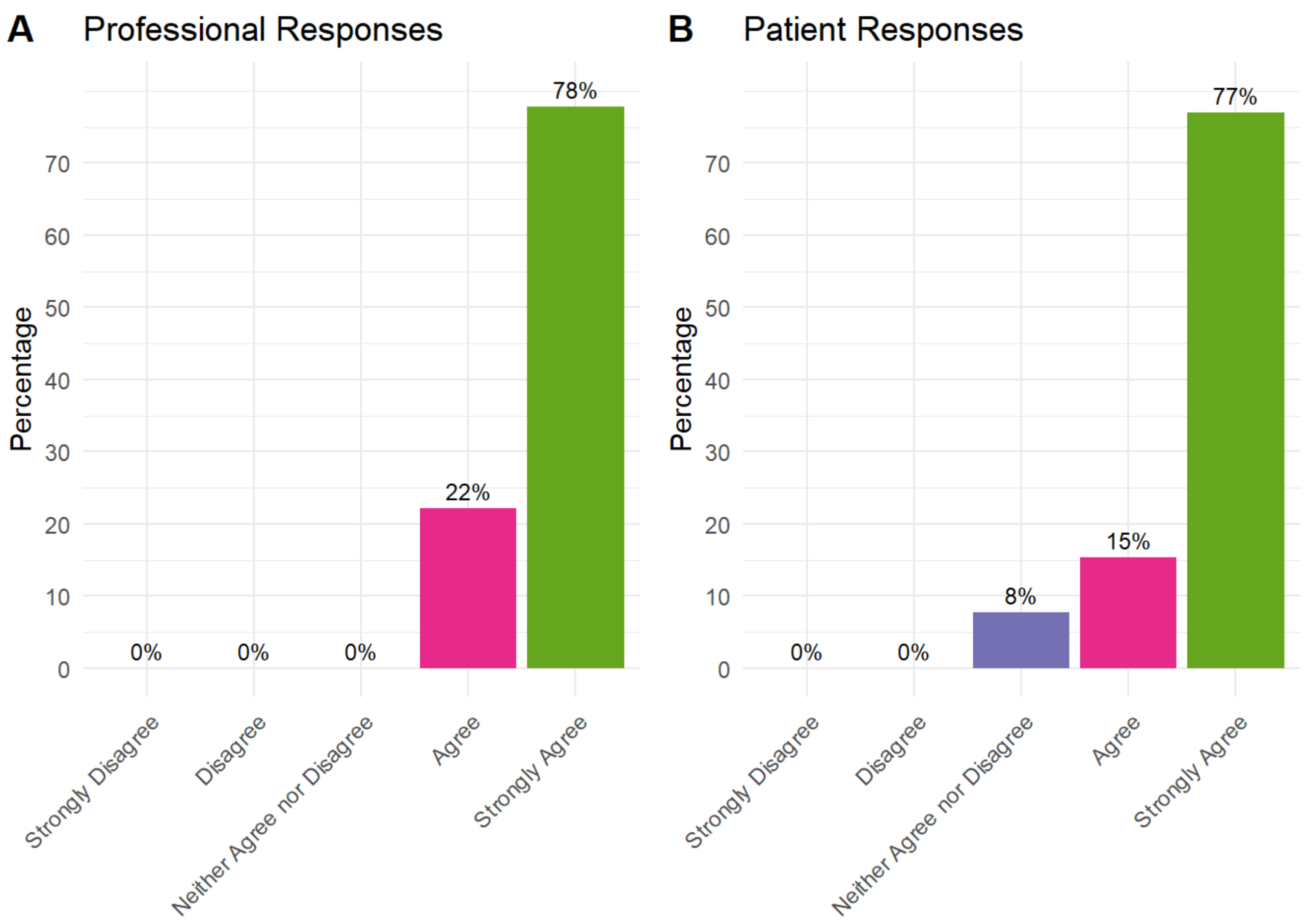


Healthy lifestyle guidance should cover: Recommended alcohol guidelines and cutting down on drinking (patients 67%; Professionals 94%)

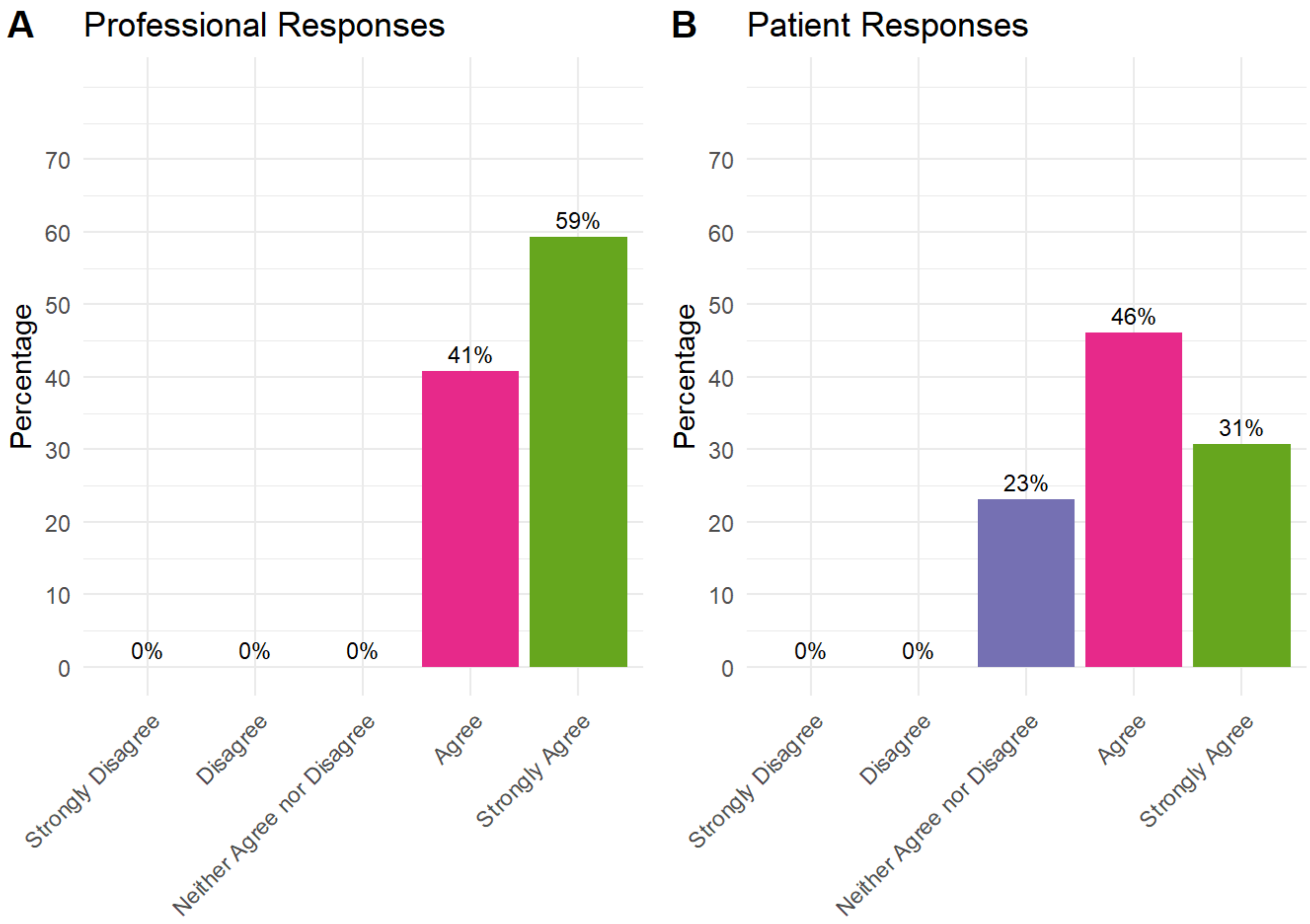


Healthy lifestyle guidance should cover: Mental wellbeing

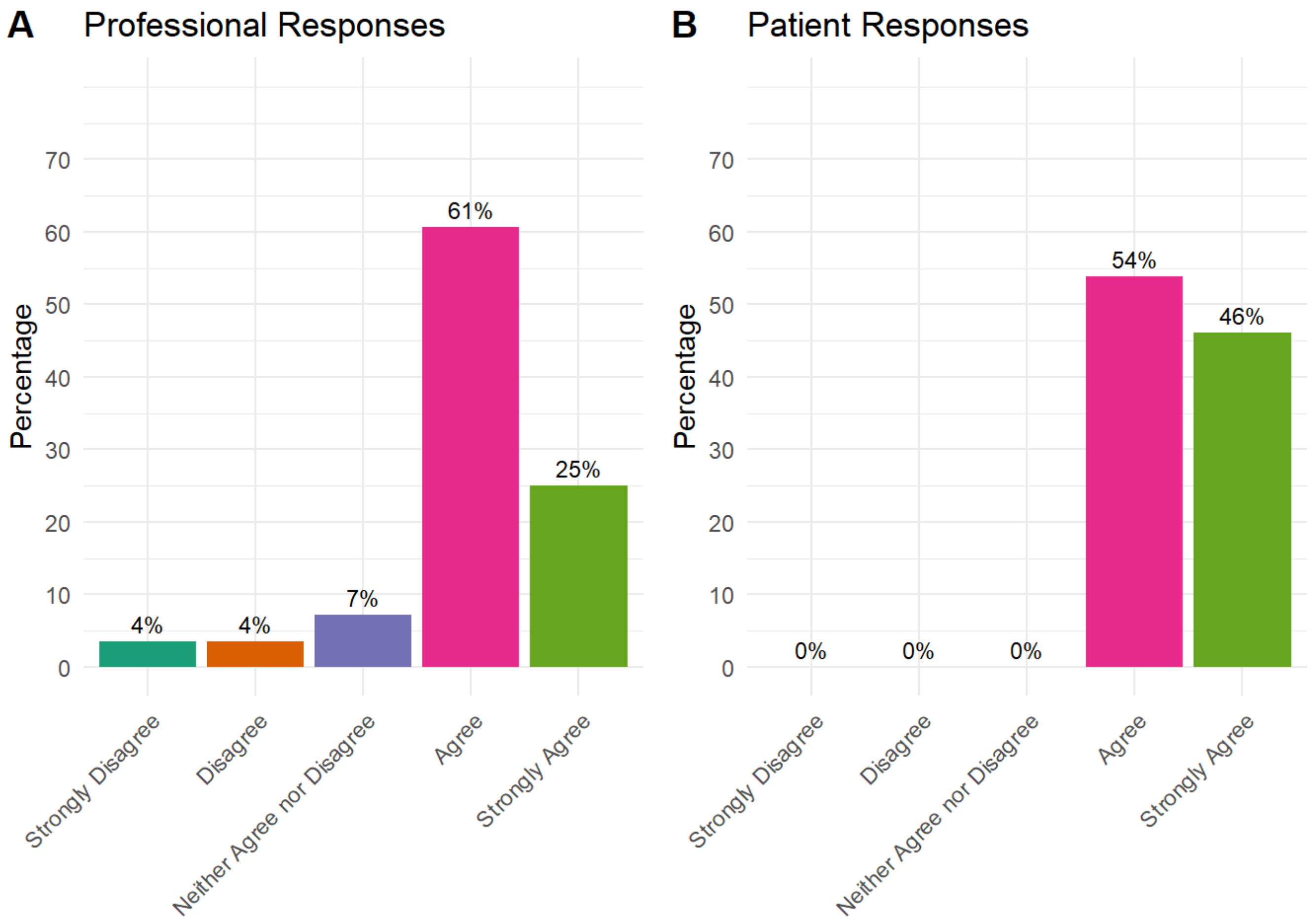


Healthy lifestyle guidance should cover: Sleep hygiene

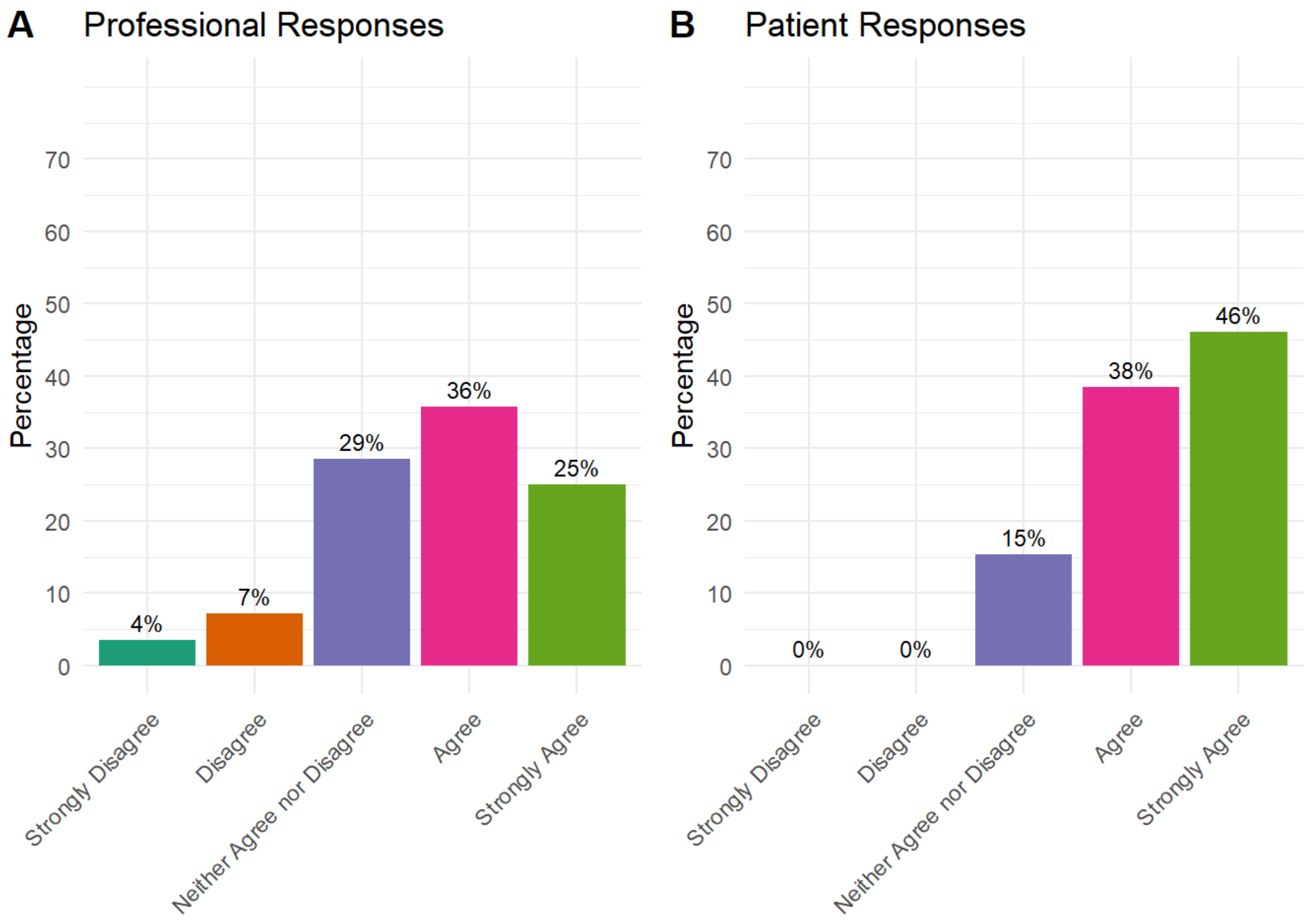


Healthy lifestyle guidance should cover: Signposting to information and sources of support for healthy lifestyle guidance

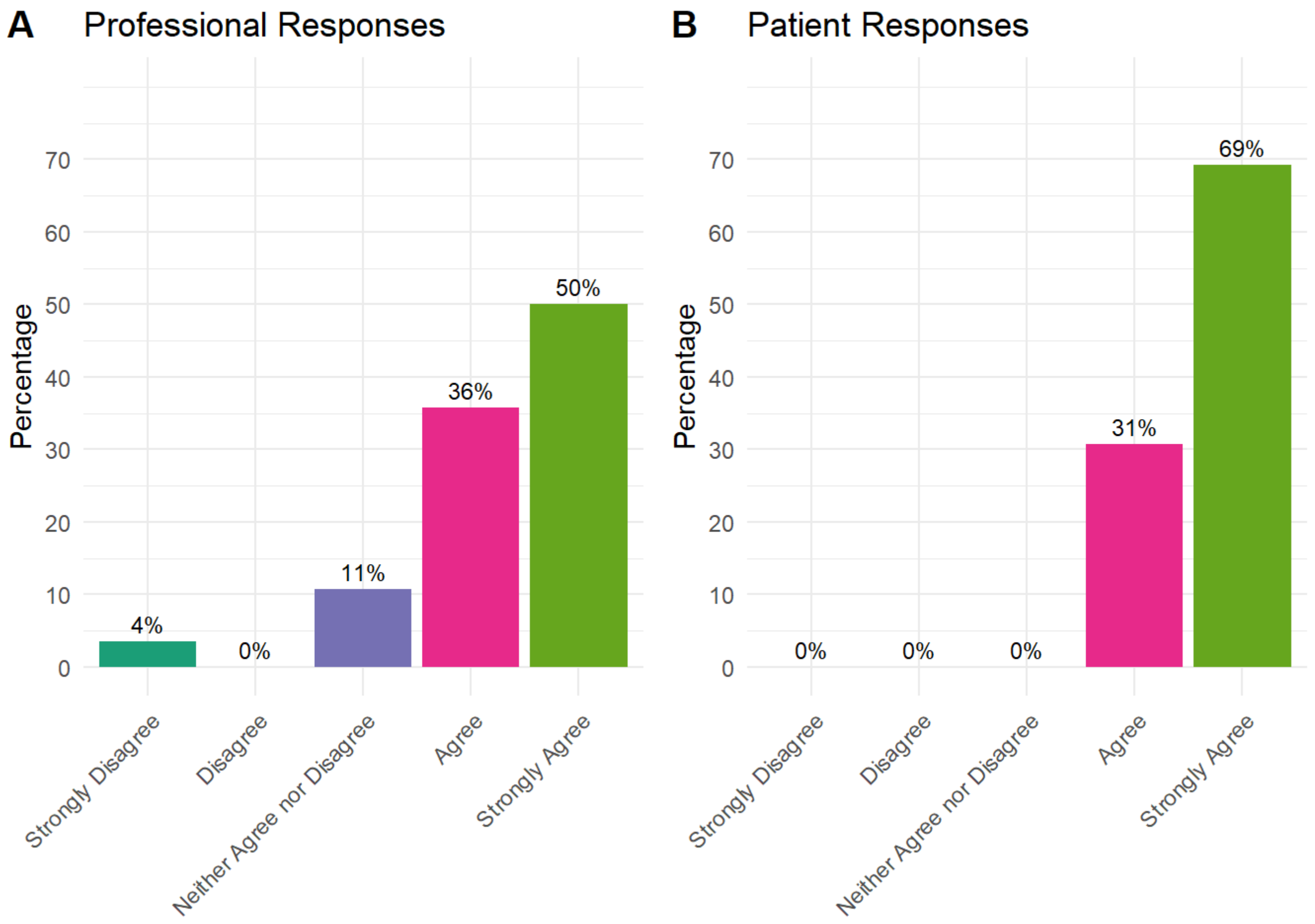

A
Professional Responses
Percentage
70
60
50
40
30
20
10
0
4%
0%
11%
36%
50%
Strongly Disagree
Disagree
Neither Agree nor Disagree
Agree
Strongly Agree
B
Patient Responses
Percentage
70
60
50
40
30
20
10
0
0%
0%
0%
31%
69%
Strongly Disagree
Disagree
Neither Agree nor Disagree
Agree
Strongly Agree
