## Supplementary File 3_Equator Checklist for "Consensus recommendations for supporting people waiting for total hip and knee arthroplasty in Scotland: A modified Delphi study"

**AUTHOR CHECKLIST** ***Authors of all papers reporting clinical research should submit this checklist together with their manuscript and the Reporting Guideline Checklist found on the EQUATOR site (http://www.equator-network.org/).***

This checklist identifies recognised guidelines for scientific reporting, which authors should use to prepare their manuscript *(required for systematic reviews and original research)*

| ***Standards of reporting*** | The editors require that manuscripts adhere to recognised reporting guidelines relevant to the research design used. These identify matters that should be addressed in your paper. Please indicate which guidelines you have referred to.<br><br>These are not quality assessment frameworks and your study need not meet all the criteria implied in the reporting guideline to be worthy of publication in the MATH. The checklists do identify essential matters that should be considered and reported upon. For example, a controlled trial may or may not be blinded but it is important that the paper identifies whether or not participants, clinicians and outcome assessors were aware of treatment assignments.<br><br>**You are also required to submit a checklist from the appropriate reporting guideline (available on the EQUATOR website (http://www.equator-network.org/) together with your paper as a guide to the editors.<br><br>*Reporting guidelines endorsed by MATH are listed below:* | **Guideline referred to** | **Checklist submitted**** |
|---|---|---|---|
| Randomised (and quasi-randomised) controlled trial | CONSORT – Consolidated Standards of Reporting Trials http://www.equator-network.org/reporting-guidelines/consort/ | | |
| Study of Diagnostic accuracy / assessment scale | STARD Standards for the Reporting of Diagnostic Accuracy studies http://www.equator-network.org/reporting-guidelines/stard/ | | |
| Systematic Review of Controlled Trials | PRISMA - Preferred Reporting Items for Systematic Reviews and Meta-Analyses http://www.equator-network.org/reporting-guidelines/prisma/ | | |
| Observational cohort, case control and cross sectional studies | STROBE **St**rengthening the **R**eporting of **Ob**servational Studies in **E**pidemiology http://www.equator-network.org/reporting-guidelines/strobe/ | | |
| Case Reports | CARE - Case Reports - http://www.care-statement.org/downloads/CAREchecklist-English.pdf | | |
| Statistical reporting | SAMPL - guidelines for statistical reporting – *no checklist exists currently but authors are encouraged to view the guidelines on the EQUATOR website http://www.equator-network.org/reporting-guidelines/sampl/* | | |
| | *Qualitative researchers might wish to consult the guideline listed below* | | |
| Qualitative studies | COREQ: Consolidated criteria for reporting qualitative research (http://www.equator-network.org/reporting-guidelines/coreq/) | | |
| Other (please give source) | CREDES<br>Jünger S, Payne SA, Brine J, Radbruch L, Brearley SG. Guidance on Conducting and REporting DElphi Studies (CREDES) in palliative care: Recommendations based on a methodological systematic review. Palliat Med. 2017;31: 684–706. doi:10.1177/0269216317690685 | | Y |
| Not applicable (please elaborate) | | | |
