## Supplementary File_CREDES Checklist for "Consensus recommendations for supporting people waiting for total hip and knee arthroplasty in Scotland: A modified Delphi study"

**CREDES Checklist:**
**Recommendations for the Conducting and REporting of DElphi Studies (CREDES)**

| Items of reporting | Reported on page |
|---|---|
| *Purpose and rationale.* The purpose of the study should be clearly defined and demonstrate the appropriateness of the use of the Delphi technique as a method to achieve the research aim. A rationale for the choice of the Delphi technique as the most suitable method needs to be provided. | Page 3-4 |
| *Expert panel.* Criteria for the selection of experts and transparent information on recruitment of the expert panel, sociodemographic details including information on expertise regarding the topic in question, (non)response and response rates over the ongoing iterations should be reported. | Page 5-7, Table 1 and Table 2 |
| *Description of the methods.* The methods employed need to be comprehensible; this includes information on preparatory steps (How was available evidence on the topic in question synthesised?), piloting of material and survey instruments, design of the survey instrument(s), the number and design of survey rounds, methods of data analysis, processing and synthesis of experts' responses to inform the subsequent survey round and methodological decisions taken by the research team throughout the process. | Page 3-4, Page 6-9 |
| *Procedure.* Flow chart to illustrate the stages of the Delphi process, including a preparatory phase, the actual 'Delphi rounds', interim steps of data processing and analysis, and concluding steps. | Figure 1 |
| *Definition and attainment of consensus.* It needs to be comprehensible to the reader how consensus was achieved throughout the process, including strategies to deal with non-consensus. | Page 7-9 |
| *Results*. Reporting of results for each round separately is highly advisable in order to make the evolving of consensus over the rounds transparent. This includes figures showing the average group response, changes between rounds, as well as any modifications of the survey instrument such as deletion, addition or modification of survey items based on previous rounds. | Page 10-21, Tables 3-7 |
| *Discussion of limitations.* Reporting should include a critical reflection of potential limitations and their impact of the resulting guidance. | Page 23-24 |
| *Adequacy of conclusions.* The conclusions should adequately reflect the outcomes of the Delphi study with a view to the scope and applicability of the resulting practice guidance. | Page 24 |
| *Publication and dissemination*. The resulting guidance on good practice in palliative care should be clearly identifiable from the publication, including recommendations for transfer into practice and implementation. If the publication does not allow for a detailed presentation of either the resulting practice guidance or the methodological features of the applied Delphi technique, or both, reference to a more detailed presentation elsewhere should be made (e.g. availability of the full guideline from the authors or online; publication of a separate paper reporting on methodological details and particularities of the process (e.g. persistent disagreement and controversy on certain issues)). A dissemination plan should include endorsement of the guidance by professional associations and health care authorities to facilitate implementation. | Guidance is currently being developed and will be published separately |

Jünger S, Payne SA, Brine J, Radbruch L, Brearley SG. Guidance on Conducting and REporting DElphi Studies (CREDES) in palliative care: Recommendations based on a methodological systematic review. Palliat Med. 2017;31: 684–706. doi:10.1177/0269216317690685
